# Medications used among Non-Hospitalized Pregnant Women with COVID-19: a Prospective Individual Patient Data Meta-analysis in Europe and North America

**DOI:** 10.1101/2025.01.28.25321174

**Authors:** Odette de Bruin, Emeline Maisonneuve, Eimir Hurley, Hedvig ME Nordeng, Anick Bérard, Odile Sheehy, Padma Kaul, Mayura U Shinde, Austin Cosgrove, Jennifer G Lyons, Elizabeth Messenger-Jones, Maria E Kempner, Sengwee Toh, Wei Hua, José J Hernández-Muñoz, Leyla Sahin, Carolyn E Cesta, David Hägg, Rosa Gini, Olga Paoletti, Beatriz Poblador-Plou, Sue Jordan, Daniel Thayer, Clara L Rodríguez-Bernal, Francisco Sánchez-Sáez, Régis Lassalle, Marie-Agnès Bernard, Ema Alsina, Fariba Ahmadizar, Guillaume Favre, Alice Panchaud, Kitty WM Bloemenkamp, Kelly Plueschke, Corinne de Vries, Satu J Siiskonen, Miriam CJM Sturkenboom, the CONSIGN collaboration group

## Abstract

**Aim:** To estimate the prevalence of medication use in non-hospitalized pregnant women with COVID-19

**Methods:** A prospective two-stage individual patient meta-analysis across 10 data sources in Europe and North America studied medication use among non-hospitalized pregnant women with COVID-19 between January 2020 to December 2022. Comparisons were made between medication use within 30 days pre- and post-COVID-19 diagnosis in this cohort and two comparator groups: pregnant women without COVID-19, and non-pregnant women with COVID-19. Prevalence estimates were pooled using a random-effects model stratified by trimester.

**Results:** 50,335 non-hospitalized pregnant women with COVID-19 were identified. The pooled prevalence of antibacterial use in 3^rd^ trimester was higher post-COVID-19 diagnosis (6.8%, 95%CI 5.5-8.4, I^2^=94%) compared with the same women pre-COVID-19 (3.9%, 95%CI 3.1-4.9, I^2^=89%). Overall, pregnant women with COVID-19 had higher medication use compared to pregnant women without COVID-19, although these differences were not statistically significant. Post-COVID-19, antithrombotics prevalence was 4.5% (95%CI 1.1-16.5, I^2^=100%) among pregnant women with COVID-19 in 3^rd^ trimester, compared to 2.1% (95%CI 1.2-3.6, I^2^=99%) among those without COVID-19 in 3^rd^ trimester. Compared to non-pregnant women with COVID-19, pregnant women with COVID-19 were less likely to be prescribed analgesics, antiprotozoals, corticosteroids, psychoanaleptics and psycholeptics, and more likely to be prescribed antithrombotics, cough and cold and nasal preparations across all trimesters. High heterogeneity existed in nearly all analyses.

**Conclusion:** This international meta-analysis reveals low medication use and country-specific variations, enhancing insight into the management of COVID-19 in non-hospitalized pregnant women. Higher antithrombotics use post-COVID-19 suggests prophylactic treatment in this population.

## INTRODUCTION

The pandemic of the coronavirus disease 2019 (COVID-19) has raised significant concerns for pregnant women, their healthcare providers, governmental authorities, and medicines regulators. Pregnant women have been notably absent from most pivotal clinical trials assessing the effectiveness and safety of medications for treating and preventing COVID-19.^1,2^ Consequently, substantial knowledge gaps exist in understanding the effects of these treatments across all trimesters of pregnancy and their impact on maternal and perinatal outcomes. Moreover, there is a lack of understanding of the specific medications utilized by pregnant women with COVID-19 in real-world settings.

A systematic review published in October 2021 examined the pharmacological treatment of COVID-19 during pregnancy based on six studies involving 599 women. The treatments included antivirals, systemic corticosteroids, antibiotics, and immunotherapy. However, the review focused on inpatient prescriptions, and anticoagulation treatment was not considered. Despite large denominators, the number of pregnant women exposed to these interventions was small, making it impossible to conduct a meta-analysis and draw conclusions about pharmacological interventions for treating pregnant women with COVID-19.^3^ Another meta-analysis reported the prevalence of medications used during pregnancy among 1,742 patients, but only two of the studies with very low numbers of pregnant women were able to stratify by disease severity.^4^ To the best of our knowledge, no other published meta-analysis assesses medication use prevalence among non-hospitalized pregnant women with COVID-19.

The COVID-19 infectiOn aNd medicineS In preGnancy (CONSIGN) project was conducted by the EU PE&PV (Pharmacoepidemiology and Pharmacovigilance) Research Network and funded by the European Medicines Agency (EMA) to investigate the usage of medications among pregnant women with COVID-19 and its potential effects on unborn children. The project gathered evidence from the secondary use of data provided by the IMI-funded ConcePTION tools and network, referred to as CONSIGN electronic health records (EHR) study, as well as through primary data collections from the COVI-PREG and International Network of Obstetric Survey Systems (INOSS).^5–10^ The findings indicated that the utilization of medications to treat COVID-19 during pregnancy is rare, strongly associated with the severity of the disease, and subject to change over time since the onset of the pandemic.^5–11^ However, due to the limited and varying number of pregnant women receiving treatment for COVID-19, drawing definitive conclusions remains highly challenging.

To provide more precise estimates with reduced bias, following a landscape analysis of ongoing studies and discussions with the International Coalition for Medicines Regulatory Agencies (ICMRA), an international prospective two-stage individual patient data (IPD) meta-analysis was conducted within the CONSIGN project, incorporating all identified healthcare data sources that implemented the CONSIGN study protocol and standardized Statistical Analysis Plan.^12–14^ The focus of this meta-analysis was on outpatient medication usage, aiming to describe the utilization of medications for treating COVID-19 among non-hospitalized pregnant women across trimesters of pregnancy.

## METHODS

We conducted a prospective two-stage IPD meta-analysis, pooling the results from analyses conducted in different data sources based on secondary use of administrative and EHR data, as well as medical claims data containing information on pregnancies affected by COVID-19 between January 2020 and December 2022.^15,16^ To the extent possible, we adhered to the PRISMA-IPD and Cochrane guidelines for prospective meta-analysis.^17,18^

### Data sources

The EMA and CONSIGN leadership engaged through ICMRA with the U.S. Food and Drug Administration (FDA) and Health Canada to foster international collaboration on using medications and their effects on managing COVID-19 during pregnancy. Identified initiatives were requested to share their analysis plans and additional materials, and meetings were scheduled to discuss the eligibility of participation in the meta-analysis. To be eligible, networks or individual data sources needed access to population-based healthcare databases capable of identifying the start and end of pregnancies and linking them to COVID-19 diagnosis and medication records. Furthermore, they needed to implement fully or partially the CONSIGN EHR study protocol (EUPAS39438). ^12,13^ The CONSIGN EHR study is a multinational EHR and registry-based study conducted in Europe by the EU PE&PV Research Network.^6,10^

Fifty-one networks and data sources containing information on pregnant women with COVID-19 and medication use were identified. Among these, 16 were based on the secondary use of data. Five of the 16 healthcare data sources were excluded as they could not implement the CONSIGN protocol (Figure 1). In total, results from 11 data sources, available through three research initiatives were included in the meta-analysis and their characteristics are presented in Table S2.

**Figure 1.**
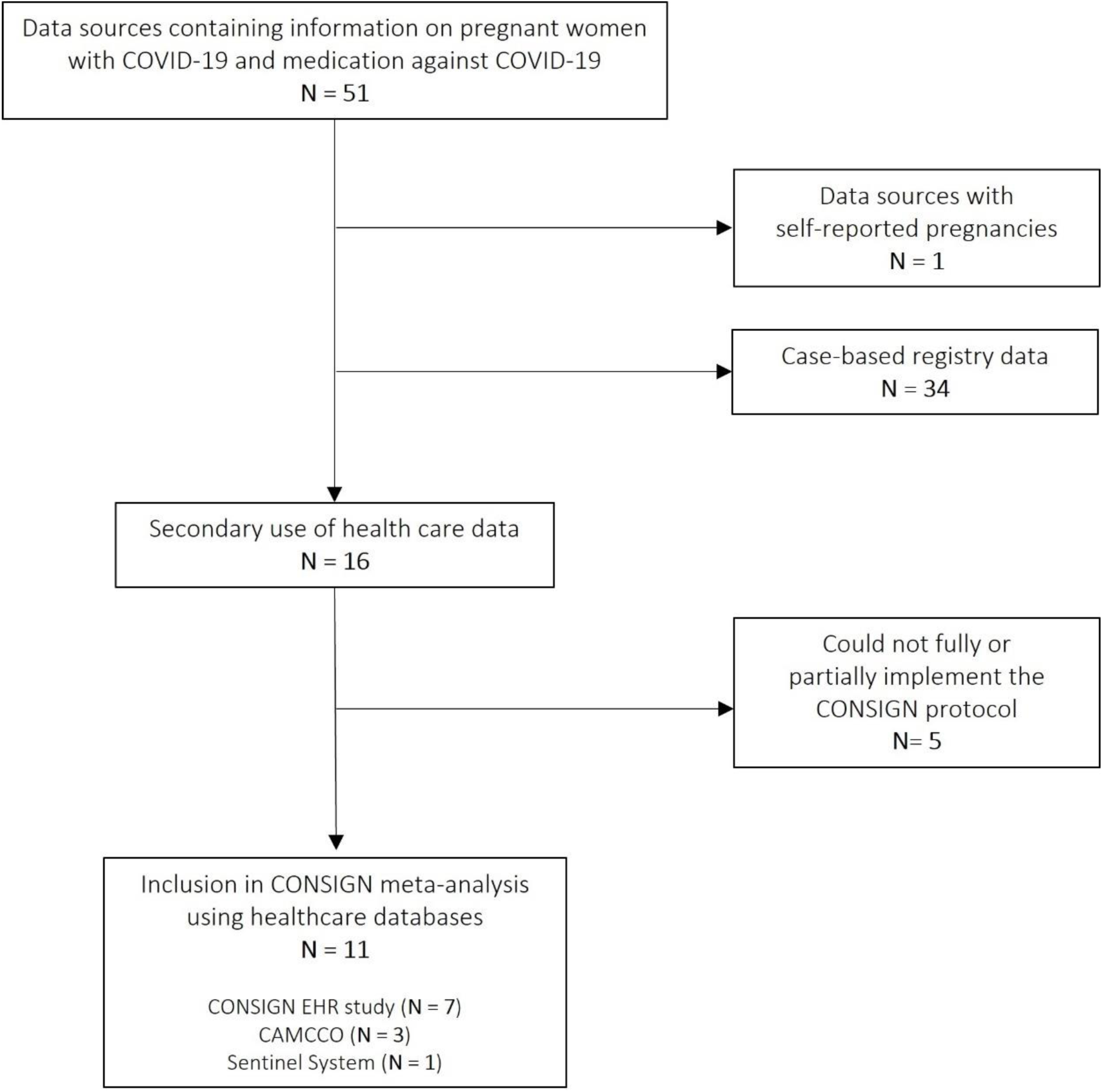
Flowchart of the selection process of networks and data sources participating in the meta-analysis using secondary healthcare data

The CONSIGN EHR study included data from seven electronic healthcare registries in six European countries (Tuscany, Italy; France; Valencia, Spain; Aragon, Spain; Wales, UK; Norway; Sweden). The data sources included general practice (primary care) databases and record linkage of demographic data, registers covering primary and secondary care, and prescribing or dispensing registers (Table S2). The data were analysed in a distributed manner using a common data model (CDM) and common analytics developed by the IMI-funded ConcePTION project.^5,6,10^ The Canadian Mother-Child Cohort (CAMCCO) Active Surveillance Initiative included data from three provinces, Alberta, Manitoba, and Ontario, and their results are presented separately. Data came from medical services, prescription drugs, hospitalization archives, and COVID-19 testing program results (Table S2). CAMCCO aims to study the impact of prescription drug use during pregnancy and short and long-term health outcomes in the mother and child. CAMCCO has developed standardized and harmonized tools based on the Quebec Pregnancy/Child Cohort (QPC).^19,20^

The U.S. FDA Sentinel System included the most recently refreshed data from seven Data Partners (DP) participating in the Rapid COVID-19 Sentinel Distributed Database. The database included four national claims-insurers and three DP with regional coverage in Colorado, Oregon, Minnesota, and Washington States (Table S2). Sentinel wrote its own study protocol focusing on COVID-19 in pregnancy and implemented the key aims and methods of the CONSIGN EHR study protocol.^21^

### Study population

The study population comprised an index cohort of pregnant women with a recorded COVID-19 diagnosis during their pregnancies overlapping January 2020 and extending until the last available data from each included data source, ranging from December 2020 to December 2022 (Table S2). Two external comparator cohorts were used: i) pregnant women without COVID-19 and ii) non-pregnant women of reproductive age with COVID-19. In CONSIGN EHR and the Sentinel System, the external comparator cohorts were matched, when appropriate, to the index cohort using pregnancy trimester, calendar month of COVID-19 infection and maternal age group. When matching pregnant women with and without COVID-19, pregnant women without COVID-19 were assigned the index date as the date of the matched individual’s COVID-19 diagnosis. CAMCCO did not deploy matching.

### Variables

Data source-specific information to determine pregnancy start and end dates and COVID-19 diagnosis is provided in Table S3. Pregnant women were identified according to the three initiatives. Across all sites, the start of a pregnancy was defined as the estimated first day of the last menstrual period (LMP), while the end of a pregnancy was defined as the date of birth (or non-live births in sites collecting this information). The Sentinel System algorithm used to detect pregnancies was based solely on the identification of live births. The definitions of trimesters of pregnancy are slightly different among the three included networks (Table S4).

COVID-19 cases were identified through records of COVID-19 in surveillance systems, diagnostic codes in health care records, and/or laboratory results (Table S3). In all data sources, except for Bordeaux PharmacoEpi (BPE) using the “Système National des Données de Santé (SNDS)” data in France, the identification of confirmed COVID-19 diagnoses was possible through the use of polymerase chain reaction (PCR) or antigen tests. COVID-19 severity was categorized into two levels: non-hospitalized and hospitalized for COVID-19 event. Non-hospitalized women had a recording of a positive COVID-19 test or diagnosis, with no subsequent hospital admission with a recording (primary or secondary) of COVID-19 within a 4-week period. Sentinel used a slightly different definition for assessing COVID-19 severity since reliable information on admission reason was available (Table S4).

The medication groups considered of special relevance to COVID-19 were obtained from the World Health Organization (WHO) and National Institutes of Health (NIH) guidelines and included analgesics, anthelminthics, anti-inflammatory and antirheumatic products, antibacterials for systemic use, antigout preparations, antihypertensives, antimycobacterials, antimycotics for systematic use, antineoplastic agents, antivirals for systematic use, corticosteroids for systematic use, cough and cold preparations, drugs for obstructive airway diseases, drugs used in diabetes, immune sera and immunoglobulins, immunostimulants, immunosuppressants, nasal preparations, psychoanaleptics, and psycholeptics.^22,23^ The one-month prevalence of each medication was assessed, with the numerator counting whether the medication was prescribed or dispensed within the 30 day risk window using the Anatomical Therapeutic Classification (ATC) at level 2 (Table S5). The denominator was the number of women in the 30 days prior to the date of COVID-19 diagnosis and separately 30 days after the diagnosis. In nearly all participating data sources, medication use during inpatient treatment periods was not available (Table S3).

Covariates of interest are listed in Table S4 and included: maternal age, trimester of pregnancy, at risk medical conditions for severe COVID-19, risk conditions for obstetric complications and calendar month of COVID-19 diagnosis.

### Statistical Analysis

The protocol and Statistical Analysis Plan (SAP) are publicly available elsewhere.^14^ A common R-script based on the ConcePTION CDM structure was created for the data sources participating in CONSIGN EHR study, except at Karolinska Institutet in Sweden, where they programmed their own analysis in SAS according to the SAP. CAMCCO and Sentinel used the CONSIGN EHR study SAP and ran their own scripts in SAS. All data sources delivered aggregated results, including counts, proportions, and 95% confidence intervals (CI), in prespecified shell table formats.

The statistical analyses were undertaken with R software, version 4.2.2.^24^ A meta-analysis of proportions was conducted using the “metaprop” function, enabling the calculation of a combined effect estimate with corresponding 95% CI. The Generalized Linear Mixed Model (GLMM) method assumes a random effects model and was employed to account for potential heterogeneity among individual study estimates.^25^ Specifically, the PLOGIT function was used to estimate the logit-transformed proportions. The use of the PLOGIT function allows for a more accurate estimation of proportion, especially when dealing with small sample sizes and sparse data.^26,27^ Comparisons were made between prevalence of medication use within 30 days pre- and 30 days post-COVID-19 diagnosis in pregnant women with COVID-19 and in two comparator groups: pregnant women without COVID-19, and non-pregnant women with COVID-19. Analyses were stratified by pregnancy trimester when the COVID-19 infection occurred and were limited to non-hospitalized COVID-19 cases to avoid misclassification of exposure during hospitalization. Forest plots were used to show individual site estimates (with 95% CI), and a diamond to represent the pooled estimate (95% CI) for each outcome of interest. Statistical significance was assessed by examining the overlap of confidence intervals, rather than by applying a statistical test. We evaluated heterogeneity with the I^2^ statistics.

Some sites were not allowed to share non-zero event counts <10 or, <6 or <5. In those cases, we imputed one event to calculate the prevalence. Otherwise, all sites with <10 events would have been excluded from the analyses, which would have introduced a bias. Where zero events were observed at a site, the model included the prevalence of 0%.

## RESULTS

The study identified 11 data sources in eight countries comprising 59,037 pregnant women diagnosed with COVID-19 during their pregnancy. Most of the countries had pregnancy data during 2020 and 2021; the US had data until the end of 2022, while France and Sweden had data on pregnancies in 2020 only (Table S2). Of the 59,037 pregnant women, 50,335 (85.3%) were not hospitalized for COVID-19. France had access to inpatient data only and was excluded from the analyses. More than half of the recorded COVID-19 infections (50.6%) occurred during the third trimester of pregnancy. The distribution of COVID-19 infection across the pregnancy trimesters differed considerably between regions. Sweden and Manitoba, Canada had lower numbers of first trimester infections compared with other regions (Table 1).

**Table 1.** Description of the different pregnancy cohorts positive for COVID-19.

|  | Total number of pregnant women with COVID-19 | Total number of non-hospitalized pregnant women with COVID-19 | Non-hospitalized pregnant women with COVID-19 trimester 1 | Non-hospitalized pregnant women with COVID-19 in trimester 2 | Non-hospitalized pregnant women with COVID-19 in trimester 3 |
| --- | --- | --- | --- | --- | --- |
| Tuscany, Italy | 995 | 739 | 181 (24.5%) | 261 (35.3%) | 297 (40.2%) |
| France* | 1,069 | 0 | 0 (0%) | 0 (0%) | 0 (0%) |
| Valencia, Spain | 3,654 | 3,231 | 1,241 (38.4%) | 1,032 (31.9%) | 958 (29.7%) |
| Aragon, Spain | 951 | 754 | 201 (26.7%) | 272 (36.1%) | 281 (37.3%) |
| Wales, UK | 1,941 | 1,642 | 555 (33.8%) | 599 (36.5%) | 488 (29.7%) |
| Norway | 1,146 | 879 | 217 (24.7%) | 361 (41.1%) | 301 (34.2%) |
| Sweden | 5,030 | 4,199 | 259 (6.2%) | 1,455 (34.7%) | 2,485 (59.2%) |
| Alberta, Canada | 2,296 | 1,968 | 571 (29.0%) | 706 (35.9%) | 691 (35.1%) |
| Manitoba, Canada | 235 | 170 | 19 (11.2%) | 62 (36.5%) | 89 (52.4%) |
| Ontario, Canada | 933 | 933 | 270 (28.9%) | 303 (32.5%) | 360 (38.6%) |
| US | 40,787 | 35,820 | 6,726 (18.8%) | 9,562 (26.7%) | 19,532 (54.5%) |
| **Total** | **59,037** | **50,335** | **10,240 (20.3%)** | **14,613 (29.0%)** | **25,482 (50.6%)** |

\*COVID-19 cases in France were identified through hospital admissions records with a COVID-19 diagnosis. Consequently, pregnant women not requiring hospitalization are not included.

### Pre- and post-COVID-19 diagnosis medication use in pregnant women with COVID-19

Figure 2 displays the pooled prevalence of medication use in the 30 days pre- and post-COVID-19 diagnosis in non-hospitalized pregnant women with COVID-19 by medication group and pregnancy trimester of infection. All medication groups exhibited a low monthly prevalence across the different pregnancy trimesters, with values consistently below 7%. Notably, the pooled prevalence pre- and post-COVID-19 for anthelminthics, antigout preparations, antimycobacterials, antimycotics, antineoplastic agents, immune sera and immunoglobulins, immunostimulants and immunosuppressants was found to be <0.2% across all trimesters. Due to this extremely low prevalence, these medication groups are not depicted in Figure 2. The forest plots showing the pooled prevalence of all medication groups are available in Figures S1-S22. For nearly all medication groups and trimesters, the heterogeneity of the pooled prevalence pre- and post-COVID-19 diagnosis was high.

**Figure 2.**
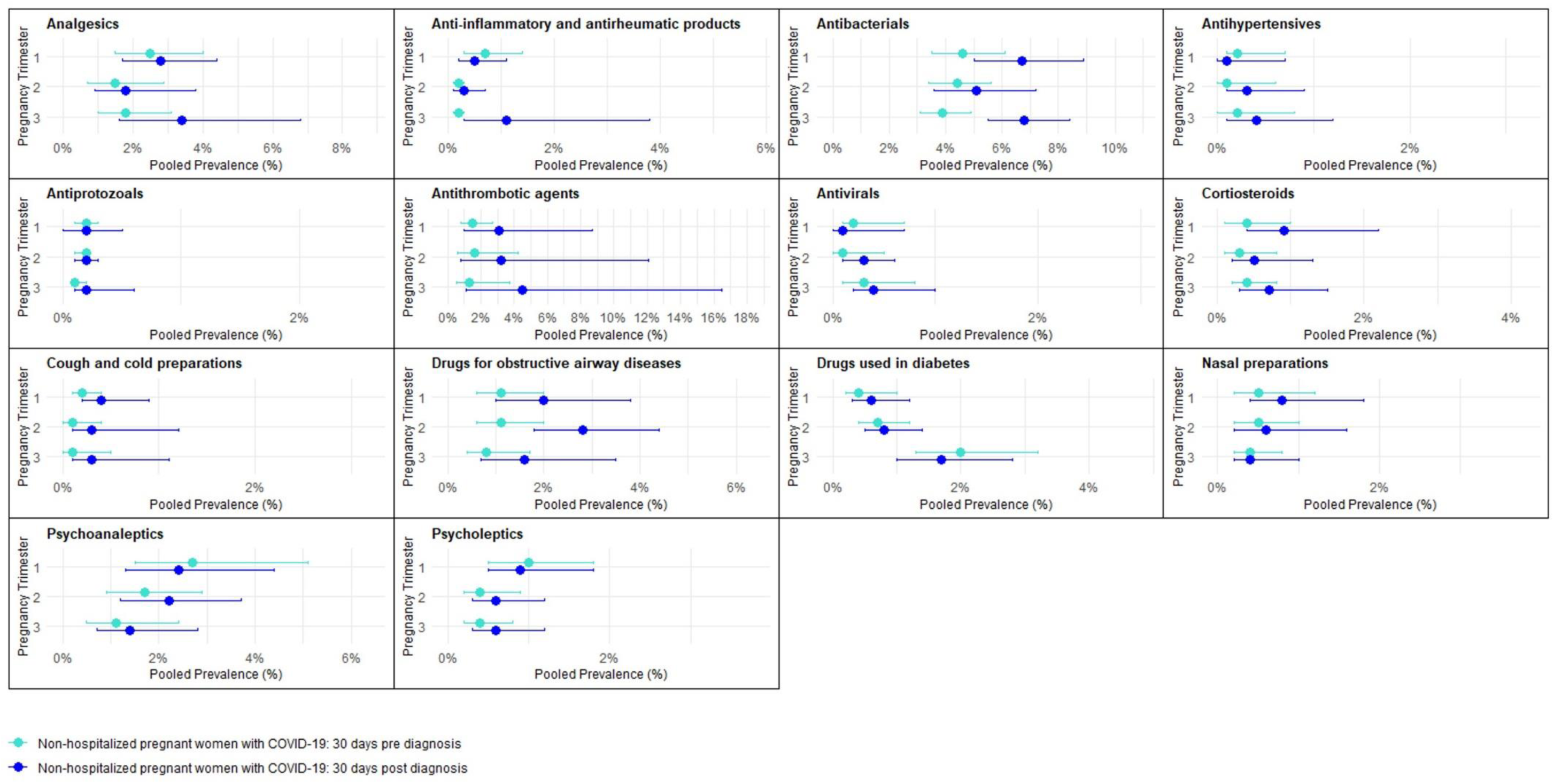
Pooled prevalence of medication use in the 30 days pre- and post-COVID-19 diagnosis in non-hospitalized pregnant women with COVID-19 by medication group and pregnancy trimester

Comparisons between the 30 days pre- and post-COVID-19 diagnosis periods showed a significant increase in the pooled prevalence of the use of antibacterials after COVID-19 diagnosis in the third pregnancy trimester (pre-COVID-19 3.9%, 95% CI 3.1 - 4.9, I^2^ = 89% versus post-COVID-19 6.8%, 95% CI 5.5 - 8.4, I^2^ = 94%). This increase of antibacterials in the month following COVID-19 diagnosis was also seen in the first and second pregnancy trimesters, although not statistically significant (Figure S4). With other medications, including analgesics, antithrombotic agents, corticosteroids, cough and cold preparations, and drugs for obstructive airway diseases, we also observed an increase in the pooled prevalence across all trimesters, but the confidence intervals of the prevalence pre- and post-COVID-19 diagnosis overlapped (Figure 2). The increase in antithrombotic agents following a COVID-19 diagnosis was most prominent in third trimester with a pooled prevalence of 1.3% (95% CI 0.5 - 3.7, I^2^ = 97%) pre-COVID-19 and 4.5% (95% CI 1.1 - 16.5, I^2^ = 100%) post-COVID-19, mostly due to the increase in dispensing in European sites (Figure S11).

Additionally, a non-statistically significant increase was observed for anti-inflammatory and antirheumatic products, antihypertensives, antivirals, psychoanaleptics, and psycholeptics in the second and third pregnancy trimesters, and for drugs used in diabetes and nasal preparations in the first and second pregnancy trimesters (Figure 2). The increase in anti-inflammatory and antirheumatic products prescription following a COVID-19 diagnosis was most prominent in the third pregnancy trimester (pre-COVID-19 0.2%, 95% CI 0.1 - 0.3, I^2^ = 8% versus post-COVID-19 1.1%, 95% CI 0.3 - 3.8, I^2^ = 98%), mainly attributed to increase in prevalence in US, Manitoba, Canada, and Spain (Figure S3).

### Medication use in pregnant women with COVID-19 and pregnant women without COVID-19

The baseline characteristics of the cohorts of pregnant women with and without COVID-19 across the study sites are presented in Table S6. In CONSIGN and Sentinel, the pregnant women were matched on pregnancy trimester, calendar month of COVID-19 infection and maternal age group. Therefore, the distribution of age between pregnant women with and without COVID-19 was comparable between groups, with most women aged 25-39 years. In CAMCCO, age distribution between the two groups was also comparable even though they did not apply matching. The level of co-morbidities differed across sites, with the highest prevalence of co-morbidity in Canada. The prevalence of obesity was highest in the US.

Figure 3 shows the pooled prevalence of medication use in the 30 days post-COVID-19 diagnosis in non-hospitalized pregnant women with COVID-19 and pregnant women without COVID-19 by medication group and pregnancy trimester. The corresponding forest plots showing the pooled prevalence of all medication groups are provided in Figures S1-S22 and indicated high heterogeneity in nearly all analyses. None of the pooled prevalence rates were significantly different between pregnant women with COVID-19 and those without COVID-19. However, pooled prevalence rates of the use of analgesics, anti-inflammatory and antirheumatic products, antibacterials, antithrombotic agents, corticosteroids, cough and cold preparations, drugs for obstructive airway diseases, drugs used in diabetes, nasal preparations, and psycholeptics were generally higher across all trimesters in pregnant women with COVID-19 compared with those without COVID-19 (Figure 3).

**Figure 3.**
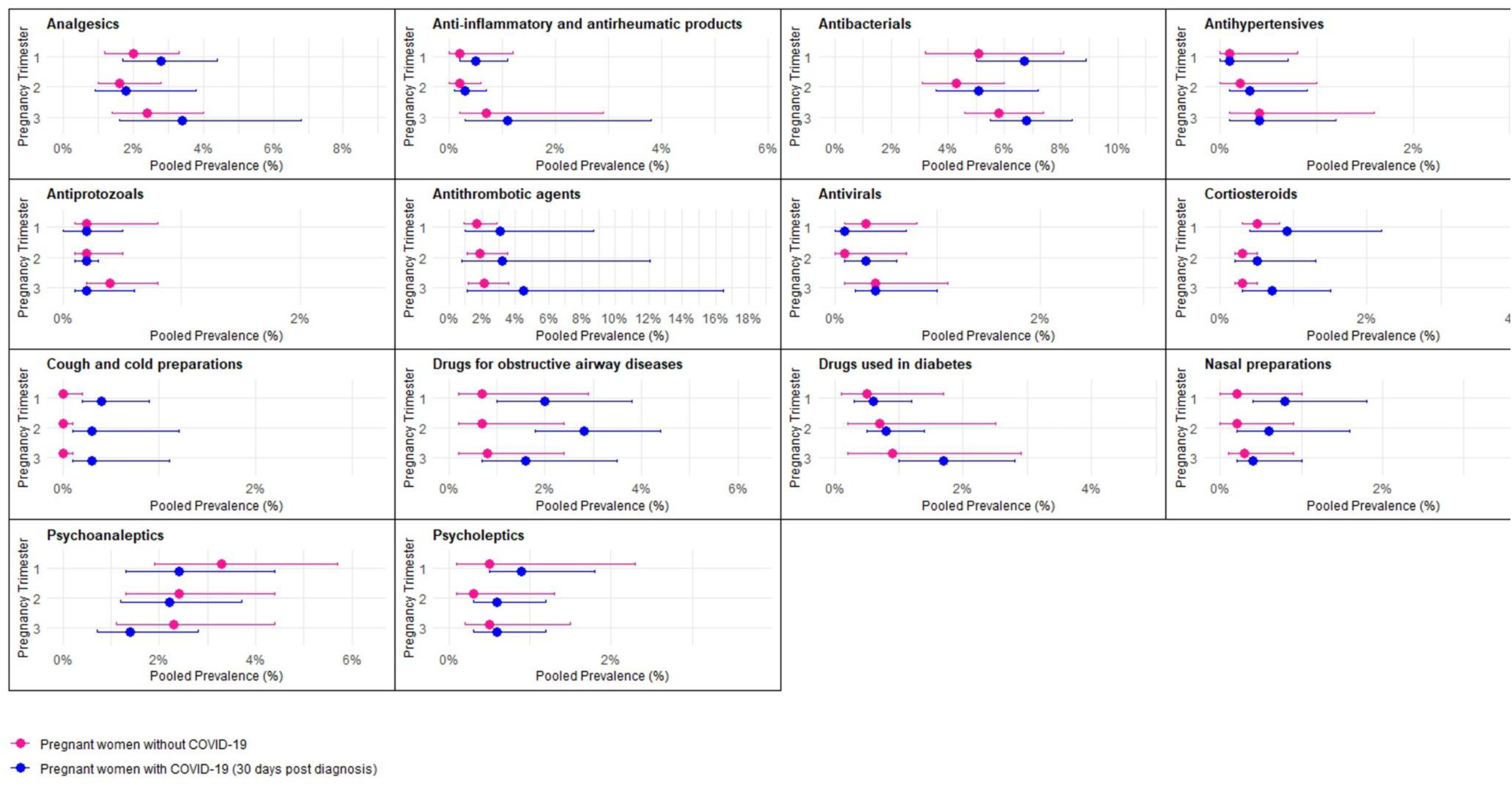
Pooled prevalence of medication use in the 30 days post COVID-19 diagnosis in non-hospitalized pregnant women with COVID-19 compared to pregnant women without COVID-19 by medication group and pregnancy trimester

The greatest difference in 30 days post-COVID-19 pooled prevalence of medication use between non-hospitalized pregnant women with and without COVID-19 was observed for antithrombotic agents. In the third trimester, the pooled prevalence of antithrombotic agents 30 days post-COVID-19 was 4.5% (95% CI 1.1 - 16.5, I^2^ = 100%) in pregnant women with COVID-19, compared to 2.1% (95% CI 1.2 - 3.6, I^2^ = 99%) in pregnant women without COVID-19 in the third trimester (Figure S11). The higher prescription post-COVID-19 diagnosis of drugs for obstructive airway diseases observed among pregnant women with COVID-19 than those without COVID-19 was most prominent in the second trimester (2.8%, 95% CI 1.8 - 4.4, I^2^ = 80% and 0.7%, 95% CI 0.2 - 2.4, I^2^ = 96%, respectively) (Figure S15). Regarding drugs used in diabetes, the difference was most prominent in the third trimester with a pooled prevalence of 1.7% (95% CI 1.0 - 2.8, I^2^ = 80%) in pregnant women with COVID-19 compared to 0.9% (95% CI 0.2 - 2.9, I^2^ = 99%) in those without COVID-19 (Figure S16). In contrast, the pooled prevalence of psychoanaleptics use was lower post-COVID-19 across all trimesters in pregnant women with COVID-19 than amongst those without COVID-19 (Figure S21).

### Medication use in pregnant women with COVID-19 and non-pregnant women with COVID-19

The baseline characteristics of the cohorts of pregnant women with COVID-19 and non-pregnant women with COVID-19 across the study sites are presented in Table S7. Wales, UK did not have adequate non-pregnant comparator group and was excluded from the analyses. In CONSIGN and Sentinel, the distribution of age between pregnant women with COVID-19 and non-pregnant women with COVID-19 was comparable due to matching on age, with most women aged 25-39 years. In CAMCCO, no matching was applied, and the age distribution differed slightly between cohorts, with higher proportions of women aged 15-24 years and 40-45 years in the non-pregnant group. The level of co-morbidities differed across sites, with the highest prevalence of any co-morbidities in both cohorts in Canada.

Figure 4 shows the pooled prevalence of medication use in the 30 days post-COVID-19 diagnosis in non-hospitalized pregnant women with COVID-19 compared to non-pregnant women with COVID-19 by medication group and pregnancy trimester. The corresponding forest plots showing the pooled prevalence of all medication groups are provided in Figures S23-S44 and indicated high heterogeneity in nearly all analyses. Comparisons between the 30 days post-COVID-19 prevalence rates in pregnant women with COVID-19 and non-pregnant women with COVID-19 showed a statistically significant lower pooled prevalence of the use of psychoanaleptics in pregnant women with COVID-19 in the third trimester (1.4%, 95% CI 0.8 - 2.5, I^2^ = 93%) compared to non-pregnant women with COVID-19 (4.8%, 95% CI 3.1 - 7.4, I^2^ = 99%). This difference was also seen in the first and second trimesters, although not statistically significant (Figure S43). The pooled prevalence rates of the use of analgesics, antiprotozoals, corticosteroids, and psycholeptics were also generally lower across all trimesters in pregnant women with COVID-19 compared with non-pregnant women with COVID-19, although confidence intervals overlapped (Figure 4). The lower prescription post-COVID-19 diagnosis of corticosteroids observed among pregnant women with COVID-19 than non-pregnant with COVID-19 was most prominent in the second trimester (0.5%, 95% CI 0.2 - 1.5, I^2^ = 89% and 2.1%, 95% CI 1.0 - 4.2, I^2^ = 100%, respectively) (Figure S35).

**Figure 4.**
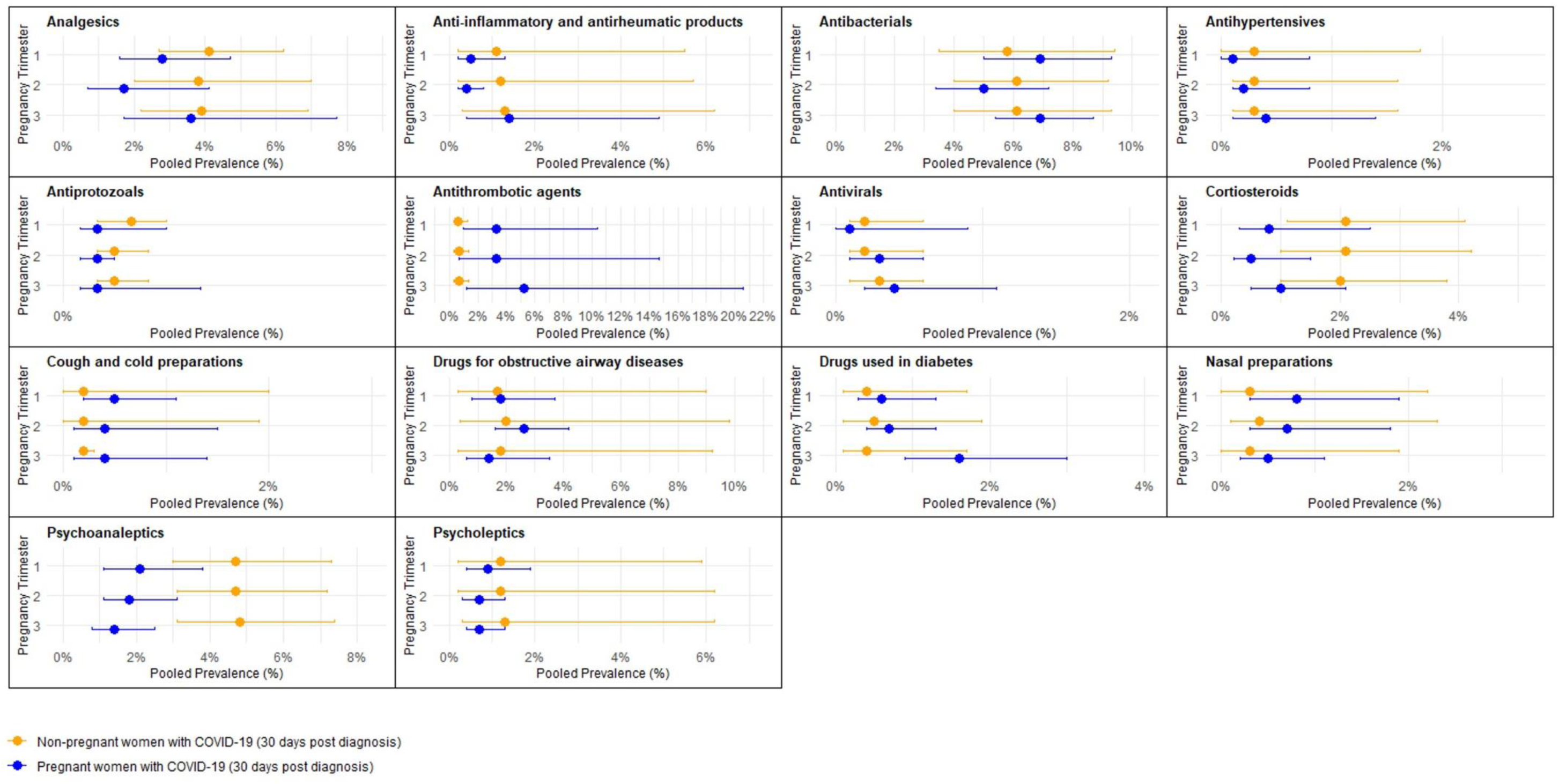
Pooled prevalence of medication use in the 30 days post COVID-19 diagnosis in non-hospitalized pregnant women with COVID-19 compared to non-pregnant women with COVID-19 by medication group and pregnancy trimester

In contrast, pooled prevalence rates of the use of antithrombotic agents, cough and cold preparations, drugs used in diabetes, and nasal preparations were generally higher across all trimesters in pregnant women with COVID-19 compared with non-pregnant women with COVID-19 (Figure 4). The greatest difference in 30 days post-COVID-19 pooled prevalence of medication use between non-hospitalized pregnant women with COVID-19 and non-pregnant women with COVID-19 was observed for antithrombotic agents. The 30 days post-COVID-19 pooled prevalence of antithrombotic agents was 5.2% (95% CI 1.2 - 20.6, I^2^ = 100%) in pregnant women with COVID-19 in third trimester compared with 0.7% (95% CI 0.3 - 1.4, I^2^ = 99%) in non-pregnant women with COVID-19 (Figure S33).

## DISCUSSION

This international prospective two-stage IPD meta-analysis, combining healthcare data from 10 data sources in seven countries, presents findings on medication utilization in a cohort of 50,335 pregnant women not hospitalized for their COVID-19 event. Comparisons were made between prevalence of medication use 30 days pre- and post-COVID-19 diagnosis in this cohort and in two comparator groups: pregnant women without COVID-19, and non-pregnant women with COVID-19.

Overall, the prevalence of medication use in pregnant women with COVID-19 who were not hospitalized was low and varied widely across data sources. Among non-hospitalized pregnant women with COVID-19, antibacterials, antithrombotic agents, and analgesics were the medications used most frequently, with antibacterials being statistically significantly more frequently dispensed medication group in the 30 days after COVID-19 diagnosis than 30 days prior. The prevalence of medication use among non-hospitalized pregnant women with COVID-19 was often higher than the prevalence among pregnant women without COVID-19, although none of these differences reached statistical significance.

For some medications, the pooled prevalence was lower among non-hospitalized pregnant women with COVID-19 compared with non-pregnant women with COVID-19, e.g., analgesics, antiprotozoals, corticosteroids, psychoanaleptics and psycholeptics. Conversely, antithrombotic agents appeared to be more prevalent, although not significantly, amongst non-hospitalized pregnant women with COVID-19 compared with non-pregnant women with COVID-19. This finding probably reflects various guidelines recommending thrombosis prophylaxis in pregnant women with COVID-19.^28^ Furthermore, in our meta-analysis, post-COVID-19 diagnosis, the prevalence of antithrombotic agents was notably higher in pregnant women with COVID-19 compared to those without COVID-19 across all trimesters. It has been demonstrated that pregnant women with COVID-19 are more at risk of developing venous thromboembolism than pregnant women without COVID-19.^29,30^ The post-COVID-19 prevalence of the use of antithrombotic agents in pregnant women with COVID-19 was highest in Spain, followed by Italy and Sweden. The prevalence was much lower in Canada, the US and Wales. This may be due to strong recommendations to prescribe low molecular weight heparin (LMWH) treatment in European guidelines, while guidelines in the US recommended LMWH only for some categories of patients, e.g., those with comorbidities.^9^ Corticosteroids were mostly prescribed to non-hospitalized pregnant women in Italy and the US, with national guidelines strongly recommending them from September 2020 and April 2020, respectively.^11^ Compared to non-pregnant women, corticosteroids were less often prescribed to pregnant women indicating that pregnant women may be undertreated, potentially due to concerns about foetal safety. The prescription rate of psychoanaleptics was markedly higher in Wales, than elsewhere in Europe, as previously reported.^31,32^ The higher prevalence of drugs used in diabetes in pregnant women with COVID-19 compared to non-pregnant women with COVID-19 probably reflects gestational diabetes in the pregnant cohort. Additionally, our meta-analysis highlights that anti-inflammatory and antirheumatic products, which includes NSAIDs, were prescribed in non-hospitalized pregnant women with and without COVID-19 across all trimesters of pregnancy, particularly in the US and Canada. However, NSAIDs are not recommended after 30 weeks of gestation due to their association with premature closure of the ductus arteriosus.^33,34^ Unfortunately, we could not identify the specific medication used, nor confirm whether these medications were prescribed for COVID-19 treatment or obstetric management.

A systematic review and meta-analysis of 62 case series and cohort studies published between December 2019 and February 2021 provided data on 31,016 pregnant women diagnosed with COVID-19.^4^ The review indicated that among the studies reporting on the pharmacologic management of COVID-19, approximately half of the pregnant women received antibiotics, anticoagulants, or hydroxychloroquine, while one in three were administered antivirals, and nearly one in five were managed with either corticosteroids or immunotherapy.^6^ It is important to note that a key distinction in the studies included in that meta-analysis was that most were conducted in a hospital setting, making direct comparisons with our study challenging. Additionally, results from the Observational Health Data Sciences and Informatics (OHDSI) network, which included electronic medical records and claims data from France, Spain and the USA and included 8,598 pregnant women with COVID-19, were focused on the inpatient settings.^35^ Data from the outpatient setting were not available, making comparison with our findings impossible. The distinction between in and outpatient is important, as was already shown by previous studies within CONSIGN from the COVI-PREG and INOSS initiatives.^7,9^ They showed that medication use is strongly associated with the severity of the disease, and therefore, the restriction to include non-hospitalized pregnant women with prescriptions in outpatient or primary care settings only is a key strength of our meta-analysis. Another CONSIGN meta-analysis is collecting information directly about medication use among hospitalized patients and should be referred to for that comparison.^14^

Several limitations need to be considered when interpreting the findings of this meta-analysis. Firstly, we observed high levels of heterogeneity among the data sources, and a possible explanation for this variability could be attributed to the differences in health systems, prescription status and reimbursement practices, and clinical guidelines across the countries included. These differences may have influenced the availability, accessibility, and utilization of medications among pregnant women with COVID-19, leading to the diverse prevalence rates observed. For instance, in Canada, the provinces Alberta and Manitoba have pharmaceutical claims for the total population whereas Ontario has very few people under 65 eligible for drug reimbursement through the provincial program. In addition, some of the highest prescription rates were observed in Wales, UK, where there are no charges for medicines at the point of need and collection, for example analgesics that may be bought cheaply if often prescribed. The data analysed from the Sentinel System included primarily a commercially insured population. Therefore, publicly insured persons may be underrepresented, and uninsured persons are not represented. The impact of different healthcare systems on medication usage patterns is a crucial factor to consider when interpreting findings. We were unable to adjust for covariates such as socio-economic status, substance misuse, and smoking, which were captured in some data sources but not in others. Further research and comparative studies exploring the influence of health system structures on medication utilization during pregnancy and COVID-19 infection will be valuable for a comprehensive understanding of these observed heterogeneities.

Another potential explanation for the observed heterogeneity in our analyses could be the variation in data collection methods and alignment challenges observed during the harmonization process. To ensure comparability between the CONSIGN EHR study, CAMCCO and the Sentinel System, we made significant attempts to share shell tables, definitions, and other relevant information. However, despite our best efforts, there were instances where complete alignment proved challenging. For example, CAMCCO did not apply matching, however, we do not anticipate this to have significantly influenced the results, given the comparability of baseline characteristics. Additionally, there were differences in the definition of trimester of pregnancy and age groups. Differences in data collection methods, varying healthcare systems, and distinct approaches to defining certain variables posed obstacles to achieving perfect congruence. Nonetheless, we rigorously pursued harmonization as far as possible. Furthermore, it is important to note that the methods used for meta-analysis of proportions typically result in a large I², caused by the nature of proportional data. Therefore, high I² in the context of meta-analysis of proportions does not necessarily mean that the data are inconsistent.^36^

Secondly, due to the stratification of results by trimester and the limitation to non-hospitalized pregnant women only, the small sample sizes in some data sources resulted in less precise prevalence estimates, as evidenced by wide confidence intervals. When comparing across data sources, it is essential to consider this. Although not planned in the SAP, an analysis combining all trimesters together to investigate the influence of COVID-19 may be useful.

Thirdly, the Sentinel System identified only pregnancies resulting in live births, which may favour low-risk pregnancies. The pregnancies potentially at risk of miscarriage could have had different medication use patterns during the first trimester, that may have been detected in the other countries. However, it is worth noting that Sentinel is the largest dataset, so excluding it would inevitably reduce the sample size. Lastly, due to the rule of reporting one event for data sources that were not allowed to share very low numbers, we may sometimes have underestimated the prevalence.

## CONCLUSION

The findings of this international prospective meta-analysis, which includes 50,335 non-hospitalized pregnant women with COVID-19 from seven countries and covers a considerable period spanning from January 2020 to December 2022, offers valuable insights into medication utilization patterns. The data provides rich information on the similarities and disparities across different countries. This information contributes significantly to a comprehensive understanding of COVID-19 management strategies in non-hospitalized pregnant women, informing evidence-based decision-making in diverse healthcare settings.

## ACKNOWLEGDEMENTS

We would like to acknowledge Prof. Dr. Olaf Klungel from Utrecht University for his leadership within the EU PE&PV Research Network. We extend our gratitude to our colleagues at the Real-World Evidence department of UMCU Utrecht for their assistance in generating R scripts, conducting quality checks, and developing codelists. Additionally, we express our appreciation to RTI Health Solutions, particularly Andrea Margulis and Elena Rivero, for their support and feedback throughout the development of this manuscript.

## DATA AVAILABILITY STATEMENT

The aggregated results from the individual data sources that support the findings of this study are available on request from the corresponding author. The individual level data in each data source are not publicly available due to privacy, governance or ethical restrictions.

## AUTHOR CONTRIBUTION STATEMENT

The CONSIGN core research team (OB, EM, EH, HMEN, EA, FA, GF, AP, KWMB, KP, CV, SJS, and MCJM) drafted the protocol and SAP. The protocol and SAP were revised and finalized based on feedback from all co-authors. OB and EM coordinated contact with all data access providers. The following authors had the main responsibility for local data analysis: Tuscany, Italy (RG, OP), France (RL, MAB), Valencia, Spain (CLRB, FSS), Aragon, Spain (BPP), Wales, UK (SJ, DT), Norway (HMEN), Sweden (CEC, DH), Canada (AB, OS, PK), US (MUS, AC, JGL, EMJ, MEK, ST, WH, JJHM, LS). OB developed the R-scripts for meta-analysis, created the figures and tables for all the results. All authors participated in interpreting the data, reviewing the manuscript, and approved the final version. Each author accepts accountability for their part of the paper as published.

