## Supplementary material for "Medications used among Non-Hospitalized Pregnant Women with COVID-19: a Prospective Individual Patient Data Meta-analysis in Europe and North America": Table S1

*Collaborators of CONSIGN are listed in Table S1

Table S1. Members of the CONSIGN collaboration group

| Organisation | Name | Affiliation | ORCID ID |
|---|---|---|---|
| University of Oslo (CONSIGN WP1) | Benjamin P. Geisler | Pharmacoepidemiology and Drug Safety Research Group, Department of Pharmacy, University Oslo (UiO), Oslo, Norway | 0000-0003-1704-6067 |
| CAMCCO | Mark Walker | University of Ottawa, Ottawa, Ontario, Canada | 0000-0001-8974-4548 |
| CAMCCO | Steven Hawken | University of Ottawa, Ottawa, Ontario, Canada | 0000-0002-3341-9022 |
| CAMCCO | Sasha Bernatsky | Department of Epidemiology, Biostatistics and Occupational Health, McGill University, Montreal, Quebec, Canada | 0000-0002-9515-2802 |
| CAMCCO | Sherif Eltonsy | University of Manitoba, Winnipeg, Manitoba, Canada | 0000-0002-0520-5406 |
| Sentinel System | Emma Hoffman | Department of Population Medicine, Harvard Pilgrim Health Care Institute, Boston, Massachusetts, USA | N.A. |
| Sentinel System | Andrew B. Petrone | Department of Population Medicine, Harvard Pilgrim Health Care Institute, Boston, Massachusetts, USA | 0000-0001-8413-6236 |
| Sentinel System | Jolene Mosley | Department of Population Medicine, Harvard Pilgrim Health Care Institute, Boston, Massachusetts, USA | N.A. |
| Sentinel System | Jenice Ko | Department of Population Medicine, Harvard Pilgrim Health Care Institute, Boston, Massachusetts, USA | N.A. |
| ARS Toscana | Claudia Bartolini | Tuscan Regional Healthcare Agency, Florence, Italy | 0000-0001-8630-4598 |
| ARS Toscana | Giuseppe Roberto | Tuscan Regional Healthcare Agency, Florence, Italy | N.A. |
| ARS Toscana | Giorgio Limoncella | Tuscan Regional Healthcare Agency, Florence, Italy | N.A. |
| ARS Toscana | Anna Girardi | Tuscan Regional Healthcare Agency, Florence, Italy | N.A. |
| ARS Toscana | Giulia Hyeraci | Tuscan Regional Healthcare Agency, Florence, Italy | 0000-0002-6536-2083 |
| IACS | Antonio Gimeno-Miguel | EpiChron Research Group, Aragon Health Sciences Institute (IACS), IIS Aragón, Miguel Servet University Hospital, Zaragoza, Spain | 0000-0002-5440-1710 |
| IACS | Jonás Carmona-Pírez | EpiChron Research Group, Aragon Health Sciences Institute (IACS), IIS Aragón, Miguel Servet University Hospital, Zaragoza, Spain | 0000-0002-6268-8803 |
| IACS | Antonio Poncel-Falcó | EpiChron Research Group, Aragon Health Sciences Institute (IACS), IIS Aragón, Miguel Servet University Hospital, Zaragoza, Spain | N.A. |
| IACS | Aida Moreno-Juste | EpiChron Research Group, Aragon Health Sciences Institute (IACS), IIS Aragón, Miguel Servet University Hospital, Zaragoza, Spain | 0000-0002-8819-3278 |
| IACS | Alexandra Prados-Torres | EpiChron Research Group, Aragon Health Sciences Institute (IACS), IIS Aragón, Miguel Servet University Hospital, Zaragoza, Spain | 0000-0002-5704-6056 |
| SWANSEA | Ian Farr | Faculty of Medicine, Health and Life Science, Swansea University, Wales, UK | N.A. |
| SWANSEA | Saira Ahmed | Faculty of Medicine, Health and Life Science, Swansea University, Wales, UK | N.A. |
| SWANSEA | Ieuan Scanlon | Faculty of Medicine, Health and Life Science, Swansea University, Wales, UK | N.A. |

Table S1 continued. Members of the CONSIGN collaboration group

| Organisation | Name | Affiliation | ORCID ID |
|---|---|---|---|
| FISABIO-HSRU | Gabriel Sanfélix-Gimeno | Health Services Research and Pharmacoepidemiology Unit, Foundation for the Promotion of Health and Biomedical Research of Valencia Region, Valencia, Spain | 0000-0001-7098-4576 |
| FISABIO-HSRU | Isabel Hurtado | Health Services Research and Pharmacoepidemiology Unit, Foundation for the Promotion of Health and Biomedical Research of Valencia Region, Valencia, Spain | 0000-0002-8475-8112 |
| FISABIO-HSRU | Anibal Garcia-Sempere | Health Services Research and Pharmacoepidemiology Unit, Foundation for the Promotion of Health and Biomedical Research of Valencia Region, Valencia, Spain | N.A. |
| FISABIO-HSRU | Salvador Peiro | Health Services Research and Pharmacoepidemiology Unit, Foundation for the Promotion of Health and Biomedical Research of Valencia Region, Valencia, Spain | 0000-0002-3902-569X |
| BPE | Jérémy Jové | Bordeaux PharmacoEpi, INSERM CIC-P1401, Université de Bordeaux, Bordeaux, France | N.A. |
| BPE | Dunia Sakr | Bordeaux PharmacoEpi, INSERM CIC-P1401, Université de Bordeaux, Bordeaux, France | N.A. |
| BPE | Cécile Droz-Perroteau | Bordeaux PharmacoEpi, INSERM CIC-P1401, Université de Bordeaux, Bordeaux, France | 0000-0002-7697-1167 |
| COVI-PREG (CONSIGN WP2) | David Baum | Materno-Fetal and Obstetrics Research Unit, Woman-Mother-Child Department, Lausanne University Hospital, Lausanne, Switzerland | N.A. |
| INOSS (CONSIGN WP3) | Hilde M. Engjom | Department for Health Promotion and Department for Health Registry Research and Development, Norwegian Institute of Public Health, Bergen, Norway | 0000-0003-1582-4283 |
| Vall d'Hebron & UMC Utrecht | Riera-Arnau | Department of Clinical Pharmacology, Vall d'Hebron Hospital Universitari, Vall Hebron Institut de Recerca Barcelona, Spain & Department of Data Science & Biostatistics, Julius Global Health, University Medical Center Utrecht (UMCU), Utrecht, the Netherlands | 0000-0001-7591-0218 |
| Vall d'Hebron | Mònica Sabaté Gallego | Department of Clinical Pharmacology, Vall d'Hebron Hospital Universitari, Vall Hebron Institut de Recerca Barcelona, Spain | 0000-0001-6206-1085 |
| Vall d'Hebron | Elena Ballarín Alins | Department of Clinical Pharmacology, Vall d'Hebron Hospital Universitari, Vall Hebron Institut de Recerca Barcelona, Spain | 0000-0001-9786-6617 |
| Vall d'Hebron | Cristina Aguilera Martin | Department of Clinical Pharmacology, Vall d'Hebron Hospital Universitari, Vall Hebron Institut de Recerca Barcelona, Spain | 0000-0002-7985-7327 |
| Health Canada | Melissa Kampman | Data Analytics and Real-world Evidence Division, Health Products and Food Branch, Health Canada, Ottawa, Ontario, Canada | N.A. |
| Health Canada | Celline Brasil | Data Analytics and Real-world Evidence Division, Health Products and Food Branch, Health Canada, Ottawa, Ontario, Canada | N.A. |

N.A. = not available.

Table S2. Description of the data sources included in the meta-analysis

| Data access provider (data source) | Country (area) | Estimated births per year | Type of data source | Medical birth registry | Diagnosis | Data availability |
|---|---|---|---|---|---|---|
| CONSIGN EHR study | | | | | | |
| ARS Toscana (ARS database) | Italy (Tuscany) | 25 000 | Record linkage | Yes | In-hospital, emergency room | Mar 2020 – Dec 2021 |
| BPE (SNDS) | France (national) | 700 000 | Health insurance | No | In-hospital | Mar 2020 – Dec 2020 |
| FISABIO-HSRU (VID) | Spain (Valencia) | 32 000 | Record linkage | Yes | GP, In-hospital, Outpatient Specialists | Mar 2020 – Dec 2021 |
| IACS (PRECOVID study aNd EpiChron Cohort) | Spain (Aragon) | 10 000 | Record linkage | Yes | GP, In-hospital | Mar 2020 – Dec 2021 |
| SWANSEA (SAIL) | UK (Wales) | 33 000 | Record linkage | Yes | GP, In-hospital | Mar 2020 – Dec 2021 |
| UiO (Linked national registries) | Norway (national) | 60 000 | Record linkage | Yes | GP, In-hospital | Mar 2020 – Dec 2021 |
| Karolinska Institutet (Linked national registers) | Sweden (national) | 100 000 | Record linkage | Yes | In-hospital, Outpatient Specialists | Mar 2020 – Dec 2020 |
| Total estimated births Europe: | | 960 000 births/year | | | | |
| CAMCCO | | | | | | |
| Alberta (Linked databases) | Canada (Alberta) | 49 000 | Record linkage | Yes | GP, In-hospital, Outpatient Specialists, COVID-19 database | Mar 2020 - Aug 2021 |
| Manitoba (Linked databases) | Canada (Manitoba) | 16 000 | Record linkage | Yes | GP, In-hospital, Outpatient Specialists, COVID-19 database | Mar 2020 - Feb 2021 |
| Ontario (Linked databases) | Canada (Ontario) | 136 000 | Record linkage | Yes | GP, In-hospital, Outpatient Specialists, COVID-19 database | Mar 2020 - June 2021 |
| Total estimated births in Canada: | | 201 000 births/year | | | | |
| Sentinel System | | | | | | |
| Sentinel System (Sentinel Distributed Database) | US (national and regional coverage in Colorado, Oregon, Minnesota, and Washington States) | 485 000 | Health insurance | No | In-Hospital, Outpatient Specialists | Jan 2020 - Dec 2022 |
| Total estimated births in US: | | 485 000 births/year | | | | |
| Total estimated births: | | 1 685 000 births/year | | | | |

Abbreviations: ARS = Agenzia Regionale di Sanita' della Toscana; BPE = Bordeaux PharmacoEpi platform; CAMCCO = Canadian Mother-Child Cohort; EHR = electronic health record; FISABIO-HSRU = Foundation for the Promotion of Health and Biomedical Research of Valencia Region - Health Services Research Unit; GP: general practitioner, primary care; IACS = Instituto Aragones de Ciencias de la Salud; SAIL = Secure Anonymised Information Linkage; SNDS = Système National des Données de Santé; SWANSEA = Swansea University; UiO = University of Oslo; VID = Valencia Integrated Database.

Table S3. Data source specific methods for determining pregnancy start and end, COVID-19 diagnosis, and medication use

| DAP (data source) | Pregnancy start | Pregnancy end | COVID-19 diagnosis | Medications |
|---|---|---|---|---|
| CONSIGN EHR study | | | | |
| ARS Toscana (ARS database) | Date of record minus gestational age OR estimated on diagnostic code or period of pregnancy when a procedure is first expected | Date of record OR estimated from due date based on estimation of start date | Registry of positive COVID-19 tests (official surveillance system) | Outpatient dispensing |
| BPE (SNDS) | Date estimated from pregnancy algorithm which includes LMP and gestational age | Date of delivery | Inpatient data (PMSI) with ICD10 codes for COVID-19 diagnoses; no laboratory positive test result available. | Out + inpatient dispensing |
| FISABIO-HSRU (VID) | Date of delivery/ end of pregnancy minus gestational age (based on ultrasound/LMP) (95% pregnancies) OR Date of delivery minus 40 weeks when new-borns weight is 2500 gr or above and through a linear model when weight <2500 gr | Date of delivery or abortion | All PCR or antigen test results are recorded in RedMIVA (Microbiological Surveillance Network of the Valencian Community) | Outpatient dispensing |
| IACS (PRECOVID study aNd EpiChron Cohort) | Date of the LMP | Date of the pregnancy delivery or abortion recorded in primary care or hospital discharge | Registry developed for monitoring the evolution of COVID-19 disease in the region, includes all PCR or antigen test results | Outpatient dispensing |
| SWANSEA (SAIL) | Date of delivery (Monday of the week of birth as part of the irrevocable anonymisation process) minus gestational age at birth | Monday of the infant's week of delivery (to avoid use of identifiable data) | COVID-19 test results dataset available from healthdatagateay,org which include all test results and symptom trackers | Primary care |
| UiO (Linked national registries) | Date of delivery minus gestational length in days based on ultrasound or LMP | Date of delivery (all pregnancies > gestational week 12, live or non-live) | Laboratory confirmed positive test recorded in MSIS (Norwegian surveillance system for communicable diseases) | Outpatient dispensing |
| Karolinska Institutet (Linked national registries) | Date of delivery minus gestational length in days based on ultrasound or LMP | Date of delivery | All positive PCR test results registered in SmiNet (The Infectious Disease Register) and ICD-10 diagnosis for COVID-19 (National Patient register) | Outpatient dispensing |
| CAMCCO | | | | |
| Alberta (Linked databases) | First day of LMP (for deliveries) using gestational age reported in the delivery hospitalization chart summary AND algorithm to estimate first day of LMP (for spontaneous and planned abortions) | Clinically detected spontaneous or induced/planned abortion or delivery | PCR test | Outpatient dispensing |
| Manitoba (Linked databases) | First day of LMP (for deliveries) using gestational age reported in the delivery hospitalization chart summary AND algorithm to estimate first day of LMP (for spontaneous and planned abortions) | Clinically detected spontaneous or induced/planned abortion; or delivery | PCR test | Outpatient dispensing |
| Ontario (Linked databases) | First day of LMP (for deliveries) using gestational age reported in the delivery hospitalization chart summary AND algorithm to estimate first day of LMP (for spontaneous and planned abortions) | Clinically detected spontaneous or induced/planned abortion; or delivery | PCR test | Outpatient dispensing and some in-patient collected at delivery |
| Sentinel System | | | | |
| Sentinel System (Sentinel Distributed Database) | Estimated based on date of live-birth delivery and gestational age codes surrounding delivery date | ICD-10 diagnosis code indicating live-birth delivery | ICD-10 diagnosis code for COVID-19 and/or positive COVID-19 NAAT test | Outpatient dispensing |

Abbreviations: ARS = Agenzia Regionale di Sanita' della Toscana; BPE = Bordeaux PharmacoEpi platform; CAMCCO = Canadian Mother-Child Cohort; EHR = electronic health record; FISABIO-HSRU = Foundation for the Promotion of Health and Biomedical Research of Valencia Region - Health Services Research Unit; IACS = Instituto Aragones de Ciencias de la Salud; LMP = last menstrual period; NAAT = Nucleic Acid Amplification Tests; PCR = polymerase chain reaction; SAIL = Secure Anonymised Information Linkage; SNDS = Système National des Données de Santé; SWANSEA = Swansea University; UiO = University of Oslo; VID = Valencia Integrated Database.

**Table S4.** Covariates of interest

| Covariate | Definition | Identification of covariate |
|---|---|---|
| Age of mother | Categorized into three groups: 12-24 years of age, 25-39 years of age, 40-55 years of age in CONSIGN EHR study and Sentinel. In CAMCCO, the age groups are 15-24 years of age, 25-39 years of age, and 40-45 years of age. | Date of birth |
| Trimester of pregnancy | CONSIGN EHR: The ACOG definition of timing in pregnancy is used:<br>1. Trimester 1: from Last Menstrual Period (LMP) to day 97 after LMP; or end of pregnancy, whichever earlier<br>2. Trimester 2: from day 98 after LMP to day 195 after LMP; or end of pregnancy, whichever earlier<br>3. Trimester 3: from day 196 after LMP onwards until end of pregnancy<br>For Sentinel:<br>1. First trimester: days 0 to 90 of gestation (13 weeks)<br>2. Second trimester: days 91 to 180 (13+1 to 25+5 weeks)<br>3. Third trimester: days 181 (≥25+6 weeks through the day of the hospital admission for live-birth delivery.<br>For CAMCCO:<br>1. 1st trimester: 0-98 days (> 0 to ≥14 weeks gestation)<br>2. 2nd trimester: 99-182 days (>14 to ≥26 weeks' gestation)<br>3. 3rd trimester: ≥183 days (≥26+1 weeks' gestation to end of the pregnancy) | Pregnancy algorithm |
| Covid-19 positive test or diagnosis | Recording of a positive COVID-19 test (PCR or antigen test), or presence of a diagnostic code in health care records or mandatory notification to the surveillance system | Recording of "COVID-19" in registry data OR diagnostic codes AND/OR laboratory results |
| COVID-19 severity | Non-hospitalized women had a recording of a positive COVID-19 test or diagnosis, with no subsequent hospital admission with a recording (primary or secondary) of COVID-19 (within a 4-week period). Hospitalized women had any recording of 'COVID-19 positive test', or 'COVID-19 complication' in any of the diagnostic fields in hospital records, not just the principal diagnosis. However, if the COVID-19 test positive test was two days of delivery date/hospitalization for obstetric reasons, and no codes of severe symptoms (pneumonia, respiratory aid/use of ventilator) were found subsequently, these women were excluded from the severe COVID-19 group. This was not conducted in Sentinel, where severe COVID-19 was defined as hospitalized patients with COVID-19 complications, ICU admission, ventilation, or death; and non-severe COVID-19 as diagnosis of COVID-19 or positive SARS-COV-2 test. | |
| At-risk medical conditions for severe COVID-19 | At risk conditions for severe COVID-19 were divided into following subcategories:<br>- Cardiovascular disease/serious heart conditions include heart failure, coronary artery disease, cardiac myopathies<br>- Hypertension<br>- Sickle cell disease<br>- Chronic lung disease including COPD, cystic fibrosis, severe asthma, interstitial lung disease, pulmonary hypertension, bronchiectasis<br>- Type 1 & 2 Diabetes<br>- Obesity diagnosis or having a BMI ≥30 kg/m2<br>- Chronic kidney disease<br>- Chronic liver disease diagnosis (cirrhosis, non-alcoholic fatty liver disease, alcoholic liver disease, autoimmune hepatitis)<br>- HIV<br>- Common rheumatic diseases<br>- Immunosuppression & solid organ transplants<br>- Cancer<br>- Mental health disease (depression, dementia, and schizophrenia spectrum disorders) | Recording of the at-risk medical conditions for severe COVID-19 OR diagnostic codes AND medicines proxies |
| Risk conditions for obstetric complications | Prior maternal history of gestational diabetes or pre-eclampsia and prior history of stillbirth or late miscarriage, small for gestational age (SGA) child or foetal growth restriction (FGR), or child with congenital anomaly. | Recording of risk conditions for obstetric complications OR diagnostic codes |
| Calendar month of COVID-19 diagnosis | December 2019 till [according to each data source] | Date of diagnosis/positive test |

Abbreviations: ACOG = American College of Obstetricians and Gynaecologists; BMI = body mass index; CAMCCO = Canadian Mother-Child Cohort; EHR = electronic health record; LMP = last menstrual period; PCR = polymerase chain reaction.

Table S5. Medication groups of special relevance to COVID-19 at ATC level 2

| Name | Anatomical Therapeutic Classification (ATC) level 2 |
|---|---|
| Analgesics | N02 |
| Anthelminthics | P02 |
| Anti-inflammatory and antirheumatic products | M01 |
| Antibacterials for systemic use | J01 |
| Antigout preparations | M04 |
| Antihypertensives | C02, C03, C04, C07, C08 and/or C09 |
| Antimycobacterials | J04 |
| Antimycotics for systemic use | J02 |
| Antineoplastic agents | L01 |
| Antiprotozoals | P01 |
| Antithrombotic agents | B01 |
| Antivirals for systemic use | J05 |
| Corticosteroids for systemic use | H02 |
| Cough and cold preparations | R05 |
| Drugs for obstructive airway diseases | R03 |
| Drugs used in diabetes | A10 |
| Immune sera and immunoglobulins | J06 |
| Immunostimulants | L03 |
| Immunosuppressants | L04 |
| Nasal preparations | R01 |
| Psychoanaleptics | N06 |
| Psycholeptics | N05 |

Table S6. Baseline characteristics of pregnant women with and without COVID-19

| Study site | Trimester at COVID-19 infection | COVID-19 severity | Age | | Co-morbidities[1] | | Obstetric risk[2] | |
|---|---|---|---|---|---|---|---|---|
| | Pregnant with COVID-19 | Pregnant with COVID-19[3] | Pregnant with COVID-19 | Pregnant without COVID-19 | Pregnant with COVID-19 | Pregnant without COVID-19 | Pregnant with COVID-19 | Pregnant without COVID-19 |
| Tuscany, Italy | 1st trimester: 20.4%<br>2nd trimester: 27.8%<br>3rd trimester: 51.8% | Hospitalized: 13.3%<br>Non-hospitalized: 74.2% | 12-24 years: 11.1%<br>25-39 years: 81.6%<br>40-55 years: 7.3% | 12-24 years: 11.1%<br>25-39 years: 81.6%<br>40-55 years: 7.3% | Any: 20.8%<br>Cardiovascular: 4.2%<br>Chronic lung: 4.6%<br>Severe obesity: 0.2% | Any: 22.1%<br>Cardiovascular: 4.2%<br>Chronic lung: 5.7%<br>Severe obesity: 0.2% | Any: 8.4%<br>Adverse pregnancy outcomes: 7.0% | Any: 6.4%<br>Adverse pregnancy outcomes: 5.2% |
| Valencia, Spain | 1st trimester: 34.8%<br>2nd trimester: 28.7%<br>3rd trimester: 36.5% | Hospitalized: 0.9%<br>Non-hospitalized: 88.4% | 12-24 years: 13.2%<br>25-39 years: 76.9%<br>40-55 years: 9.9% | 12-24 years: 13.2%<br>25-39 years: 76.9%<br>40-55 years: 9.9% | Any: 23.6%<br>Cardiovascular: 5.9%<br>Chronic lung: 9.9%<br>Severe obesity: 1.7% | Any: 21.5%<br>Cardiovascular: 4.4%<br>Chronic lung: 9.0%<br>Severe obesity: 1.4% | Any: 12.0%<br>Adverse pregnancy outcomes: 9.6% | Any: 7.1%<br>Adverse pregnancy outcomes: 5.5% |
| Aragon, Spain | 1st trimester: 20.6%<br>2nd trimester: 29.0%<br>3rd trimester: 50.3% | Hospitalized: 2.6%<br>Non-hospitalized: 78.3% | 12-24 years: 13.9%<br>25-39 years: 78.3%<br>40-55 years: 7.8% | 12-24 years: 13.9%<br>25-39 years: 78.3%<br>40-55 years: 7.8% | Any: 24.6%<br>Cardiovascular: 4.4%<br>Chronic lung: 7.5%<br>Severe obesity: 0.4% | Any: 24.4%<br>Cardiovascular: 3.3%<br>Chronic lung: 8.3%<br>Severe obesity: 0.6% | Any: 10.4%<br>Adverse pregnancy outcomes: 8.5% | Any: 9.1%<br>Adverse pregnancy outcomes: 7.0% |
| Wales, UK | 1st trimester: 29.5%<br>2nd trimester: 32.6%<br>3rd trimester: 37.8% | Hospitalized: 10.3%<br>Non-hospitalized: 84.6% | 12-24 years: N.A.<br>25-39 years: N.A.<br>40-55 years: N.A. | 12-24 years: N.A.<br>25-39 years: N.A.<br>40-55 years: N.A. | Any: 29.8%<br>Cardiovascular: 1.2%<br>Chronic lung: 10.0%<br>Severe obesity: 1.2% | Any: 28.5%<br>Cardiovascular: 1.1%<br>Chronic lung: 9.5%<br>Severe obesity: 0.7% | Any: 19.2%<br>Adverse pregnancy outcomes: N.A. | Any: 17.4%<br>Adverse pregnancy outcomes: N.A. |
| Norway | 1st trimester: 20.5%<br>2nd trimester: 35.7%<br>3rd trimester: 43.8% | Hospitalized: 16.8%<br>Non-hospitalized: 76.7% | 12-24 years: 10.6%<br>25-39 years: 86.2%<br>40-55 years: 3.2% | 12-24 years: 10.6%<br>25-39 years: 86.2%<br>40-55 years: 3.2% | Any: 19.4%<br>Cardiovascular: 3.6%<br>Chronic lung: 4.5%<br>Severe obesity: 1.7% | Any: 21.5%<br>Cardiovascular: 2.7%<br>Chronic lung: 5.8%<br>Severe obesity: 1.3% | Any: 2.0%<br>Adverse pregnancy outcomes: 0.4% | Any: 1.7%<br>Adverse pregnancy outcomes: 0.3% |
| Sweden | 1st trimester: 5.9%<br>2nd trimester: 34.6%<br>3rd trimester: 59.4% | Hospitalized: 16.7%<br>Non-hospitalized: 83.3% | 12-24 years: 6.2%<br>25-39 years: 88.5%<br>40-55 years: 5.3% | 12-24 years: 6.2%<br>25-39 years: 88.5%<br>40-55 years: 5.3% | Any: 6.2%<br>Cardiovascular: 0.4%<br>Chronic lung: 0.8%<br>Severe obesity: 3.7% | Any: 5.5%<br>Cardiovascular: 0.6%<br>Chronic lung: 0.7%<br>Severe obesity: 3.0% | Any: N.A.<br>Adverse pregnancy outcomes: N.A. | Any: N.A.<br>Adverse pregnancy outcomes: N.A. |
| Alberta, Canada | 1st trimester: 25.3%<br>2nd trimester: 32.3%<br>3rd trimester: 42.5% | Hospitalized: 14.3%<br>Non-hospitalized: 85.7% | 15-24 years: 15.0%<br>25-39 years: 81.7%<br>40-45 years: 3.3% | 12-24 years: 13.7%<br>25-39 years: 82.9%<br>40-55 years: 3.5% | Any: 41.9%<br>Cardiovascular: 2.5%<br>Respiratory: 7.6%<br>Severe obesity: 1.8% | Any: 43.0%<br>Cardiovascular: 2.7%<br>Respiratory: 9.0%<br>Severe obesity: 1.4% | Any: N.A.<br>Adverse reproductive history outcomes: 5.4% | Any: N.A.<br>Adverse reproductive history outcomes: 5.2% |
| Manitoba, Canada | 1st trimester: 8.1%<br>2nd trimester: 26.4%<br>3rd trimester: 65.5% | Hospitalized: 27.7%<br>Non-hospitalized: 72.3% | 15-24 years: 27.3%<br>25-39 years: 68.9%<br>40-45 years: 3.8% | 12-24 years: 22.5%<br>25-39 years: 74.2%<br>40-55 years: 3.4% | Any: 38.7%<br>Cardiovascular: <2.6%<br>Respiratory: 7.2%<br>Severe obesity: 3.4% | Any: 42.6%<br>Cardiovascular: 1.4%<br>Respiratory: 9.2%<br>Severe obesity: 2.0% | Any: N.A.<br>Adverse reproductive history outcomes: 4.3% | Any: N.A.<br>Adverse reproductive history outcomes: 2.7% |
| Ontario, Canada | 1st trimester: 28.9%<br>2nd trimester: 32.5%<br>3rd trimester: 38.6% | Hospitalized: 0%<br>Non-hospitalized: 100% | 15-24 years: 34.7%<br>25-39 years: 61.7%<br>40-45 years: 3.5% | 12-24 years: 34.8%<br>25-39 years: 61.2%<br>40-55 years: 4.0% | Any: 28.9%<br>Cardiovascular: 0%<br>Respiratory: 2.6%<br>Severe obesity: 1.2% | Any: 33.8%<br>Cardiovascular: 0.03%<br>Respiratory: 2.7%<br>Severe obesity: 1.6% | Any: N.A.<br>Adverse reproductive history outcomes: 2.8% | Any: N.A.<br>Adverse reproductive history outcomes: 3.5% |
| U.S. | 1st trimester: 19.6%<br>2nd trimester: 27.1%<br>3rd trimester: 53.3% | Hospitalized: 3.2%<br>Non-hospitalized: 87.8% | 12-24 years: 16.7%<br>25-39 years: 79.9%<br>40-55 years: 3.4% | 12-24 years: 16.7%<br>25-39 years: 79.9%<br>40-55 years: 3.4% | Any: N.A.<br>Cardiovascular: 0.9%<br>Chronic lung: 6.3%<br>Severe obesity: 15.4% | Any: N.A.<br>Cardiovascular: 0.8%<br>Chronic lung: 6.2%<br>Severe obesity: 15.4% | Any: N.A.<br>Adverse pregnancy outcomes: 5.1% | Any: N.A.<br>Adverse pregnancy outcomes: 5.0% |

[1]Any co-morbidities included: cancer, cardiovascular disease, chronic kidney disease, chronic liver disease, chronic lung disease, common rheumatic disease, diabetes, HIV, hypertension, mental disorders, severe obesity, sickle cell disease, and use of immunosuppressants.[2]Any obstetric risk included: prior history of the following: gestational diabetes, gestational hypertension, pre-eclampsia, HELLP and adverse pregnancy outcomes. Adverse pregnancy outcomes included: stillbirth, spontaneous abortion, SGA, FGR, and major congenital abnormalities. [3]Cases in which the COVID-19 test/positive diagnosis was +/- 2 days of delivery date did not contribute to analyses by hospitalization. They did however contribute to the analyses undertaken on 'total cases'. Therefore, in some sites hospitalized and non-hospitalized cases do not add up to 100%. Abbreviations: N.A. = not available.

Figure S1. Forest plots showing the pooled prevalence of analgesics in the 30 days pre-COVID (left) and 30 days post-COVID (right) in non-hospitalized pregnant women with COVID-19 (upper) and pregnant women without COVID-19 (lower), by pregnancy trimester

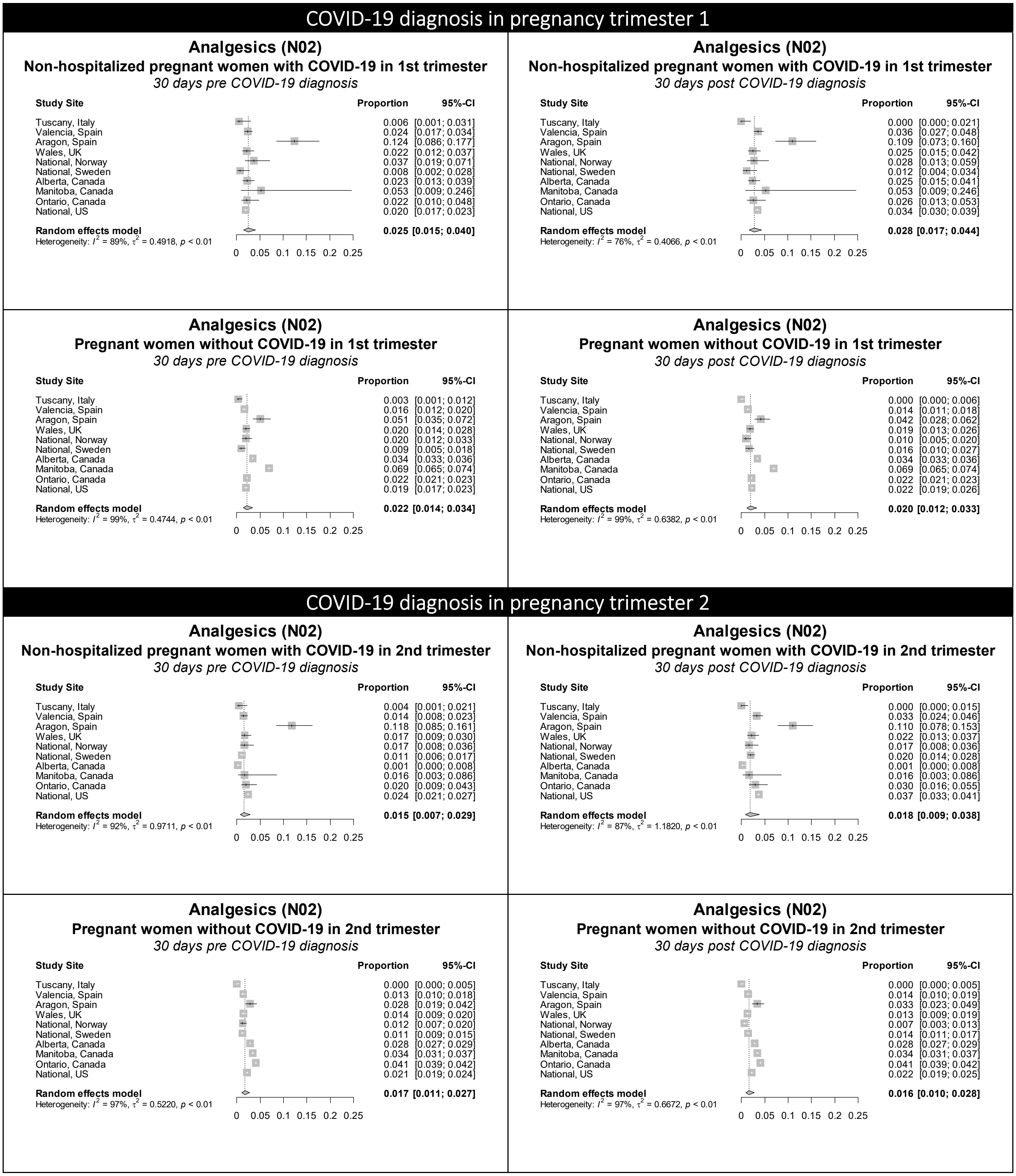

**Figure S1 continued.** Forest plots showing the pooled prevalence of analgesics in the 30 days pre-COVID (left) and 30 days post-COVID (right) in non-hospitalized pregnant women with COVID-19 (upper) and pregnant women without COVID-19 (lower), by pregnancy trimester

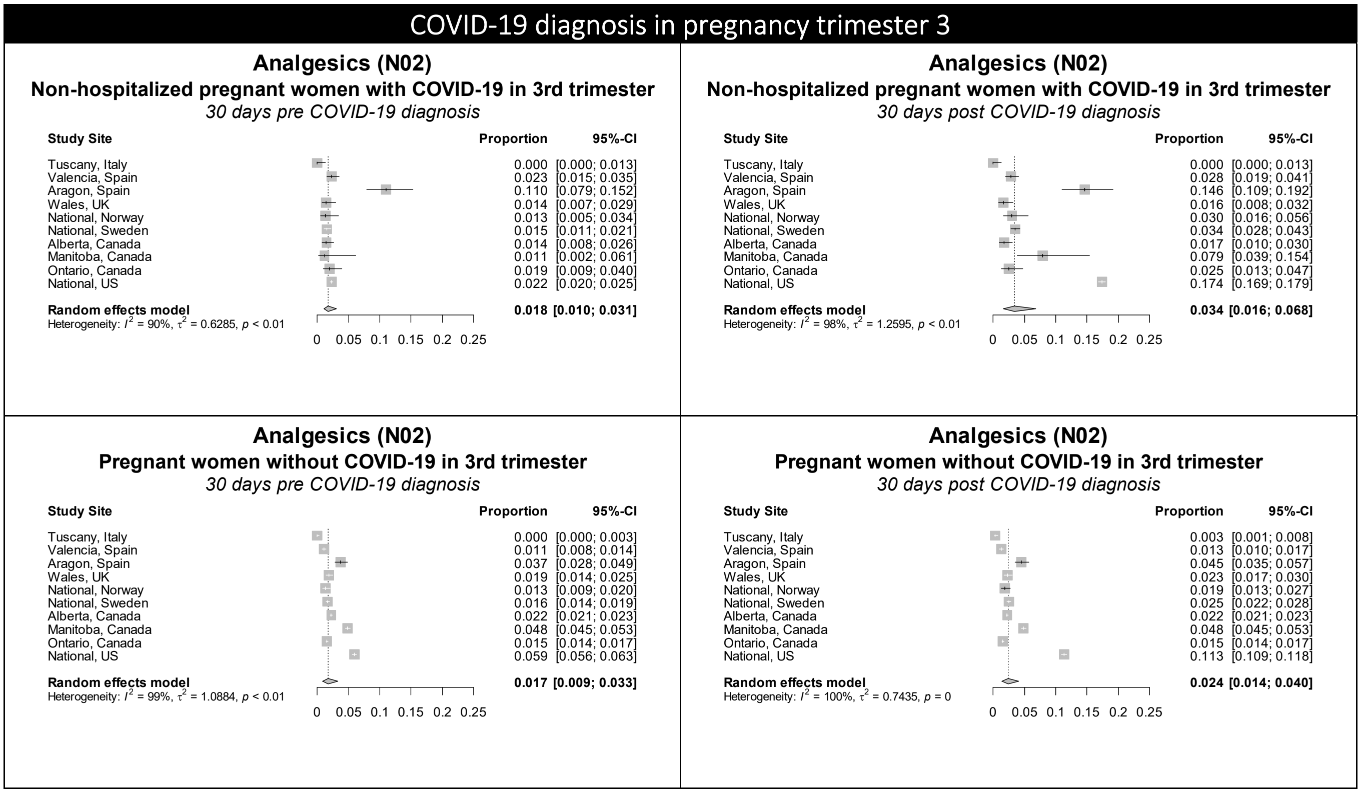

Figure S2. Forest plots showing the pooled prevalence of anthelminthics in the 30 days pre-COVID (left) and 30 days post-COVID (right) in non-hospitalized pregnant women with COVID-19 (upper) and pregnant women without COVID-19 (lower), by pregnancy trimester

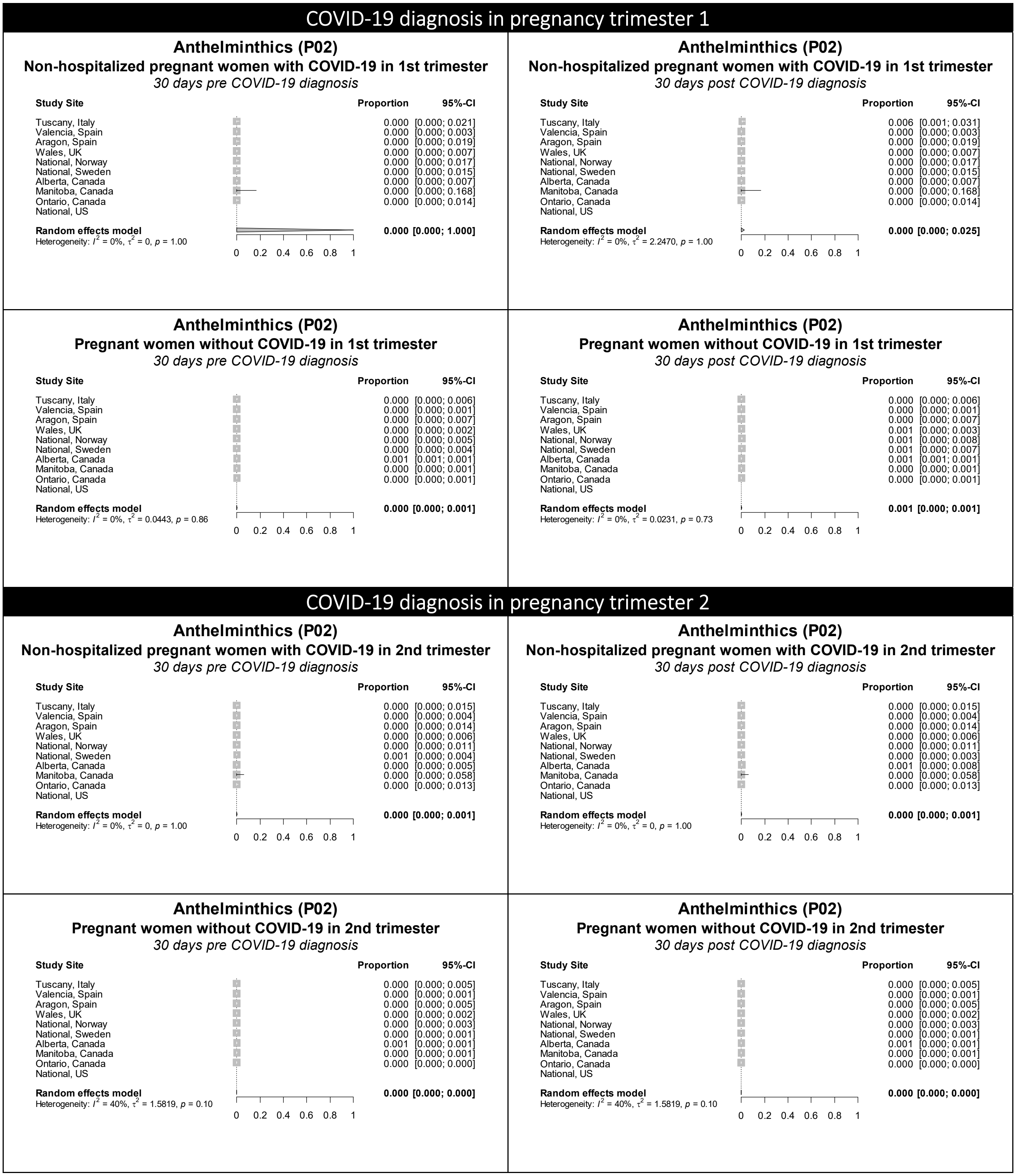

Figure S2 continued. Forest plots showing the pooled prevalence of anthelminthics in the 30 days pre-COVID (left) and 30 days post-COVID (right) in non-hospitalized pregnant women with COVID-19 (upper) and pregnant women without COVID-19 (lower), by pregnancy trimester

### COVID-19 diagnosis in pregnancy trimester 3

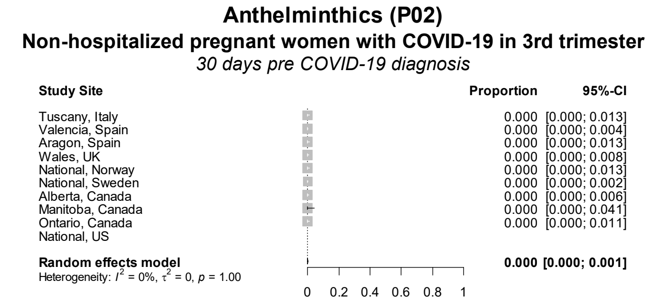

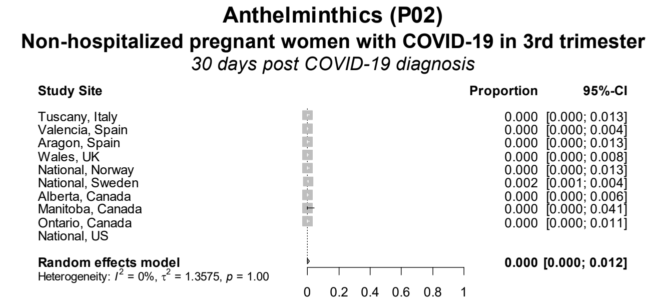

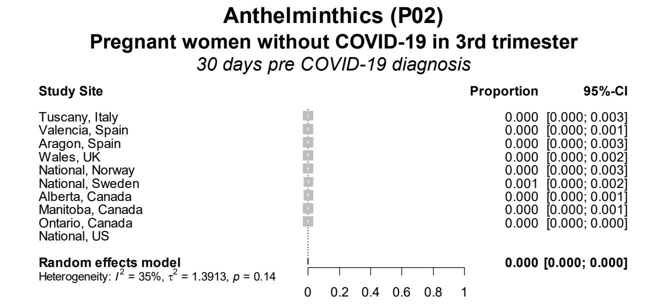

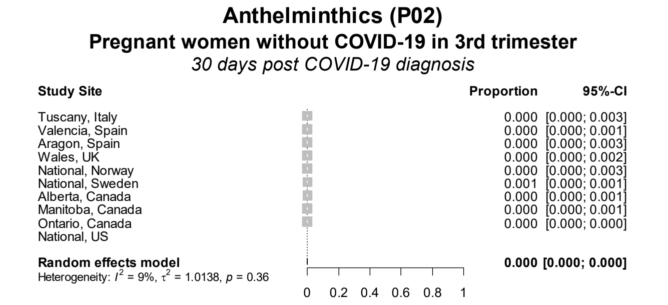

Figure S3. Forest plots showing the pooled prevalence of anti-inflammatory and antirheumatic products in the 30 days pre-COVID (left) and 30 days post-COVID (right) in non-hospitalized pregnant women with COVID-19 (upper) and pregnant women without COVID-19 (lower), by pregnancy trimester

### COVID-19 diagnosis in pregnancy trimester 1

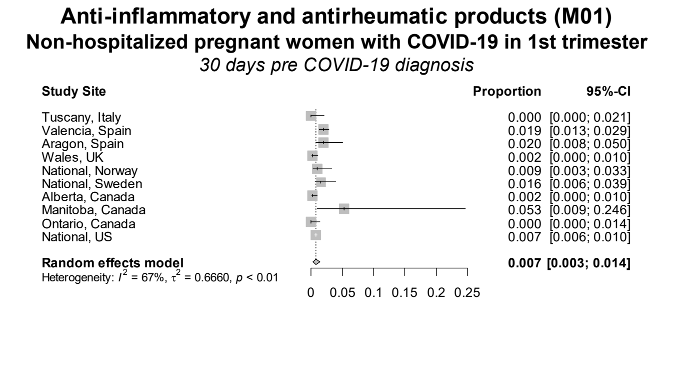

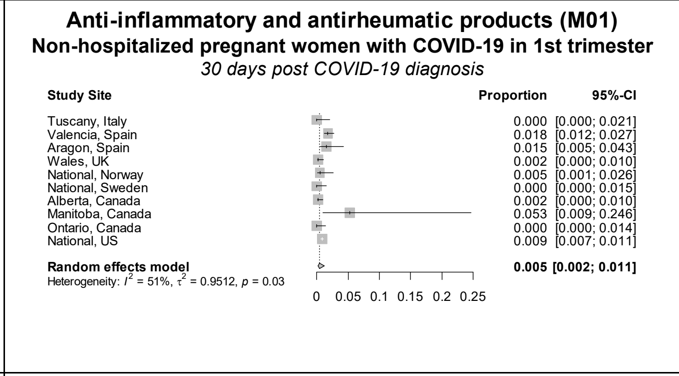

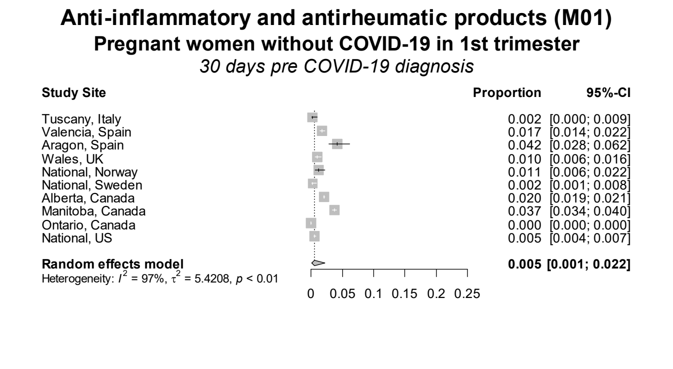

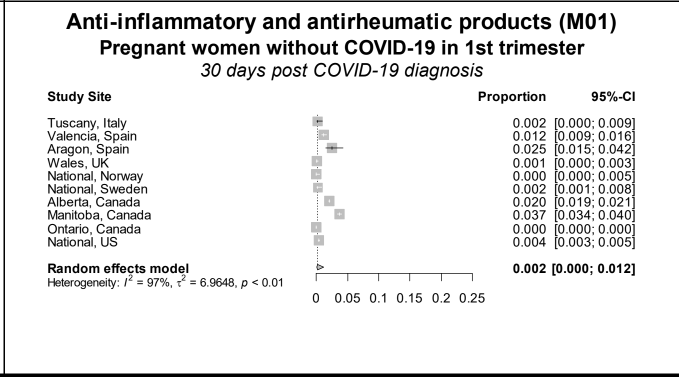

### COVID-19 diagnosis in pregnancy trimester 2

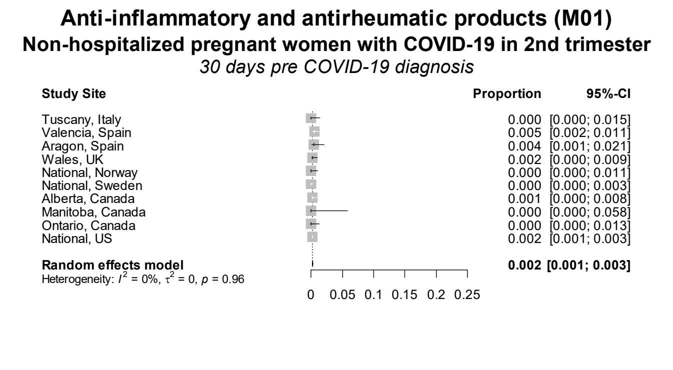

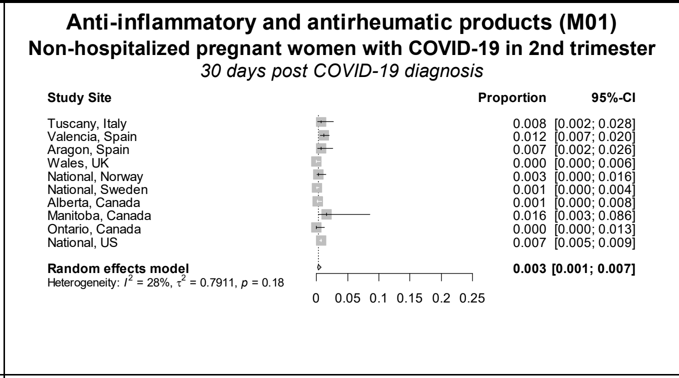

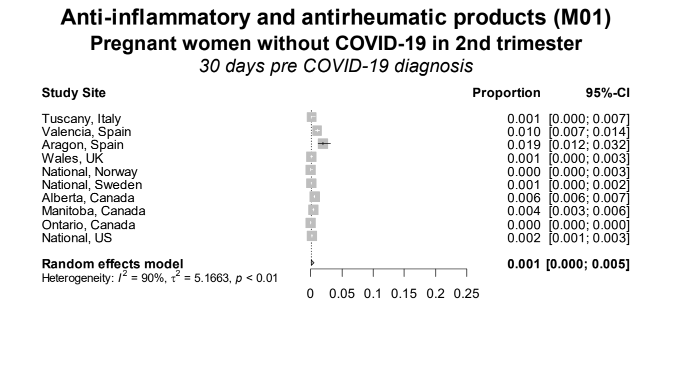

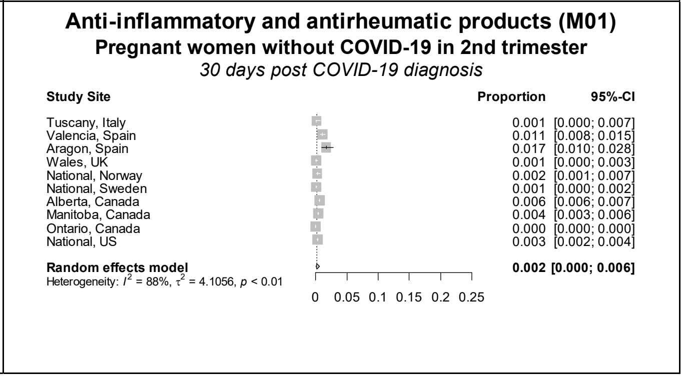

**Figure S3 continued.** Forest plots showing the pooled prevalence of anti-inflammatory and antirheumatic products in the 30 days pre-COVID (left) and 30 days post-COVID (right) in non-hospitalized pregnant women with COVID-19 (upper) and pregnant women without COVID-19 (lower), by pregnancy trimester

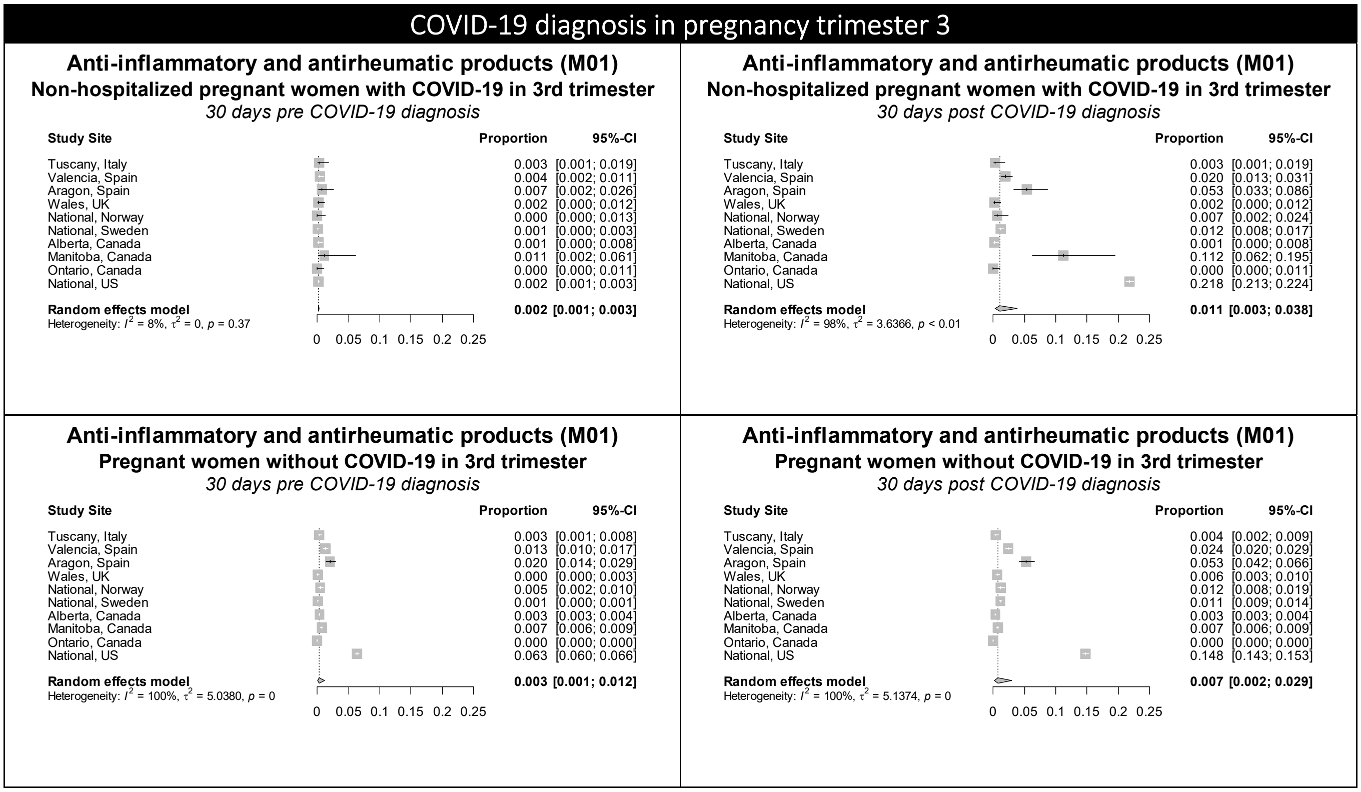

Figure S4. Forest plots showing the pooled prevalence of antibacterials in the 30 days pre-COVID (left) and 30 days post-COVID (right) in non-hospitalized pregnant women with COVID-19 (upper) and pregnant women without COVID-19 (lower), by pregnancy trimester

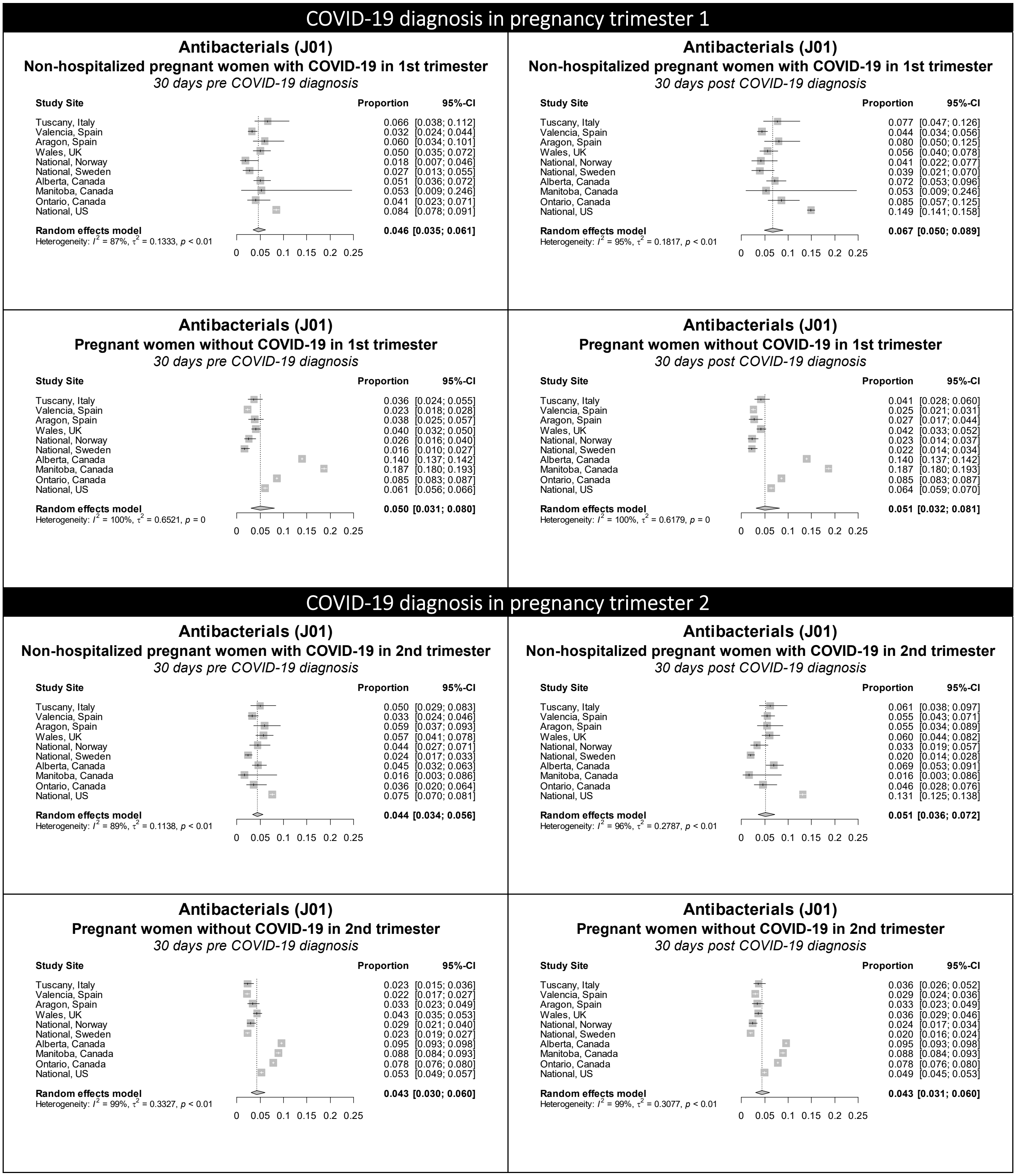

**Figure S4 continued.** Forest plots showing the pooled prevalence of antibacterials in the 30 days pre-COVID (left) and 30 days post-COVID (right) in non-hospitalized pregnant women with COVID-19 (upper) and pregnant women without COVID-19 (lower), by pregnancy trimester

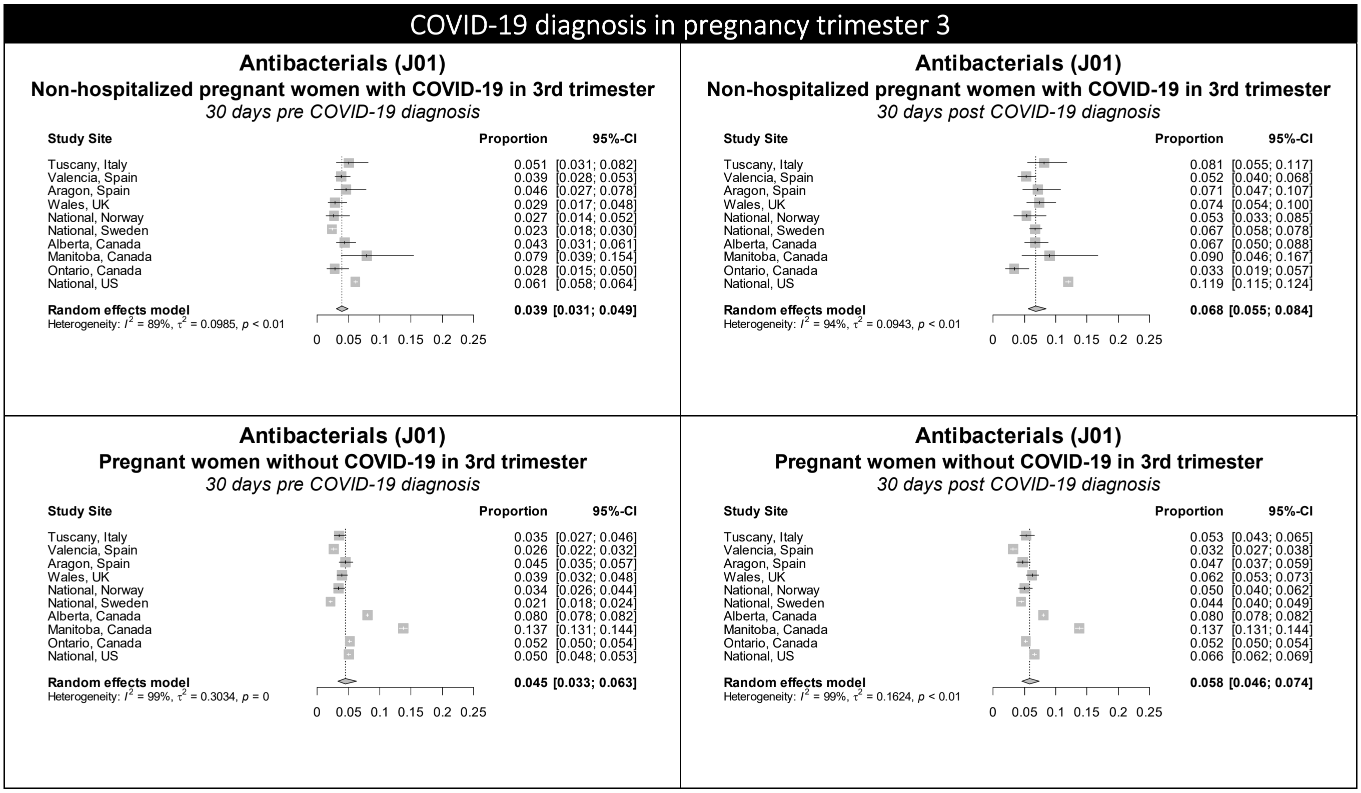

Figure S5. Forest plots showing the pooled prevalence of antigout preparations in the 30 days pre-COVID (left) and 30 days post-COVID (right) in non-hospitalized pregnant women with COVID-19 (upper) and pregnant women without COVID-19 (lower), by pregnancy trimester

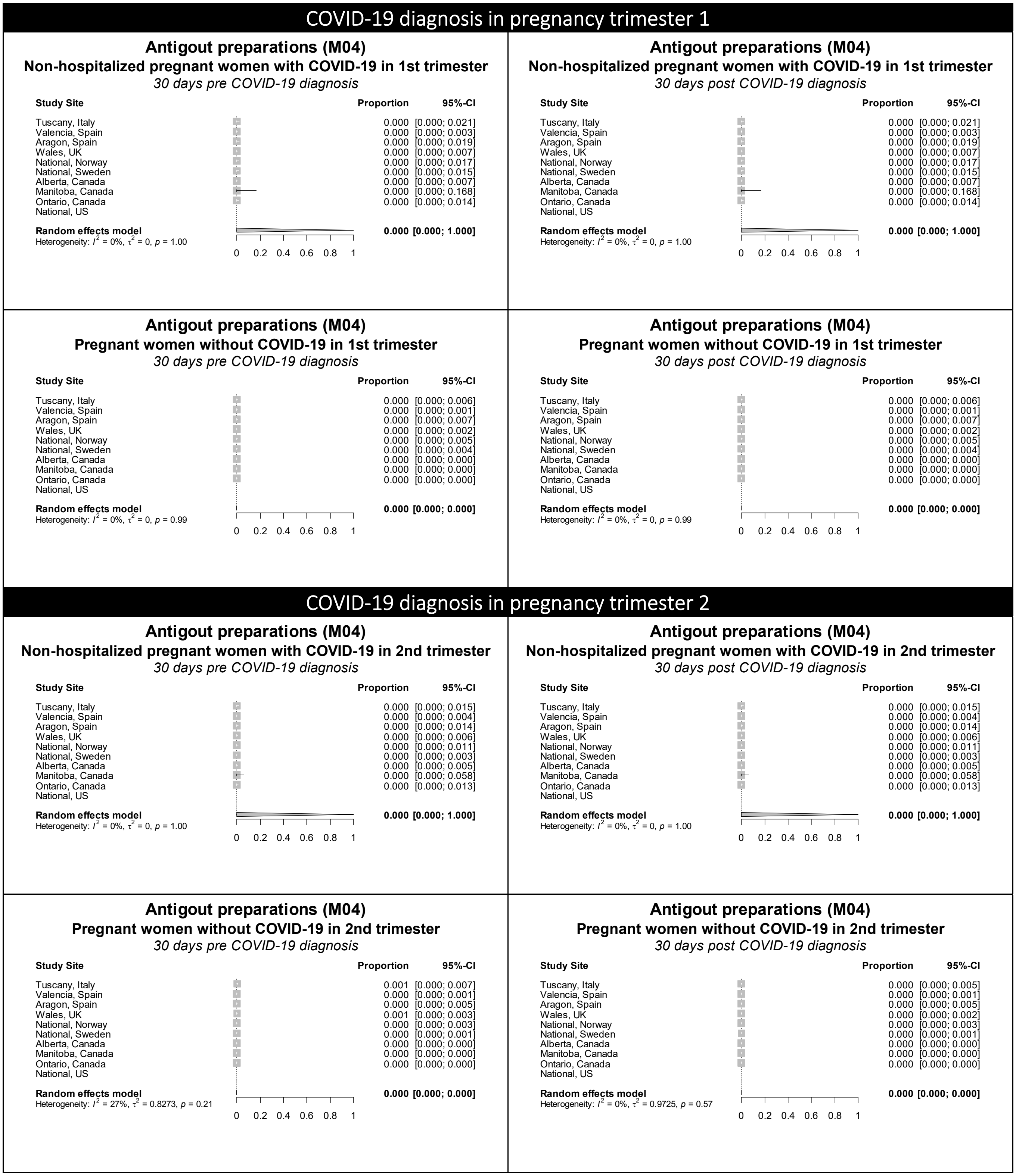

**Figure S5 continued.** Forest plots showing the pooled prevalence of antigout preparations in the 30 days pre-COVID (left) and 30 days post-COVID (right) in non-hospitalized pregnant women with COVID-19 (upper) and pregnant women without COVID-19 (lower), by pregnancy trimester

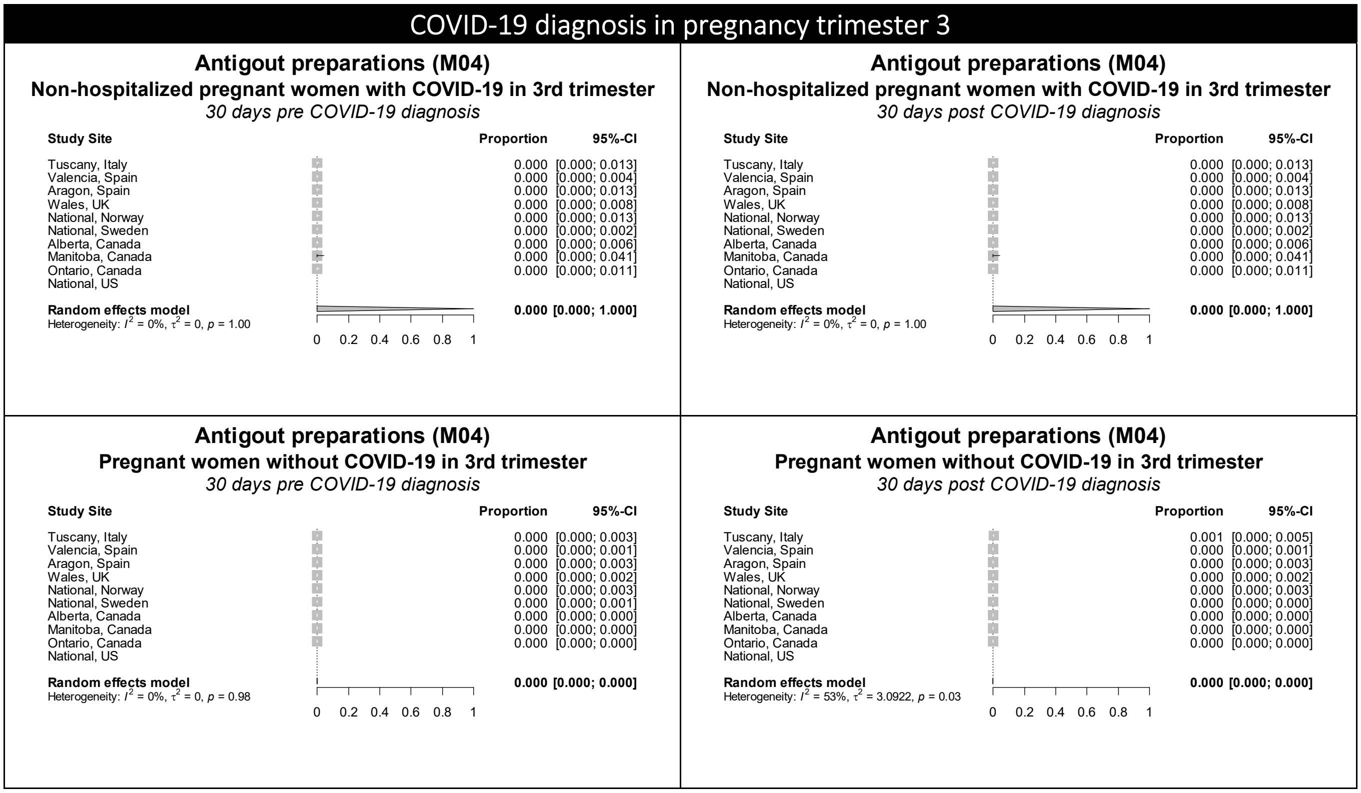

Figure S6. Forest plots showing the pooled prevalence of antihypertensives in the 30 days pre-COVID (left) and 30 days post-COVID (right) in non-hospitalized pregnant women with COVID-19 (upper) and pregnant women without COVID-19 (lower), by pregnancy trimester

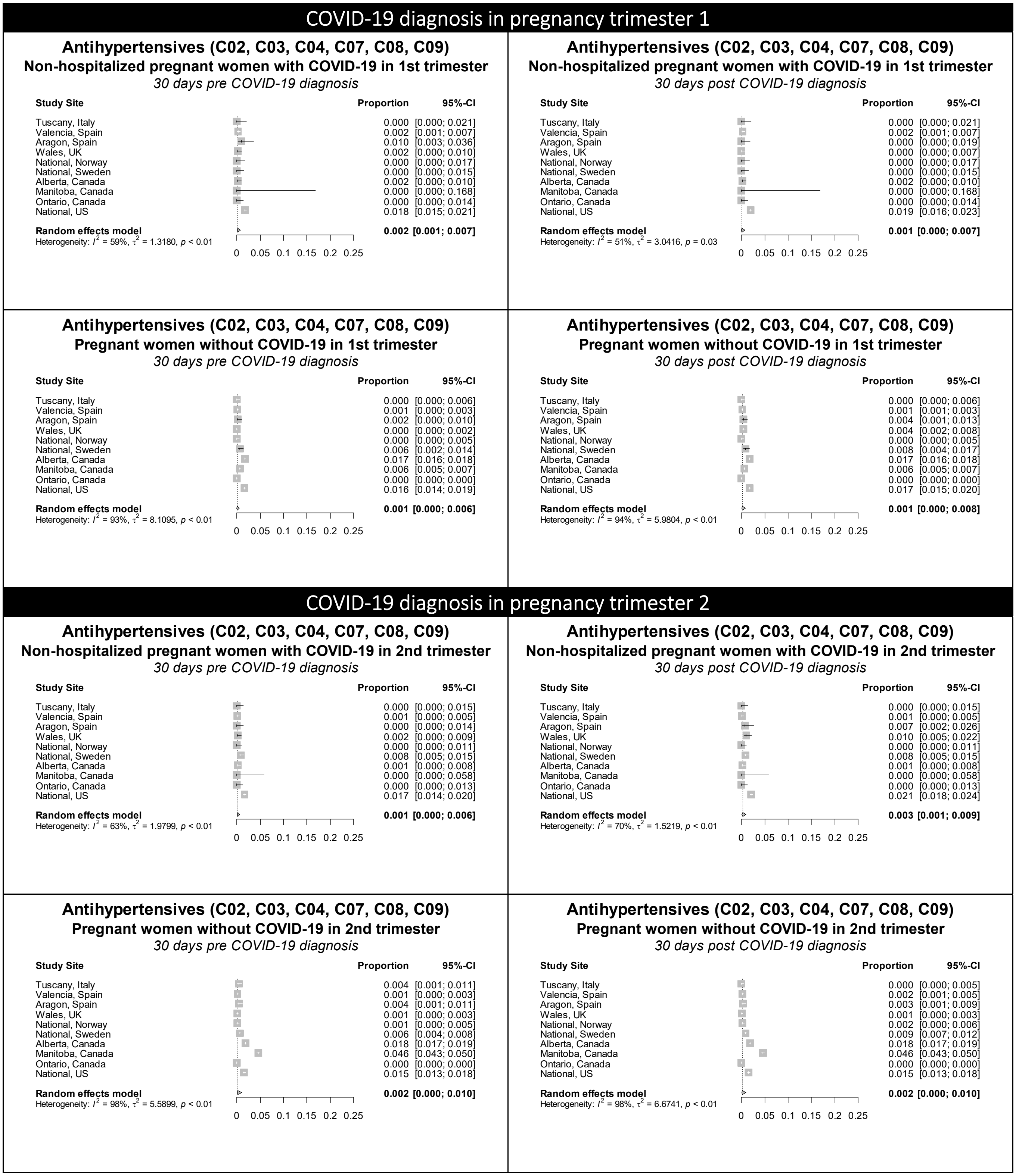

Figure S6 continued. Forest plots showing the pooled prevalence of antihypertensives in the 30 days pre-COVID (left) and 30 days post-COVID (right) in non-hospitalized pregnant women with COVID-19 (upper) and pregnant women without COVID-19 (lower), by pregnancy trimester

**COVID-19 diagnosis in pregnancy trimester 3**

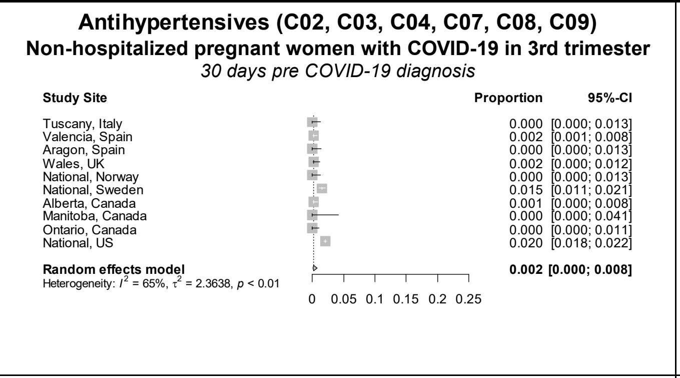

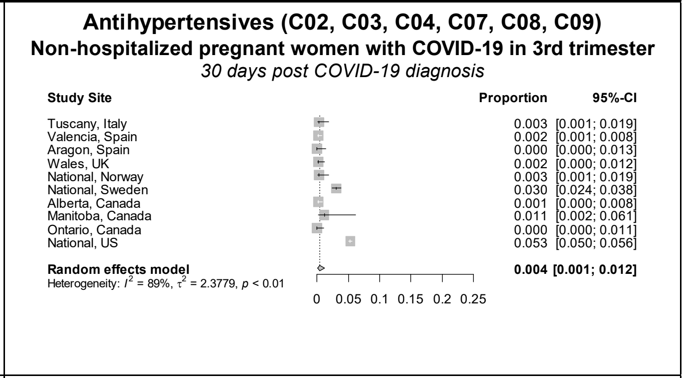

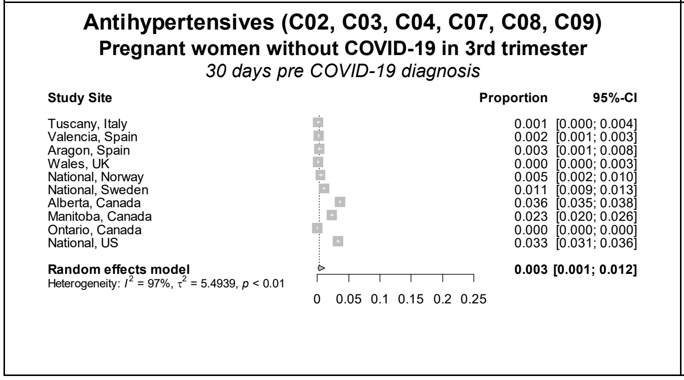

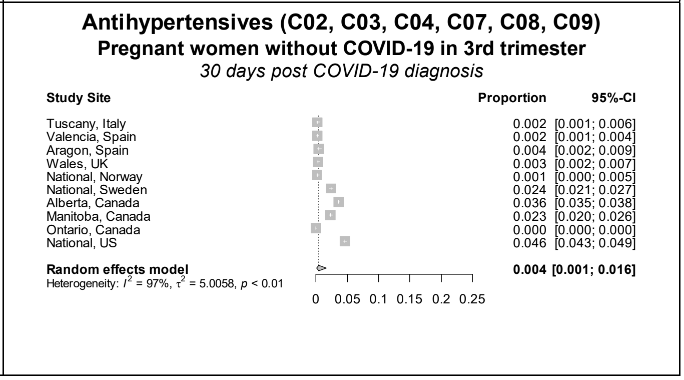

Figure S7. Forest plots showing the pooled prevalence of antimycobacterials in the 30 days pre-COVID (left) and 30 days post-COVID (right) in non-hospitalized pregnant women with COVID-19 (upper) and pregnant women without COVID-19 (lower), by pregnancy trimester

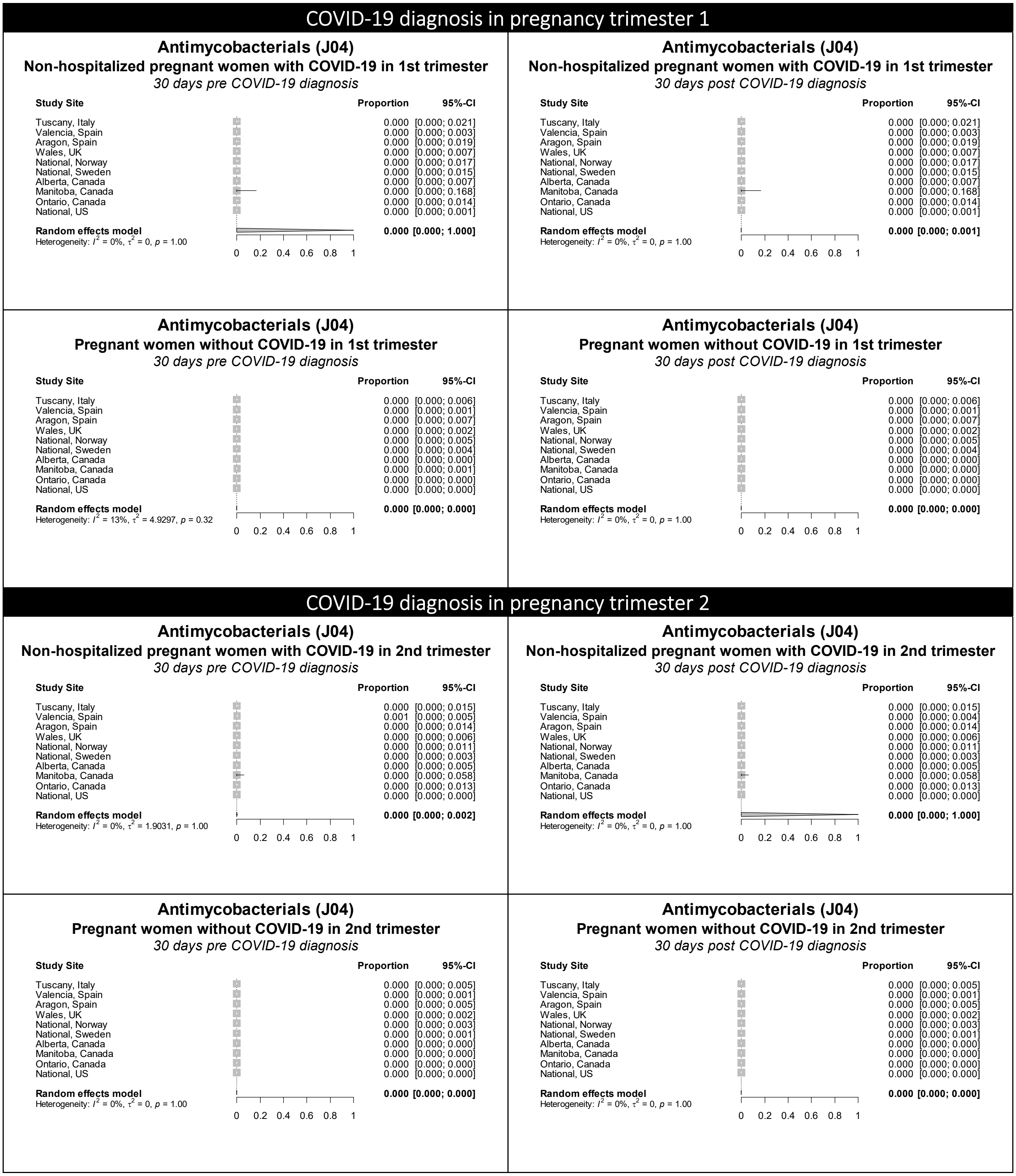

Figure S7 continued. Forest plots showing the pooled prevalence of antimycobacterials in the 30 days pre-COVID (left) and 30 days post-COVID (right) in non-hospitalized pregnant women with COVID-19 (upper) and pregnant women without COVID-19 (lower), by pregnancy trimester

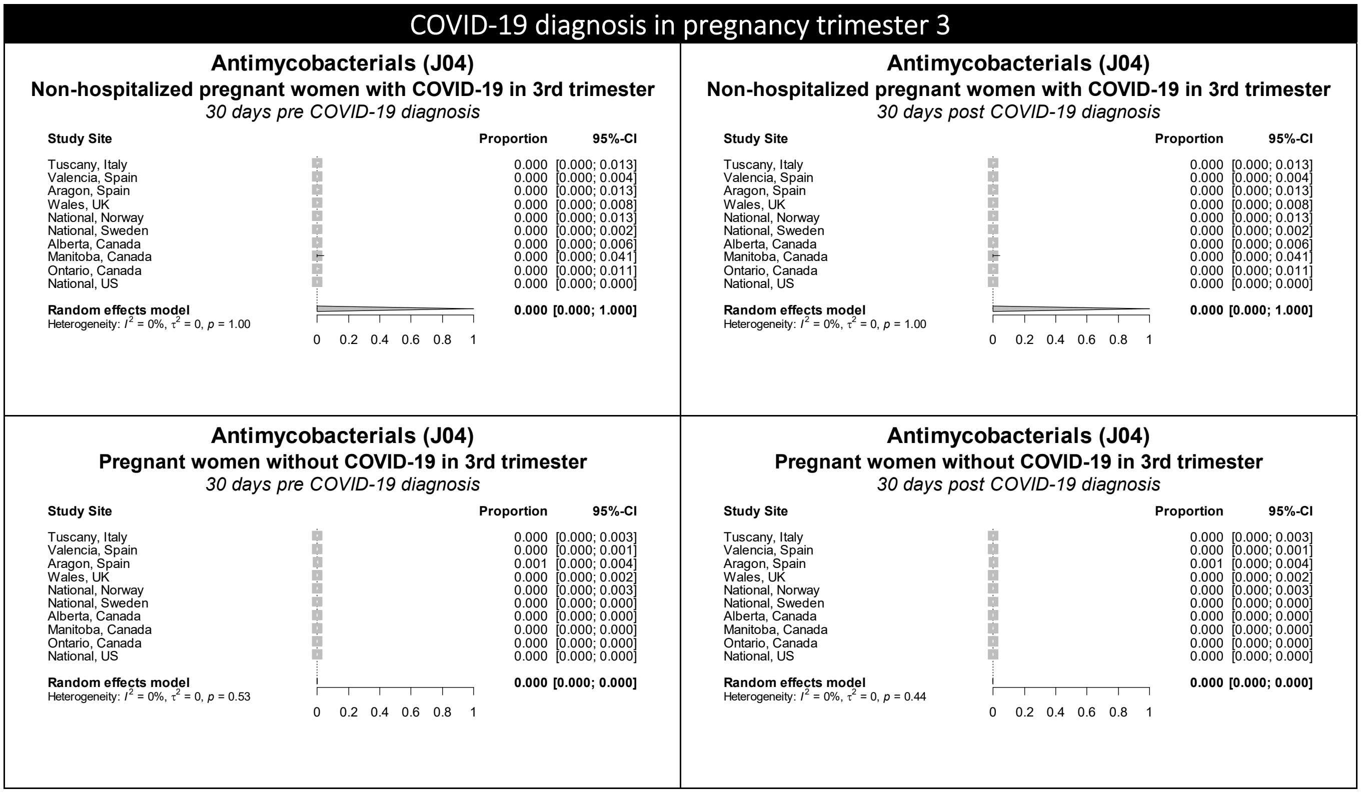

Figure S8. Forest plots showing the pooled prevalence of antimycotics in the 30 days pre-COVID (left) and 30 days post-COVID (right) in non-hospitalized pregnant women with COVID-19 (upper) and pregnant women without COVID-19 (lower), by pregnancy trimester

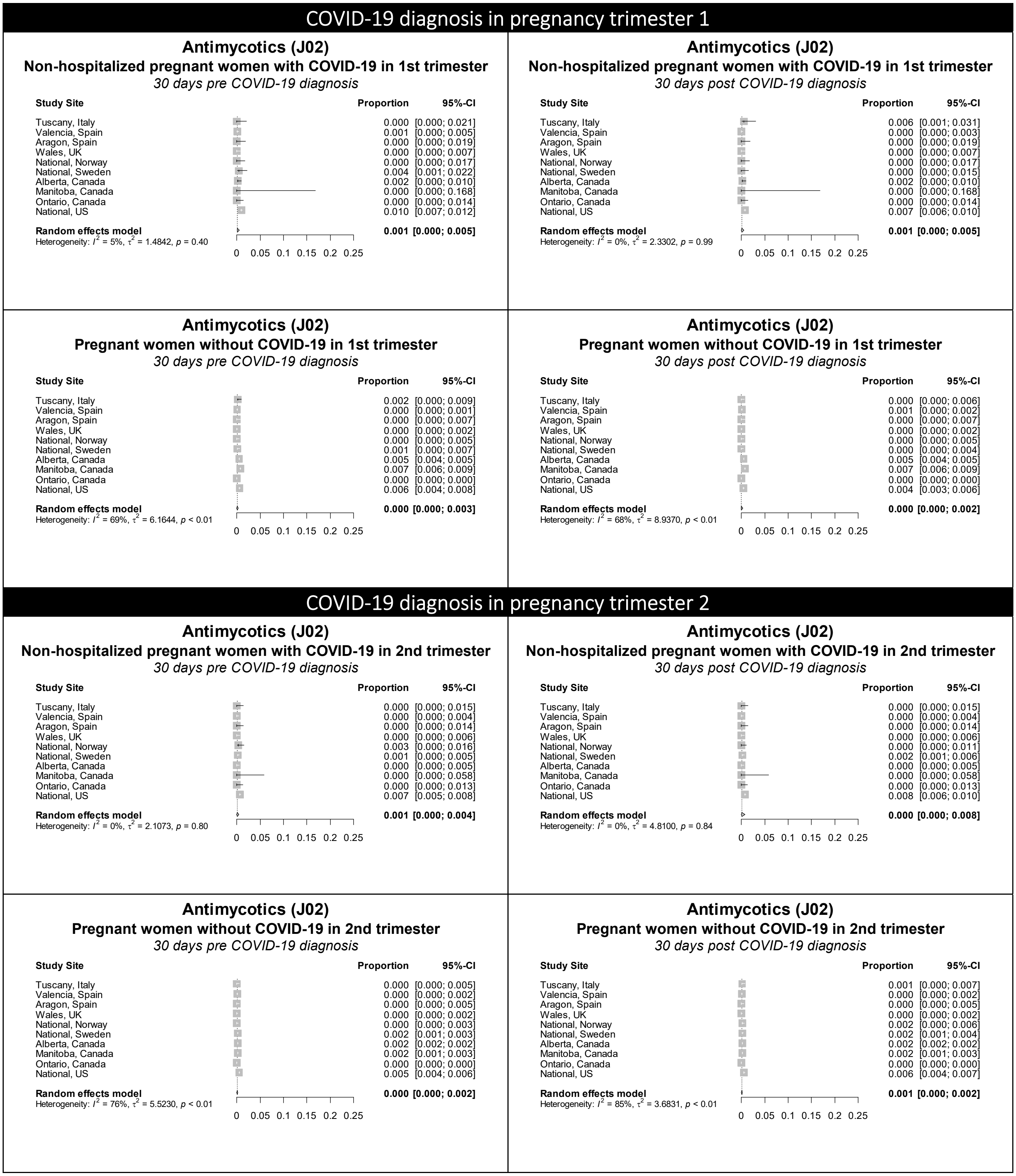

**Figure S8 continued.** Forest plots showing the pooled prevalence of antimycotics in the 30 days pre-COVID (left) and 30 days post-COVID (right) in non-hospitalized pregnant women with COVID-19 (upper) and pregnant women without COVID-19 (lower), by pregnancy trimester

### COVID-19 diagnosis in pregnancy trimester 3

#### Antimycotics (J02)
**Non-hospitalized pregnant women with COVID-19 in 3rd trimester**
*30 days pre COVID-19 diagnosis*

| Study Site | Proportion | 95%-CI |
|---|---|---|
| Tuscany, Italy | 0.000 | [0.000; 0.013] |
| Valencia, Spain | 0.000 | [0.000; 0.004] |
| Aragon, Spain | 0.000 | [0.000; 0.013] |
| Wales, UK | 0.000 | [0.000; 0.008] |
| National, Norway | 0.007 | [0.002; 0.024] |
| National, Sweden | 0.004 | [0.002; 0.008] |
| Alberta, Canada | 0.001 | [0.000; 0.008] |
| Manitoba, Canada | 0.000 | [0.000; 0.041] |
| Ontario, Canada | 0.000 | [0.000; 0.011] |
| National, US | 0.008 | [0.007; 0.010] |
| **Random effects model** | **0.001** | **[0.000; 0.006]** |

Heterogeneity: $I^2$ = 0%, $\tau^2$ = 1.6038, $p$ = 0.59

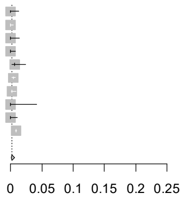

#### Antimycotics (J02)
**Non-hospitalized pregnant women with COVID-19 in 3rd trimester**
*30 days post COVID-19 diagnosis*

| Study Site | Proportion | 95%-CI |
|---|---|---|
| Tuscany, Italy | 0.003 | [0.001; 0.019] |
| Valencia, Spain | 0.000 | [0.000; 0.004] |
| Aragon, Spain | 0.004 | [0.001; 0.020] |
| Wales, UK | 0.000 | [0.000; 0.008] |
| National, Norway | 0.000 | [0.000; 0.013] |
| National, Sweden | 0.002 | [0.001; 0.004] |
| Alberta, Canada | 0.000 | [0.000; 0.006] |
| Manitoba, Canada | 0.000 | [0.000; 0.041] |
| Ontario, Canada | 0.000 | [0.000; 0.011] |
| National, US | 0.011 | [0.010; 0.013] |
| **Random effects model** | **0.001** | **[0.000; 0.005]** |

Heterogeneity: $I^2$ = 47%, $\tau^2$ = 2.1226, $p$ = 0.05

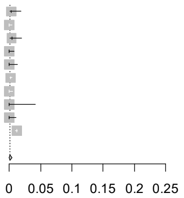

#### Antimycotics (J02)
**Pregnant women without COVID-19 in 3rd trimester**
*30 days pre COVID-19 diagnosis*

| Study Site | Proportion | 95%-CI |
|---|---|---|
| Tuscany, Italy | 0.001 | [0.000; 0.005] |
| Valencia, Spain | 0.001 | [0.000; 0.002] |
| Aragon, Spain | 0.001 | [0.000; 0.004] |
| Wales, UK | 0.000 | [0.000; 0.002] |
| National, Norway | 0.001 | [0.000; 0.005] |
| National, Sweden | 0.003 | [0.002; 0.005] |
| Alberta, Canada | 0.002 | [0.002; 0.002] |
| Manitoba, Canada | 0.002 | [0.001; 0.003] |
| Ontario, Canada | 0.000 | [0.000; 0.000] |
| National, US | 0.007 | [0.006; 0.008] |
| **Random effects model** | **0.001** | **[0.000; 0.002]** |

Heterogeneity: $I^2$ = 93%, $\tau^2$ = 2.5813, $p$ < 0.01

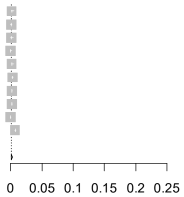

#### Antimycotics (J02)
**Pregnant women without COVID-19 in 3rd trimester**
*30 days post COVID-19 diagnosis*

| Study Site | Proportion | 95%-CI |
|---|---|---|
| Tuscany, Italy | 0.001 | [0.000; 0.004] |
| Valencia, Spain | 0.001 | [0.000; 0.002] |
| Aragon, Spain | 0.000 | [0.000; 0.003] |
| Wales, UK | 0.003 | [0.002; 0.007] |
| National, Norway | 0.002 | [0.001; 0.006] |
| National, Sweden | 0.004 | [0.003; 0.005] |
| Alberta, Canada | 0.002 | [0.002; 0.002] |
| Manitoba, Canada | 0.002 | [0.001; 0.003] |
| Ontario, Canada | 0.000 | [0.000; 0.000] |
| National, US | 0.011 | [0.010; 0.012] |
| **Random effects model** | **0.001** | **[0.000; 0.003]** |

Heterogeneity: $I^2$ = 97%, $\tau^2$ = 2.8329, $p$ < 0.01

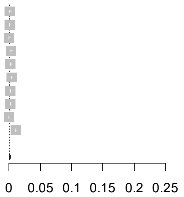

Figure S9. Forest plots showing the pooled prevalence of antineoplastic agents in the 30 days pre-COVID (left) and 30 days post-COVID (right) in non-hospitalized pregnant women with COVID-19 (upper) and pregnant women without COVID-19 (lower), by pregnancy trimester

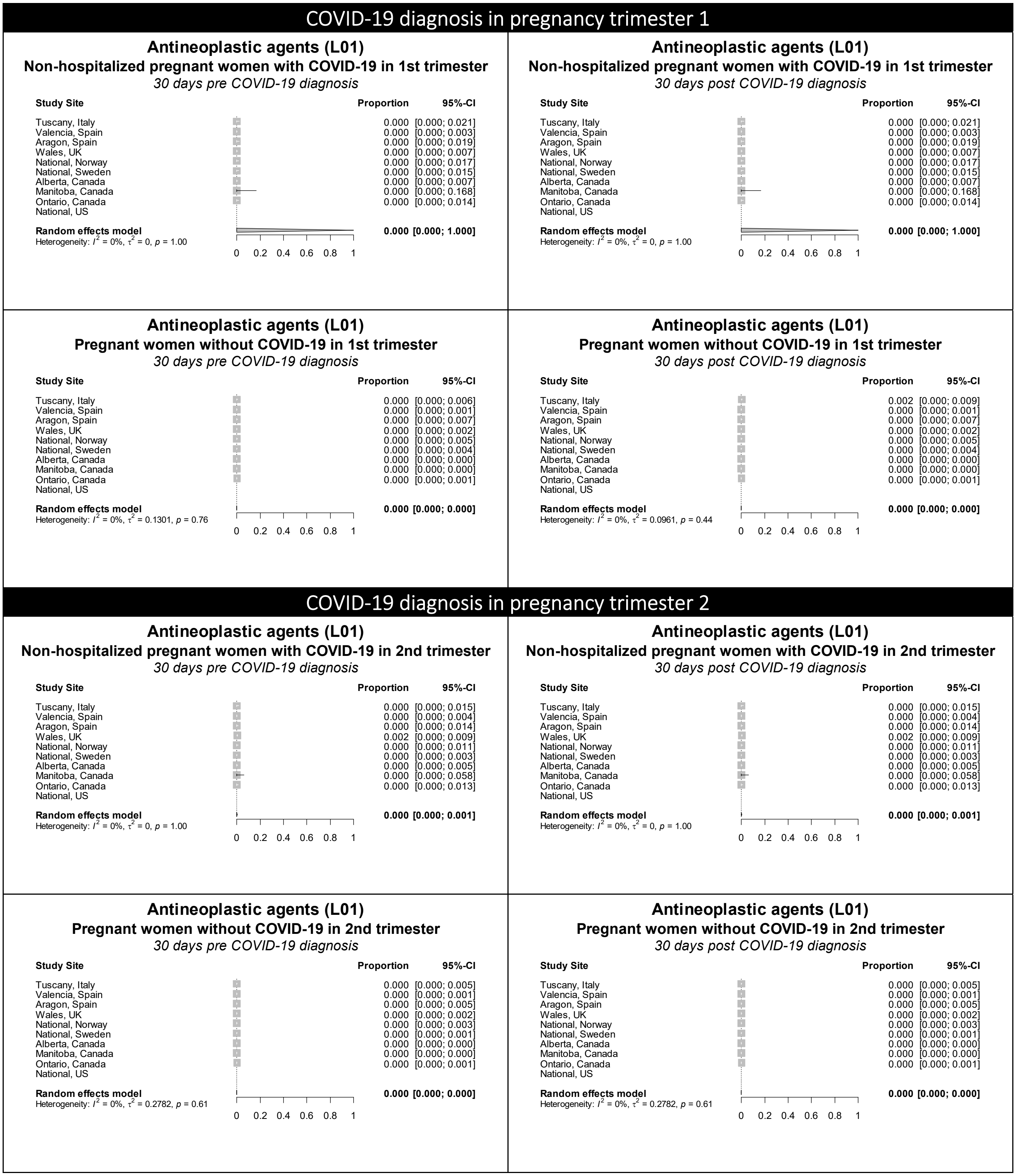

**Figure S9 continued.** Forest plots showing the pooled prevalence of antineoplastic agents in the 30 days pre-COVID (left) and 30 days post-COVID (right) in non-hospitalized pregnant women with COVID-19 (upper) and pregnant women without COVID-19 (lower), by pregnancy trimester

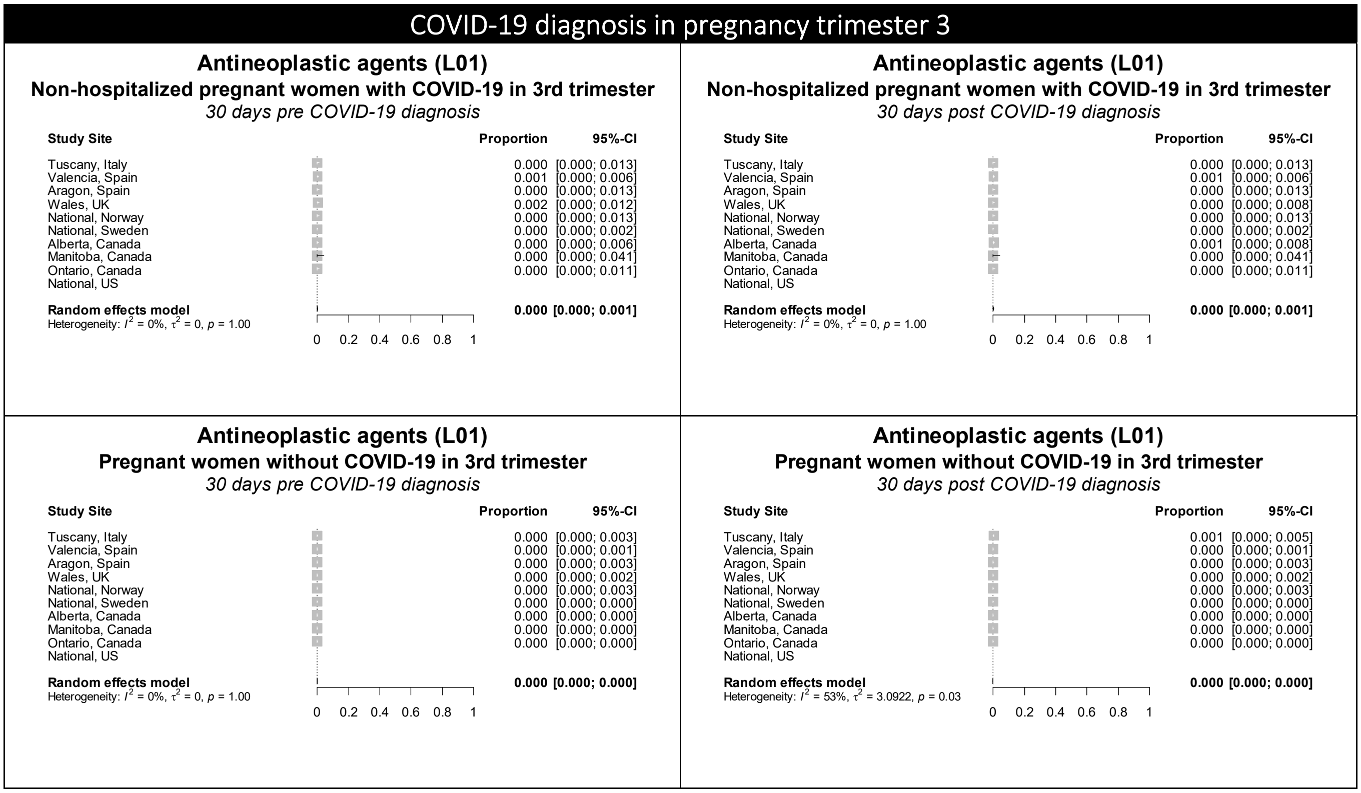

Figure S10. Forest plots showing the pooled prevalence of antiprotozoals in the 30 days pre-COVID (left) and 30 days post-COVID (right) in non-hospitalized pregnant women with COVID-19 (upper) and pregnant women without COVID-19 (lower), by pregnancy trimester

### COVID-19 diagnosis in pregnancy trimester 1

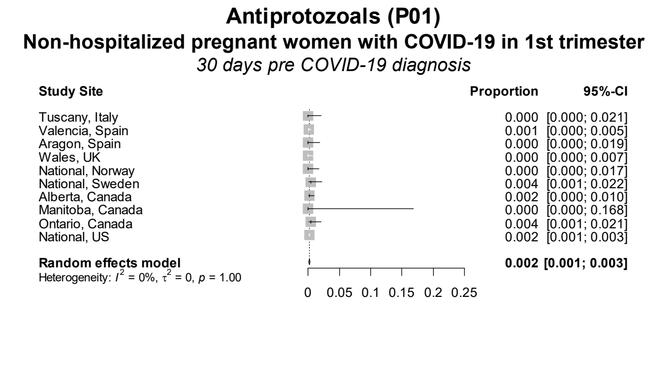

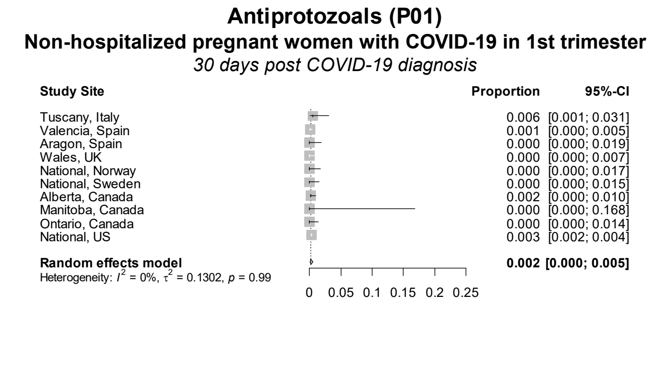

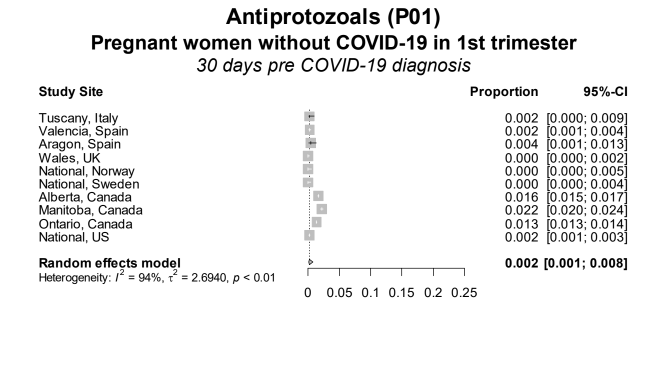

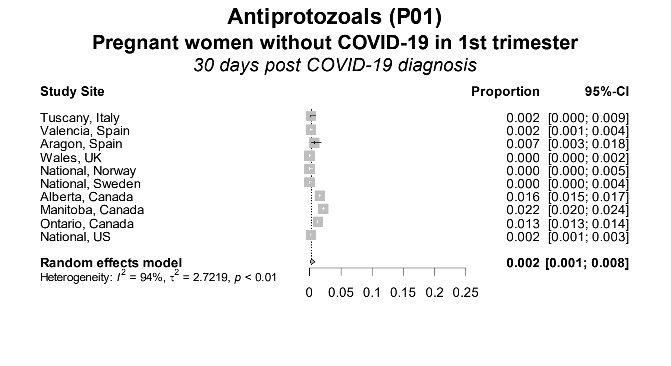

### COVID-19 diagnosis in pregnancy trimester 2

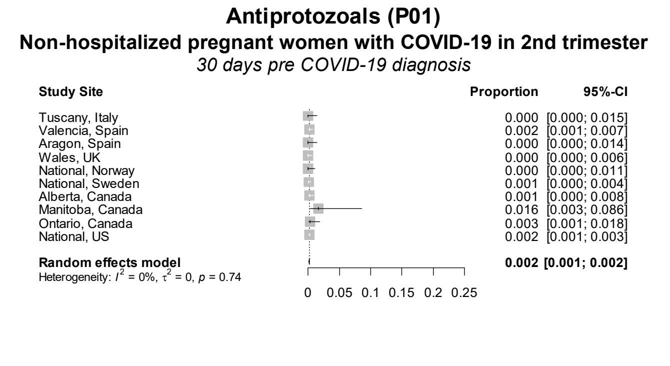

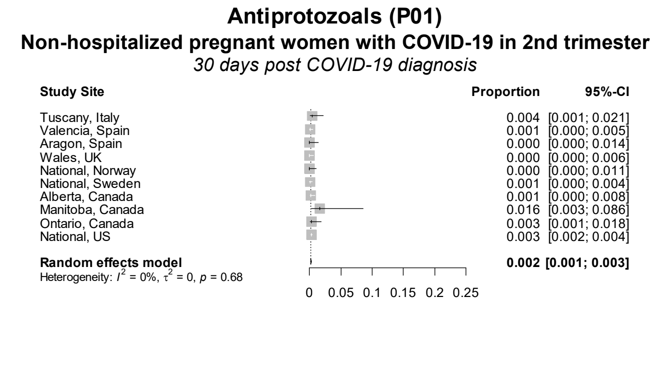

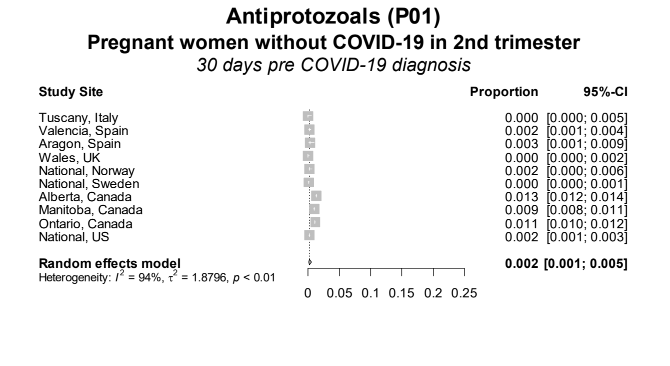

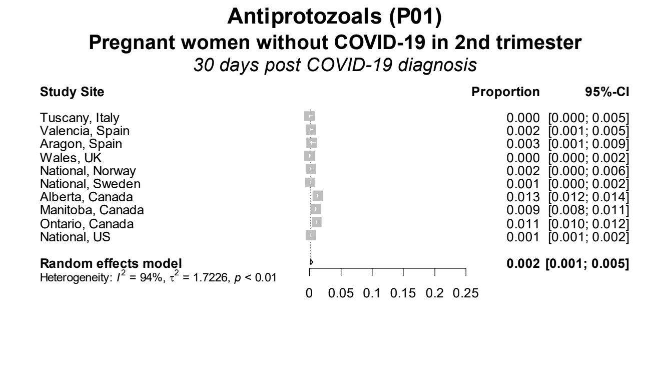

**Figure S10 continued.** Forest plots showing the pooled prevalence of antiprotozoals in the 30 days pre-COVID (left) and 30 days post-COVID (right) in non-hospitalized pregnant women with COVID-19 (upper) and pregnant women without COVID-19 (lower), by pregnancy trimester

### COVID-19 diagnosis in pregnancy trimester 3

#### Antiprotozoals (P01)
**Non-hospitalized pregnant women with COVID-19 in 3rd trimester**
*30 days pre COVID-19 diagnosis*

| Study Site | Proportion | 95%-CI |
|---|---|---|
| Tuscany, Italy | 0.000 | [0.000; 0.013] |
| Valencia, Spain | 0.002 | [0.001; 0.008] |
| Aragon, Spain | 0.004 | [0.001; 0.020] |
| Wales, UK | 0.000 | [0.000; 0.008] |
| National, Norway | 0.003 | [0.001; 0.019] |
| National, Sweden | 0.000 | [0.000; 0.002] |
| Alberta, Canada | 0.001 | [0.000; 0.008] |
| Manitoba, Canada | 0.011 | [0.002; 0.061] |
| Ontario, Canada | 0.003 | [0.000; 0.016] |
| National, US | 0.001 | [0.001; 0.002] |
| **Random effects model** | **0.001** | **[0.001; 0.002]** |

Heterogeneity: $I^2 = 0\%$, $\tau^2 = 0$, $p = 0.45$

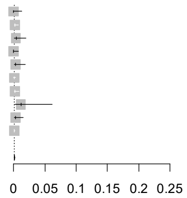

#### Antiprotozoals (P01)
**Non-hospitalized pregnant women with COVID-19 in 3rd trimester**
*30 days post COVID-19 diagnosis*

| Study Site | Proportion | 95%-CI |
|---|---|---|
| Tuscany, Italy | 0.000 | [0.000; 0.013] |
| Valencia, Spain | 0.000 | [0.000; 0.004] |
| Aragon, Spain | 0.004 | [0.001; 0.020] |
| Wales, UK | 0.000 | [0.000; 0.008] |
| National, Norway | 0.013 | [0.005; 0.034] |
| National, Sweden | 0.020 | [0.015; 0.027] |
| Alberta, Canada | 0.001 | [0.000; 0.008] |
| Manitoba, Canada | 0.011 | [0.002; 0.061] |
| Ontario, Canada | 0.000 | [0.000; 0.011] |
| National, US | 0.001 | [0.001; 0.002] |
| **Random effects model** | **0.002** | **[0.001; 0.006]** |

Heterogeneity: $I^2 = 93\%$, $\tau^2 = 2.2103$, $p < 0.01$

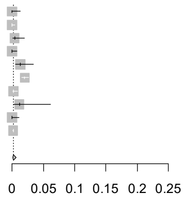

#### Antiprotozoals (P01)
**Pregnant women without COVID-19 in 3rd trimester**
*30 days pre COVID-19 diagnosis*

| Study Site | Proportion | 95%-CI |
|---|---|---|
| Tuscany, Italy | 0.002 | [0.001; 0.006] |
| Valencia, Spain | 0.002 | [0.001; 0.004] |
| Aragon, Spain | 0.004 | [0.002; 0.010] |
| Wales, UK | 0.000 | [0.000; 0.002] |
| National, Norway | 0.003 | [0.001; 0.007] |
| National, Sweden | 0.000 | [0.000; 0.001] |
| Alberta, Canada | 0.010 | [0.009; 0.011] |
| Manitoba, Canada | 0.015 | [0.013; 0.018] |
| Ontario, Canada | 0.007 | [0.006; 0.008] |
| National, US | 0.001 | [0.001; 0.002] |
| **Random effects model** | **0.003** | **[0.001; 0.006]** |

Heterogeneity: $I^2 = 96\%$, $\tau^2 = 1.3470$, $p < 0.01$

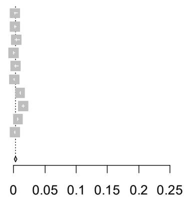

#### Antiprotozoals (P01)
**Pregnant women without COVID-19 in 3rd trimester**
*30 days post COVID-19 diagnosis*

| Study Site | Proportion | 95%-CI |
|---|---|---|
| Tuscany, Italy | 0.002 | [0.001; 0.006] |
| Valencia, Spain | 0.003 | [0.002; 0.005] |
| Aragon, Spain | 0.004 | [0.002; 0.010] |
| Wales, UK | 0.000 | [0.000; 0.002] |
| National, Norway | 0.003 | [0.001; 0.007] |
| National, Sweden | 0.012 | [0.010; 0.014] |
| Alberta, Canada | 0.010 | [0.009; 0.011] |
| Manitoba, Canada | 0.015 | [0.013; 0.018] |
| Ontario, Canada | 0.007 | [0.006; 0.008] |
| National, US | 0.001 | [0.001; 0.002] |
| **Random effects model** | **0.004** | **[0.002; 0.008]** |

Heterogeneity: $I^2 = 96\%$, $\tau^2 = 1.0555$, $p < 0.01$

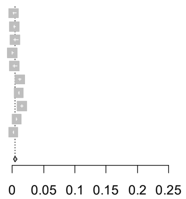

Figure S11. Forest plots showing the pooled prevalence of antithrombotic agents in the 30 days pre-COVID (left) and 30 days post-COVID (right) in non-hospitalized pregnant women with COVID-19 (upper) and pregnant women without COVID-19 (lower), by pregnancy trimester

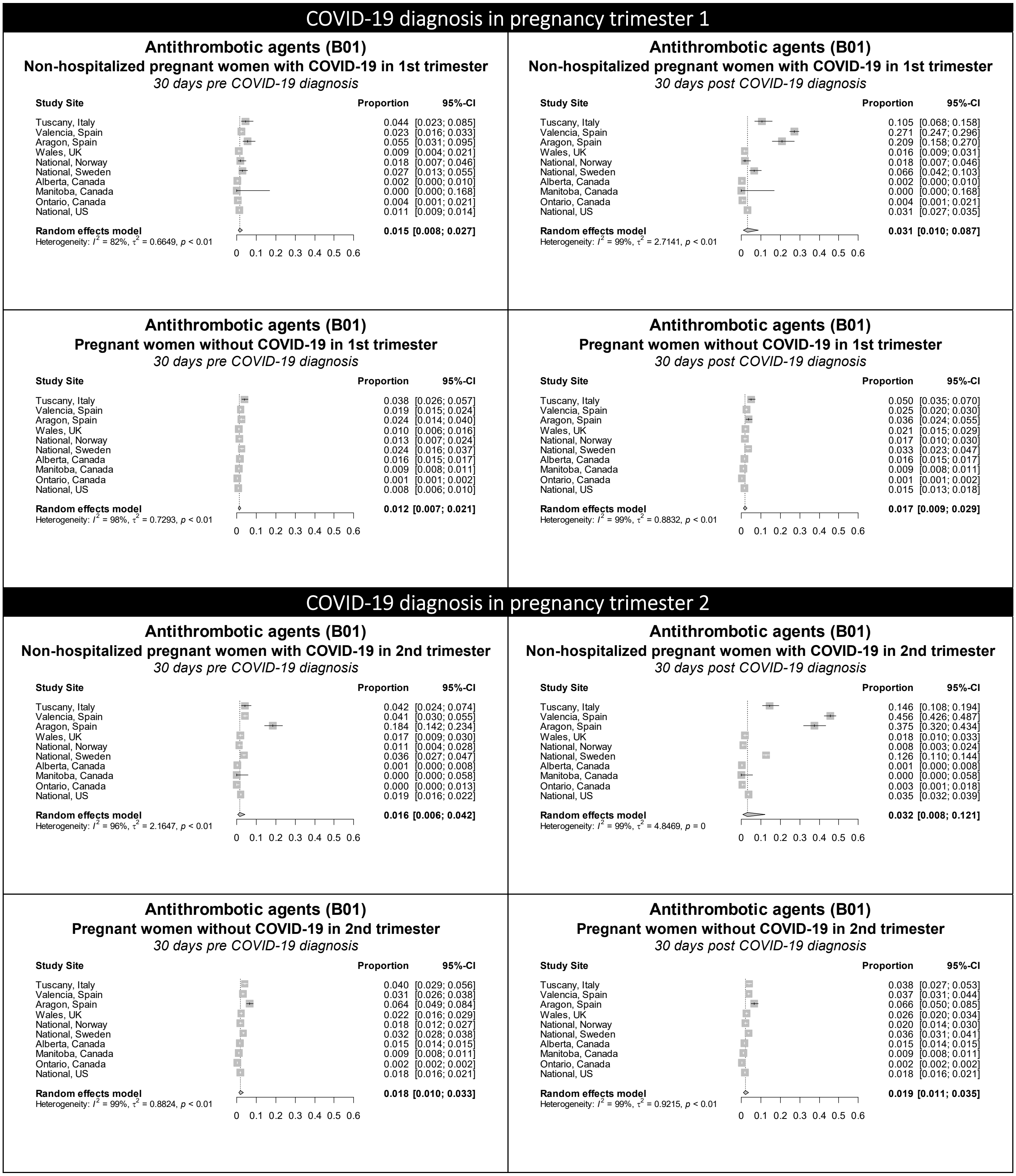

Figure S11 continued. Forest plots showing the pooled prevalence of antithrombotic agents in the 30 days pre-COVID (left) and 30 days post-COVID (right) in non-hospitalized pregnant women with COVID-19 (upper) and pregnant women without COVID-19 (lower), by pregnancy trimester

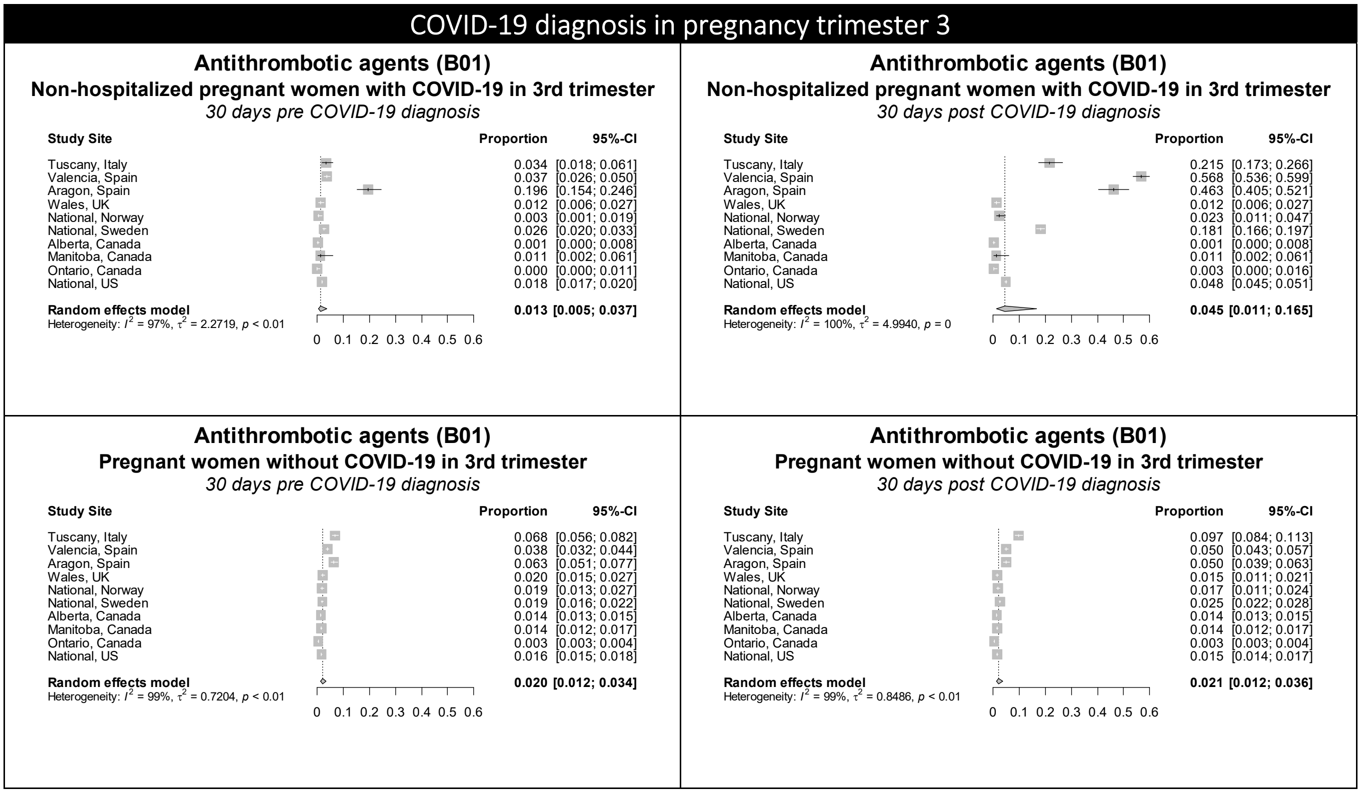

Figure S12. Forest plots showing the pooled prevalence of antivirals in the 30 days pre-COVID (left) and 30 days post-COVID (right) in non-hospitalized pregnant women with COVID-19 (upper) and pregnant women without COVID-19 (lower), by pregnancy trimester

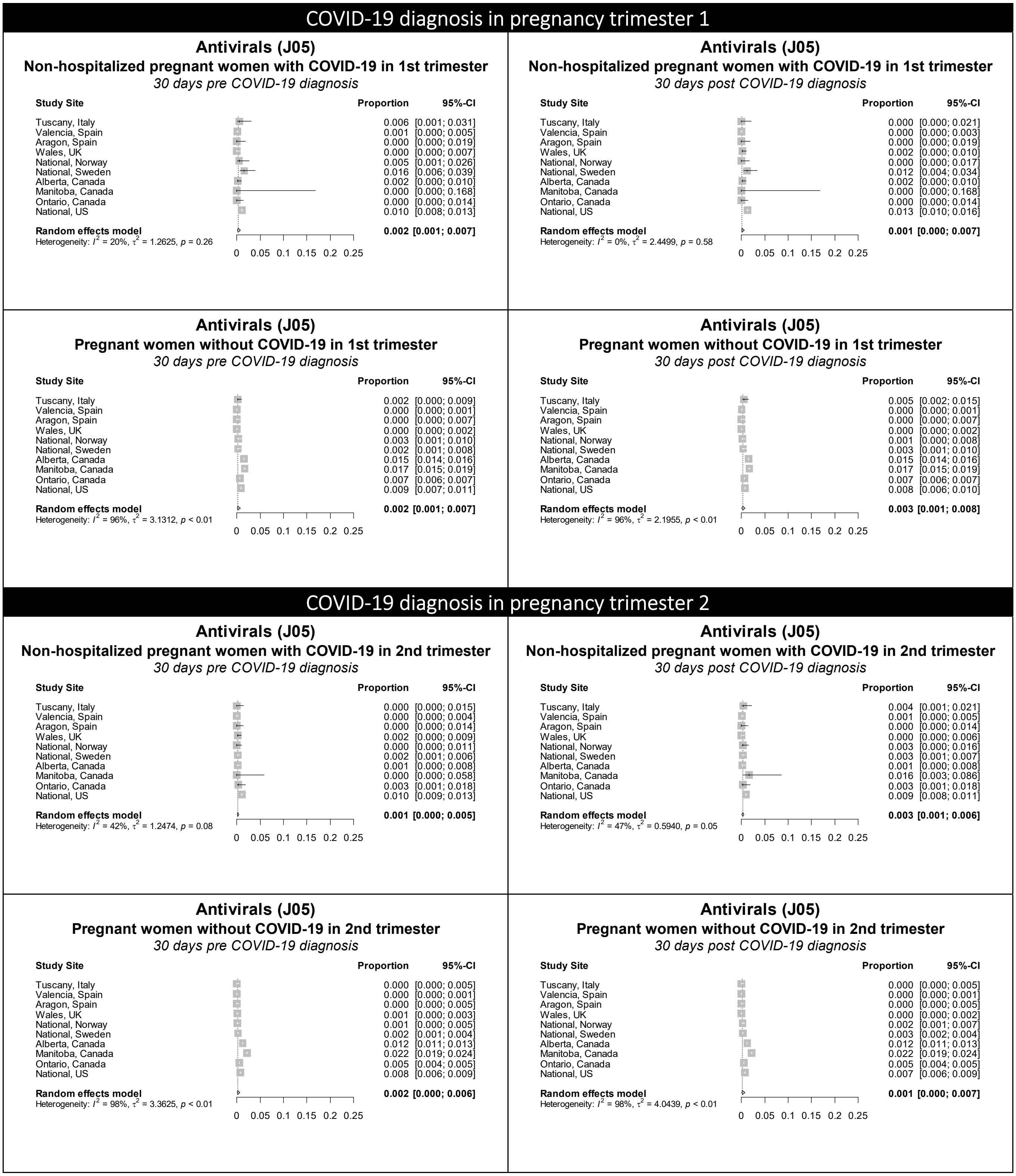

Figure S12 continued. Forest plots showing the pooled prevalence of antivirals in the 30 days pre-COVID (left) and 30 days post-COVID (right) in non-hospitalized pregnant women with COVID-19 (upper) and pregnant women without COVID-19 (lower), by pregnancy trimester

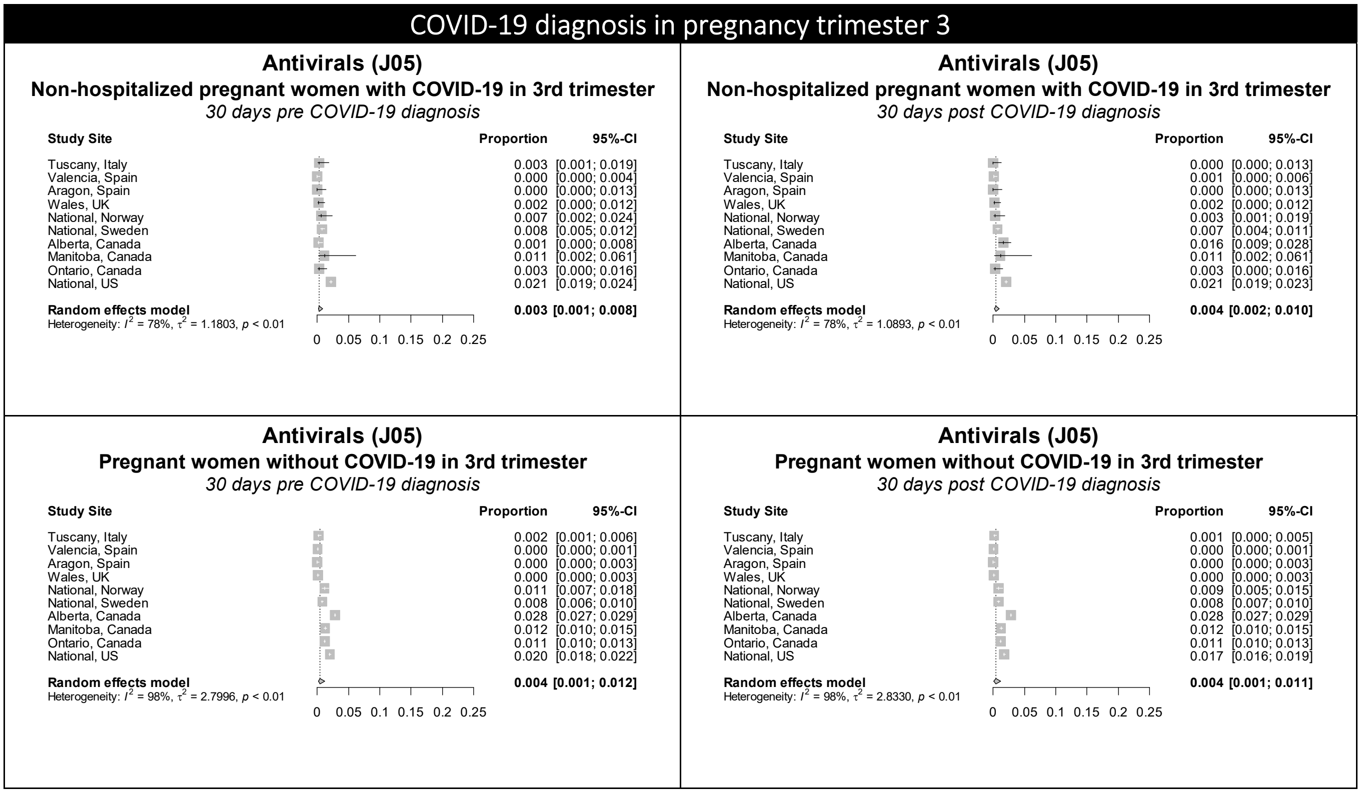

Figure S13. Forest plots showing the pooled prevalence of corticosteroids in the 30 days pre-COVID (left) and 30 days post-COVID (right) in non-hospitalized pregnant women with COVID-19 (upper) and pregnant women without COVID-19 (lower), by pregnancy trimester

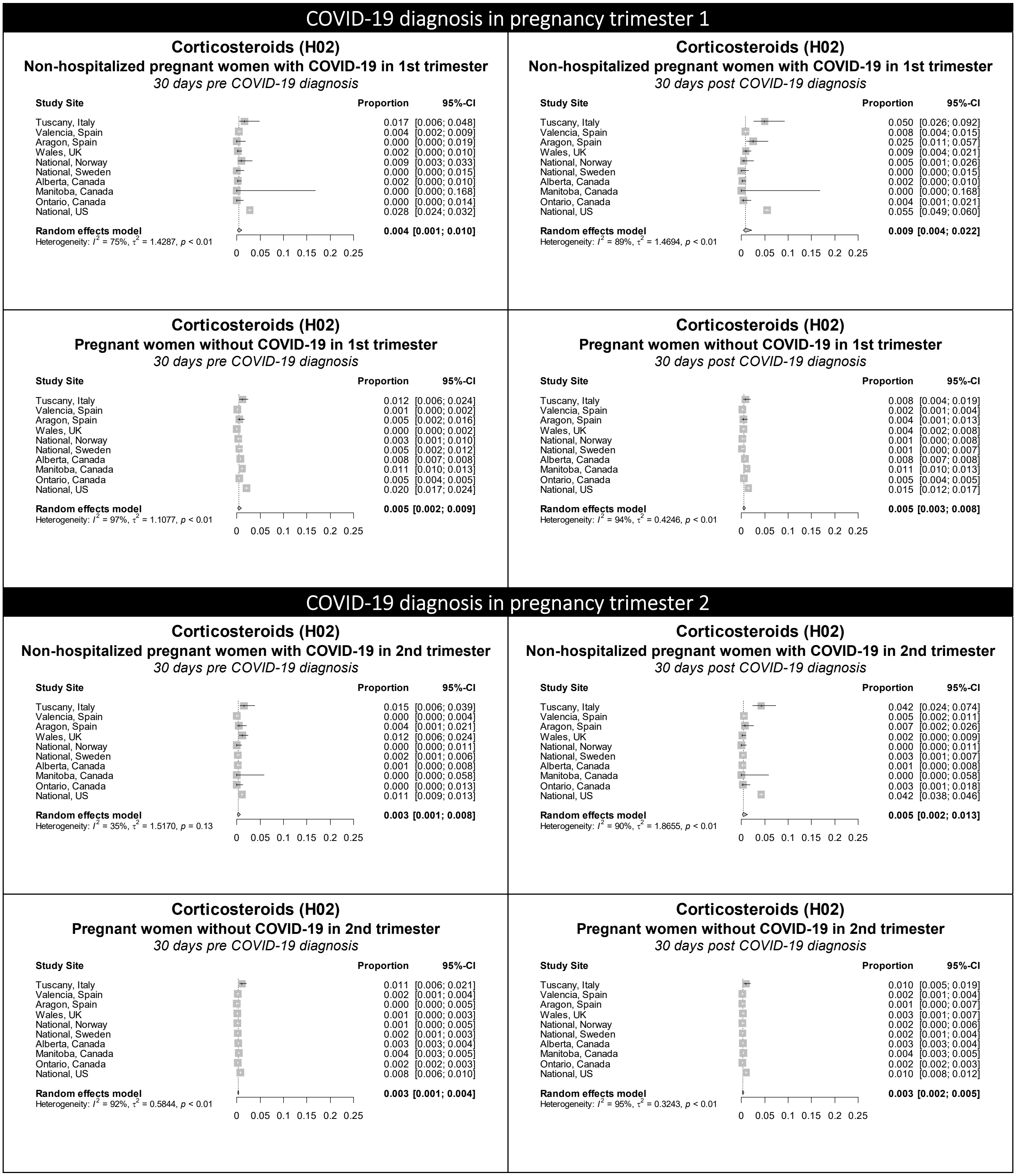

Figure S13 continued. Forest plots showing the pooled prevalence of corticosteroids in the 30 days pre-COVID (left) and 30 days post-COVID (right) in non-hospitalized pregnant women with COVID-19 (upper) and pregnant women without COVID-19 (lower), by pregnancy trimester

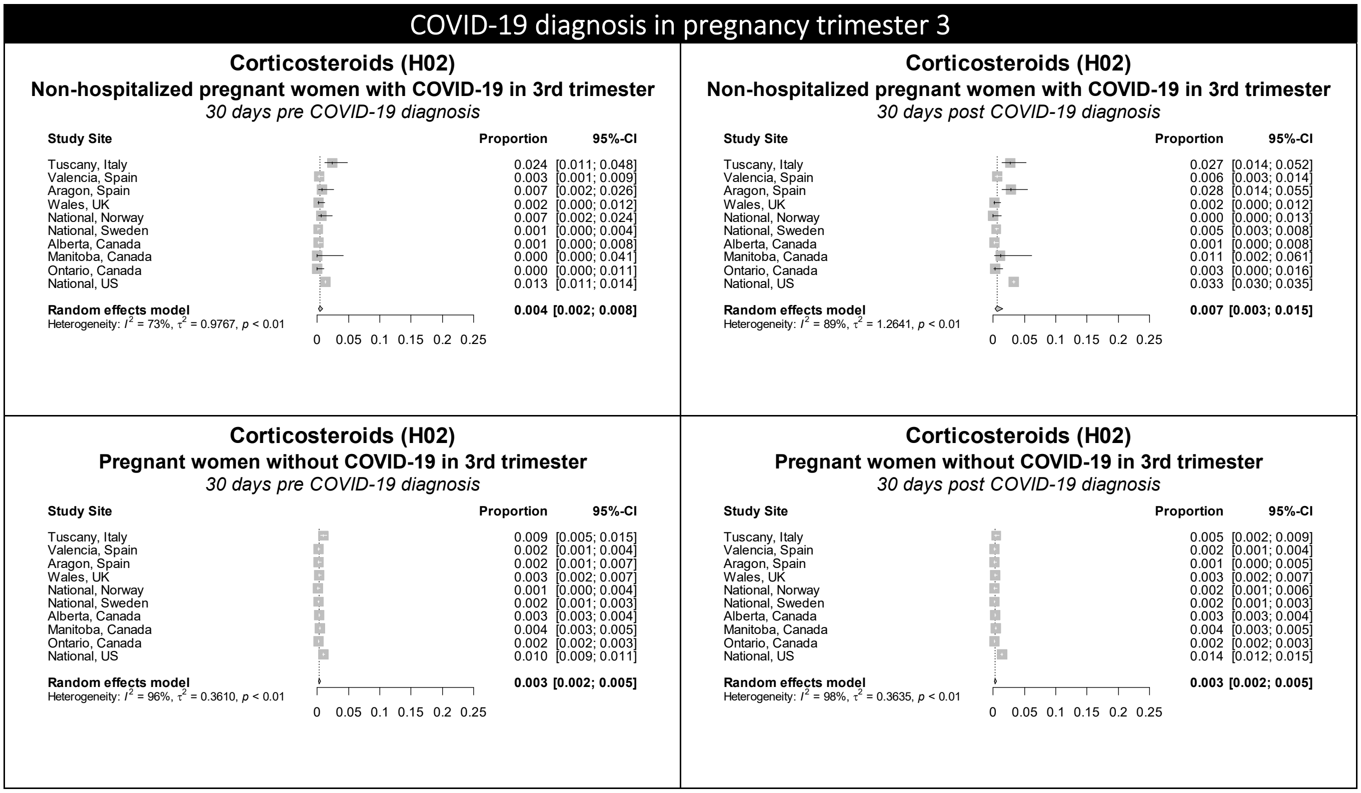

Figure S14. Forest plots showing the pooled prevalence of cough and cold preparations in the 30 days pre-COVID (left) and 30 days post-COVID (right) in non-hospitalized pregnant women with COVID-19 (upper) and pregnant women without COVID-19 (lower), by pregnancy trimester

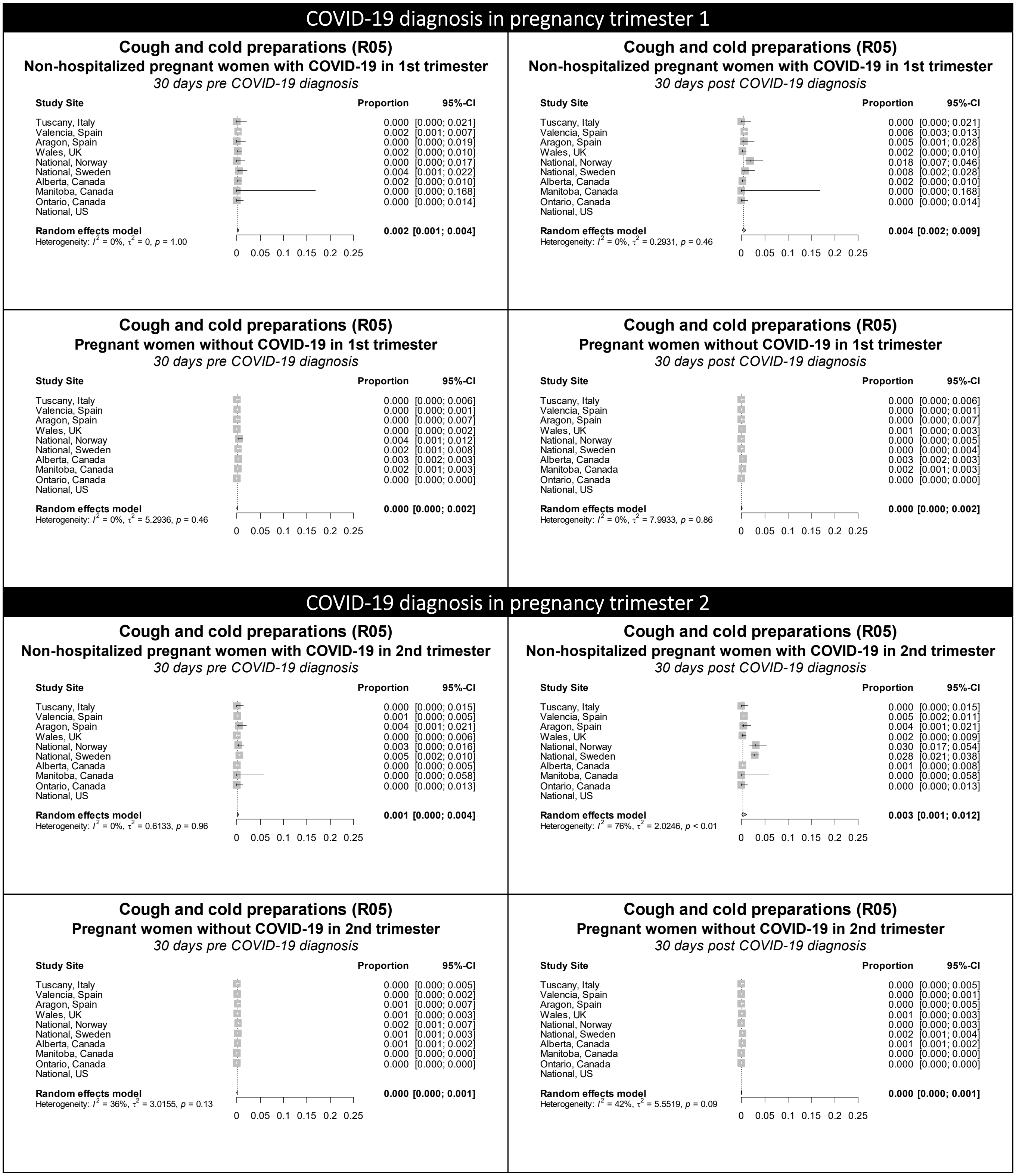

Figure S14 continued. Forest plots showing the pooled prevalence of cough and cold preparations in the 30 days pre-COVID (left) and 30 days post-COVID (right) in non-hospitalized pregnant women with COVID-19 (upper) and pregnant women without COVID-19 (lower), by pregnancy trimester

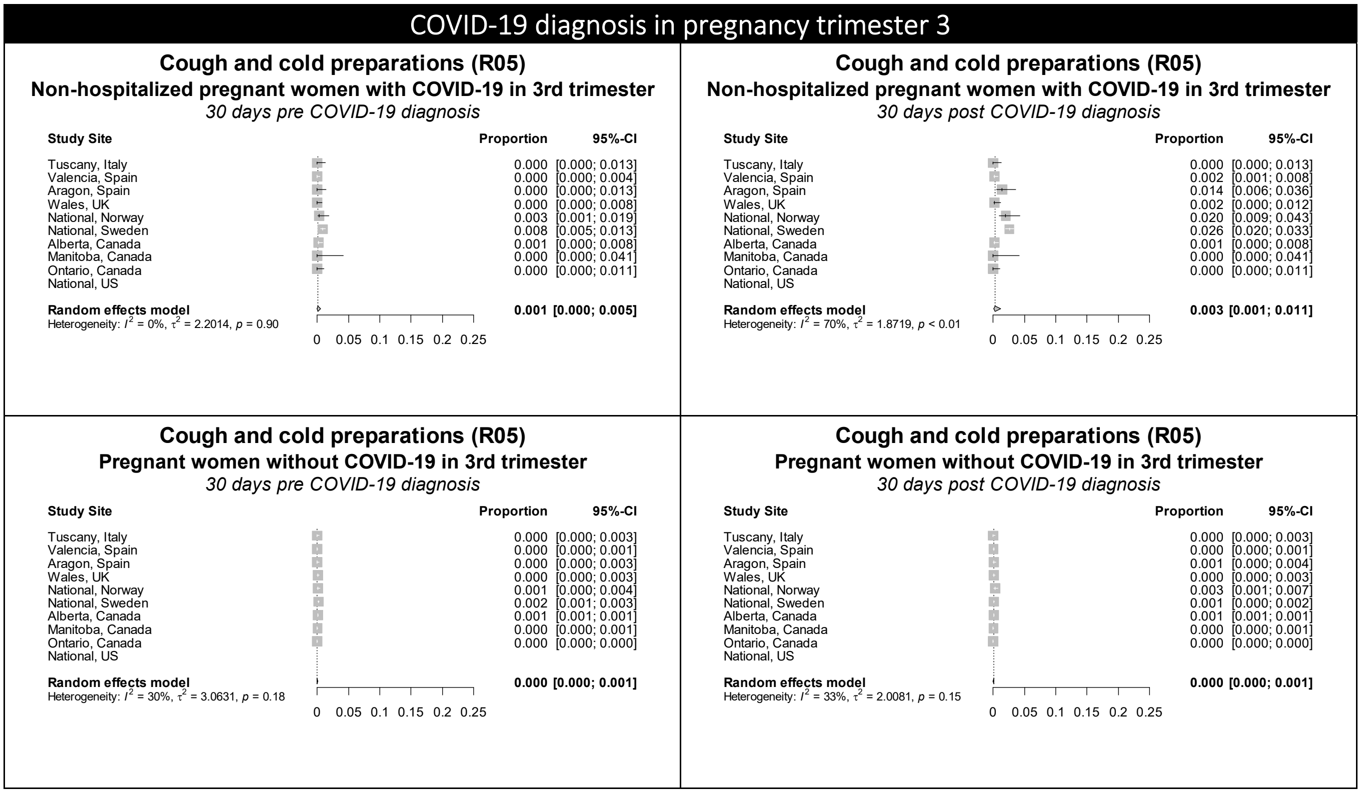

Figure S15. Forest plots showing the pooled prevalence of drugs for obstructive airway diseases in the 30 days pre-COVID (left) and 30 days post-COVID (right) in non-hospitalized pregnant women with COVID-19 (upper) and pregnant women without COVID-19 (lower), by pregnancy trimester

### COVID-19 diagnosis in pregnancy trimester 1

#### Drugs for obstructive airway diseases (R03)
**Non-hospitalized pregnant women with COVID-19 in 1st trimester**
*30 days pre COVID-19 diagnosis*

| Study Site | Proportion | 95%-CI |
|---|---|---|
| Tuscany, Italy | 0.006 | [0.001; 0.031] |
| Valencia, Spain | 0.018 | [0.012; 0.027] |
| Aragon, Spain | 0.015 | [0.005; 0.043] |
| Wales, UK | 0.022 | [0.012; 0.037] |
| National, Norway | 0.009 | [0.003; 0.033] |
| National, Sweden | 0.012 | [0.004; 0.034] |
| Alberta, Canada | 0.002 | [0.000; 0.010] |
| Manitoba, Canada | 0.053 | [0.009; 0.246] |
| Ontario, Canada | 0.000 | [0.000; 0.014] |
| National, US | 0.023 | [0.019; 0.027] |
| **Random effects model** | **0.011** | **[0.006; 0.020]** |

Heterogeneity: $I^2 = 33\%$, $\tau^2 = 0.4016$, $p = 0.15$

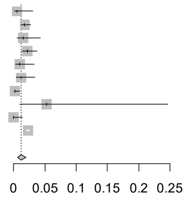

#### Drugs for obstructive airway diseases (R03)
**Non-hospitalized pregnant women with COVID-19 in 1st trimester**
*30 days post COVID-19 diagnosis*

| Study Site | Proportion | 95%-CI |
|---|---|---|
| Tuscany, Italy | 0.006 | [0.001; 0.031] |
| Valencia, Spain | 0.044 | [0.034; 0.057] |
| Aragon, Spain | 0.015 | [0.005; 0.043] |
| Wales, UK | 0.040 | [0.026; 0.059] |
| National, Norway | 0.005 | [0.001; 0.026] |
| National, Sweden | 0.023 | [0.011; 0.050] |
| Alberta, Canada | 0.028 | [0.017; 0.045] |
| Manitoba, Canada | 0.053 | [0.009; 0.246] |
| Ontario, Canada | 0.000 | [0.000; 0.014] |
| National, US | 0.059 | [0.054; 0.065] |
| **Random effects model** | **0.020** | **[0.010; 0.038]** |

Heterogeneity: $I^2 = 75\%$, $\tau^2 = 0.7584$, $p < 0.01$

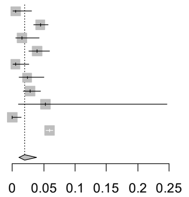

#### Drugs for obstructive airway diseases (R03)
**Pregnant women without COVID-19 in 1st trimester**
*30 days pre COVID-19 diagnosis*

| Study Site | Proportion | 95%-CI |
|---|---|---|
| Tuscany, Italy | 0.012 | [0.006; 0.024] |
| Valencia, Spain | 0.010 | [0.008; 0.014] |
| Aragon, Spain | 0.011 | [0.005; 0.024] |
| Wales, UK | 0.030 | [0.023; 0.039] |
| National, Norway | 0.014 | [0.008; 0.026] |
| National, Sweden | 0.010 | [0.006; 0.020] |
| Alberta, Canada | 0.035 | [0.033; 0.036] |
| Manitoba, Canada | 0.044 | [0.041; 0.047] |
| Ontario, Canada | 0.000 | [0.000; 0.000] |
| National, US | 0.015 | [0.013; 0.018] |
| **Random effects model** | **0.008** | **[0.002; 0.033]** |

Heterogeneity: $I^2 = 96\%$, $\tau^2 = 4.8485$, $p < 0.01$

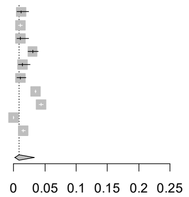

#### Drugs for obstructive airway diseases (R03)
**Pregnant women without COVID-19 in 1st trimester**
*30 days post COVID-19 diagnosis*

| Study Site | Proportion | 95%-CI |
|---|---|---|
| Tuscany, Italy | 0.005 | [0.002; 0.015] |
| Valencia, Spain | 0.010 | [0.007; 0.013] |
| Aragon, Spain | 0.011 | [0.005; 0.024] |
| Wales, UK | 0.029 | [0.022; 0.038] |
| National, Norway | 0.013 | [0.007; 0.024] |
| National, Sweden | 0.009 | [0.005; 0.018] |
| Alberta, Canada | 0.035 | [0.033; 0.036] |
| Manitoba, Canada | 0.044 | [0.041; 0.047] |
| Ontario, Canada | 0.000 | [0.000; 0.000] |
| National, US | 0.015 | [0.013; 0.018] |
| **Random effects model** | **0.007** | **[0.002; 0.029]** |

Heterogeneity: $I^2 = 96\%$, $\tau^2 = 4.9179$, $p < 0.01$

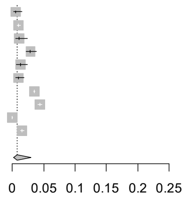

### COVID-19 diagnosis in pregnancy trimester 2

#### Drugs for obstructive airway diseases (R03)
**Non-hospitalized pregnant women with COVID-19 in 2nd trimester**
*30 days pre COVID-19 diagnosis*

| Study Site | Proportion | 95%-CI |
|---|---|---|
| Tuscany, Italy | 0.000 | [0.000; 0.015] |
| Valencia, Spain | 0.014 | [0.008; 0.023] |
| Aragon, Spain | 0.011 | [0.004; 0.032] |
| Wales, UK | 0.042 | [0.028; 0.061] |
| National, Norway | 0.006 | [0.002; 0.020] |
| National, Sweden | 0.008 | [0.005; 0.015] |
| Alberta, Canada | 0.017 | [0.010; 0.029] |
| Manitoba, Canada | 0.016 | [0.003; 0.086] |
| Ontario, Canada | 0.000 | [0.000; 0.013] |
| National, US | 0.019 | [0.016; 0.022] |
| **Random effects model** | **0.011** | **[0.006; 0.020]** |

Heterogeneity: $I^2 = 70\%$, $\tau^2 = 0.5360$, $p < 0.01$

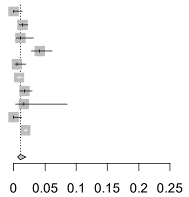

#### Drugs for obstructive airway diseases (R03)
**Non-hospitalized pregnant women with COVID-19 in 2nd trimester**
*30 days post COVID-19 diagnosis*

| Study Site | Proportion | 95%-CI |
|---|---|---|
| Tuscany, Italy | 0.015 | [0.006; 0.039] |
| Valencia, Spain | 0.039 | [0.029; 0.052] |
| Aragon, Spain | 0.040 | [0.023; 0.071] |
| Wales, UK | 0.047 | [0.033; 0.067] |
| National, Norway | 0.019 | [0.009; 0.039] |
| National, Sweden | 0.024 | [0.017; 0.033] |
| Alberta, Canada | 0.048 | [0.035; 0.067] |
| Manitoba, Canada | 0.016 | [0.003; 0.086] |
| Ontario, Canada | 0.000 | [0.000; 0.013] |
| National, US | 0.057 | [0.052; 0.061] |
| **Random effects model** | **0.028** | **[0.018; 0.044]** |

Heterogeneity: $I^2 = 80\%$, $\tau^2 = 0.3515$, $p < 0.01$

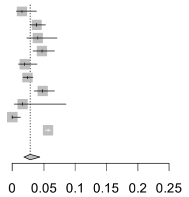

#### Drugs for obstructive airway diseases (R03)
**Pregnant women without COVID-19 in 2nd trimester**
*30 days pre COVID-19 diagnosis*

| Study Site | Proportion | 95%-CI |
|---|---|---|
| Tuscany, Italy | 0.004 | [0.001; 0.011] |
| Valencia, Spain | 0.014 | [0.010; 0.018] |
| Aragon, Spain | 0.005 | [0.002; 0.013] |
| Wales, UK | 0.019 | [0.014; 0.026] |
| National, Norway | 0.009 | [0.005; 0.016] |
| National, Sweden | 0.010 | [0.008; 0.013] |
| Alberta, Canada | 0.032 | [0.031; 0.033] |
| Manitoba, Canada | 0.023 | [0.020; 0.025] |
| Ontario, Canada | 0.000 | [0.000; 0.000] |
| National, US | 0.013 | [0.011; 0.015] |
| **Random effects model** | **0.006** | **[0.002; 0.022]** |

Heterogeneity: $I^2 = 97\%$, $\tau^2 = 4.4962$, $p < 0.01$

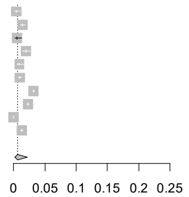

#### Drugs for obstructive airway diseases (R03)
**Pregnant women without COVID-19 in 2nd trimester**
*30 days post COVID-19 diagnosis*

| Study Site | Proportion | 95%-CI |
|---|---|---|
| Tuscany, Italy | 0.006 | [0.003; 0.014] |
| Valencia, Spain | 0.009 | [0.006; 0.013] |
| Aragon, Spain | 0.012 | [0.006; 0.022] |
| Wales, UK | 0.019 | [0.014; 0.026] |
| National, Norway | 0.007 | [0.004; 0.014] |
| National, Sweden | 0.012 | [0.009; 0.015] |
| Alberta, Canada | 0.032 | [0.031; 0.033] |
| Manitoba, Canada | 0.023 | [0.020; 0.025] |
| Ontario, Canada | 0.000 | [0.000; 0.000] |
| National, US | 0.016 | [0.013; 0.018] |
| **Random effects model** | **0.007** | **[0.002; 0.024]** |

Heterogeneity: $I^2 = 96\%$, $\tau^2 = 4.3878$, $p < 0.01$

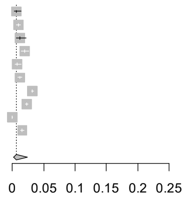

**Figure S15 continued.** Forest plots showing the pooled prevalence of drugs for obstructive airway diseases in the 30 days pre-COVID (left) and 30 days post-COVID (right) in non-hospitalized pregnant women with COVID-19 (upper) and pregnant women without COVID-19 (lower), by pregnancy trimester

### COVID-19 diagnosis in pregnancy trimester 3

#### Drugs for obstructive airway diseases (R03)
**Non-hospitalized pregnant women with COVID-19 in 3rd trimester**
*30 days pre COVID-19 diagnosis*

| Study Site | Proportion | 95%-CI |
|---|---|---|
| Tuscany, Italy | 0.010 | [0.003; 0.029] |
| Valencia, Spain | 0.011 | [0.006; 0.020] |
| Aragon, Spain | 0.025 | [0.012; 0.051] |
| Wales, UK | 0.027 | [0.016; 0.045] |
| National, Norway | 0.000 | [0.000; 0.013] |
| National, Sweden | 0.008 | [0.005; 0.013] |
| Alberta, Canada | 0.001 | [0.000; 0.008] |
| Manitoba, Canada | 0.011 | [0.002; 0.061] |
| Ontario, Canada | 0.000 | [0.000; 0.011] |
| National, US | 0.019 | [0.017; 0.021] |
| **Random effects model** | **0.008** | **[0.004; 0.017]** |

Heterogeneity: $I^2 = 66\%$, $\tau^2 = 0.7805$, $p < 0.01$

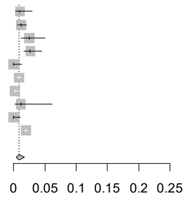

#### Drugs for obstructive airway diseases (R03)
**Non-hospitalized pregnant women with COVID-19 in 3rd trimester**
*30 days post COVID-19 diagnosis*

| Study Site | Proportion | 95%-CI |
|---|---|---|
| Tuscany, Italy | 0.000 | [0.000; 0.013] |
| Valencia, Spain | 0.044 | [0.033; 0.059] |
| Aragon, Spain | 0.043 | [0.025; 0.073] |
| Wales, UK | 0.033 | [0.020; 0.053] |
| National, Norway | 0.007 | [0.002; 0.024] |
| National, Sweden | 0.014 | [0.010; 0.020] |
| Alberta, Canada | 0.041 | [0.028; 0.058] |
| Manitoba, Canada | 0.011 | [0.002; 0.061] |
| Ontario, Canada | 0.000 | [0.000; 0.011] |
| National, US | 0.041 | [0.038; 0.044] |
| **Random effects model** | **0.016** | **[0.007; 0.035]** |

Heterogeneity: $I^2 = 80\%$, $\tau^2 = 1.1816$, $p < 0.01$

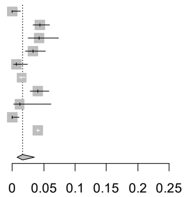

#### Drugs for obstructive airway diseases (R03)
**Pregnant women without COVID-19 in 3rd trimester**
*30 days pre COVID-19 diagnosis*

| Study Site | Proportion | 95%-CI |
|---|---|---|
| Tuscany, Italy | 0.007 | [0.004; 0.013] |
| Valencia, Spain | 0.010 | [0.007; 0.013] |
| Aragon, Spain | 0.007 | [0.004; 0.013] |
| Wales, UK | 0.023 | [0.017; 0.030] |
| National, Norway | 0.011 | [0.007; 0.018] |
| National, Sweden | 0.008 | [0.007; 0.011] |
| Alberta, Canada | 0.029 | [0.028; 0.031] |
| Manitoba, Canada | 0.034 | [0.030; 0.037] |
| Ontario, Canada | 0.000 | [0.000; 0.000] |
| National, US | 0.015 | [0.014; 0.017] |
| **Random effects model** | **0.007** | **[0.002; 0.024]** |

Heterogeneity: $I^2 = 97\%$, $\tau^2 = 3.6360$, $p < 0.01$

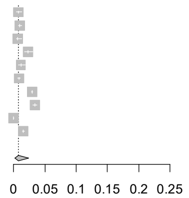

#### Drugs for obstructive airway diseases (R03)
**Pregnant women without COVID-19 in 3rd trimester**
*30 days post COVID-19 diagnosis*

| Study Site | Proportion | 95%-CI |
|---|---|---|
| Tuscany, Italy | 0.008 | [0.005; 0.014] |
| Valencia, Spain | 0.010 | [0.007; 0.013] |
| Aragon, Spain | 0.009 | [0.005; 0.016] |
| Wales, UK | 0.024 | [0.018; 0.031] |
| National, Norway | 0.009 | [0.006; 0.016] |
| National, Sweden | 0.008 | [0.007; 0.010] |
| Alberta, Canada | 0.029 | [0.028; 0.031] |
| Manitoba, Canada | 0.034 | [0.030; 0.037] |
| Ontario, Canada | 0.000 | [0.000; 0.000] |
| National, US | 0.013 | [0.012; 0.015] |
| **Random effects model** | **0.008** | **[0.002; 0.024]** |

Heterogeneity: $I^2 = 98\%$, $\tau^2 = 3.5975$, $p < 0.01$

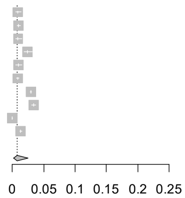

Figure S16. Forest plots showing the pooled prevalence of drugs used in diabetes in the 30 days pre-COVID (left) and 30 days post-COVID (right) in non-hospitalized pregnant women with COVID-19 (upper) and pregnant women without COVID-19 (lower), by pregnancy trimester

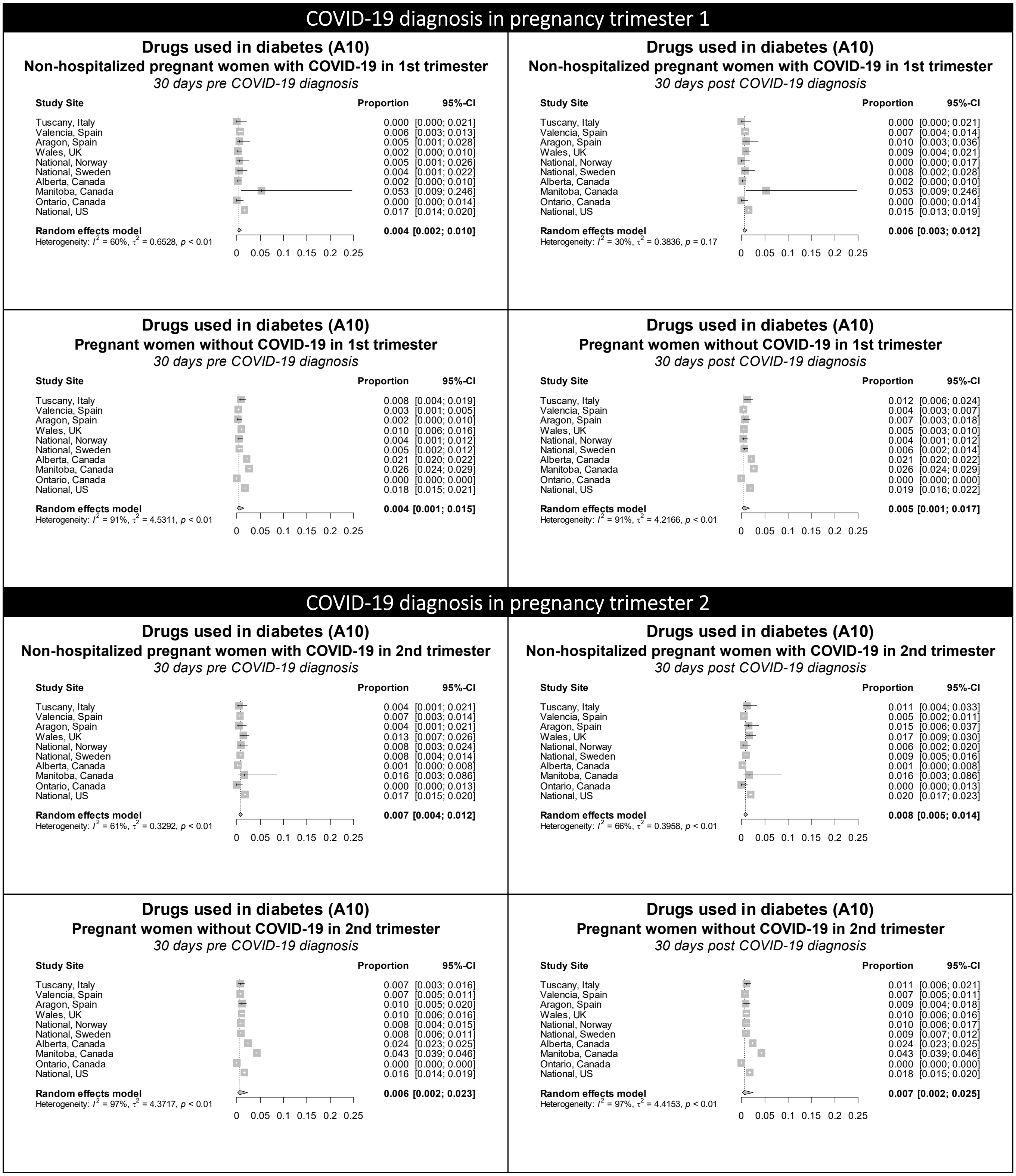

**Figure S16 continued.** Forest plots showing the pooled prevalence of drugs used in diabetes in the 30 days pre-COVID (left) and 30 days post-COVID (right) in non-hospitalized pregnant women with COVID-19 (upper) and pregnant women without COVID-19 (lower), by pregnancy trimester

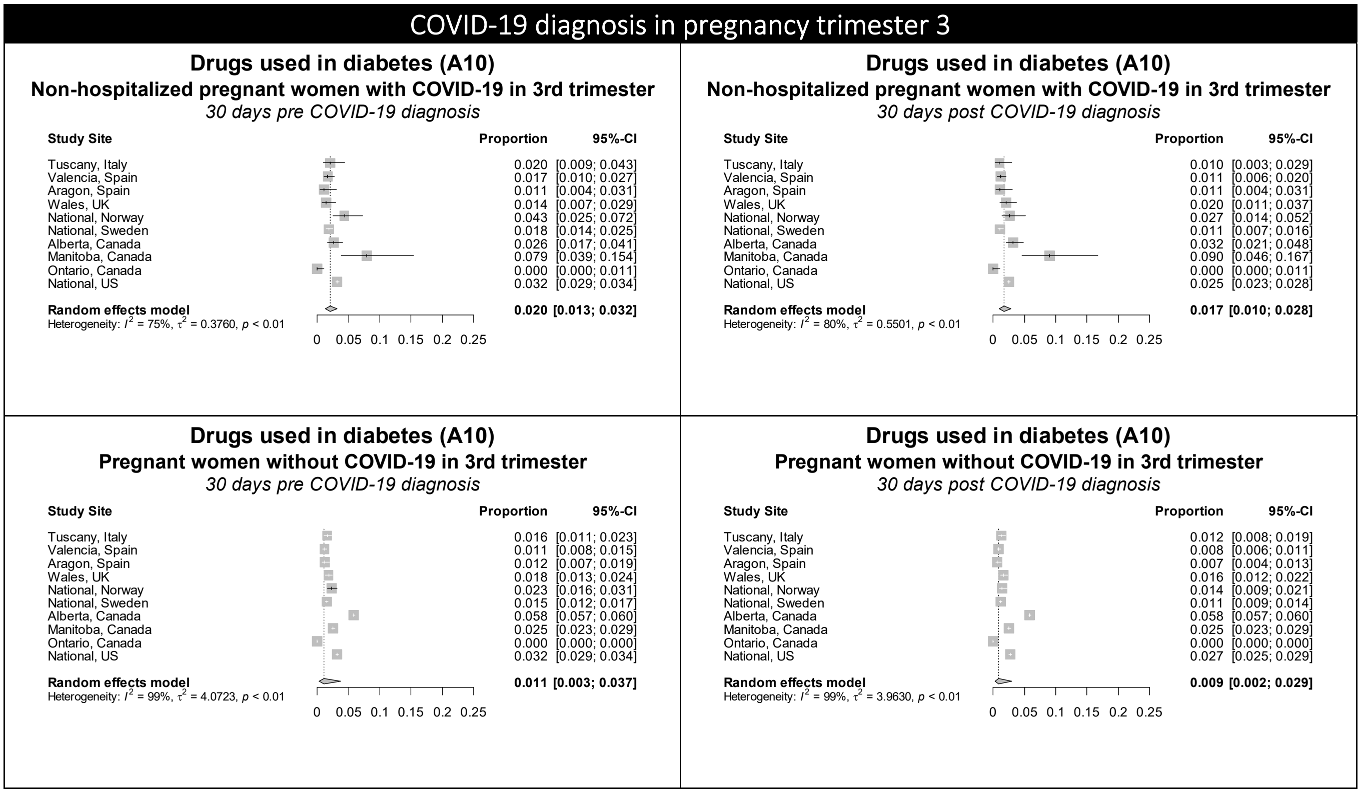

Figure S17. Forest plots showing the pooled prevalence of immune sera and immunoglobulins in the 30 days pre-COVID (left) and 30 days post-COVID (right) in non-hospitalized pregnant women with COVID-19 (upper) and pregnant women without COVID-19 (lower), by pregnancy trimester

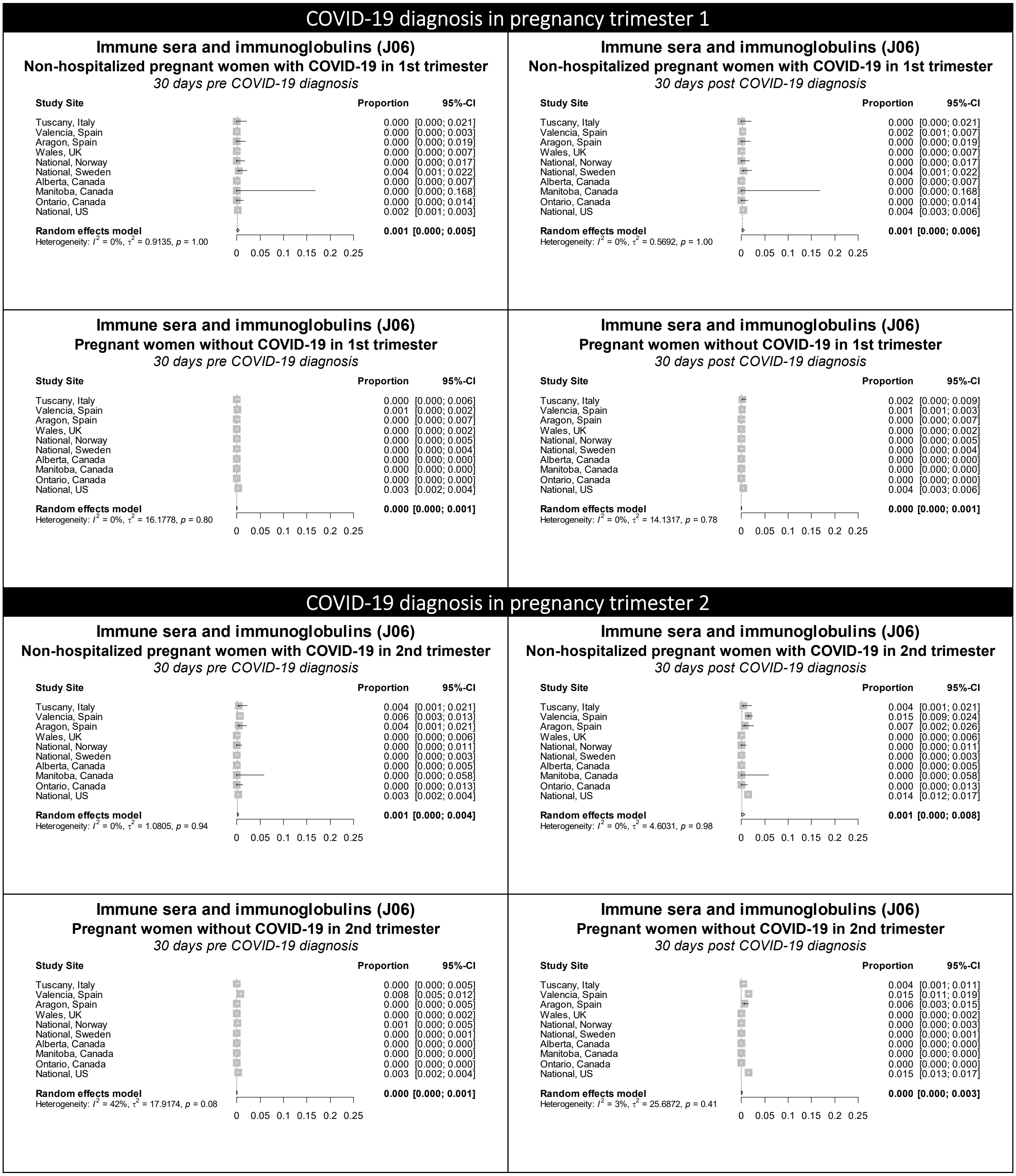

Figure S17 continued. Forest plots showing the pooled prevalence of immune sera and immunoglobulins in the 30 days pre-COVID (left) and 30 days post-COVID (right) in non-hospitalized pregnant women with COVID-19 (upper) and pregnant women without COVID-19 (lower), by pregnancy trimester

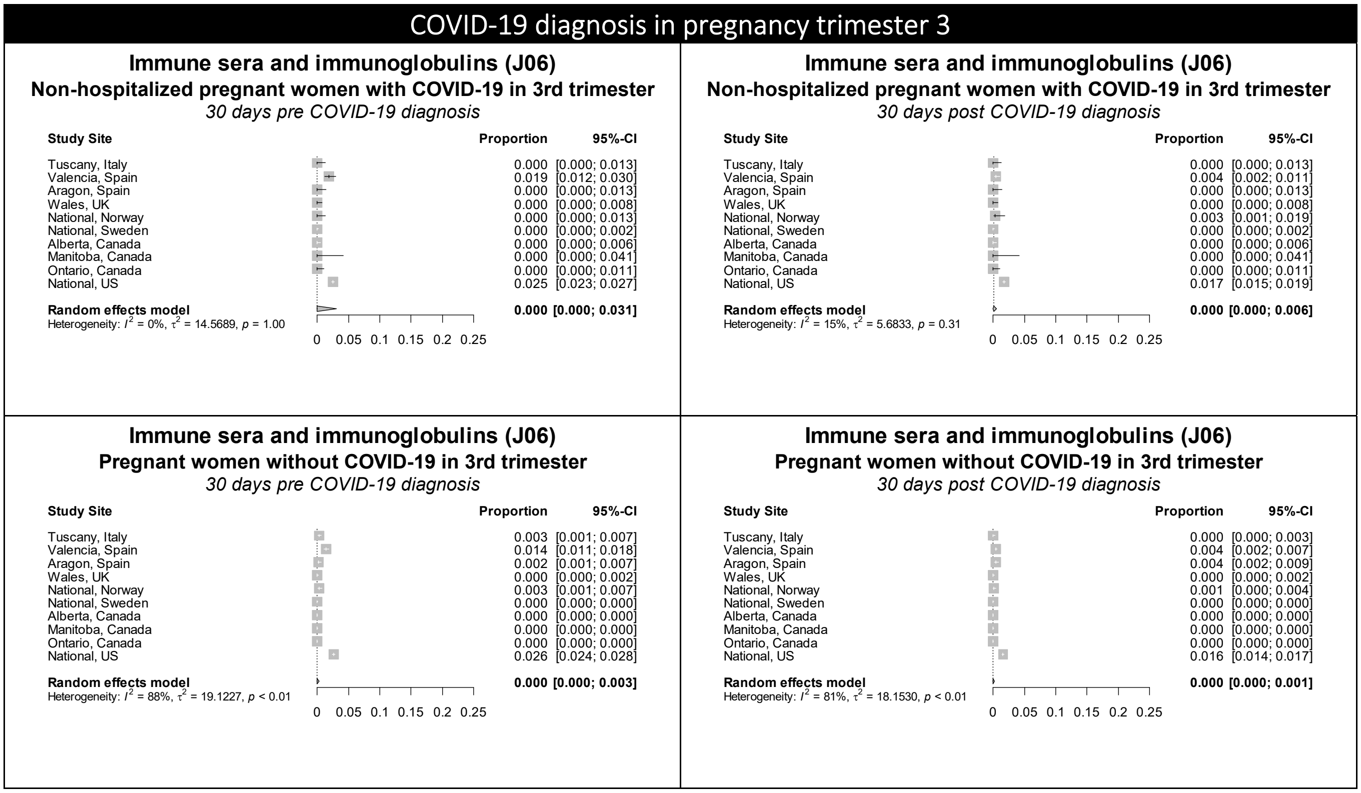

Figure S18. Forest plots showing the pooled prevalence of immunostimulants in the 30 days pre-COVID (left) and 30 days post-COVID (right) in non-hospitalized pregnant women with COVID-19 (upper) and pregnant women without COVID-19 (lower), by pregnancy trimester

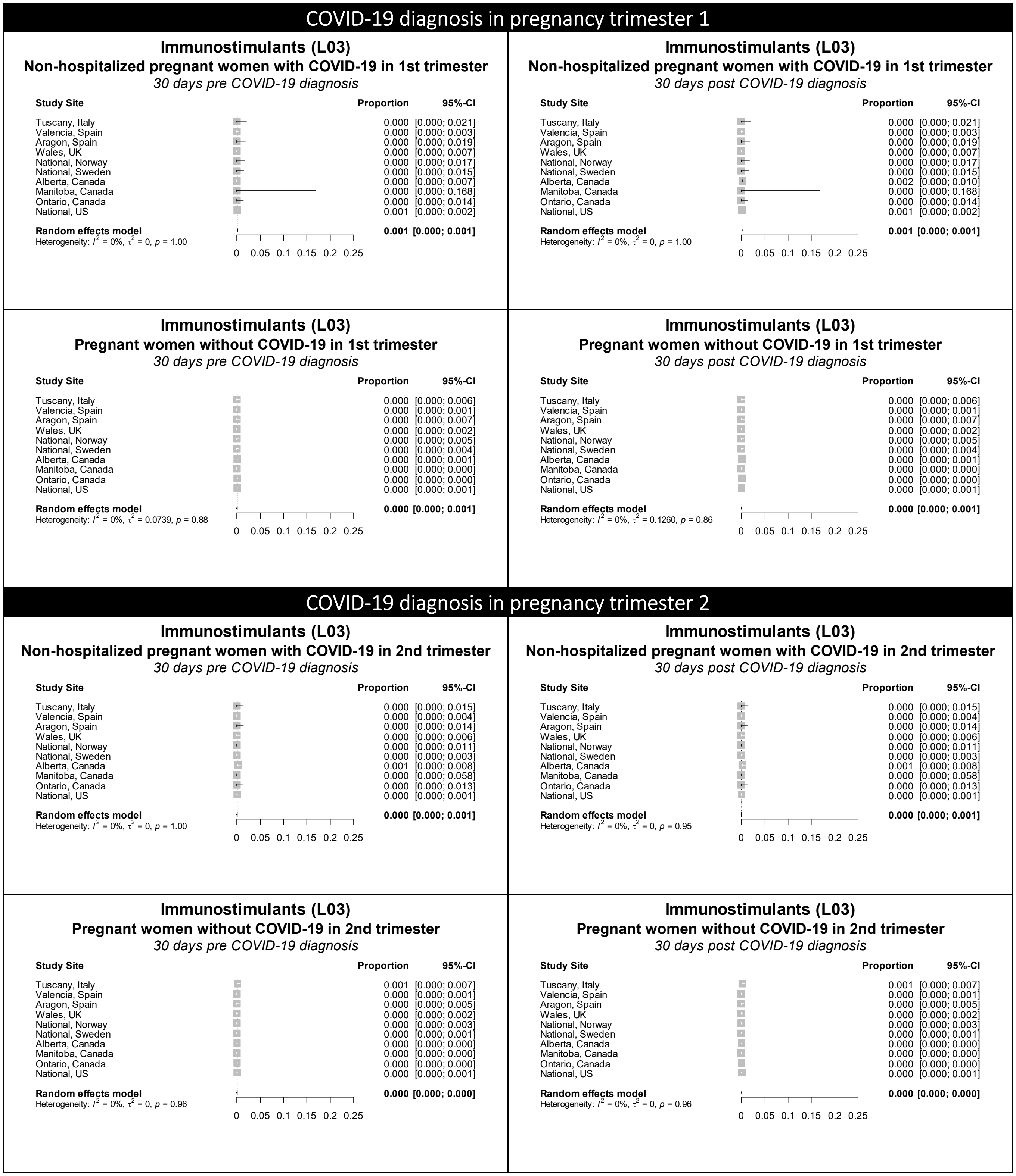

Figure S18 continued. Forest plots showing the pooled prevalence of immunostimulants in the 30 days pre-COVID (left) and 30 days post-COVID (right) in non-hospitalized pregnant women with COVID-19 (upper) and pregnant women without COVID-19 (lower), by pregnancy trimester

### COVID-19 diagnosis in pregnancy trimester 3

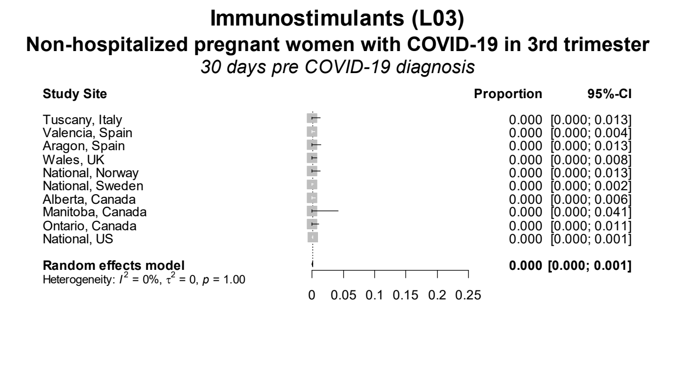

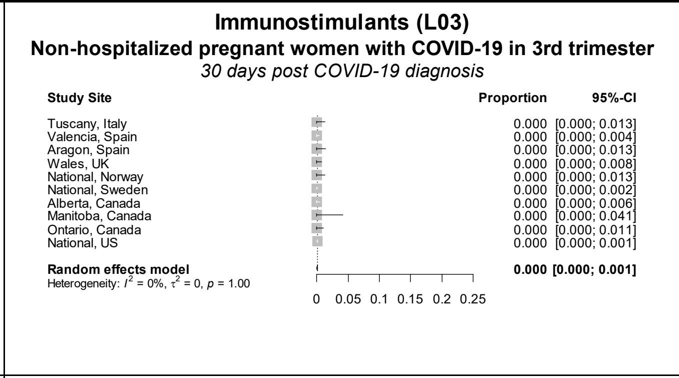

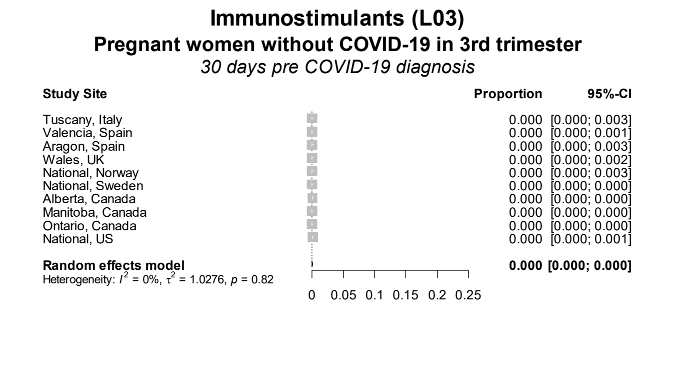

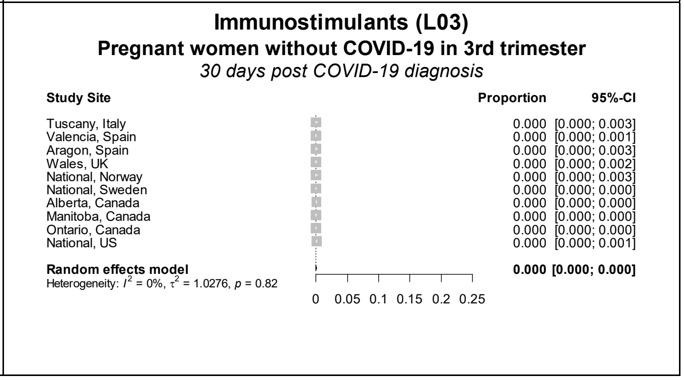

Figure S19. Forest plots showing the pooled prevalence of immunosuppressants in the 30 days pre-COVID (left) and 30 days post-COVID (right) in non-hospitalized pregnant women with COVID-19 (upper) and pregnant women without COVID-19 (lower), by pregnancy trimester

### COVID-19 diagnosis in pregnancy trimester 1

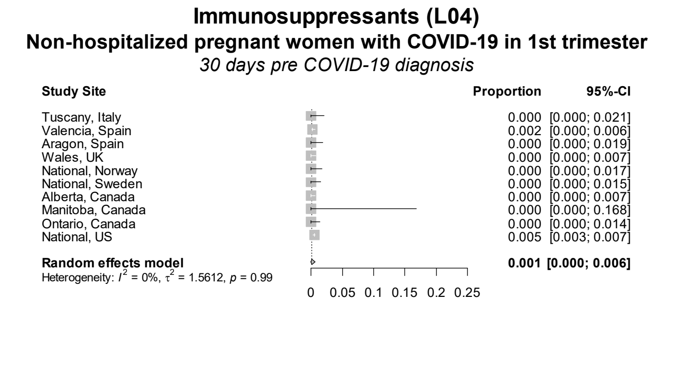

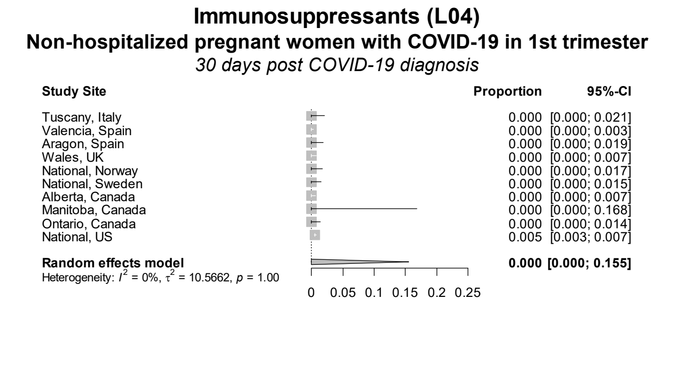

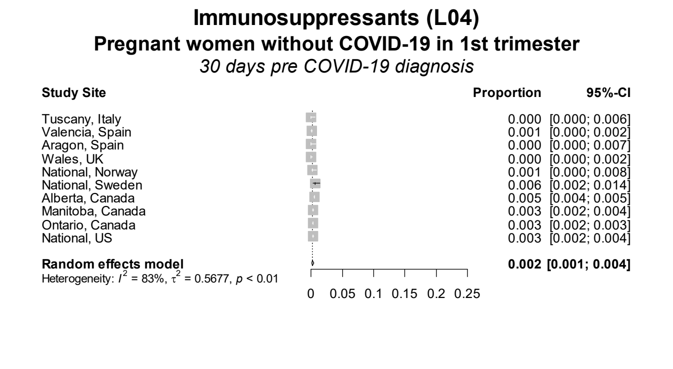

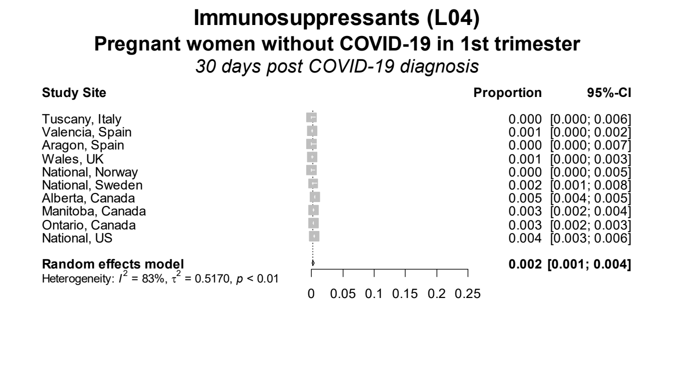

### COVID-19 diagnosis in pregnancy trimester 2

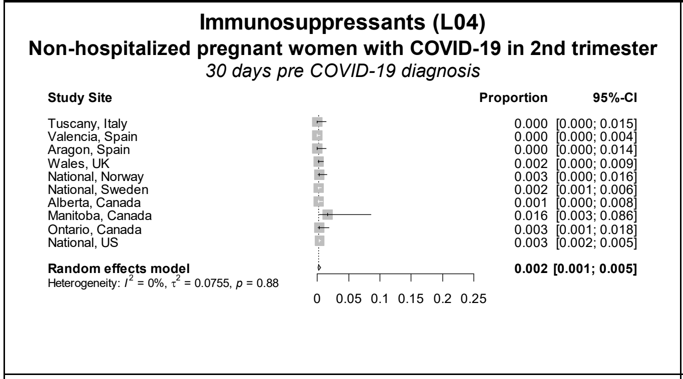

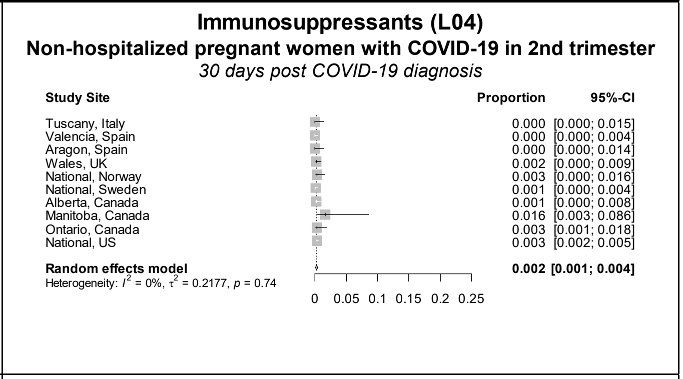

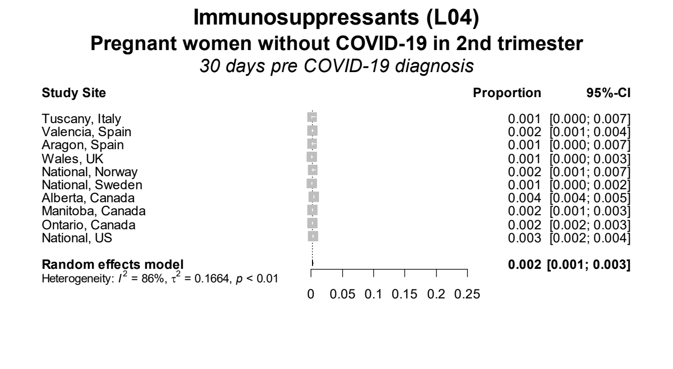

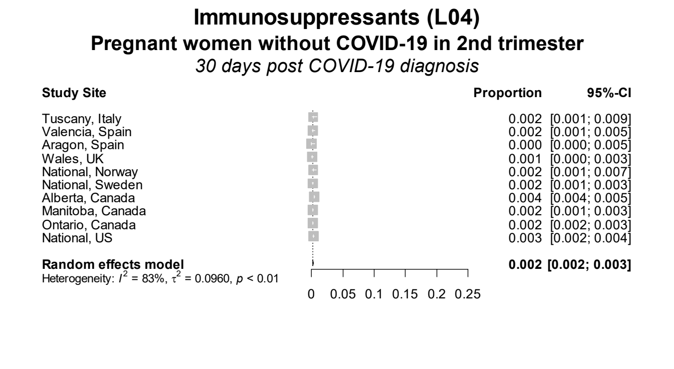

**Figure S19 continued.** Forest plots showing the pooled prevalence of immunosuppressants in the 30 days pre-COVID (left) and 30 days post-COVID (right) in non-hospitalized pregnant women with COVID-19 (upper) and pregnant women without COVID-19 (lower), by pregnancy trimester

### COVID-19 diagnosis in pregnancy trimester 3

#### Immunosuppressants (L04)
**Non-hospitalized pregnant women with COVID-19 in 3rd trimester**
*30 days pre COVID-19 diagnosis*

| Study Site | Proportion | 95%-CI |
|---|---|---|
| Tuscany, Italy | 0.000 | [0.000; 0.013] |
| Valencia, Spain | 0.001 | [0.000; 0.006] |
| Aragon, Spain | 0.004 | [0.001; 0.020] |
| Wales, UK | 0.000 | [0.000; 0.008] |
| National, Norway | 0.000 | [0.000; 0.013] |
| National, Sweden | 0.001 | [0.000; 0.003] |
| Alberta, Canada | 0.001 | [0.000; 0.008] |
| Manitoba, Canada | 0.000 | [0.000; 0.041] |
| Ontario, Canada | 0.003 | [0.000; 0.016] |
| National, US | 0.003 | [0.002; 0.004] |
| **Random effects model** | **0.002** | **[0.001; 0.004]** |

Heterogeneity: $I^2 = 0\%$, $\tau^2 = 0.1330$, $p = 0.92$

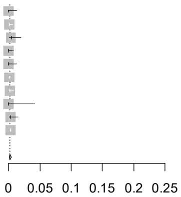

#### Immunosuppressants (L04)
**Non-hospitalized pregnant women with COVID-19 in 3rd trimester**
*30 days post COVID-19 diagnosis*

| Study Site | Proportion | 95%-CI |
|---|---|---|
| Tuscany, Italy | 0.000 | [0.000; 0.013] |
| Valencia, Spain | 0.001 | [0.000; 0.006] |
| Aragon, Spain | 0.000 | [0.000; 0.013] |
| Wales, UK | 0.000 | [0.000; 0.008] |
| National, Norway | 0.000 | [0.000; 0.013] |
| National, Sweden | 0.001 | [0.000; 0.004] |
| Alberta, Canada | 0.001 | [0.000; 0.008] |
| Manitoba, Canada | 0.000 | [0.000; 0.041] |
| Ontario, Canada | 0.003 | [0.000; 0.016] |
| National, US | 0.003 | [0.002; 0.004] |
| **Random effects model** | **0.002** | **[0.001; 0.004]** |

Heterogeneity: $I^2 = 0\%$, $\tau^2 = 0.2181$, $p = 0.91$

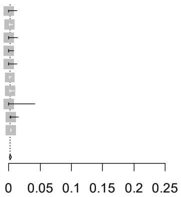

#### Immunosuppressants (L04)
**Pregnant women without COVID-19 in 3rd trimester**
*30 days pre COVID-19 diagnosis*

| Study Site | Proportion | 95%-CI |
|---|---|---|
| Tuscany, Italy | 0.001 | [0.000; 0.005] |
| Valencia, Spain | 0.001 | [0.000; 0.003] |
| Aragon, Spain | 0.000 | [0.000; 0.003] |
| Wales, UK | 0.000 | [0.000; 0.003] |
| National, Norway | 0.001 | [0.000; 0.004] |
| National, Sweden | 0.001 | [0.001; 0.002] |
| Alberta, Canada | 0.004 | [0.003; 0.004] |
| Manitoba, Canada | 0.002 | [0.001; 0.003] |
| Ontario, Canada | 0.002 | [0.002; 0.003] |
| National, US | 0.003 | [0.002; 0.003] |
| **Random effects model** | **0.002** | **[0.001; 0.003]** |

Heterogeneity: $I^2 = 82\%$, $\tau^2 = 0.2134$, $p < 0.01$

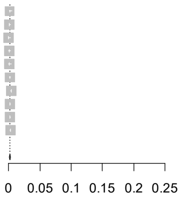

#### Immunosuppressants (L04)
**Pregnant women without COVID-19 in 3rd trimester**
*30 days post COVID-19 diagnosis*

| Study Site | Proportion | 95%-CI |
|---|---|---|
| Tuscany, Italy | 0.003 | [0.001; 0.007] |
| Valencia, Spain | 0.001 | [0.000; 0.003] |
| Aragon, Spain | 0.000 | [0.000; 0.003] |
| Wales, UK | 0.000 | [0.000; 0.003] |
| National, Norway | 0.000 | [0.000; 0.003] |
| National, Sweden | 0.001 | [0.001; 0.002] |
| Alberta, Canada | 0.004 | [0.003; 0.004] |
| Manitoba, Canada | 0.002 | [0.001; 0.003] |
| Ontario, Canada | 0.002 | [0.002; 0.003] |
| National, US | 0.003 | [0.002; 0.004] |
| **Random effects model** | **0.002** | **[0.001; 0.003]** |

Heterogeneity: $I^2 = 80\%$, $\tau^2 = 0.2916$, $p < 0.01$

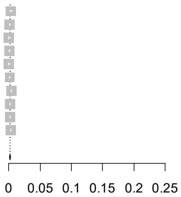

Figure S20. Forest plots showing the pooled prevalence of nasal preparations in the 30 days pre-COVID (left) and 30 days post-COVID (right) in non-hospitalized pregnant women with COVID-19 (upper) and pregnant women without COVID-19 (lower), by pregnancy trimester

### COVID-19 diagnosis in pregnancy trimester 1

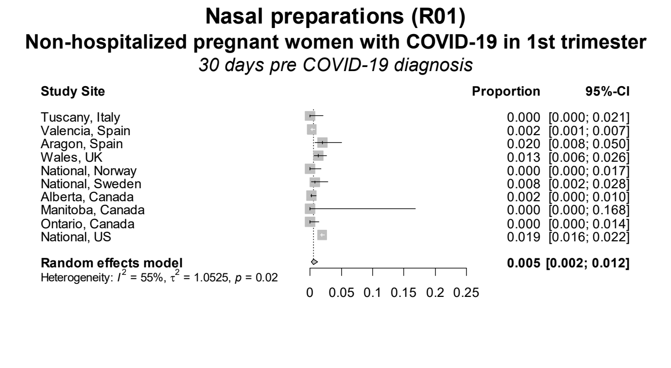

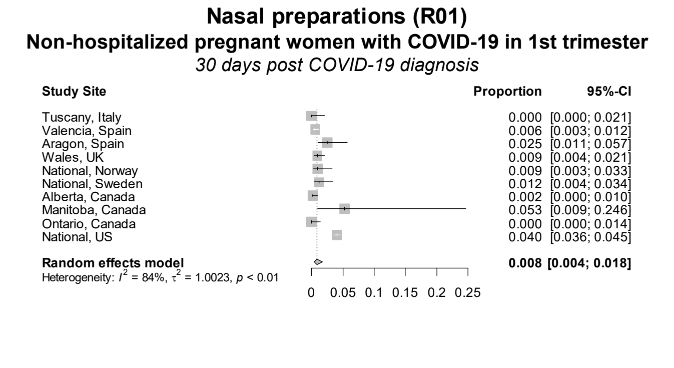

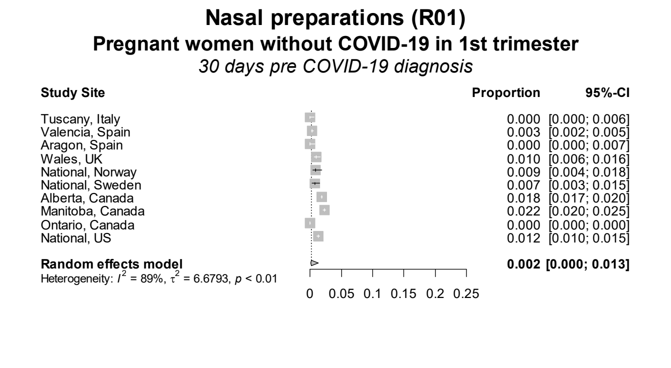

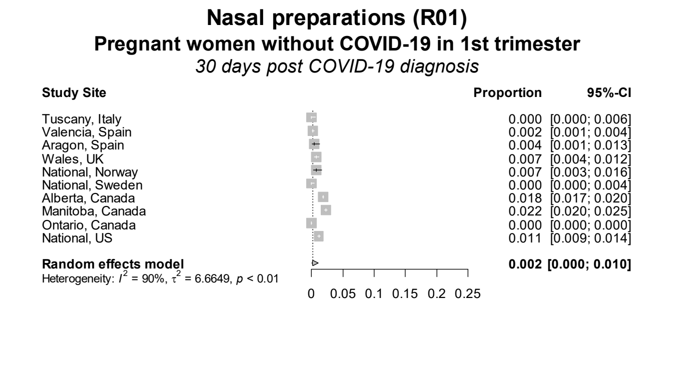

### COVID-19 diagnosis in pregnancy trimester 2

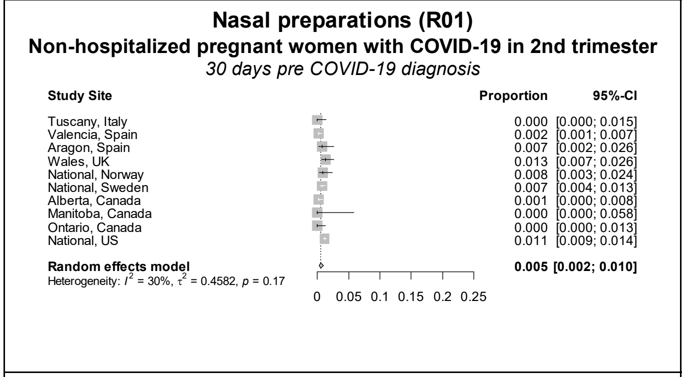

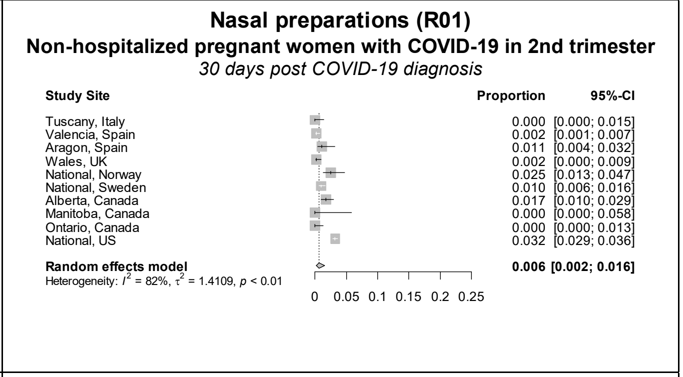

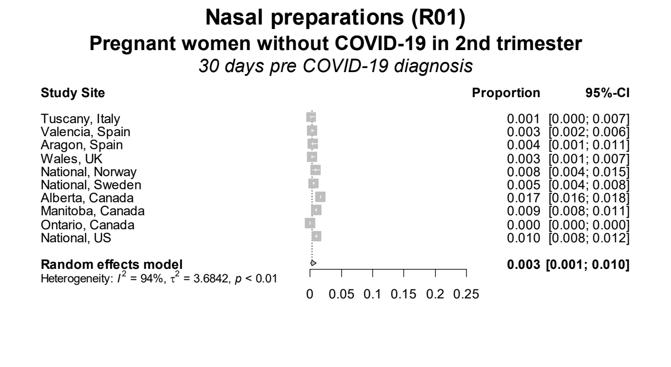

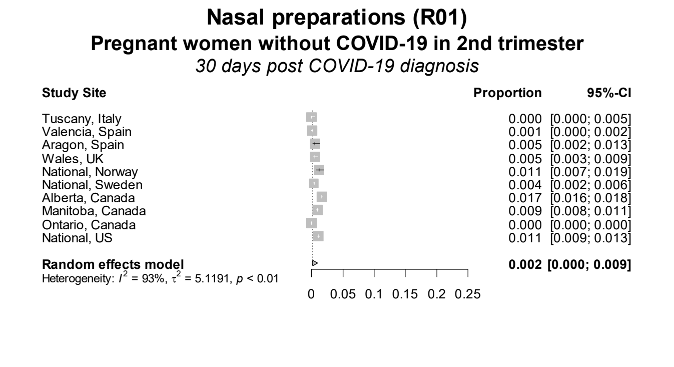

Figure S20 continued. Forest plots showing the pooled prevalence of nasal preparations in the 30 days pre-COVID (left) and 30 days post-COVID (right) in non-hospitalized pregnant women with COVID-19 (upper) and pregnant women without COVID-19 (lower), by pregnancy trimester

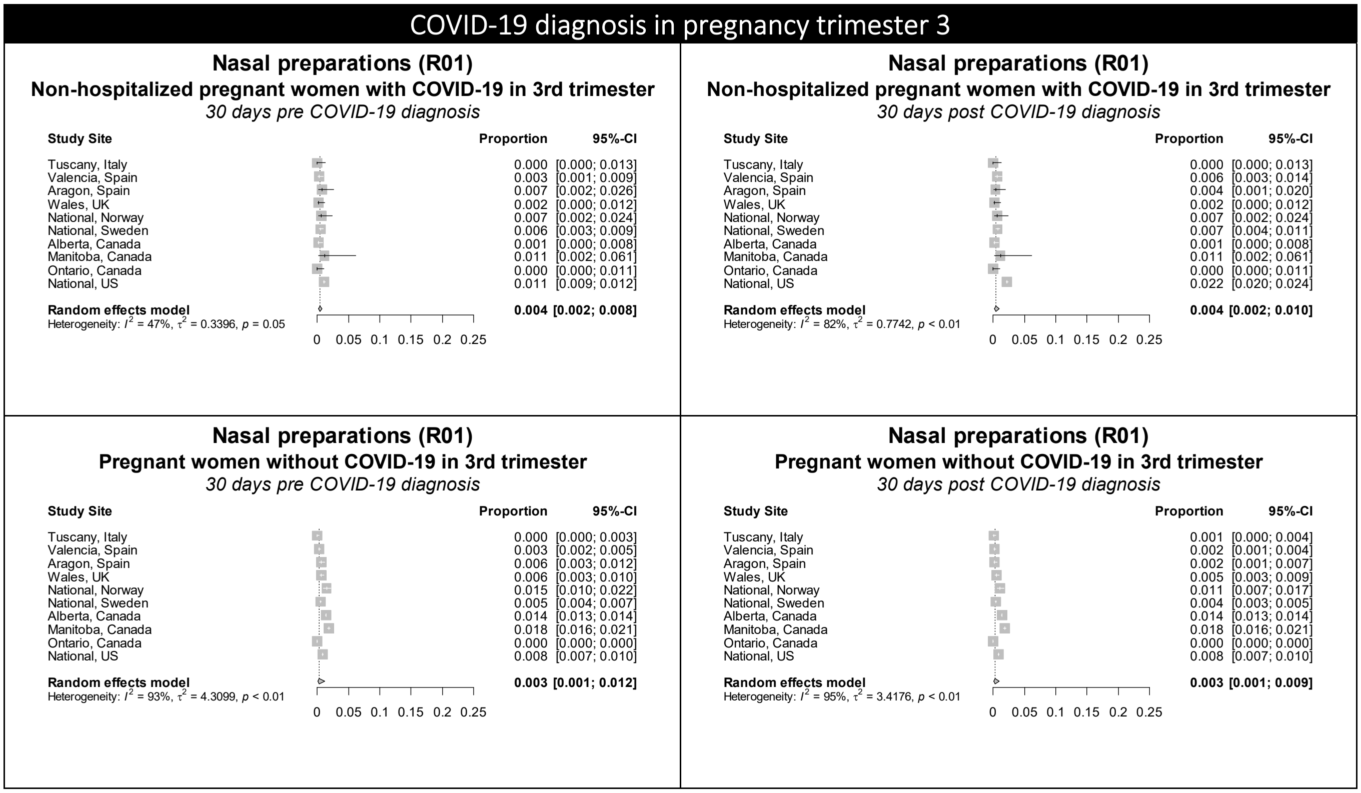

Figure S21. Forest plots showing the pooled prevalence of psychoanaleptics in the 30 days pre-COVID (left) and 30 days post-COVID (right) in non-hospitalized pregnant women with COVID-19 (upper) and pregnant women without COVID-19 (lower), by pregnancy trimester

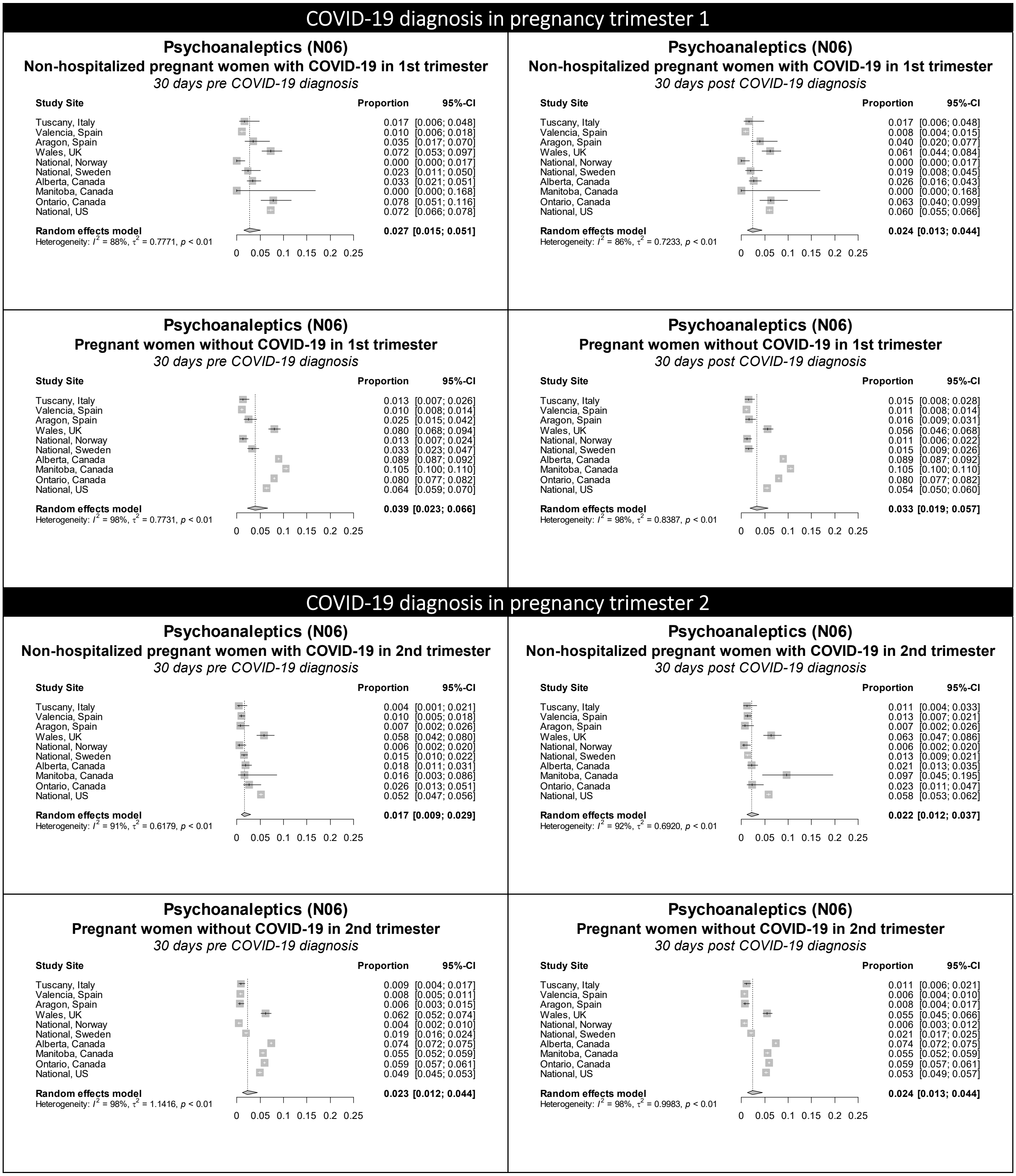

**Figure S21 continued.** Forest plots showing the pooled prevalence of psychoanaleptics in the 30 days pre-COVID (left) and 30 days post-COVID (right) in non-hospitalized pregnant women with COVID-19 (upper) and pregnant women without COVID-19 (lower), by pregnancy trimester

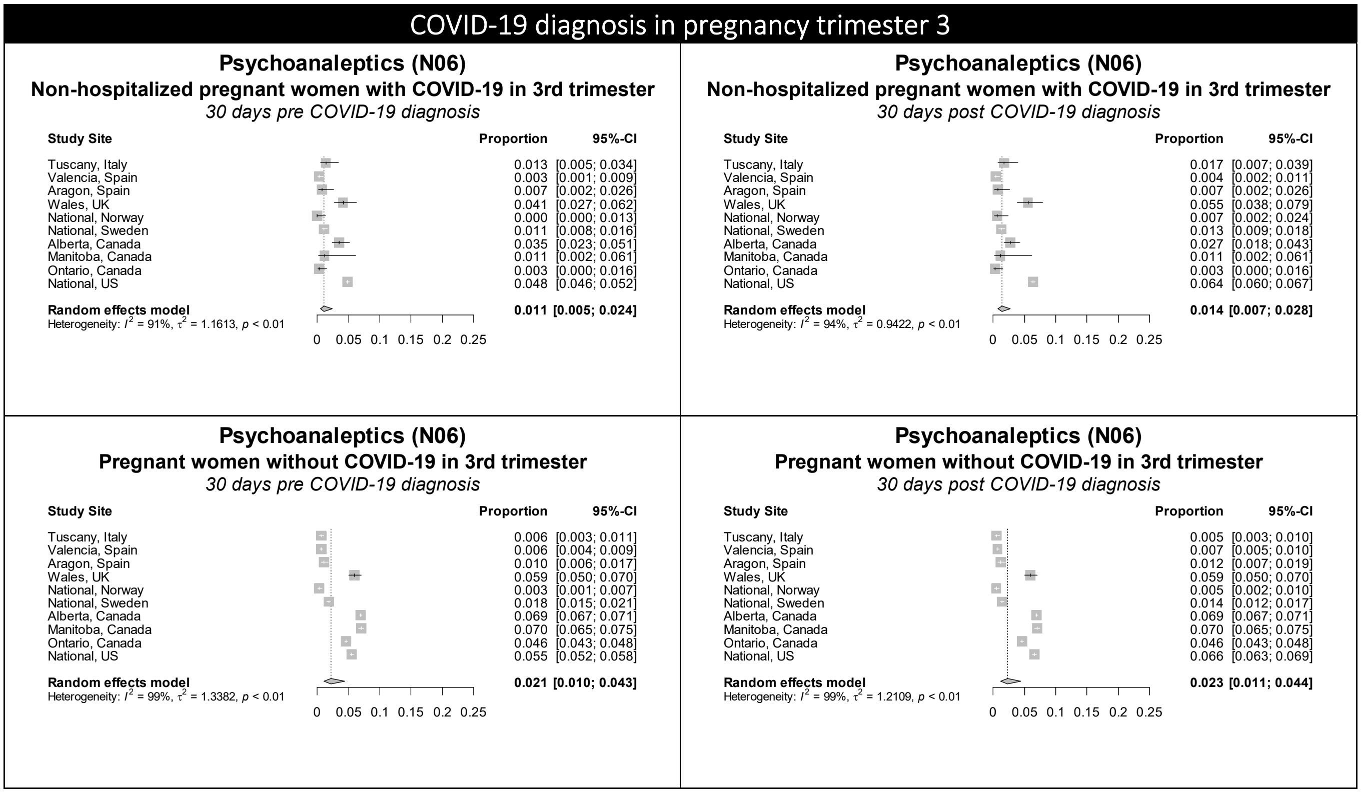

Figure S22. Forest plots showing the pooled prevalence of psycholeptics in the 30 days pre-COVID (left) and 30 days post-COVID (right) in non-hospitalized pregnant women with COVID-19 (upper) and pregnant women without COVID-19 (lower), by pregnancy trimester

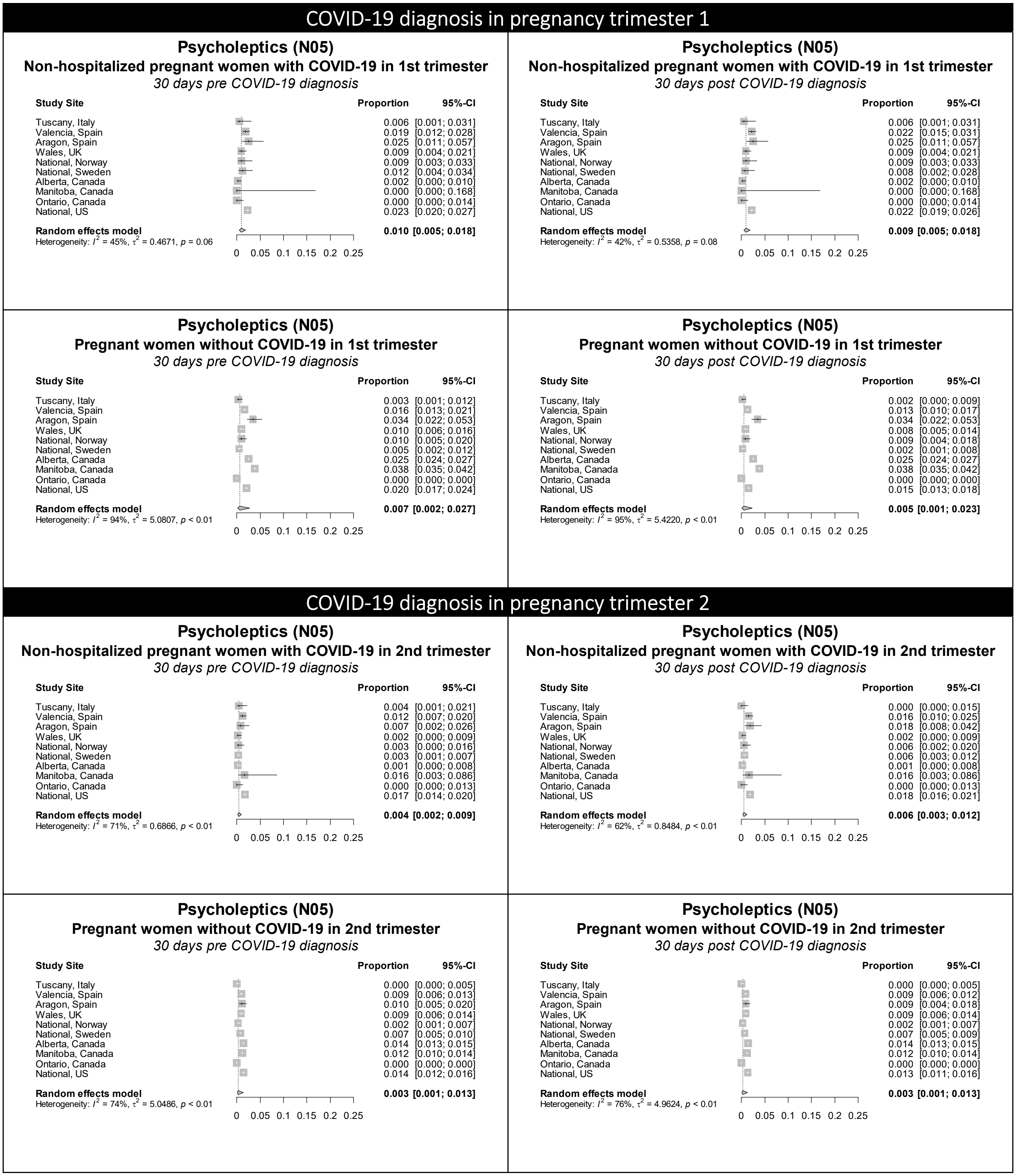

**Figure S22 continued.** Forest plots showing the pooled prevalence of psycholeptics in the 30 days pre-COVID (left) and 30 days post-COVID (right) in non-hospitalized pregnant women with COVID-19 (upper) and pregnant women without COVID-19 (lower), by pregnancy trimester

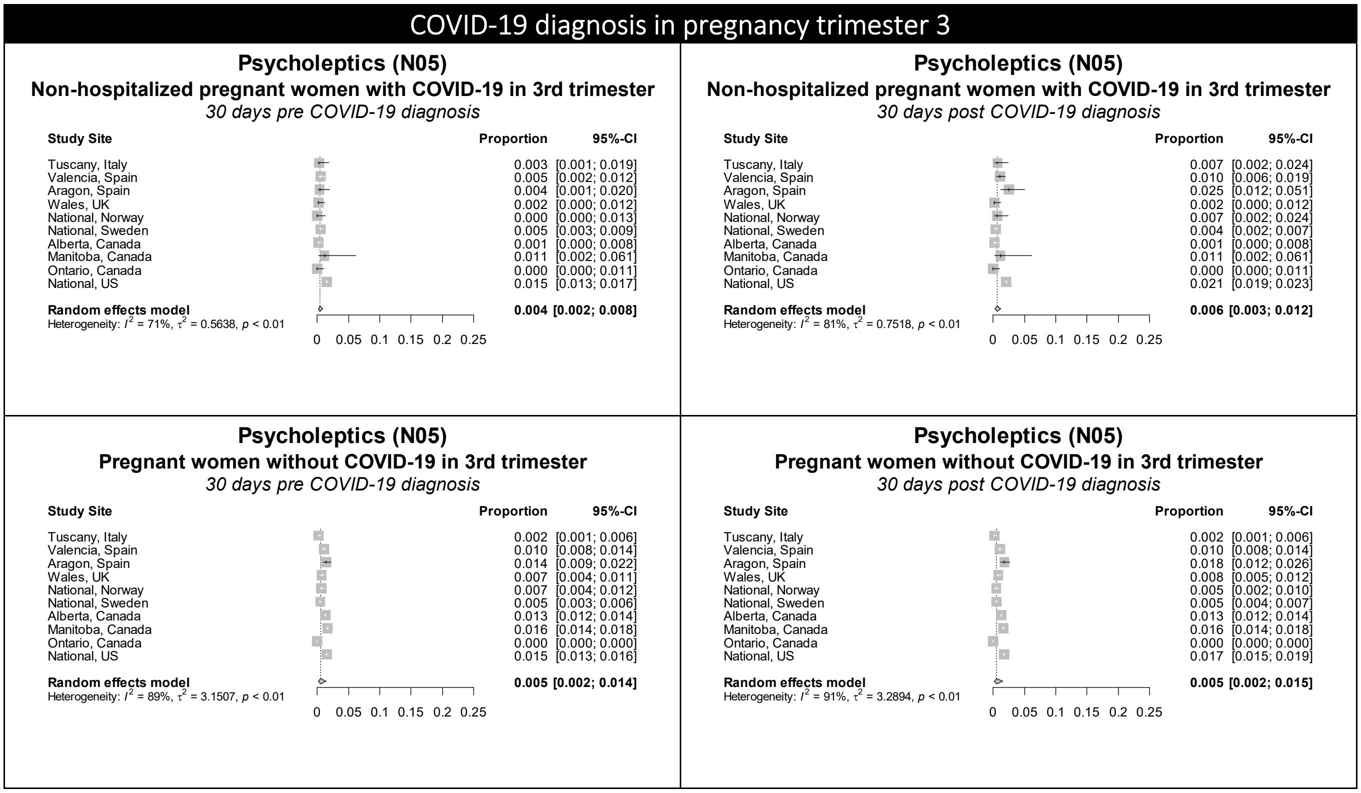

**Table S7.** Baseline characteristics of pregnant women with COVID-19 and non-pregnant women with COVID-19

| | Trimester at COVID-19 infection | COVID-19 severity | Age | | Co-morbidities[1] | |
|---|---|---|---|---|---|---|
| | Pregnant with COVID-19 | Pregnant with COVID-19[2] | Pregnant with COVID-19 | Non-pregnant with COVID-19 | Pregnant with COVID-19 | Non-pregnant with COVID-19 |
| Tuscany, Italy | 1st trimester: 20.8%<br>2nd trimester: 27.9%<br>3rd trimester: 51.3% | Hospitalized: 13.4%<br>Non-hospitalized: 74.3% | 12-24 years: 11.1%<br>25-39 years: 81.6%<br>40-55 years: 7.3% | 12-24 years: 11.1%<br>25-39 years: 81.6%<br>40-55 years: 7.3% | Any: 20.8%<br>Cardiovascular: 4.2%<br>Chronic lung: 4.6%<br>Severe obesity: 0.2% | Any: 24.8%<br>Cardiovascular: 3.9%<br>Chronic lung: 6.4%<br>Severe obesity: 0.1% |
| Valencia, Spain | 1st trimester: 35.0%<br>2nd trimester: 28.7%<br>3rd trimester: 36.4% | Hospitalized: 0.9%<br>Non-hospitalized: 88.4% | 12-24 years: 13.2%<br>25-39 years: 76.9%<br>40-55 years: 9.9% | 12-24 years: 13.2%<br>25-39 years: 76.9%<br>40-55 years: 9.9% | Any: 23.6%<br>Cardiovascular: 5.9%<br>Chronic lung: 9.9%<br>Severe obesity: 1.7% | Any: 23.8%<br>Cardiovascular: 4.5%<br>Chronic lung: 10.3%<br>Severe obesity: 1.9% |
| Aragon, Spain | 1st trimester: 22.0%<br>2nd trimester: 29.9%<br>3rd trimester: 48.2% | Hospitalized: 2.7%<br>Non-hospitalized: 79.3% | 12-24 years: 14.4%<br>25-39 years: 78.1%<br>40-55 years: 7.5% | 12-24 years: 13.8%<br>25-39 years: 78.3%<br>40-55 years: 7.9% | Any: 24.6%<br>Cardiovascular: 4.4%<br>Chronic lung: 7.5%<br>Severe obesity: 0.4% | Any: 26.7%<br>Cardiovascular: 3.0%<br>Chronic lung: 8.5%<br>Severe obesity: 1.1% |
| Norway | 1st trimester: 20.5%<br>2nd trimester: 35.7%<br>3rd trimester: 43.8% | Hospitalized: 16.8%<br>Non-hospitalized: 76.7% | 12-24 years: 10.6%<br>25-39 years: 86.2%<br>40-55 years: 3.2% | 12-24 years: 10.6%<br>25-39 years: 86.2%<br>40-55 years: 3.2% | Any: 19.4%<br>Cardiovascular: 3.6%<br>Chronic lung: 4.5%<br>Severe obesity: 1.7% | Any: 21.5%<br>Cardiovascular: 2.1%<br>Chronic lung: 6.3%<br>Severe obesity: 1.4% |
| Sweden | 1st trimester: 6.0%<br>2nd trimester: 33.9%<br>3rd trimester: 60.1% | Hospitalized: 6.8%<br>Non-hospitalized: 93.2% | 12-24 years: 6.5%<br>25-39 years: 87.5%<br>40-55 years: 6.0% | 12-24 years: 6.5%<br>25-39 years: 87.5%<br>40-55 years: 6.0% | Any: 6.1%<br>Cardiovascular: 0.4%<br>Chronic lung: 0.9%<br>Severe obesity: 3.6% | Any: 17.6%<br>Cardiovascular: 2.1%<br>Chronic lung: 6.6%<br>Severe obesity: 7.6% |
| Alberta, Canada | 1st trimester: 25.3%<br>2nd trimester: 32.3%<br>3rd trimester: 42.5% | Hospitalized: 14.3%<br>Non-hospitalized: 85.7% | 15-24 years: 15.0%<br>25-39 years: 81.7%<br>40-45 years: 3.3% | 15-24 years: 30.9%<br>25-39 years: 49.2%<br>40-45 years: 19.9% | Any: 41.9%<br>Cardiovascular: 2.5%<br>Respiratory: 7.6%<br>Severe obesity: 1.8% | Any: 47.8%<br>Cardiovascular: 2.4%<br>Respiratory: 9.0%<br>Severe obesity: 2.1% |
| Manitoba, Canada | 1st trimester: 8.1%<br>2nd trimester: 26.4%<br>3rd trimester: 65.5% | Hospitalized: 27.7%<br>Non-hospitalized: 72.3% | 15-24 years: 27.3%<br>25-39 years: 68.9%<br>40-45 years: 3.8% | 15-24 years: 32.1%<br>25-39 years: 49.1%<br>40-45 years: 18.8% | Any: 38.7%<br>Cardiovascular: <2.6%<br>Respiratory: 7.2%<br>Severe obesity: 3.4% | Any: 43.1%<br>Cardiovascular: 1.5%<br>Respiratory: 8.0%<br>Severe obesity: 1.7% |
| Ontario, Canada | 1st trimester: 28.9%<br>2nd trimester: 32.5%<br>3rd trimester: 38.6% | Hospitalized: 0%<br>Non-hospitalized: 100% | 15-24 years: 34.7%<br>25-39 years: 61.7%<br>40-45 years: 3.5% | 15-24 years: 44.1%<br>25-39 years: 37.6%<br>40-45 years: 18.3% | Any: 28.9%<br>Cardiovascular: 0%<br>Respiratory: 2.6%<br>Severe obesity: 1.2% | Any: 26.6%<br>Cardiovascular: 0.1%<br>Respiratory: 2.9%<br>Severe obesity: 2.5% |
| U.S. | 1st trimester: 20.6%<br>2nd trimester: 26.1%<br>3rd trimester: 53.3% | Hospitalized: 3.1%<br>Non-hospitalized: 87.7% | 12-24 years: 15.8%<br>25-39 years: 81.0%<br>40-55 years: 3.3% | 12-24 years: 16.0%<br>25-39 years: 80.7%<br>40-55 years: 3.3% | Any: NA<br>Cardiovascular: 0.9%<br>Chronic lung: 4.3%<br>Severe obesity: 14.3% | Any comorbidities: NA<br>Cardiovascular: 1.0%<br>Chronic lung: 4.9%<br>Severe obesity: 12.7% |

[1]Any co-morbidities included: cancer, cardiovascular disease, chronic kidney disease, chronic liver disease, chronic lung disease, common rheumatic disease, diabetes, HIV, hypertension, mental disorders, severe obesity, sickle cell disease, and use of immunosuppressants. [2]Cases in which the COVID-19 test/positive diagnosis was +/- 2 days of delivery date did not contribute to analyses by hospitalization. They did however contribute to the analyses undertaken on 'total cases'. Therefore, in some sites hospitalized and non-hospitalized cases do not add up to 100%. Abbreviations: N.A. = not available.

Figure S23. Forest plots showing the pooled prevalence of analgesics in the 30 days pre-COVID (left) and 30 days post-COVID (right) in non-hospitalized pregnant women with COVID-19 (upper) and non-hospitalized non-pregnant women with COVID-19 (lower), by pregnancy trimester

### COVID-19 diagnosis in pregnancy trimester 1

#### Analgesics (N02)
**Non-hospitalized pregnant women with COVID-19 in 1st trimester**
*30 days pre COVID-19 diagnosis*

| Study Site | Proportion | 95%-CI |
|---|---|---|
| Tuscany, Italy | 0.006 | [0.001; 0.031] |
| Valencia, Spain | 0.024 | [0.017; 0.034] |
| Aragon, Spain | 0.124 | [0.086; 0.177] |
| National, Norway | 0.037 | [0.019; 0.071] |
| National, Sweden | 0.008 | [0.002; 0.028] |
| Alberta, Canada | 0.023 | [0.013; 0.039] |
| Manitoba, Canada | 0.053 | [0.009; 0.246] |
| Ontario, Canada | 0.022 | [0.010; 0.048] |
| National, US | 0.018 | [0.015; 0.022] |
| **Random effects model** | **0.024** | **[0.014; 0.043]** |

Heterogeneity: $I^2 = 90\%$, $\tau^2 = 0.5813$, $p < 0.01$

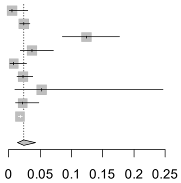

#### Analgesics (N02)
**Non-hospitalized pregnant women with COVID-19 in 1st trimester**
*30 days post COVID-19 diagnosis*

| Study Site | Proportion | 95%-CI |
|---|---|---|
| Tuscany, Italy | 0.000 | [0.000; 0.021] |
| Valencia, Spain | 0.036 | [0.027; 0.048] |
| Aragon, Spain | 0.109 | [0.073; 0.160] |
| National, Norway | 0.028 | [0.013; 0.059] |
| National, Sweden | 0.012 | [0.004; 0.034] |
| Alberta, Canada | 0.025 | [0.015; 0.041] |
| Manitoba, Canada | 0.053 | [0.009; 0.246] |
| Ontario, Canada | 0.026 | [0.013; 0.053] |
| National, US | 0.033 | [0.028; 0.038] |
| **Random effects model** | **0.028** | **[0.016; 0.047]** |

Heterogeneity: $I^2 = 78\%$, $\tau^2 = 0.4886$, $p < 0.01$

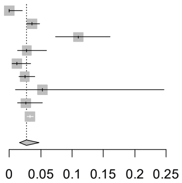

#### Analgesics (N02)
**Non-hospitalized non-pregnant women with COVID-19**
*30 days pre COVID-19 diagnosis*

| Study Site | Proportion | 95%-CI |
|---|---|---|
| Tuscany, Italy | 0.005 | [0.002; 0.015] |
| Valencia, Spain | 0.032 | [0.027; 0.038] |
| Aragon, Spain | 0.099 | [0.077; 0.128] |
| National, Norway | 0.036 | [0.024; 0.053] |
| National, Sweden | 0.025 | [0.016; 0.039] |
| Alberta, Canada | 0.025 | [0.024; 0.026] |
| Manitoba, Canada | 0.055 | [0.050; 0.061] |
| Ontario, Canada | 0.016 | [0.015; 0.017] |
| National, US | 0.036 | [0.031; 0.041] |
| **Random effects model** | **0.030** | **[0.019; 0.047]** |

Heterogeneity: $I^2 = 98\%$, $\tau^2 = 0.4700$, $p < 0.01$

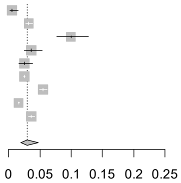

#### Analgesics (N02)
**Non-hospitalized non-pregnant women with COVID-19**
*30 days post COVID-19 diagnosis*

| Study Site | Proportion | 95%-CI |
|---|---|---|
| Tuscany, Italy | 0.010 | [0.005; 0.022] |
| Valencia, Spain | 0.061 | [0.054; 0.069] |
| Aragon, Spain | 0.116 | [0.092; 0.146] |
| National, Norway | 0.053 | [0.038; 0.073] |
| National, Sweden | 0.038 | [0.027; 0.054] |
| Alberta, Canada | 0.028 | [0.027; 0.029] |
| Manitoba, Canada | 0.076 | [0.070; 0.083] |
| Ontario, Canada | 0.022 | [0.020; 0.023] |
| National, US | 0.035 | [0.030; 0.040] |
| **Random effects model** | **0.041** | **[0.027; 0.062]** |

Heterogeneity: $I^2 = 99\%$, $\tau^2 = 0.4299$, $p < 0.01$

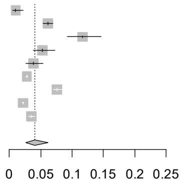

### COVID-19 diagnosis in pregnancy trimester 2

#### Analgesics (N02)
**Non-hospitalized pregnant women with COVID-19 in 2nd trimester**
*30 days pre COVID-19 diagnosis*

| Study Site | Proportion | 95%-CI |
|---|---|---|
| Tuscany, Italy | 0.004 | [0.001; 0.021] |
| Valencia, Spain | 0.014 | [0.008; 0.023] |
| Aragon, Spain | 0.118 | [0.085; 0.161] |
| National, Norway | 0.017 | [0.008; 0.036] |
| National, Sweden | 0.010 | [0.006; 0.017] |
| Alberta, Canada | 0.001 | [0.000; 0.008] |
| Manitoba, Canada | 0.016 | [0.003; 0.086] |
| Ontario, Canada | 0.020 | [0.009; 0.043] |
| National, US | 0.020 | [0.017; 0.023] |
| **Random effects model** | **0.014** | **[0.006; 0.030]** |

Heterogeneity: $I^2 = 93\%$, $\tau^2 = 1.1099$, $p < 0.01$

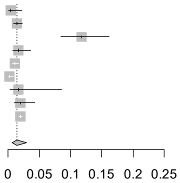

#### Analgesics (N02)
**Non-hospitalized pregnant women with COVID-19 in 2nd trimester**
*30 days post COVID-19 diagnosis*

| Study Site | Proportion | 95%-CI |
|---|---|---|
| Tuscany, Italy | 0.000 | [0.000; 0.015] |
| Valencia, Spain | 0.033 | [0.024; 0.046] |
| Aragon, Spain | 0.110 | [0.078; 0.153] |
| National, Norway | 0.017 | [0.008; 0.036] |
| National, Sweden | 0.020 | [0.014; 0.029] |
| Alberta, Canada | 0.001 | [0.000; 0.008] |
| Manitoba, Canada | 0.016 | [0.003; 0.086] |
| Ontario, Canada | 0.030 | [0.016; 0.055] |
| National, US | 0.035 | [0.031; 0.040] |
| **Random effects model** | **0.017** | **[0.007; 0.041]** |

Heterogeneity: $I^2 = 88\%$, $\tau^2 = 1.4243$, $p < 0.01$

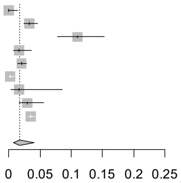

#### Analgesics (N02)
**Non-hospitalized non-pregnant women with COVID-19**
*30 days pre COVID-19 diagnosis*

| Study Site | Proportion | 95%-CI |
|---|---|---|
| Tuscany, Italy | 0.001 | [0.000; 0.007] |
| Valencia, Spain | 0.040 | [0.034; 0.048] |
| Aragon, Spain | 0.109 | [0.088; 0.133] |
| National, Norway | 0.039 | [0.029; 0.052] |
| National, Sweden | 0.023 | [0.019; 0.028] |
| Alberta, Canada | 0.025 | [0.024; 0.026] |
| Manitoba, Canada | 0.055 | [0.050; 0.061] |
| Ontario, Canada | 0.016 | [0.015; 0.017] |
| National, US | 0.035 | [0.031; 0.040] |
| **Random effects model** | **0.028** | **[0.015; 0.051]** |

Heterogeneity: $I^2 = 99\%$, $\tau^2 = 0.8424$, $p < 0.01$

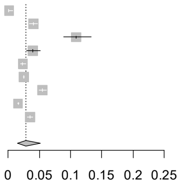

#### Analgesics (N02)
**Non-hospitalized non-pregnant women with COVID-19**
*30 days post COVID-19 diagnosis*

| Study Site | Proportion | 95%-CI |
|---|---|---|
| Tuscany, Italy | 0.002 | [0.001; 0.009] |
| Valencia, Spain | 0.070 | [0.062; 0.080] |
| Aragon, Spain | 0.141 | [0.118; 0.168] |
| National, Norway | 0.063 | [0.050; 0.078] |
| National, Sweden | 0.025 | [0.021; 0.030] |
| Alberta, Canada | 0.028 | [0.027; 0.029] |
| Manitoba, Canada | 0.076 | [0.070; 0.083] |
| Ontario, Canada | 0.022 | [0.020; 0.023] |
| National, US | 0.043 | [0.039; 0.048] |
| **Random effects model** | **0.038** | **[0.020; 0.070]** |

Heterogeneity: $I^2 = 99\%$, $\tau^2 = 0.9526$, $p < 0.01$

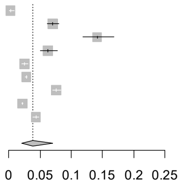

**Figure S23 continued.** Forest plots showing the pooled prevalence of analgesics in the 30 days pre-COVID (left) and 30 days post-COVID (right) in non-hospitalized pregnant women with COVID-19 (upper) and non-hospitalized non-pregnant women with COVID-19 (lower), by pregnancy trimester

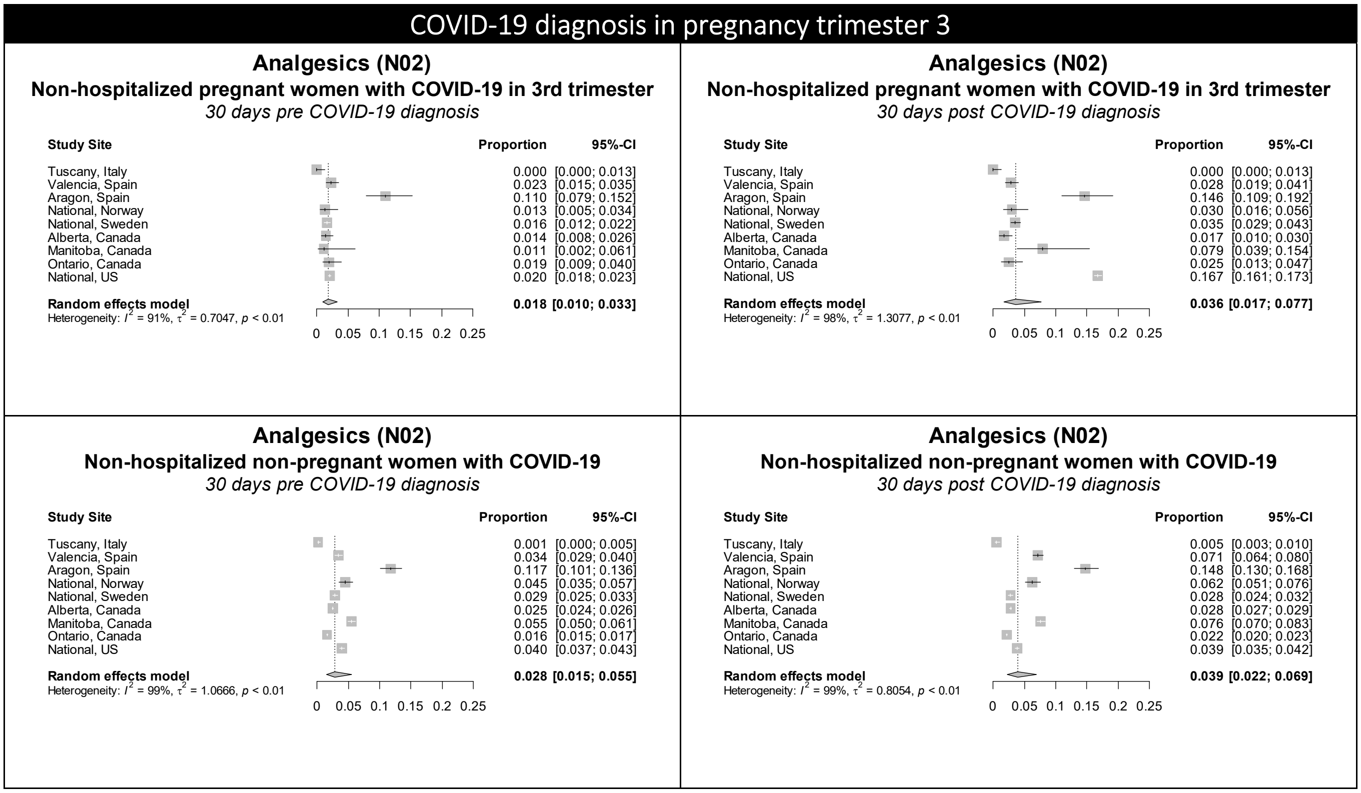

Figure S24. Forest plots showing the pooled prevalence of anthelminthics in the 30 days pre-COVID (left) and 30 days post-COVID (right) in non-hospitalized pregnant women with COVID-19 (upper) and non-hospitalized non-pregnant women with COVID-19 (lower), by pregnancy trimester

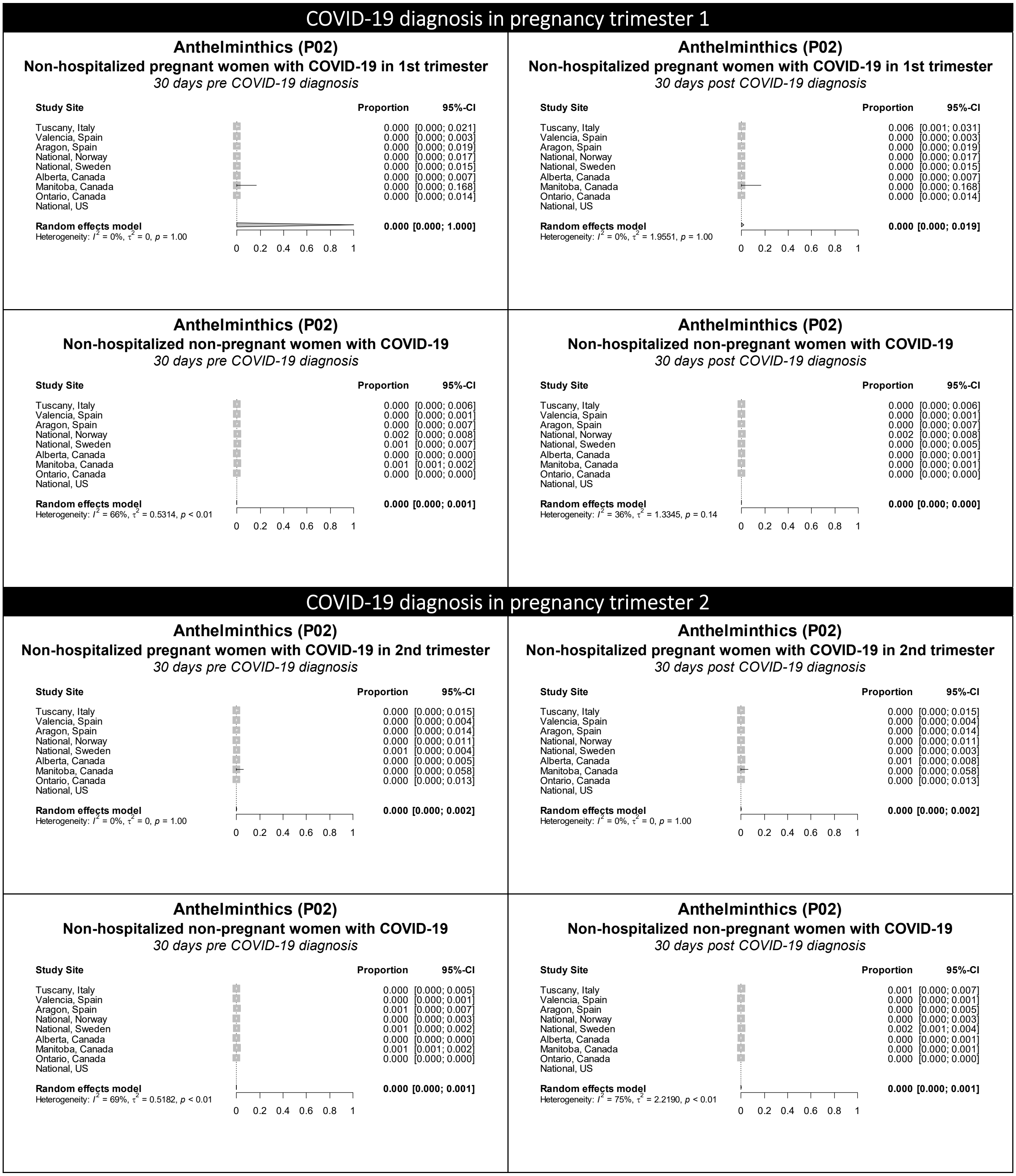

**Figure S24 continued.** Forest plots showing the pooled prevalence of anthelminthics in the 30 days pre-COVID (left) and 30 days post-COVID (right) in non-hospitalized pregnant women with COVID-19 (upper) and non-hospitalized non-pregnant women with COVID-19 (lower), by pregnancy trimester

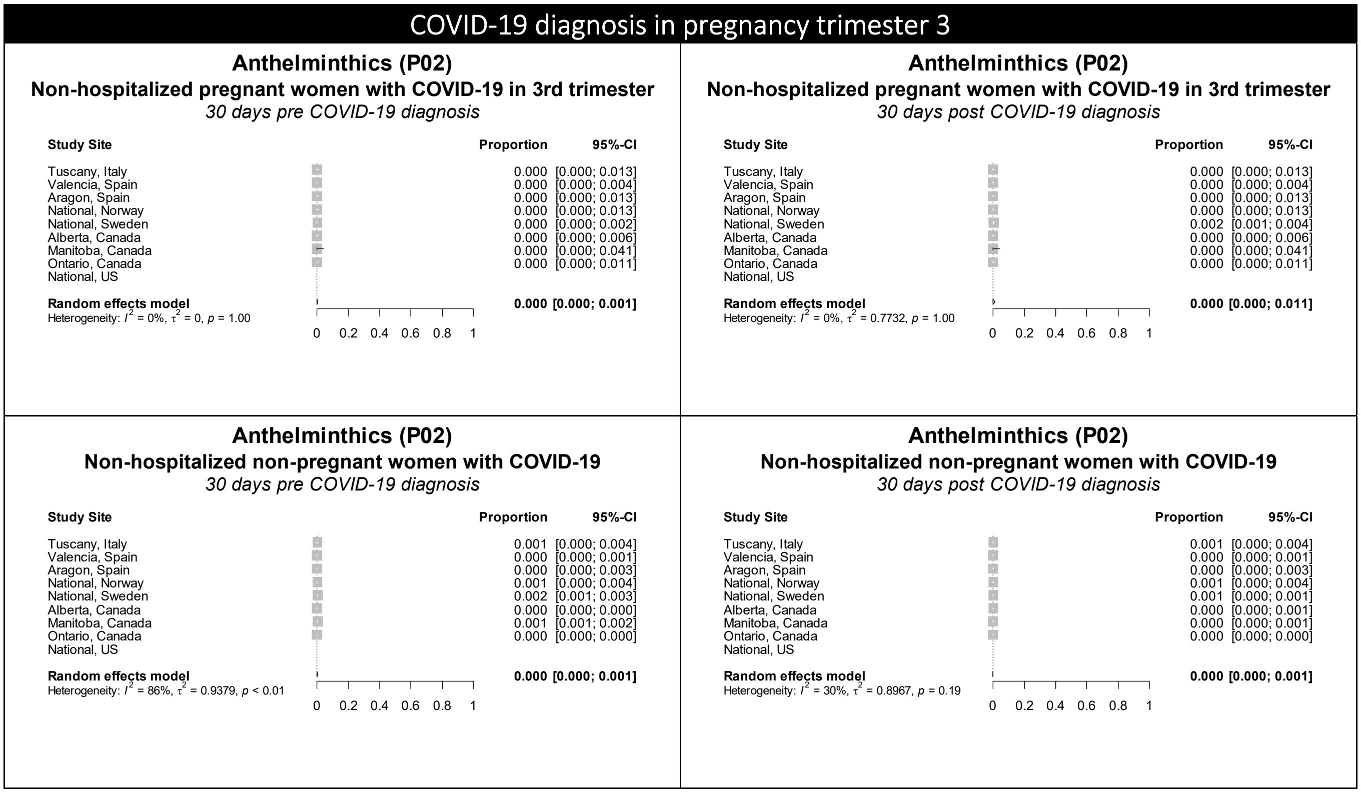

Figure S25. Forest plots showing the pooled prevalence of anti-inflammatory and antirheumatic products in the 30 days pre-COVID (left) and 30 days post-COVID (right) in non-hospitalized pregnant women with COVID-19 (upper) and non-hospitalized non-pregnant women with COVID-19 (lower), by pregnancy trimester

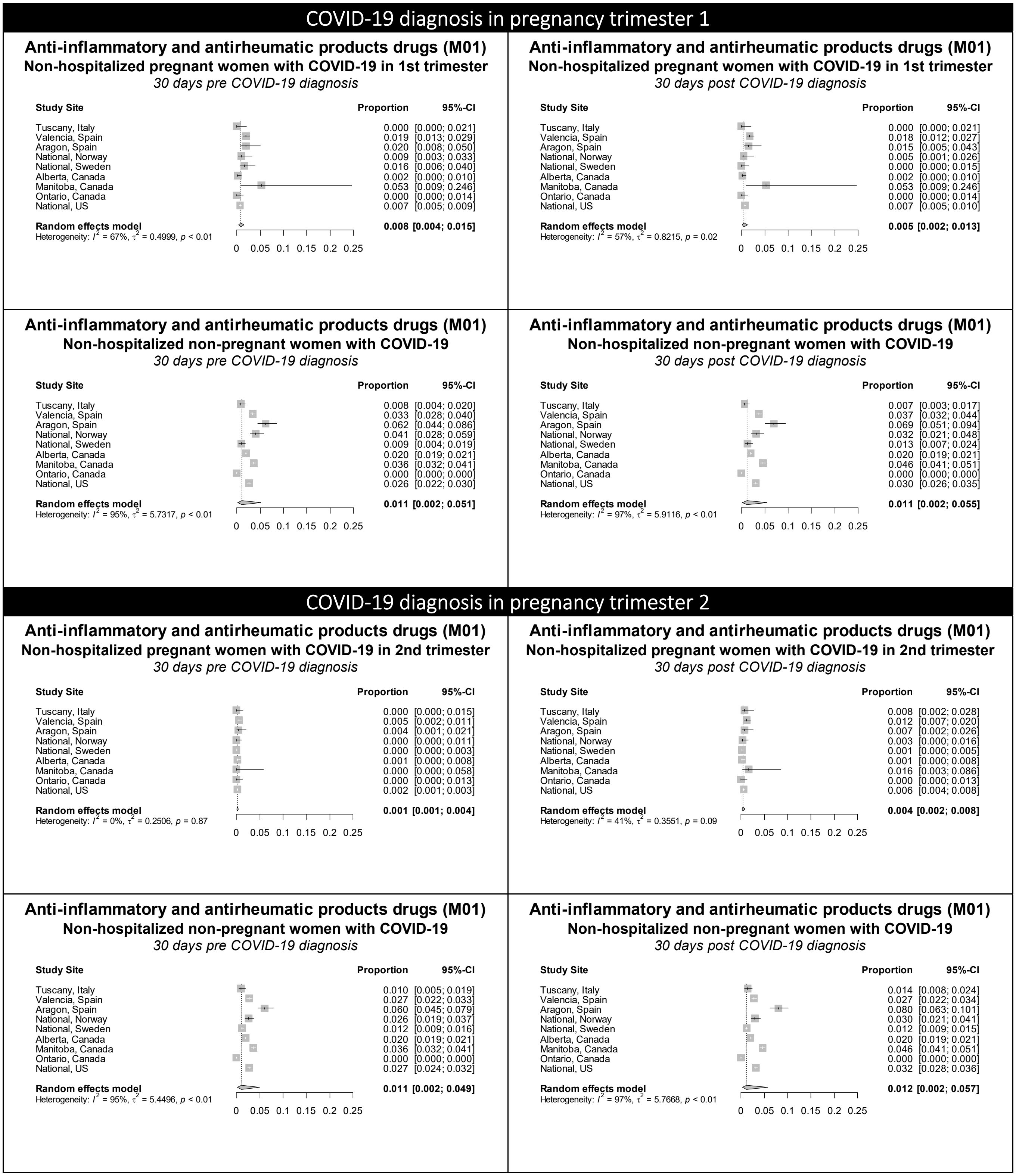

**Figure S25 continued.** Forest plots showing the pooled prevalence of anti-inflammatory and antirheumatic products in the 30 days pre-COVID (left) and 30 days post-COVID (right) in non-hospitalized pregnant women with COVID-19 (upper) and non-hospitalized non-pregnant women with COVID-19 (lower), by pregnancy trimester

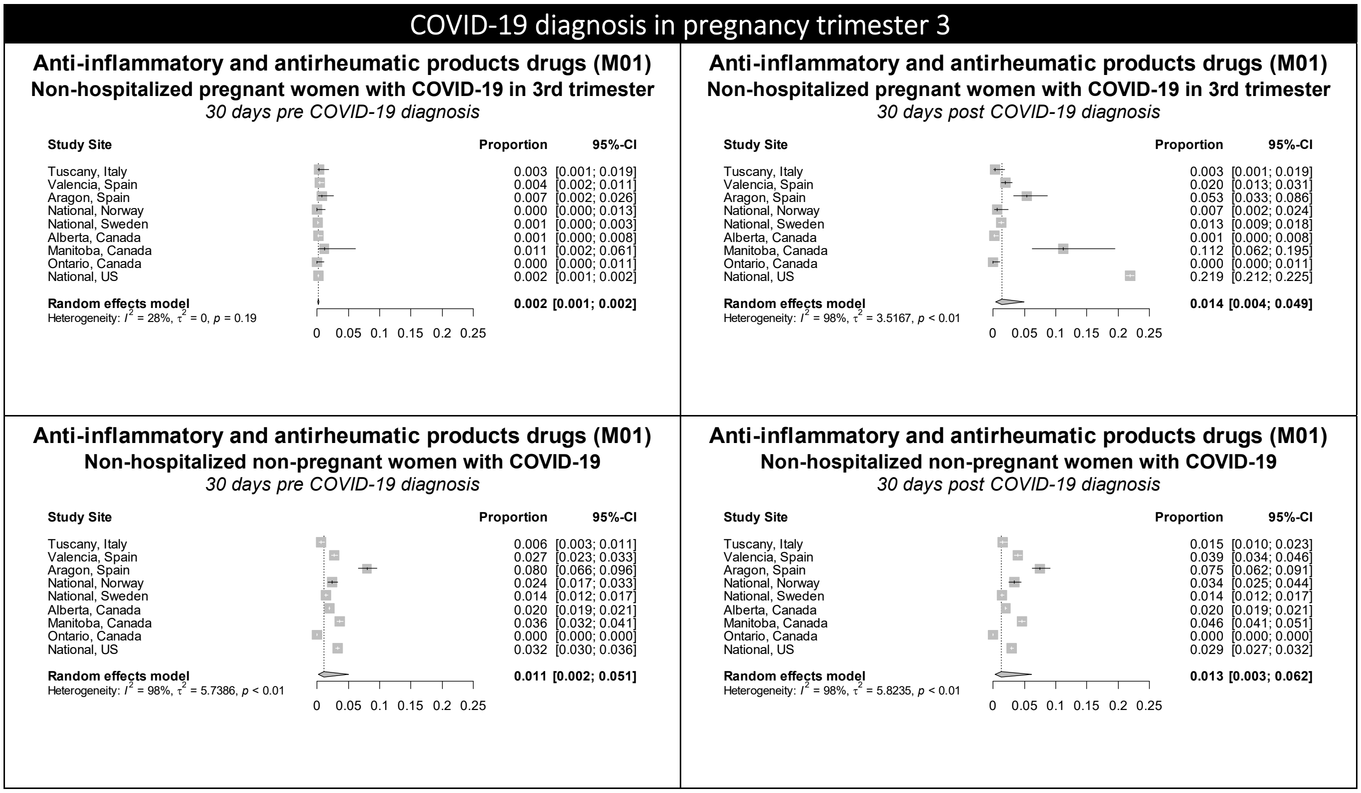

Figure S26. Forest plots showing the pooled prevalence of antibacterials in the 30 days pre-COVID (left) and 30 days post-COVID (right) in non-hospitalized pregnant women with COVID-19 (upper) and non-hospitalized non-pregnant women with COVID-19 (lower), by pregnancy trimester

### COVID-19 diagnosis in pregnancy trimester 1

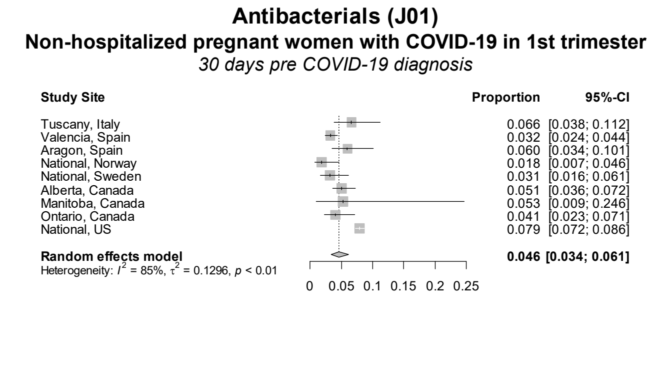

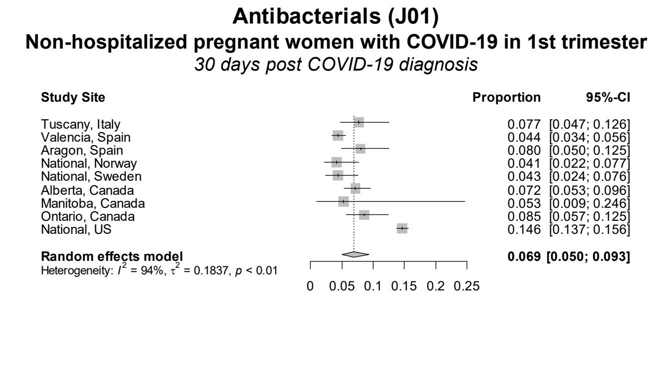

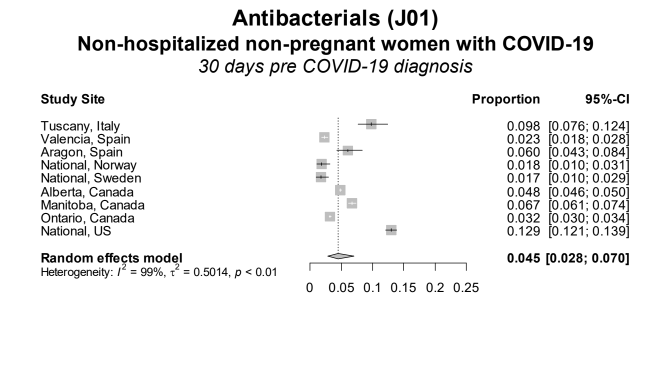

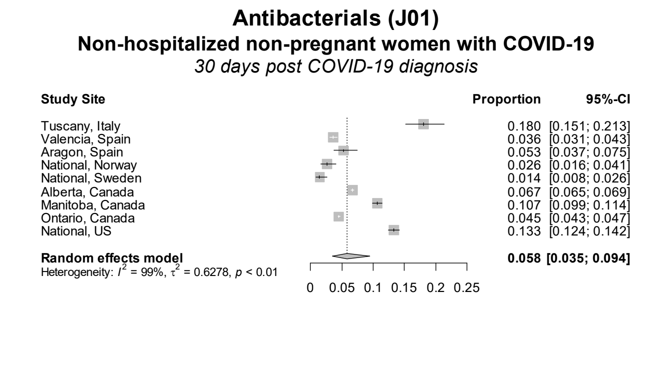

### COVID-19 diagnosis in pregnancy trimester 2

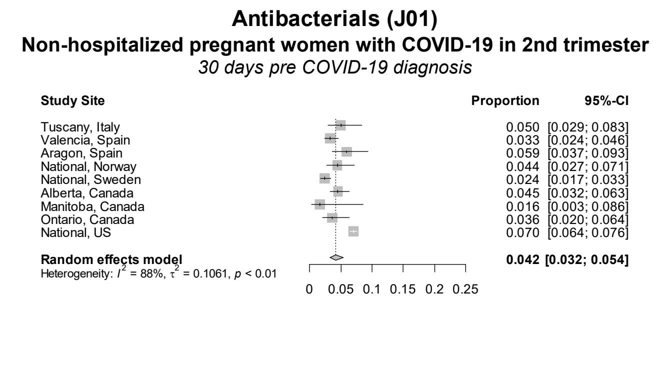

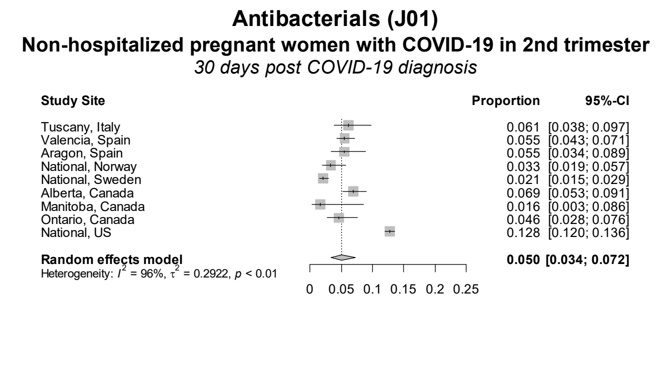

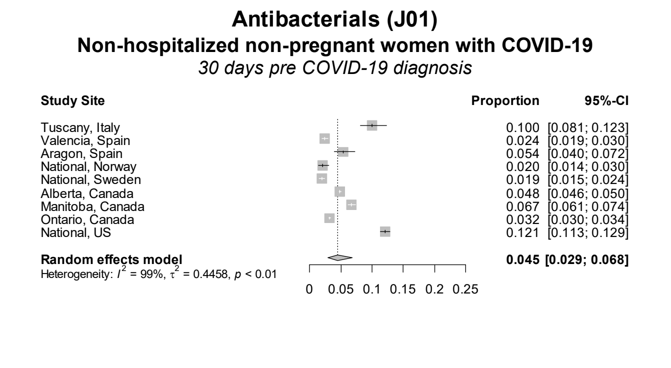

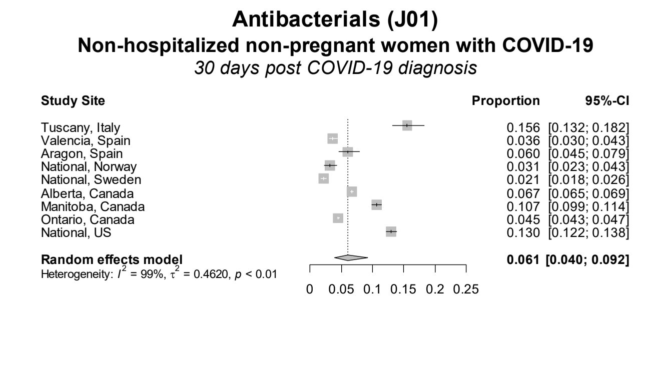

Figure S26 continued. Forest plots showing the pooled prevalence of antibacterials in the 30 days pre-COVID (left) and 30 days post-COVID (right) in non-hospitalized pregnant women with COVID-19 (upper) and non-hospitalized non-pregnant women with COVID-19 (lower), by pregnancy trimester

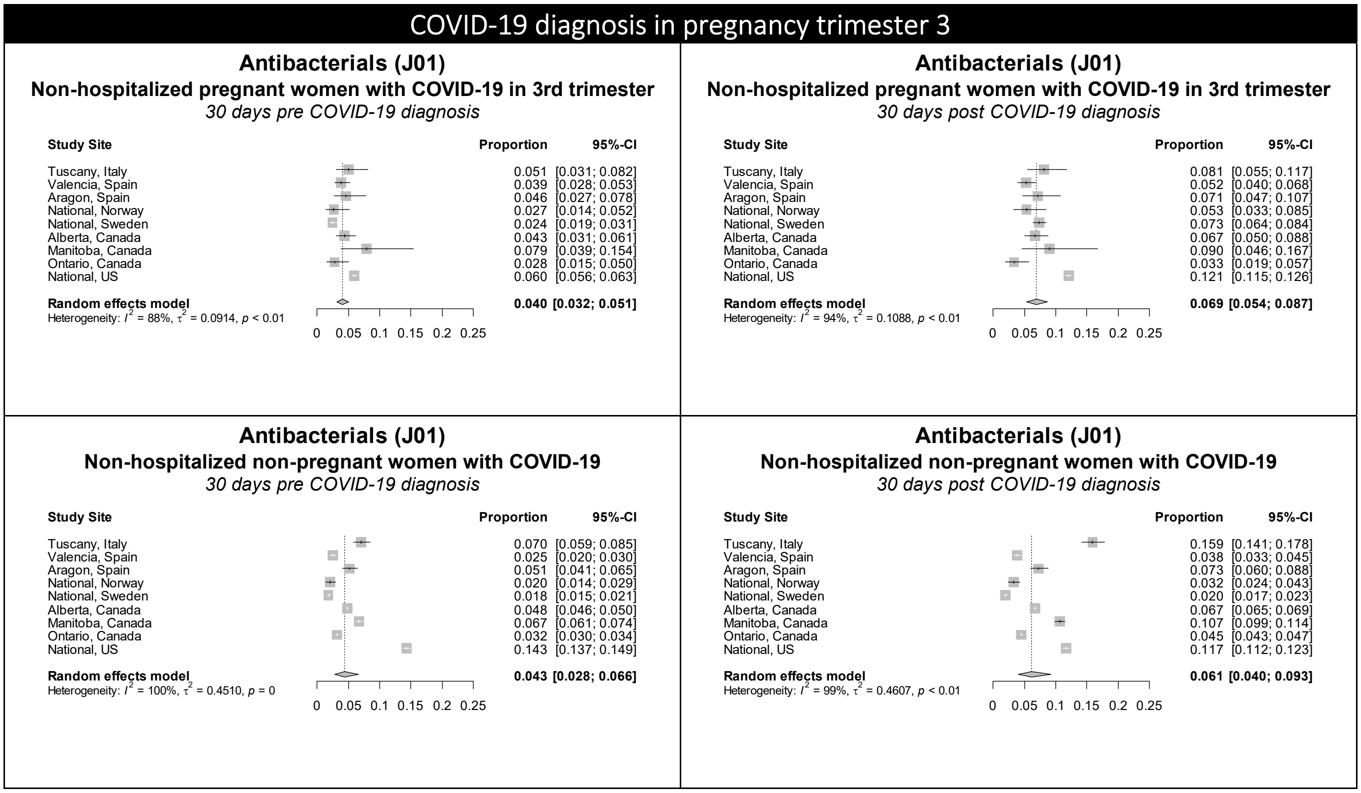

Figure S27. Forest plots showing the pooled prevalence of antigout preparations in the 30 days pre-COVID (left) and 30 days post-COVID (right) in non-hospitalized pregnant women with COVID-19 (upper) and non-hospitalized non-pregnant women with COVID-19 (lower), by pregnancy trimester

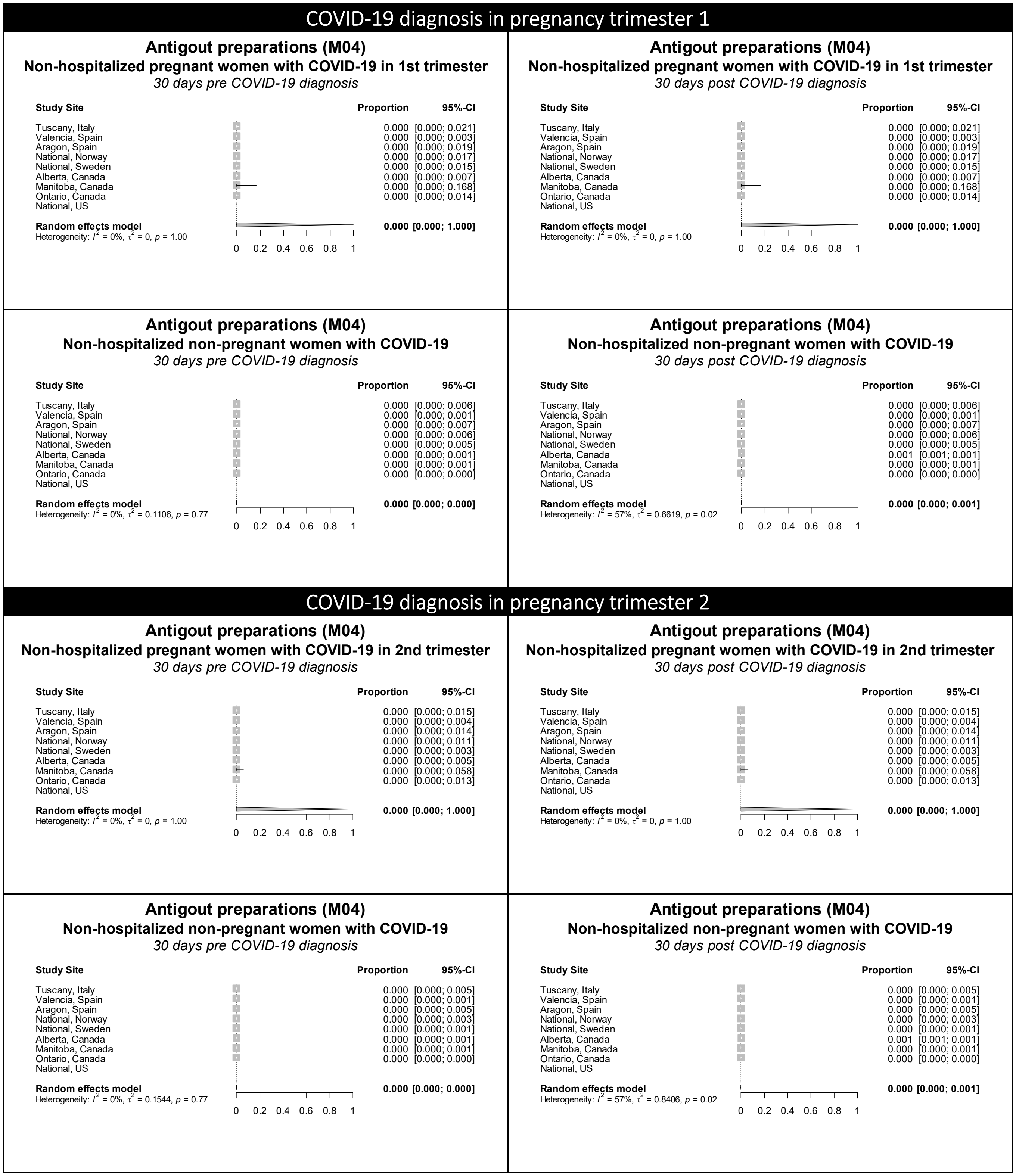

**Figure S27 continued.** Forest plots showing the pooled prevalence of antigout preparations in the 30 days pre-COVID (left) and 30 days post-COVID (right) in non-hospitalized pregnant women with COVID-19 (upper) and non-hospitalized non-pregnant women with COVID-19 (lower), by pregnancy trimester

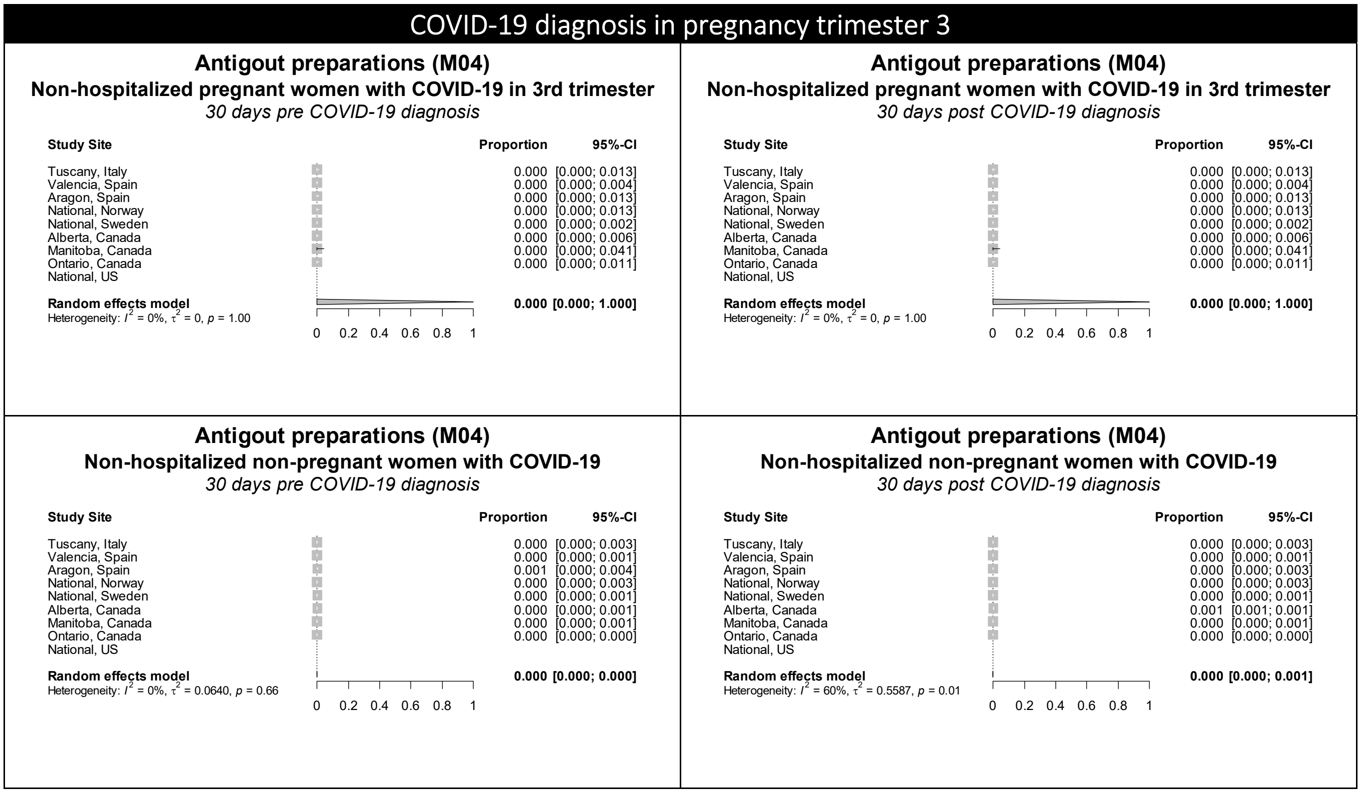

Figure S28. Forest plots showing the pooled prevalence of antihypertensives in the 30 days pre-COVID (left) and 30 days post-COVID (right) in non-hospitalized pregnant women with COVID-19 (upper) and non-hospitalized non-pregnant women with COVID-19 (lower), by pregnancy trimester

### COVID-19 diagnosis in pregnancy trimester 1

#### Antihypertensives (C02)
**Non-hospitalized pregnant women with COVID-19 in 1st trimester**
*30 days pre COVID-19 diagnosis*

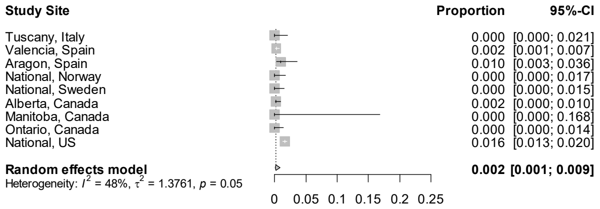

#### Antihypertensives (C02)
**Non-hospitalized pregnant women with COVID-19 in 1st trimester**
*30 days post COVID-19 diagnosis*

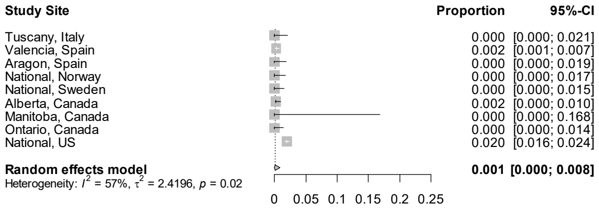

#### Antihypertensives (C02)
**Non-hospitalized non-pregnant women with COVID-19**
*30 days pre COVID-19 diagnosis*

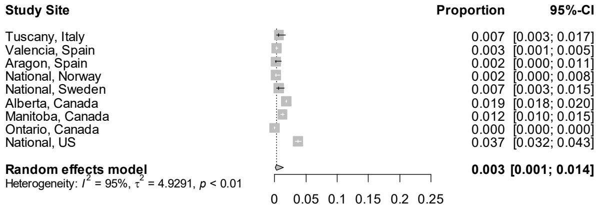

#### Antihypertensives (C02)
**Non-hospitalized non-pregnant women with COVID-19**
*30 days post COVID-19 diagnosis*

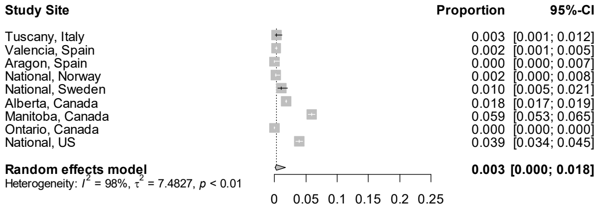

### COVID-19 diagnosis in pregnancy trimester 2

#### Antihypertensives (C02)
**Non-hospitalized pregnant women with COVID-19 in 2nd trimester**
*30 days pre COVID-19 diagnosis*

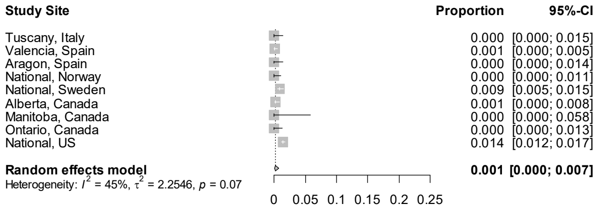

#### Antihypertensives (C02)
**Non-hospitalized pregnant women with COVID-19 in 2nd trimester**
*30 days post COVID-19 diagnosis*

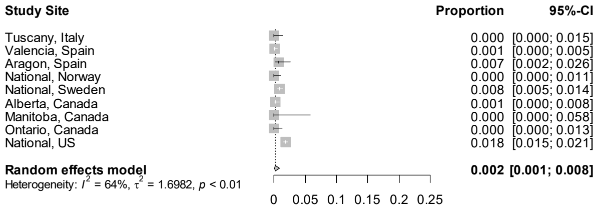

#### Antihypertensives (C02)
**Non-hospitalized non-pregnant women with COVID-19**
*30 days pre COVID-19 diagnosis*

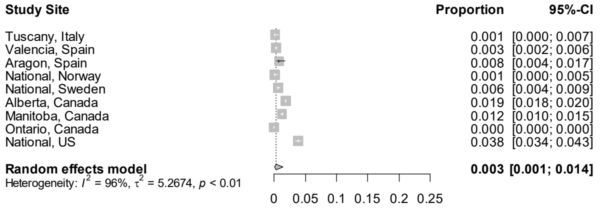

#### Antihypertensives (C02)
**Non-hospitalized non-pregnant women with COVID-19**
*30 days post COVID-19 diagnosis*

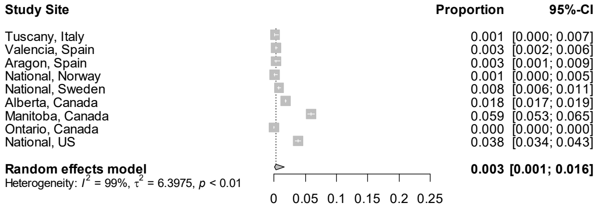

**Figure S28 continued.** Forest plots showing the pooled prevalence of antihypertensives in the 30 days pre-COVID (left) and 30 days post-COVID (right) in non-hospitalized pregnant women with COVID-19 (upper) and non-hospitalized non-pregnant women with COVID-19 (lower), by pregnancy trimester

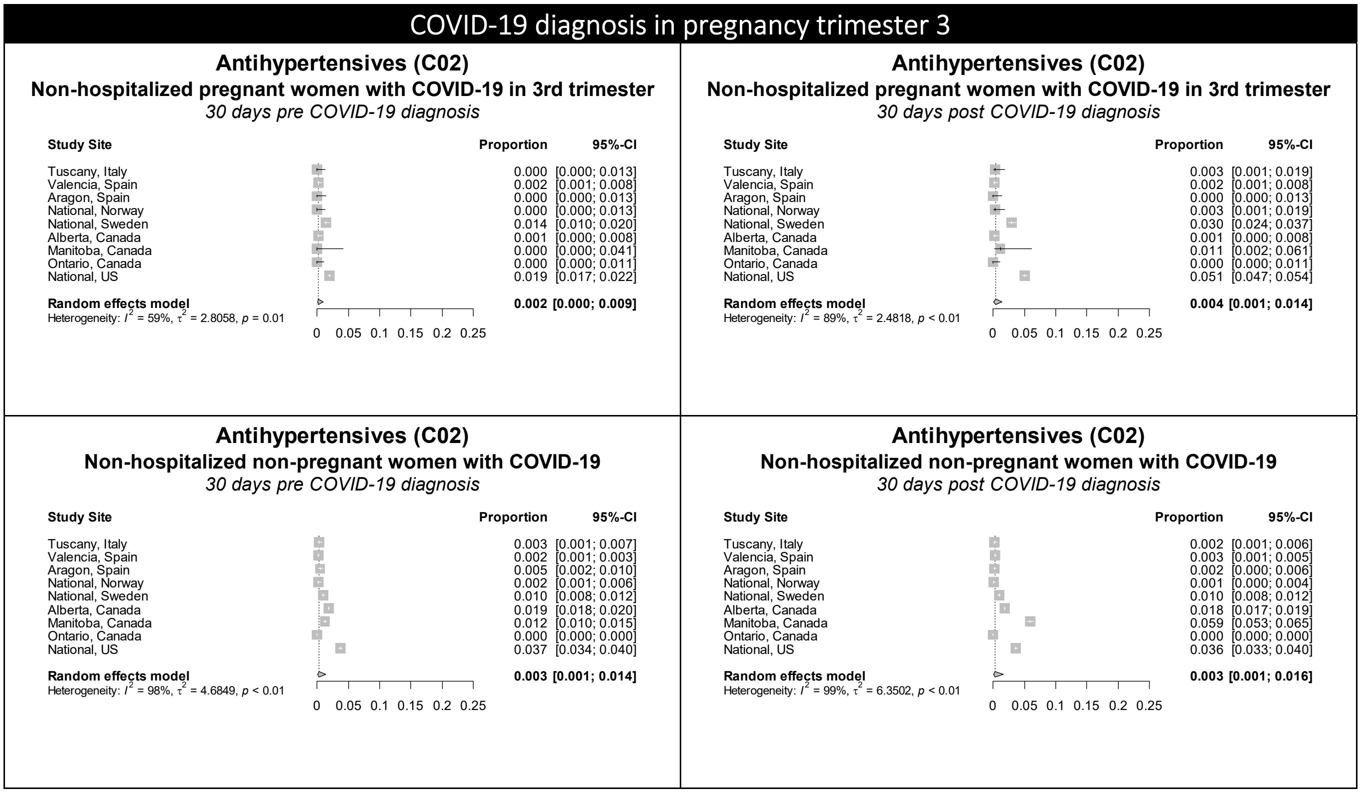

Figure S29. Forest plots showing the pooled prevalence of antimycobacterials in the 30 days pre-COVID (left) and 30 days post-COVID (right) in non-hospitalized pregnant women with COVID-19 (upper) and non-hospitalized non-pregnant women with COVID-19 (lower), by pregnancy trimester

### COVID-19 diagnosis in pregnancy trimester 1

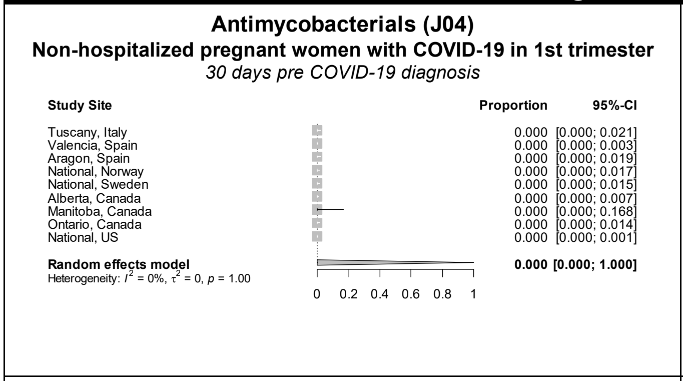

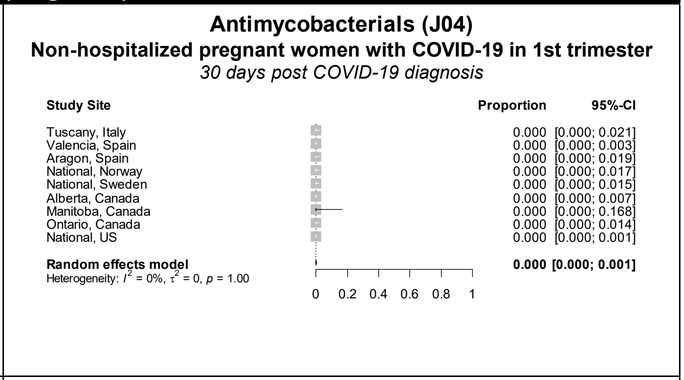

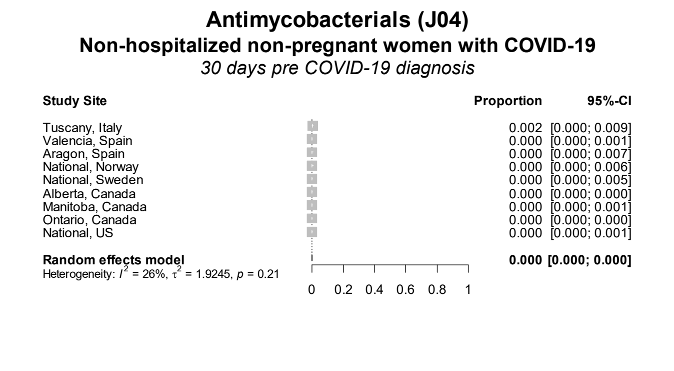

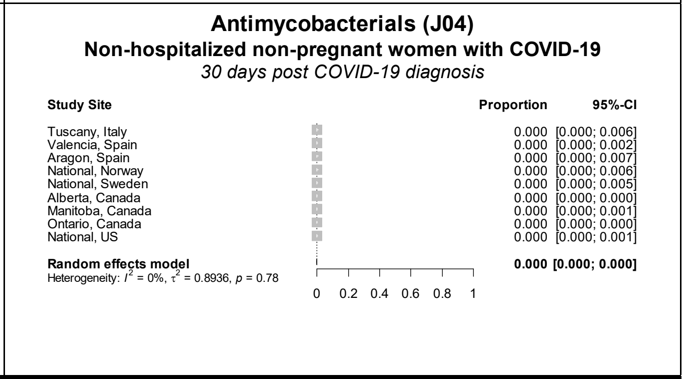

### COVID-19 diagnosis in pregnancy trimester 2

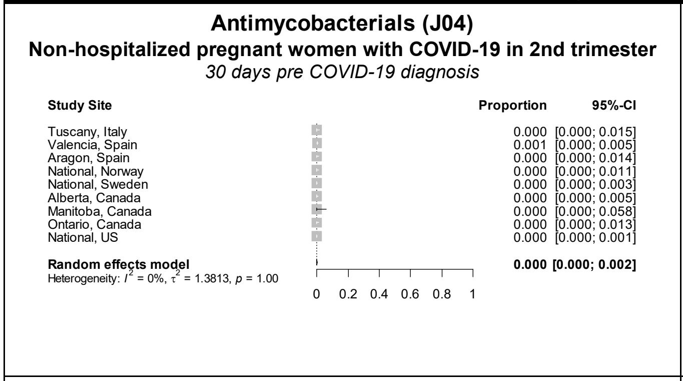

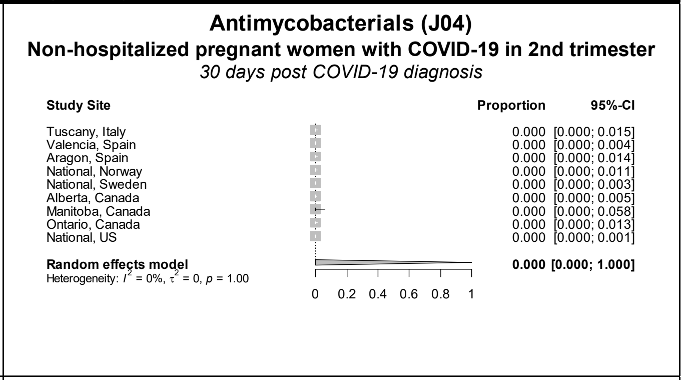

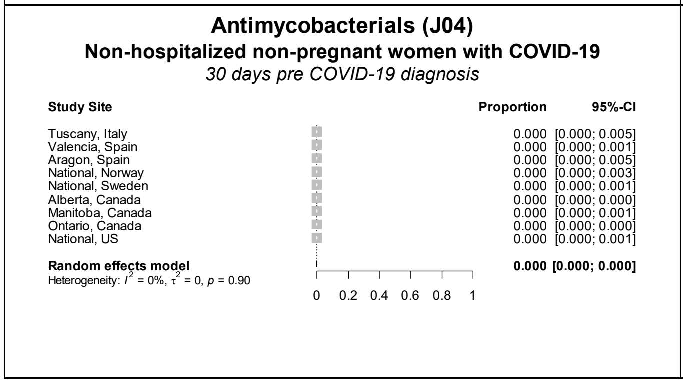

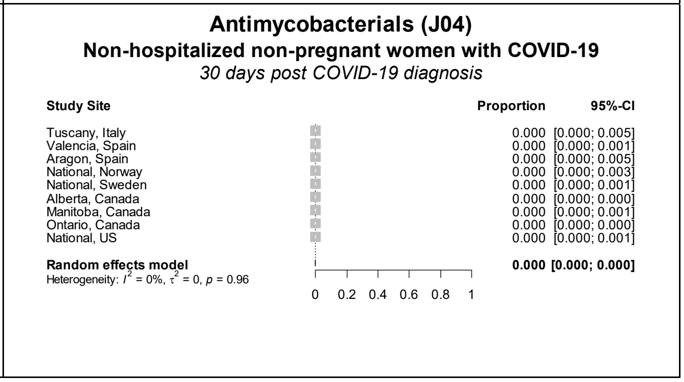

**Figure S29 continued.** Forest plots showing the pooled prevalence of antimycobacterials in the 30 days pre-COVID (left) and 30 days post-COVID (right) in non-hospitalized pregnant women with COVID-19 (upper) and non-hospitalized non-pregnant women with COVID-19 (lower), by pregnancy trimester

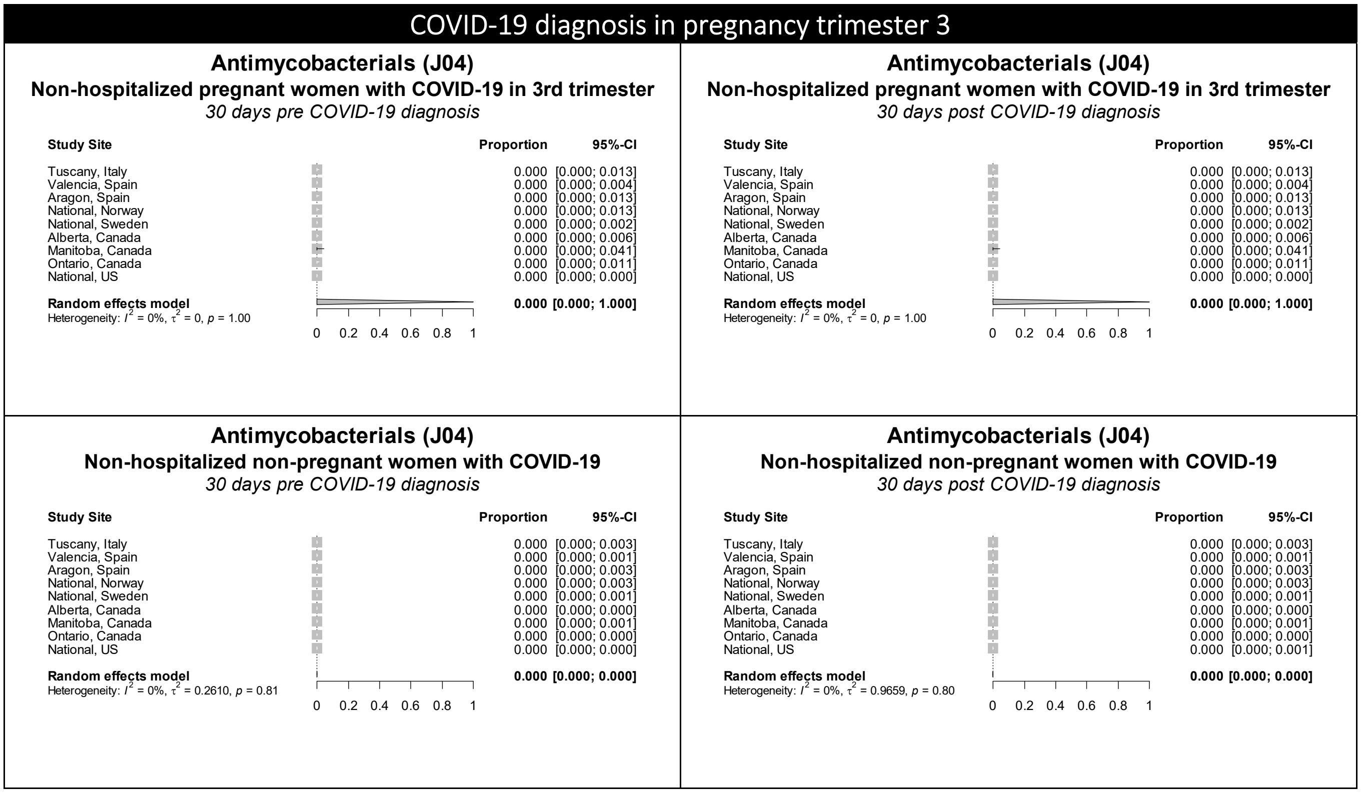

Figure S30. Forest plots showing the pooled prevalence of antimycotics in the 30 days pre-COVID (left) and 30 days post-COVID (right) in non-hospitalized pregnant women with COVID-19 (upper) and non-hospitalized non-pregnant women with COVID-19 (lower), by pregnancy trimester

### COVID-19 diagnosis in pregnancy trimester 1

**Antimycotics (J02)**
**Non-hospitalized pregnant women with COVID-19 in 1st trimester**
*30 days pre COVID-19 diagnosis*

| Study Site | Proportion | 95%-CI |
|---|---|---|
| Tuscany, Italy | 0.000 | [0.000; 0.021] |
| Valencia, Spain | 0.001 | [0.000; 0.005] |
| Aragon, Spain | 0.000 | [0.000; 0.019] |
| National, Norway | 0.000 | [0.000; 0.017] |
| National, Sweden | 0.004 | [0.001; 0.022] |
| Alberta, Canada | 0.002 | [0.000; 0.010] |
| Manitoba, Canada | 0.000 | [0.000; 0.168] |
| Ontario, Canada | 0.000 | [0.000; 0.014] |
| National, US | 0.009 | [0.007; 0.012] |
| **Random effects model** | **0.002** | **[0.000; 0.006]** |

Heterogeneity: $I^2 = 7\%$, $\tau^2 = 1.1003$, $p = 0.38$

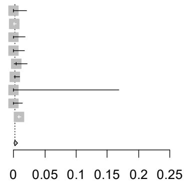

**Antimycotics (J02)**
**Non-hospitalized pregnant women with COVID-19 in 1st trimester**
*30 days post COVID-19 diagnosis*

| Study Site | Proportion | 95%-CI |
|---|---|---|
| Tuscany, Italy | 0.006 | [0.001; 0.031] |
| Valencia, Spain | 0.000 | [0.000; 0.003] |
| Aragon, Spain | 0.000 | [0.000; 0.019] |
| National, Norway | 0.000 | [0.000; 0.017] |
| National, Sweden | 0.000 | [0.000; 0.015] |
| Alberta, Canada | 0.002 | [0.000; 0.010] |
| Manitoba, Canada | 0.000 | [0.000; 0.168] |
| Ontario, Canada | 0.000 | [0.000; 0.014] |
| National, US | 0.007 | [0.005; 0.009] |
| **Random effects model** | **0.001** | **[0.000; 0.006]** |

Heterogeneity: $I^2 = 0\%$, $\tau^2 = 1.7457$, $p = 0.99$

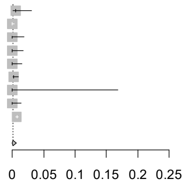

**Antimycotics (J02)**
**Non-hospitalized non-pregnant women with COVID-19**
*30 days pre COVID-19 diagnosis*

| Study Site | Proportion | 95%-CI |
|---|---|---|
| Tuscany, Italy | 0.000 | [0.000; 0.006] |
| Valencia, Spain | 0.000 | [0.000; 0.002] |
| Aragon, Spain | 0.000 | [0.000; 0.007] |
| National, Norway | 0.005 | [0.002; 0.013] |
| National, Sweden | 0.012 | [0.006; 0.022] |
| Alberta, Canada | 0.004 | [0.004; 0.005] |
| Manitoba, Canada | 0.004 | [0.003; 0.006] |
| Ontario, Canada | 0.000 | [0.000; 0.000] |
| National, US | 0.018 | [0.015; 0.023] |
| **Random effects model** | **0.001** | **[0.000; 0.006]** |

Heterogeneity: $I^2 = 95\%$, $\tau^2 = 6.7072$, $p < 0.01$

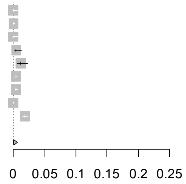

**Antimycotics (J02)**
**Non-hospitalized non-pregnant women with COVID-19**
*30 days post COVID-19 diagnosis*

| Study Site | Proportion | 95%-CI |
|---|---|---|
| Tuscany, Italy | 0.000 | [0.000; 0.006] |
| Valencia, Spain | 0.001 | [0.000; 0.002] |
| Aragon, Spain | 0.002 | [0.000; 0.011] |
| National, Norway | 0.000 | [0.000; 0.006] |
| National, Sweden | 0.004 | [0.001; 0.012] |
| Alberta, Canada | 0.003 | [0.003; 0.004] |
| Manitoba, Canada | 0.000 | [0.000; 0.001] |
| Ontario, Canada | 0.000 | [0.000; 0.000] |
| National, US | 0.020 | [0.017; 0.024] |
| **Random effects model** | **0.000** | **[0.000; 0.003]** |

Heterogeneity: $I^2 = 97\%$, $\tau^2 = 6.0092$, $p < 0.01$

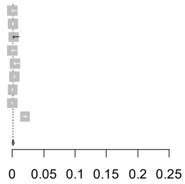

### COVID-19 diagnosis in pregnancy trimester 2

**Antimycotics (J02)**
**Non-hospitalized pregnant women with COVID-19 in 2nd trimester**
*30 days pre COVID-19 diagnosis*

| Study Site | Proportion | 95%-CI |
|---|---|---|
| Tuscany, Italy | 0.000 | [0.000; 0.015] |
| Valencia, Spain | 0.000 | [0.000; 0.004] |
| Aragon, Spain | 0.000 | [0.000; 0.014] |
| National, Norway | 0.003 | [0.000; 0.016] |
| National, Sweden | 0.001 | [0.000; 0.005] |
| Alberta, Canada | 0.000 | [0.000; 0.005] |
| Manitoba, Canada | 0.000 | [0.000; 0.058] |
| Ontario, Canada | 0.000 | [0.000; 0.013] |
| National, US | 0.006 | [0.005; 0.008] |
| **Random effects model** | **0.001** | **[0.000; 0.004]** |

Heterogeneity: $I^2 = 0\%$, $\tau^2 = 1.6504$, $p = 0.77$

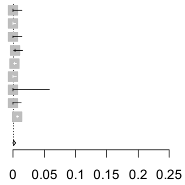

**Antimycotics (J02)**
**Non-hospitalized pregnant women with COVID-19 in 2nd trimester**
*30 days post COVID-19 diagnosis*

| Study Site | Proportion | 95%-CI |
|---|---|---|
| Tuscany, Italy | 0.000 | [0.000; 0.015] |
| Valencia, Spain | 0.000 | [0.000; 0.004] |
| Aragon, Spain | 0.000 | [0.000; 0.014] |
| National, Norway | 0.000 | [0.000; 0.011] |
| National, Sweden | 0.002 | [0.001; 0.006] |
| Alberta, Canada | 0.000 | [0.000; 0.005] |
| Manitoba, Canada | 0.000 | [0.000; 0.058] |
| Ontario, Canada | 0.000 | [0.000; 0.013] |
| National, US | 0.008 | [0.006; 0.010] |
| **Random effects model** | **0.000** | **[0.000; 0.008]** |

Heterogeneity: $I^2 = 0\%$, $\tau^2 = 3.9365$, $p = 0.76$

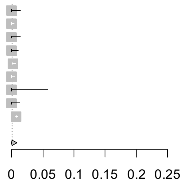

**Antimycotics (J02)**
**Non-hospitalized non-pregnant women with COVID-19**
*30 days pre COVID-19 diagnosis*

| Study Site | Proportion | 95%-CI |
|---|---|---|
| Tuscany, Italy | 0.001 | [0.000; 0.007] |
| Valencia, Spain | 0.001 | [0.000; 0.003] |
| Aragon, Spain | 0.003 | [0.001; 0.009] |
| National, Norway | 0.003 | [0.001; 0.009] |
| National, Sweden | 0.006 | [0.004; 0.009] |
| Alberta, Canada | 0.004 | [0.004; 0.005] |
| Manitoba, Canada | 0.004 | [0.003; 0.006] |
| Ontario, Canada | 0.000 | [0.000; 0.000] |
| National, US | 0.017 | [0.014; 0.020] |
| **Random effects model** | **0.002** | **[0.000; 0.006]** |

Heterogeneity: $I^2 = 96\%$, $\tau^2 = 3.4393$, $p < 0.01$

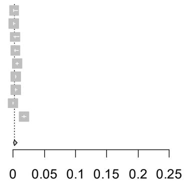

**Antimycotics (J02)**
**Non-hospitalized non-pregnant women with COVID-19**
*30 days post COVID-19 diagnosis*

| Study Site | Proportion | 95%-CI |
|---|---|---|
| Tuscany, Italy | 0.002 | [0.001; 0.009] |
| Valencia, Spain | 0.000 | [0.000; 0.002] |
| Aragon, Spain | 0.000 | [0.000; 0.005] |
| National, Norway | 0.003 | [0.001; 0.007] |
| National, Sweden | 0.003 | [0.001; 0.005] |
| Alberta, Canada | 0.003 | [0.003; 0.004] |
| Manitoba, Canada | 0.000 | [0.000; 0.001] |
| Ontario, Canada | 0.000 | [0.000; 0.000] |
| National, US | 0.017 | [0.014; 0.021] |
| **Random effects model** | **0.001** | **[0.000; 0.003]** |

Heterogeneity: $I^2 = 97\%$, $\tau^2 = 4.9797$, $p < 0.01$

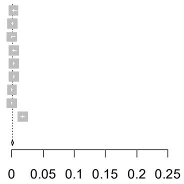

**Figure S30 continued.** Forest plots showing the pooled prevalence of antimycotics in the 30 days pre-COVID (left) and 30 days post-COVID (right) in non-hospitalized pregnant women with COVID-19 (upper) and non-hospitalized non-pregnant women with COVID-19 (lower), by pregnancy trimester

### COVID-19 diagnosis in pregnancy trimester 3

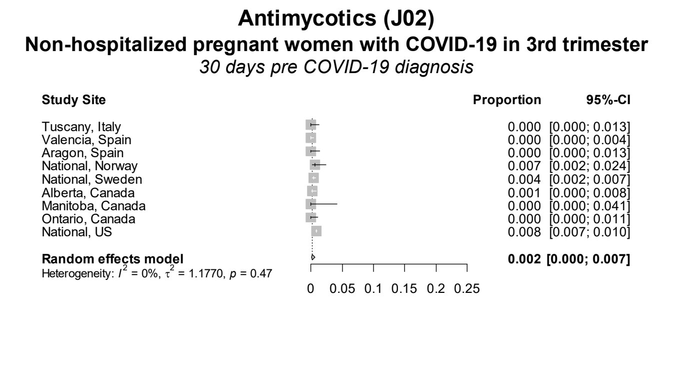

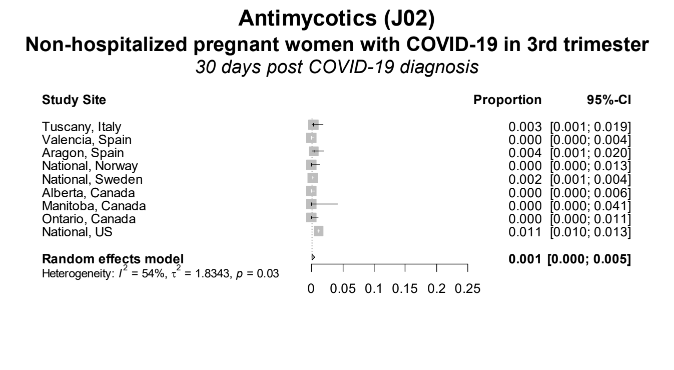

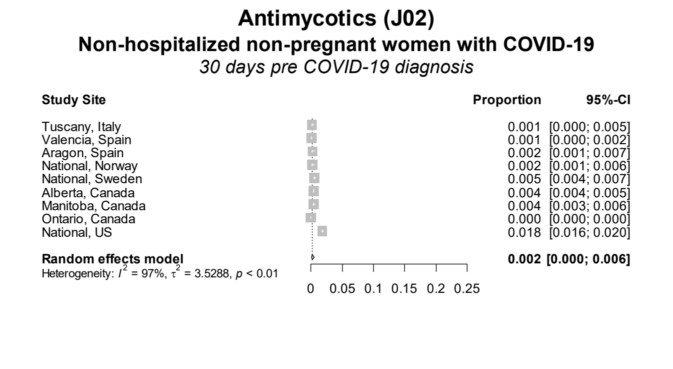

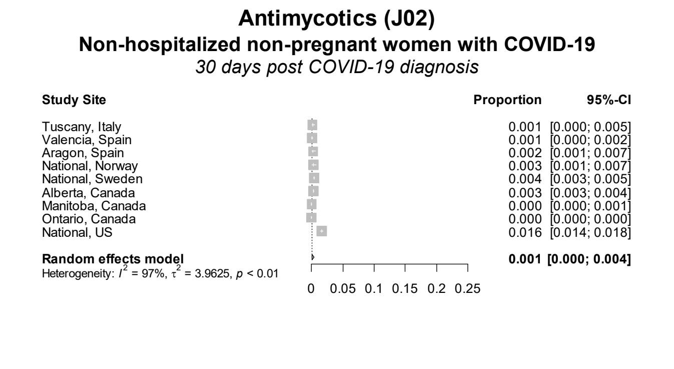

Figure S31. Forest plots showing the pooled prevalence of antineoplastic agents in the 30 days pre-COVID (left) and 30 days post-COVID (right) in non-hospitalized pregnant women with COVID-19 (upper) and non-hospitalized non-pregnant women with COVID-19 (lower), by pregnancy trimester

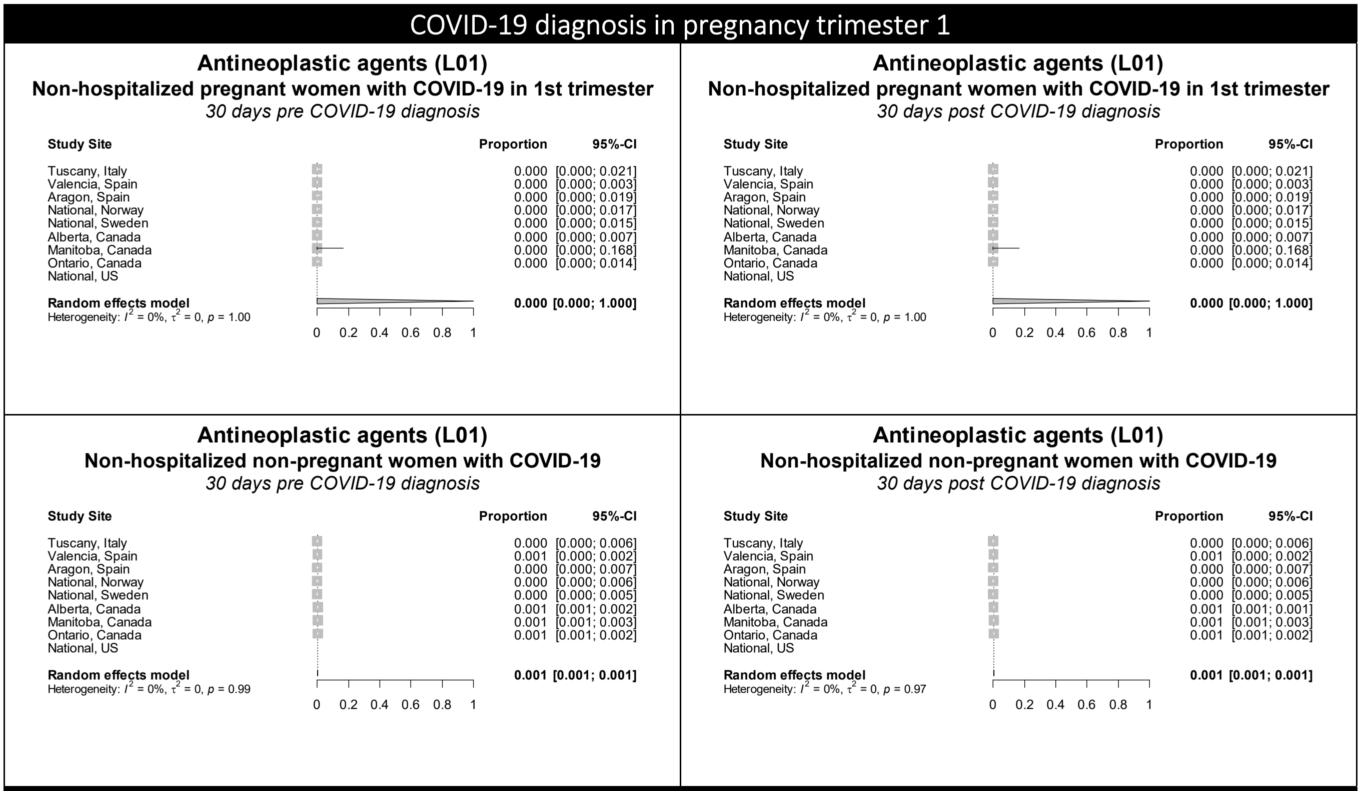

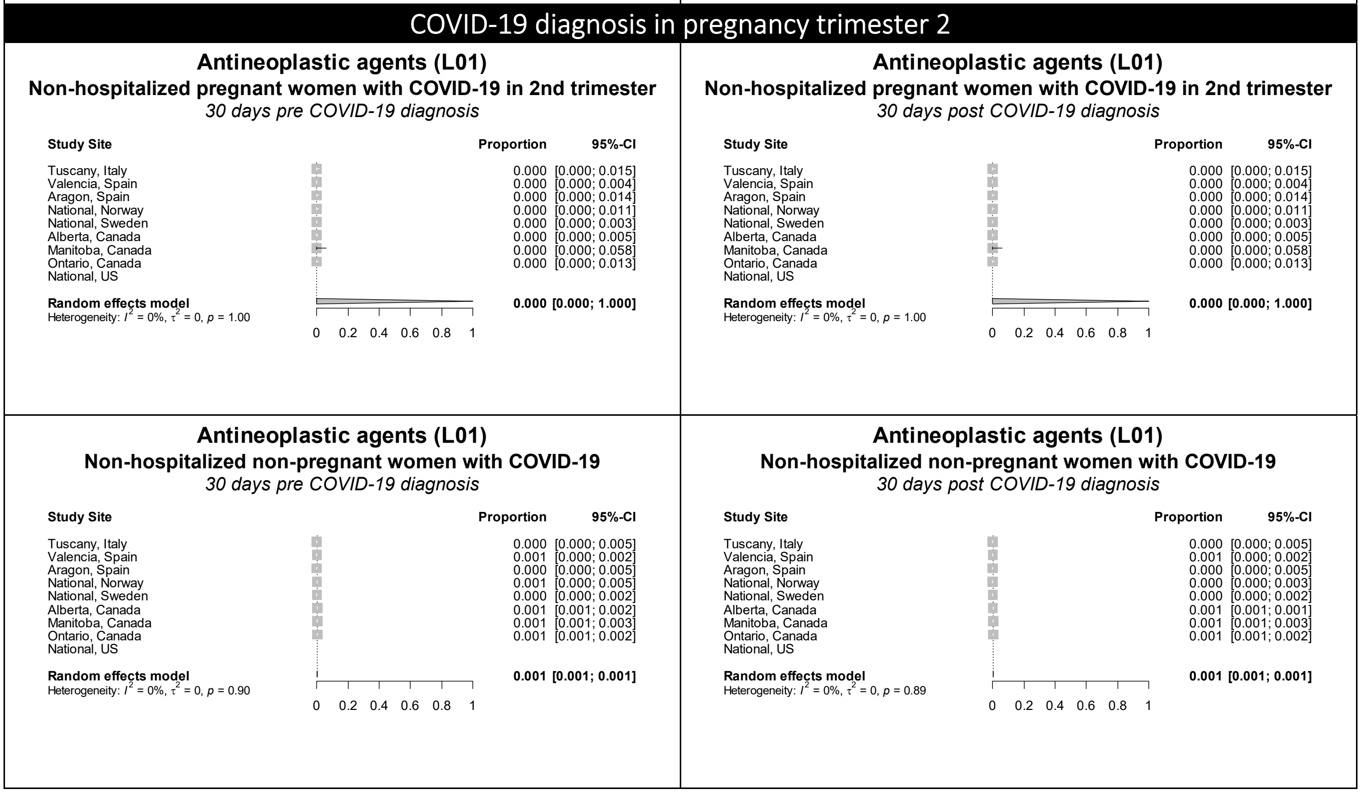

Figure S31 continued. Forest plots showing the pooled prevalence of antineoplastic agents in the 30 days pre-COVID (left) and 30 days post-COVID (right) in non-hospitalized pregnant women with COVID-19 (upper) and non-hospitalized non-pregnant women with COVID-19 (lower), by pregnancy trimester

### COVID-19 diagnosis in pregnancy trimester 3

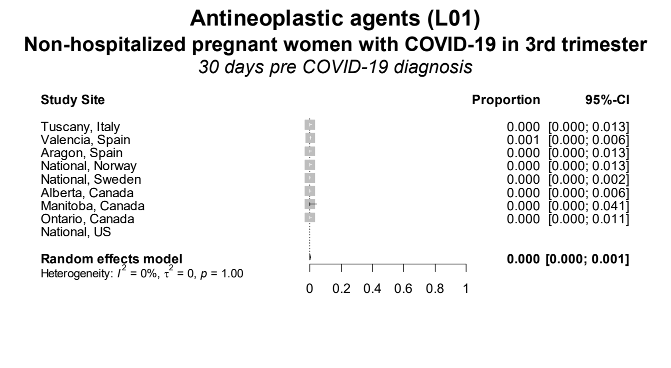

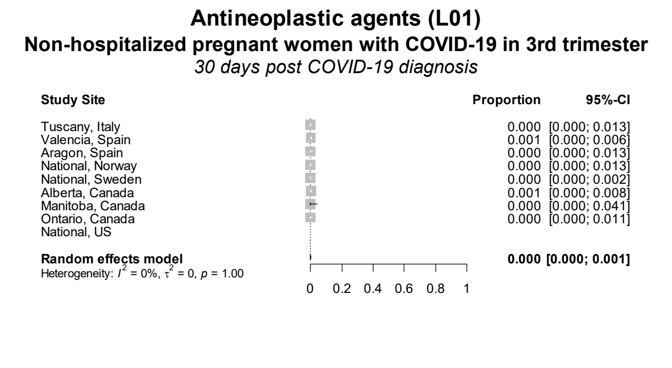

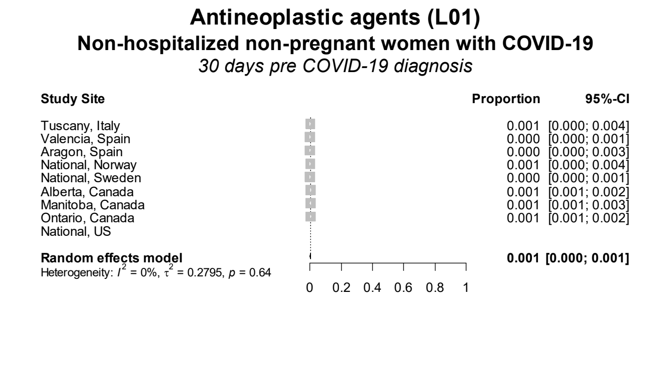

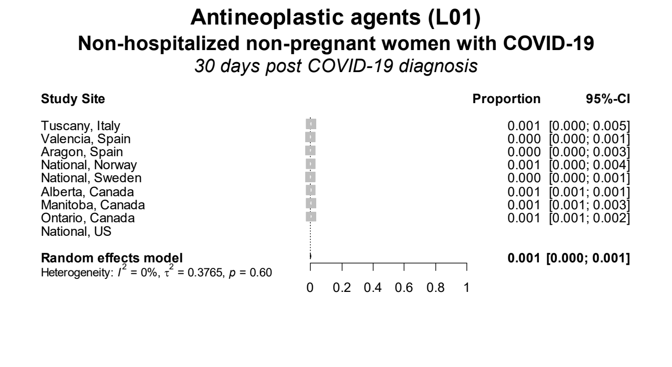

Figure S32. Forest plots showing the pooled prevalence of antiprotozoals in the 30 days pre-COVID (left) and 30 days post-COVID (right) in non-hospitalized pregnant women with COVID-19 (upper) and non-hospitalized non-pregnant women with COVID-19 (lower), by pregnancy trimester

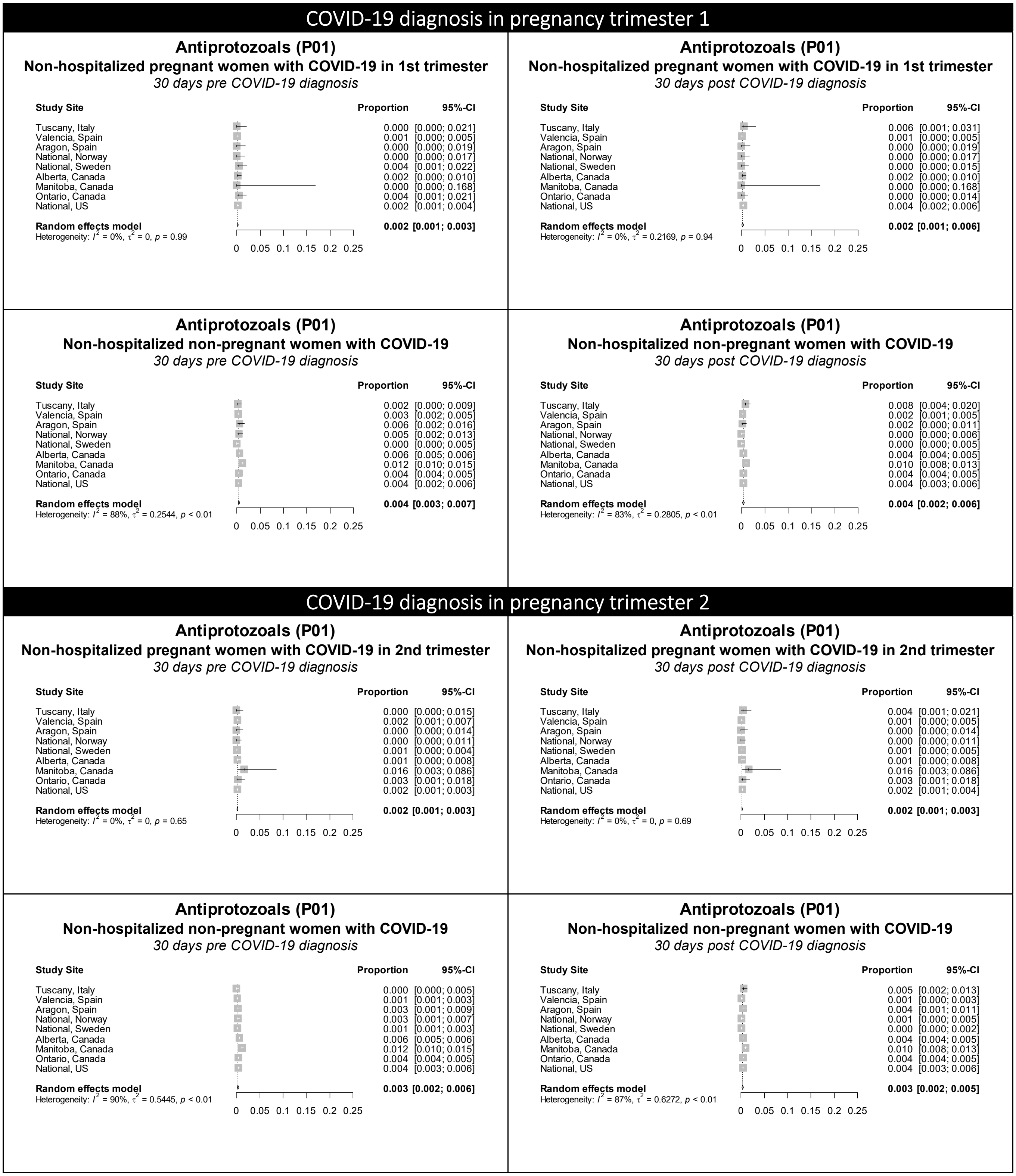

**Figure S32 continued.** Forest plots showing the pooled prevalence of antiprotozoals in the 30 days pre-COVID (left) and 30 days post-COVID (right) in non-hospitalized pregnant women with COVID-19 (upper) and non-hospitalized non-pregnant women with COVID-19 (lower), by pregnancy trimester

### COVID-19 diagnosis in pregnancy trimester 3

#### Antiprotozoals (P01)
**Non-hospitalized pregnant women with COVID-19 in 3rd trimester**
*30 days pre COVID-19 diagnosis*

| Study Site | Proportion | 95%-CI |
|---|---|---|
| Tuscany, Italy | 0.000 | [0.000; 0.013] |
| Valencia, Spain | 0.002 | [0.001; 0.008] |
| Aragon, Spain | 0.004 | [0.001; 0.020] |
| National, Norway | 0.003 | [0.001; 0.019] |
| National, Sweden | 0.000 | [0.000; 0.002] |
| Alberta, Canada | 0.001 | [0.000; 0.008] |
| Manitoba, Canada | 0.011 | [0.002; 0.061] |
| Ontario, Canada | 0.003 | [0.000; 0.016] |
| National, US | 0.001 | [0.001; 0.002] |
| **Random effects model** | **0.001** | **[0.001; 0.002]** |

Heterogeneity: $I^2 = 20\%$, $\tau^2 = 0$, $p = 0.27$

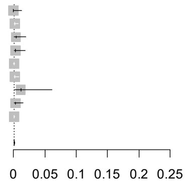

#### Antiprotozoals (P01)
**Non-hospitalized pregnant women with COVID-19 in 3rd trimester**
*30 days post COVID-19 diagnosis*

| Study Site | Proportion | 95%-CI |
|---|---|---|
| Tuscany, Italy | 0.000 | [0.000; 0.013] |
| Valencia, Spain | 0.000 | [0.000; 0.004] |
| Aragon, Spain | 0.004 | [0.001; 0.020] |
| National, Norway | 0.013 | [0.005; 0.034] |
| National, Sweden | 0.021 | [0.016; 0.028] |
| Alberta, Canada | 0.001 | [0.000; 0.008] |
| Manitoba, Canada | 0.011 | [0.002; 0.061] |
| Ontario, Canada | 0.000 | [0.000; 0.011] |
| National, US | 0.001 | [0.001; 0.002] |
| **Random effects model** | **0.002** | **[0.001; 0.008]** |

Heterogeneity: $I^2 = 93\%$, $\tau^2 = 1.9728$, $p < 0.01$

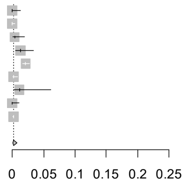

#### Antiprotozoals (P01)
**Non-hospitalized non-pregnant women with COVID-19**
*30 days pre COVID-19 diagnosis*

| Study Site | Proportion | 95%-CI |
|---|---|---|
| Tuscany, Italy | 0.003 | [0.001; 0.007] |
| Valencia, Spain | 0.002 | [0.001; 0.003] |
| Aragon, Spain | 0.002 | [0.001; 0.007] |
| National, Norway | 0.001 | [0.000; 0.004] |
| National, Sweden | 0.001 | [0.001; 0.002] |
| Alberta, Canada | 0.006 | [0.005; 0.006] |
| Manitoba, Canada | 0.012 | [0.010; 0.015] |
| Ontario, Canada | 0.004 | [0.004; 0.005] |
| National, US | 0.004 | [0.003; 0.005] |
| **Random effects model** | **0.003** | **[0.002; 0.005]** |

Heterogeneity: $I^2 = 92\%$, $\tau^2 = 0.5303$, $p < 0.01$

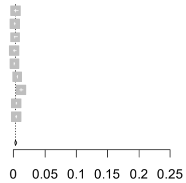

#### Antiprotozoals (P01)
**Non-hospitalized non-pregnant women with COVID-19**
*30 days post COVID-19 diagnosis*

| Study Site | Proportion | 95%-CI |
|---|---|---|
| Tuscany, Italy | 0.007 | [0.004; 0.013] |
| Valencia, Spain | 0.001 | [0.000; 0.003] |
| Aragon, Spain | 0.001 | [0.000; 0.004] |
| National, Norway | 0.001 | [0.000; 0.004] |
| National, Sweden | 0.002 | [0.001; 0.003] |
| Alberta, Canada | 0.004 | [0.004; 0.005] |
| Manitoba, Canada | 0.010 | [0.008; 0.013] |
| Ontario, Canada | 0.004 | [0.004; 0.005] |
| National, US | 0.004 | [0.003; 0.005] |
| **Random effects model** | **0.003** | **[0.002; 0.005]** |

Heterogeneity: $I^2 = 89\%$, $\tau^2 = 0.5514$, $p < 0.01$

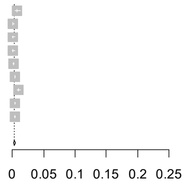

Figure S33. Forest plots showing the pooled prevalence of antithrombotic agents in the 30 days pre-COVID (left) and 30 days post-COVID (right) in non-hospitalized pregnant women with COVID-19 (upper) and non-hospitalized non-pregnant women with COVID-19 (lower), by pregnancy trimester

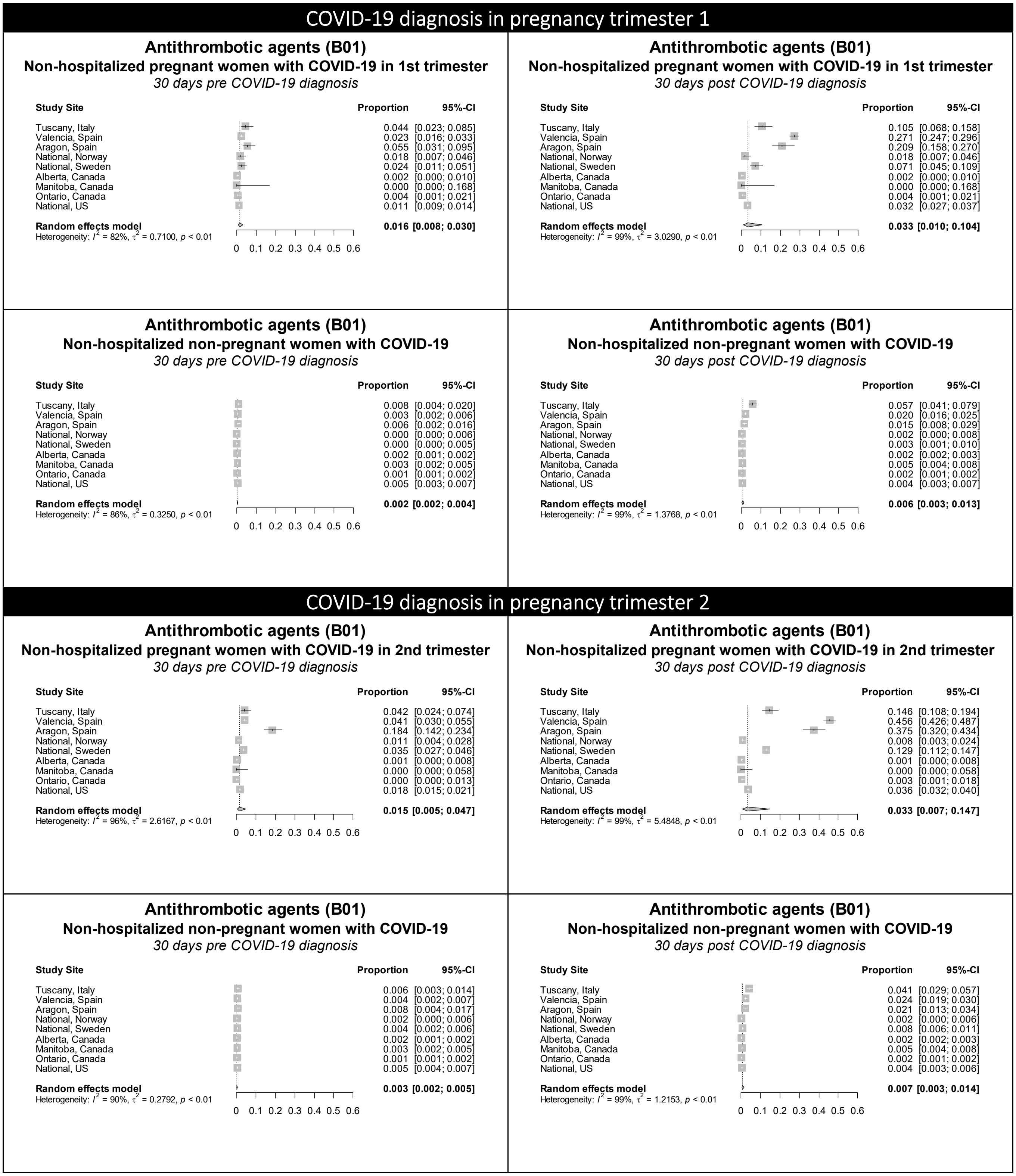

**Figure S33 continued.** Forest plots showing the pooled prevalence of antithrombotic agents in the 30 days pre-COVID (left) and 30 days post-COVID (right) in non-hospitalized pregnant women with COVID-19 (upper) and non-hospitalized non-pregnant women with COVID-19 (lower), by pregnancy trimester

### COVID-19 diagnosis in pregnancy trimester 3

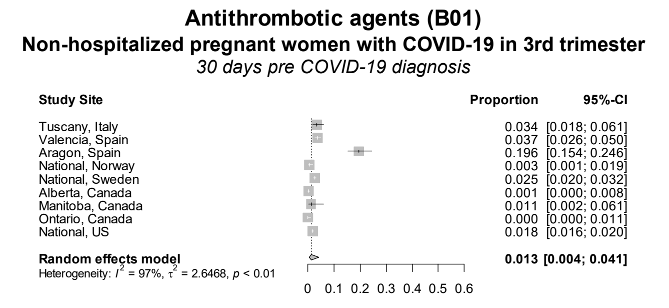

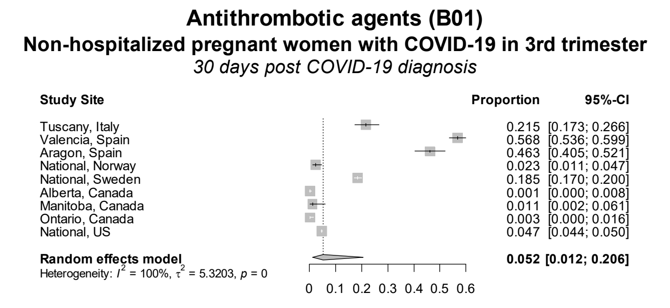

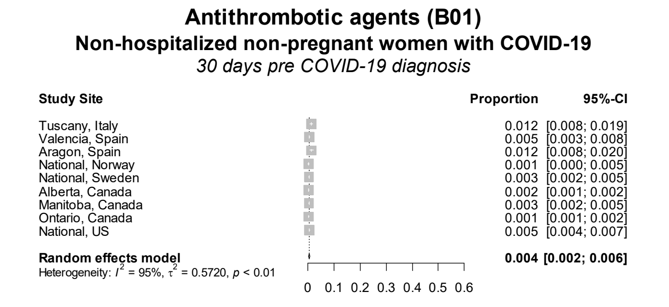

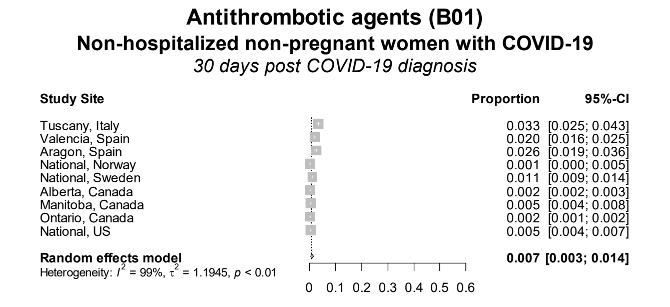

Figure S34. Forest plots showing the pooled prevalence of antivirals in the 30 days pre-COVID (left) and 30 days post-COVID (right) in non-hospitalized pregnant women with COVID-19 (upper) and non-hospitalized non-pregnant women with COVID-19 (lower), by pregnancy trimester

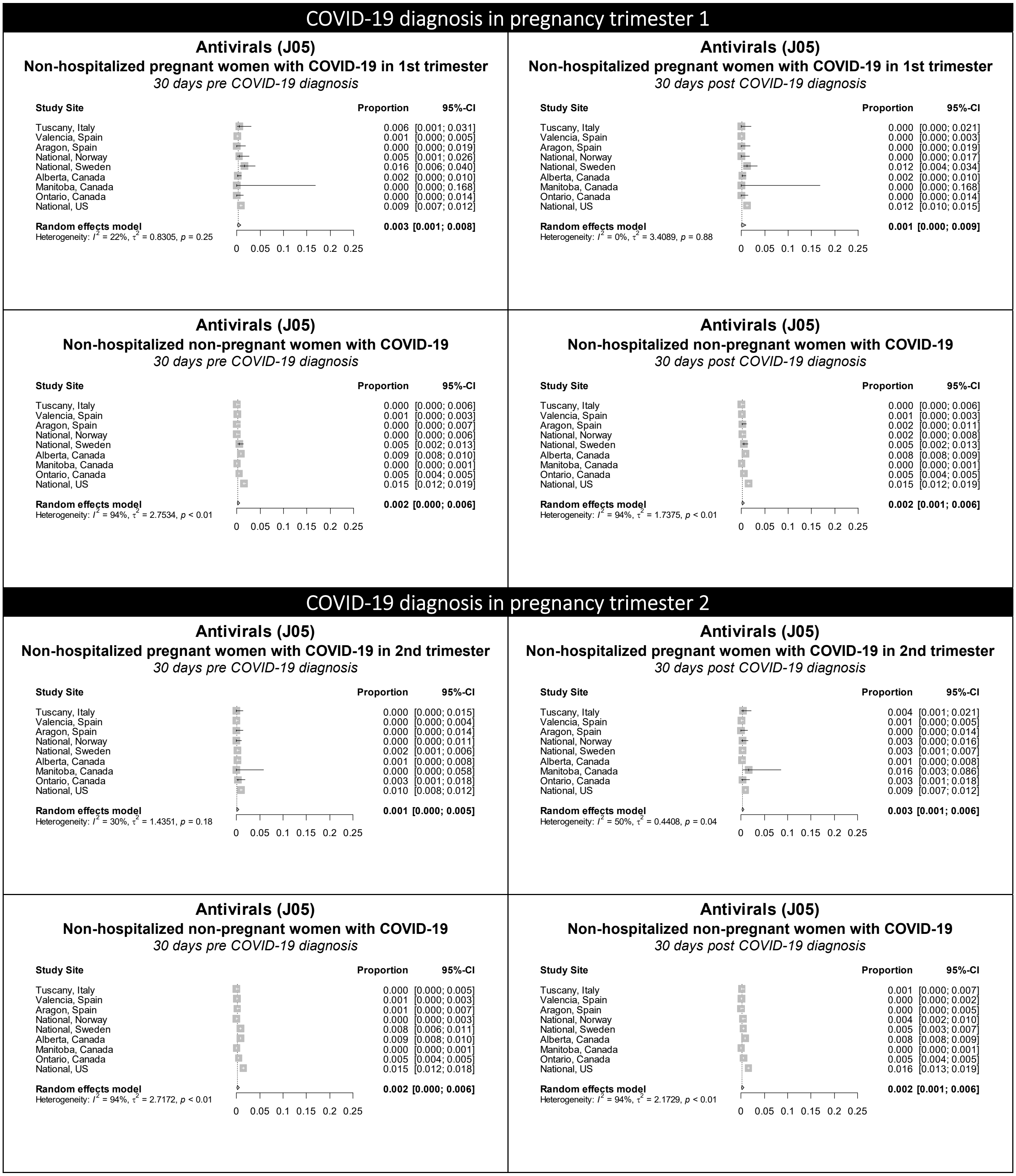

**Figure S34 continued.** Forest plots showing the pooled prevalence of antivirals in the 30 days pre-COVID (left) and 30 days post-COVID (right) in non-hospitalized pregnant women with COVID-19 (upper) and non-hospitalized non-pregnant women with COVID-19 (lower), by pregnancy trimester

### COVID-19 diagnosis in pregnancy trimester 3

#### Antivirals (J05)
**Non-hospitalized pregnant women with COVID-19 in 3rd trimester**
*30 days pre COVID-19 diagnosis*

| Study Site | Proportion | 95%-CI |
|---|---|---|
| Tuscany, Italy | 0.003 | [0.001; 0.019] |
| Valencia, Spain | 0.000 | [0.000; 0.004] |
| Aragon, Spain | 0.000 | [0.000; 0.013] |
| National, Norway | 0.007 | [0.002; 0.024] |
| National, Sweden | 0.008 | [0.005; 0.013] |
| Alberta, Canada | 0.001 | [0.000; 0.008] |
| Manitoba, Canada | 0.011 | [0.002; 0.061] |
| Ontario, Canada | 0.003 | [0.000; 0.016] |
| National, US | 0.021 | [0.018; 0.023] |
| **Random effects model** | **0.004** | **[0.001; 0.010]** |

Heterogeneity: $I^2 = 75\%$, $\tau^2 = 1.2350$, $p < 0.01$

Axis: 0 0.05 0.1 0.15 0.2 0.25

#### Antivirals (J05)
**Non-hospitalized pregnant women with COVID-19 in 3rd trimester**
*30 days post COVID-19 diagnosis*

| Study Site | Proportion | 95%-CI |
|---|---|---|
| Tuscany, Italy | 0.000 | [0.000; 0.013] |
| Valencia, Spain | 0.001 | [0.000; 0.006] |
| Aragon, Spain | 0.000 | [0.000; 0.013] |
| National, Norway | 0.003 | [0.001; 0.019] |
| National, Sweden | 0.006 | [0.004; 0.011] |
| Alberta, Canada | 0.016 | [0.009; 0.028] |
| Manitoba, Canada | 0.011 | [0.002; 0.061] |
| Ontario, Canada | 0.003 | [0.000; 0.016] |
| National, US | 0.021 | [0.019; 0.023] |
| **Random effects model** | **0.004** | **[0.002; 0.011]** |

Heterogeneity: $I^2 = 78\%$, $\tau^2 = 1.1050$, $p < 0.01$

Axis: 0 0.05 0.1 0.15 0.2 0.25

#### Antivirals (J05)
**Non-hospitalized non-pregnant women with COVID-19**
*30 days pre COVID-19 diagnosis*

| Study Site | Proportion | 95%-CI |
|---|---|---|
| Tuscany, Italy | 0.000 | [0.000; 0.003] |
| Valencia, Spain | 0.001 | [0.001; 0.003] |
| Aragon, Spain | 0.000 | [0.000; 0.003] |
| National, Norway | 0.004 | [0.002; 0.009] |
| National, Sweden | 0.006 | [0.005; 0.009] |
| Alberta, Canada | 0.009 | [0.008; 0.010] |
| Manitoba, Canada | 0.000 | [0.000; 0.001] |
| Ontario, Canada | 0.005 | [0.004; 0.005] |
| National, US | 0.014 | [0.012; 0.016] |
| **Random effects model** | **0.002** | **[0.001; 0.006]** |

Heterogeneity: $I^2 = 95\%$, $\tau^2 = 2.5442$, $p < 0.01$

Axis: 0 0.05 0.1 0.15 0.2 0.25

#### Antivirals (J05)
**Non-hospitalized non-pregnant women with COVID-19**
*30 days post COVID-19 diagnosis*

| Study Site | Proportion | 95%-CI |
|---|---|---|
| Tuscany, Italy | 0.001 | [0.000; 0.005] |
| Valencia, Spain | 0.002 | [0.001; 0.004] |
| Aragon, Spain | 0.000 | [0.000; 0.003] |
| National, Norway | 0.004 | [0.002; 0.009] |
| National, Sweden | 0.004 | [0.003; 0.006] |
| Alberta, Canada | 0.008 | [0.008; 0.009] |
| Manitoba, Canada | 0.000 | [0.000; 0.001] |
| Ontario, Canada | 0.005 | [0.004; 0.005] |
| National, US | 0.013 | [0.011; 0.015] |
| **Random effects model** | **0.003** | **[0.001; 0.006]** |

Heterogeneity: $I^2 = 94\%$, $\tau^2 = 1.4864$, $p < 0.01$

Axis: 0 0.05 0.1 0.15 0.2 0.25

Figure S35. Forest plots showing the pooled prevalence of corticosteroids in the 30 days pre-COVID (left) and 30 days post-COVID (right) in non-hospitalized pregnant women with COVID-19 (upper) and non-hospitalized non-pregnant women with COVID-19 (lower), by pregnancy trimester

### COVID-19 diagnosis in pregnancy trimester 1

#### Corticosteroids (H02)
**Non-hospitalized pregnant women with COVID-19 in 1st trimester**
*30 days pre COVID-19 diagnosis*

| Study Site | Proportion | 95%-CI |
|---|---|---|
| Tuscany, Italy | 0.017 | [0.006; 0.048] |
| Valencia, Spain | 0.004 | [0.002; 0.009] |
| Aragon, Spain | 0.000 | [0.000; 0.019] |
| National, Norway | 0.009 | [0.003; 0.033] |
| National, Sweden | 0.000 | [0.000; 0.015] |
| Alberta, Canada | 0.002 | [0.000; 0.010] |
| Manitoba, Canada | 0.000 | [0.000; 0.168] |
| Ontario, Canada | 0.000 | [0.000; 0.014] |
| National, US | 0.025 | [0.022; 0.030] |
| **Random effects model** | **0.004** | **[0.001; 0.012]** |

Heterogeneity: $I^2 = 69\%$, $\tau^2 = 1.3659$, $p < 0.01$

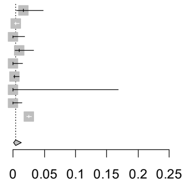

#### Corticosteroids (H02)
**Non-hospitalized pregnant women with COVID-19 in 1st trimester**
*30 days post COVID-19 diagnosis*

| Study Site | Proportion | 95%-CI |
|---|---|---|
| Tuscany, Italy | 0.050 | [0.026; 0.092] |
| Valencia, Spain | 0.008 | [0.004; 0.015] |
| Aragon, Spain | 0.025 | [0.011; 0.057] |
| National, Norway | 0.005 | [0.001; 0.026] |
| National, Sweden | 0.000 | [0.000; 0.015] |
| Alberta, Canada | 0.002 | [0.000; 0.010] |
| Manitoba, Canada | 0.000 | [0.000; 0.168] |
| Ontario, Canada | 0.004 | [0.001; 0.021] |
| National, US | 0.054 | [0.048; 0.060] |
| **Random effects model** | **0.008** | **[0.003; 0.025]** |

Heterogeneity: $I^2 = 87\%$, $\tau^2 = 1.7516$, $p < 0.01$

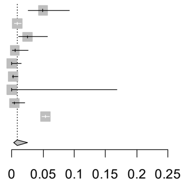

#### Corticosteroids (H02)
**Non-hospitalized non-pregnant women with COVID-19**
*30 days pre COVID-19 diagnosis*

| Study Site | Proportion | 95%-CI |
|---|---|---|
| Tuscany, Italy | 0.039 | [0.026; 0.057] |
| Valencia, Spain | 0.005 | [0.003; 0.008] |
| Aragon, Spain | 0.015 | [0.008; 0.029] |
| National, Norway | 0.005 | [0.002; 0.013] |
| National, Sweden | 0.004 | [0.001; 0.012] |
| Alberta, Canada | 0.006 | [0.006; 0.007] |
| Manitoba, Canada | 0.006 | [0.005; 0.008] |
| Ontario, Canada | 0.004 | [0.004; 0.005] |
| National, US | 0.078 | [0.071; 0.086] |
| **Random effects model** | **0.010** | **[0.005; 0.019]** |

Heterogeneity: $I^2 = 100\%$, $\tau^2 = 1.1075$, $p = 0$

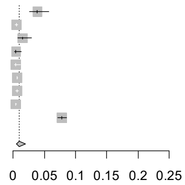

#### Corticosteroids (H02)
**Non-hospitalized non-pregnant women with COVID-19**
*30 days post COVID-19 diagnosis*

| Study Site | Proportion | 95%-CI |
|---|---|---|
| Tuscany, Italy | 0.128 | [0.103; 0.157] |
| Valencia, Spain | 0.012 | [0.009; 0.016] |
| Aragon, Spain | 0.021 | [0.012; 0.037] |
| National, Norway | 0.009 | [0.004; 0.020] |
| National, Sweden | 0.010 | [0.005; 0.021] |
| Alberta, Canada | 0.020 | [0.019; 0.021] |
| Manitoba, Canada | 0.015 | [0.012; 0.018] |
| Ontario, Canada | 0.009 | [0.008; 0.010] |
| National, US | 0.101 | [0.093; 0.109] |
| **Random effects model** | **0.021** | **[0.011; 0.041]** |

Heterogeneity: $I^2 = 100\%$, $\tau^2 = 0.9972$, $p = 0$

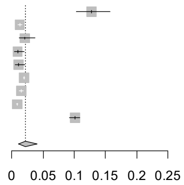

### COVID-19 diagnosis in pregnancy trimester 2

#### Corticosteroids (H02)
**Non-hospitalized pregnant women with COVID-19 in 2nd trimester**
*30 days pre COVID-19 diagnosis*

| Study Site | Proportion | 95%-CI |
|---|---|---|
| Tuscany, Italy | 0.015 | [0.006; 0.039] |
| Valencia, Spain | 0.000 | [0.000; 0.004] |
| Aragon, Spain | 0.004 | [0.001; 0.021] |
| National, Norway | 0.000 | [0.000; 0.011] |
| National, Sweden | 0.002 | [0.001; 0.006] |
| Alberta, Canada | 0.001 | [0.000; 0.008] |
| Manitoba, Canada | 0.000 | [0.000; 0.058] |
| Ontario, Canada | 0.000 | [0.000; 0.013] |
| National, US | 0.009 | [0.007; 0.011] |
| **Random effects model** | **0.002** | **[0.001; 0.007]** |

Heterogeneity: $I^2 = 29\%$, $\tau^2 = 1.4449$, $p = 0.18$

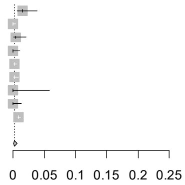

#### Corticosteroids (H02)
**Non-hospitalized pregnant women with COVID-19 in 2nd trimester**
*30 days post COVID-19 diagnosis*

| Study Site | Proportion | 95%-CI |
|---|---|---|
| Tuscany, Italy | 0.042 | [0.024; 0.074] |
| Valencia, Spain | 0.005 | [0.002; 0.011] |
| Aragon, Spain | 0.007 | [0.002; 0.026] |
| National, Norway | 0.000 | [0.000; 0.011] |
| National, Sweden | 0.003 | [0.001; 0.007] |
| Alberta, Canada | 0.001 | [0.000; 0.008] |
| Manitoba, Canada | 0.000 | [0.000; 0.058] |
| Ontario, Canada | 0.003 | [0.001; 0.018] |
| National, US | 0.038 | [0.034; 0.043] |
| **Random effects model** | **0.005** | **[0.002; 0.015]** |

Heterogeneity: $I^2 = 89\%$, $\tau^2 = 1.7889$, $p < 0.01$

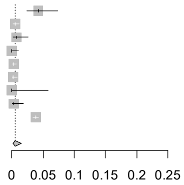

#### Corticosteroids (H02)
**Non-hospitalized non-pregnant women with COVID-19**
*30 days pre COVID-19 diagnosis*

| Study Site | Proportion | 95%-CI |
|---|---|---|
| Tuscany, Italy | 0.037 | [0.026; 0.052] |
| Valencia, Spain | 0.004 | [0.002; 0.007] |
| Aragon, Spain | 0.009 | [0.004; 0.019] |
| National, Norway | 0.003 | [0.001; 0.007] |
| National, Sweden | 0.003 | [0.002; 0.005] |
| Alberta, Canada | 0.006 | [0.006; 0.007] |
| Manitoba, Canada | 0.006 | [0.005; 0.008] |
| Ontario, Canada | 0.004 | [0.004; 0.005] |
| National, US | 0.071 | [0.065; 0.077] |
| **Random effects model** | **0.008** | **[0.004; 0.017]** |

Heterogeneity: $I^2 = 100\%$, $\tau^2 = 1.1782$, $p = 0$

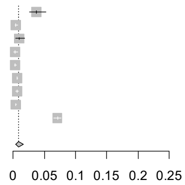

#### Corticosteroids (H02)
**Non-hospitalized non-pregnant women with COVID-19**
*30 days post COVID-19 diagnosis*

| Study Site | Proportion | 95%-CI |
|---|---|---|
| Tuscany, Italy | 0.123 | [0.103; 0.148] |
| Valencia, Spain | 0.016 | [0.012; 0.021] |
| Aragon, Spain | 0.042 | [0.030; 0.059] |
| National, Norway | 0.005 | [0.002; 0.011] |
| National, Sweden | 0.006 | [0.004; 0.009] |
| Alberta, Canada | 0.020 | [0.019; 0.021] |
| Manitoba, Canada | 0.015 | [0.012; 0.018] |
| Ontario, Canada | 0.009 | [0.008; 0.010] |
| National, US | 0.095 | [0.088; 0.102] |
| **Random effects model** | **0.021** | **[0.010; 0.042]** |

Heterogeneity: $I^2 = 100\%$, $\tau^2 = 1.2180$, $p = 0$

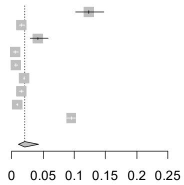

**Figure S35 continued.** Forest plots showing the pooled prevalence of corticosteroids in the 30 days pre-COVID (left) and 30 days post-COVID (right) in non-hospitalized pregnant women with COVID-19 (upper) and non-hospitalized non-pregnant women with COVID-19 (lower), by pregnancy trimester

### COVID-19 diagnosis in pregnancy trimester 3

#### Corticosteroids (H02)
**Non-hospitalized pregnant women with COVID-19 in 3rd trimester**
*30 days pre COVID-19 diagnosis*

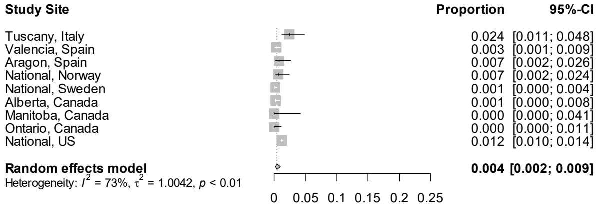

#### Corticosteroids (H02)
**Non-hospitalized pregnant women with COVID-19 in 3rd trimester**
*30 days post COVID-19 diagnosis*

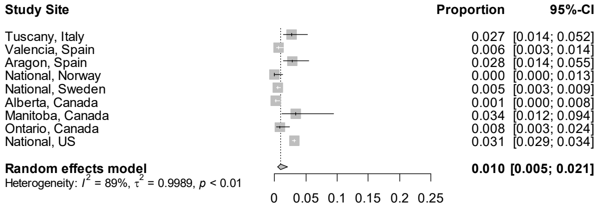

#### Corticosteroids (H02)
**Non-hospitalized non-pregnant women with COVID-19**
*30 days pre COVID-19 diagnosis*

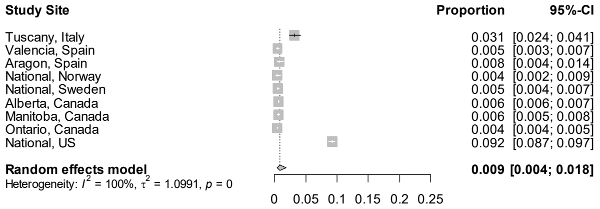

#### Corticosteroids (H02)
**Non-hospitalized non-pregnant women with COVID-19**
*30 days post COVID-19 diagnosis*

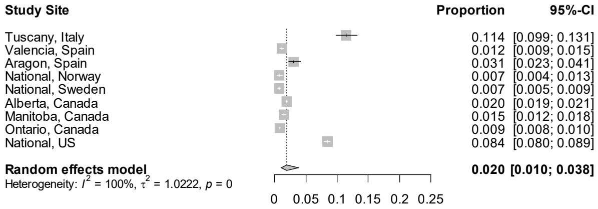

Figure S36. Forest plots showing the pooled prevalence of cough and cold preparations in the 30 days pre-COVID (left) and 30 days post-COVID (right) in non-hospitalized pregnant women with COVID-19 (upper) and non-hospitalized non-pregnant women with COVID-19 (lower), by pregnancy trimester

### COVID-19 diagnosis in pregnancy trimester 1

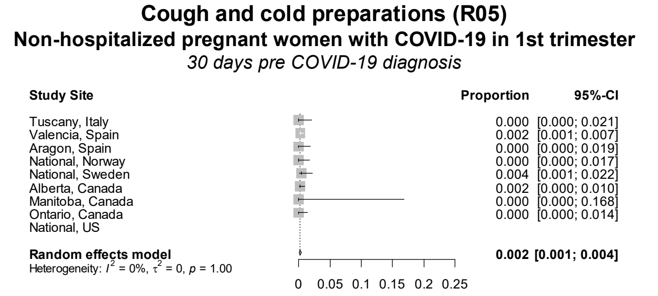

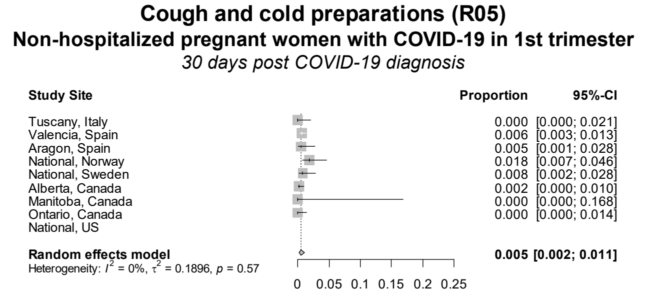

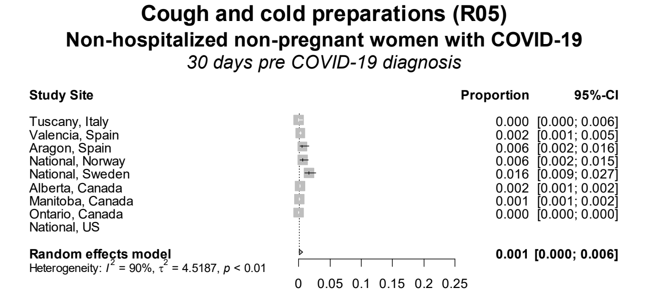

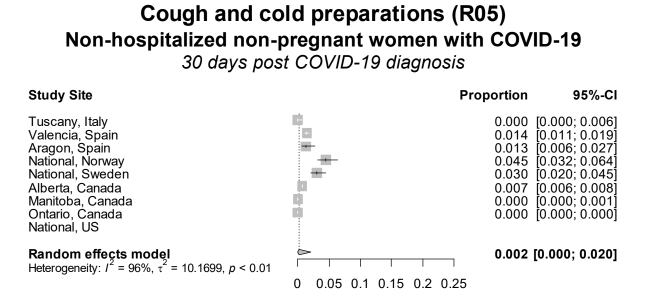

### COVID-19 diagnosis in pregnancy trimester 2

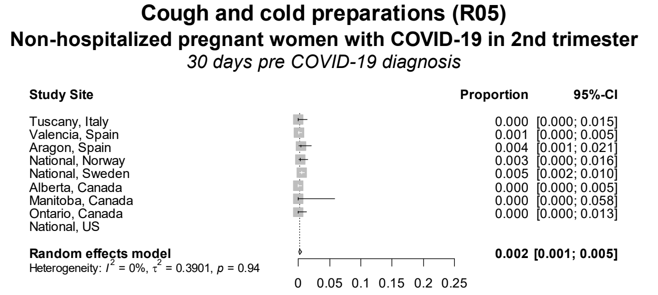

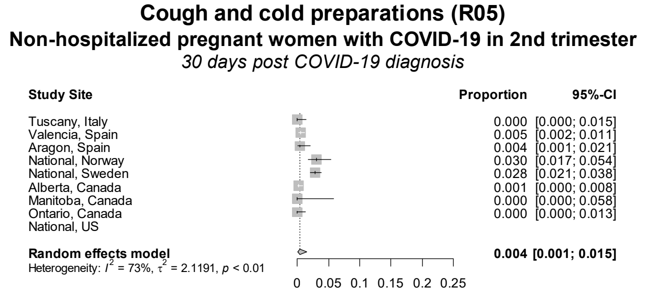

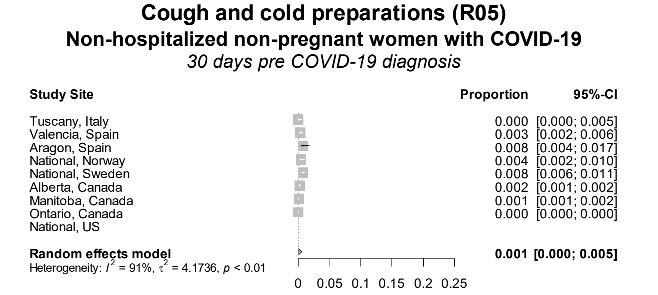

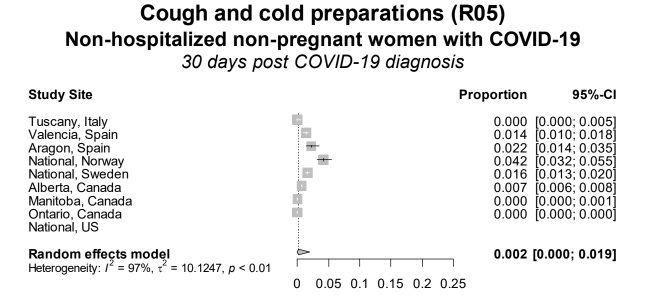

Figure S36 continued. Forest plots showing the pooled prevalence of cough and cold preparations in the 30 days pre-COVID (left) and 30 days post-COVID (right) in non-hospitalized pregnant women with COVID-19 (upper) and non-hospitalized non-pregnant women with COVID-19 (lower), by pregnancy trimester

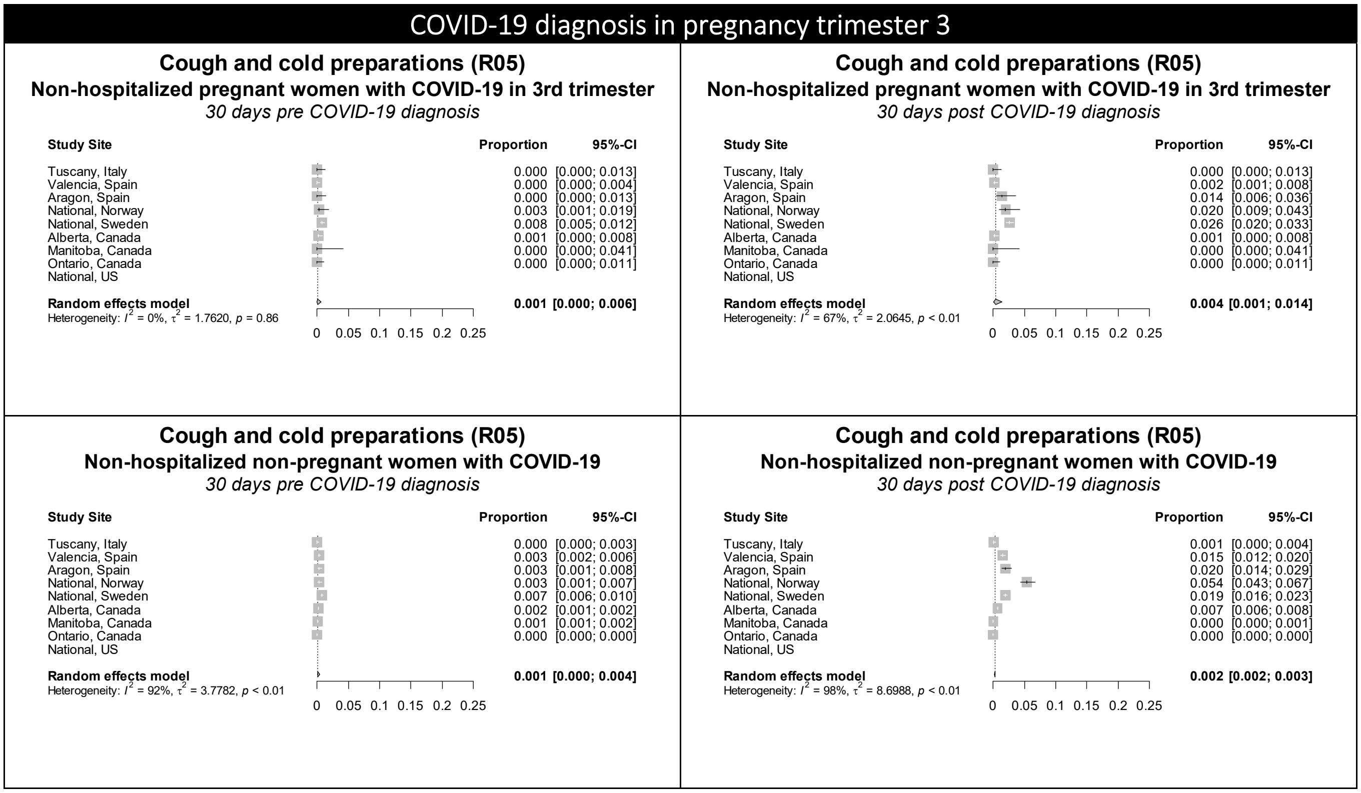

Figure S37. Forest plots showing the pooled prevalence of drugs for obstructive airway diseases in the 30 days pre-COVID (left) and 30 days post-COVID (right) in non-hospitalized pregnant women with COVID-19 (upper) and non-hospitalized non-pregnant women with COVID-19 (lower), by pregnancy trimester

### COVID-19 diagnosis in pregnancy trimester 1

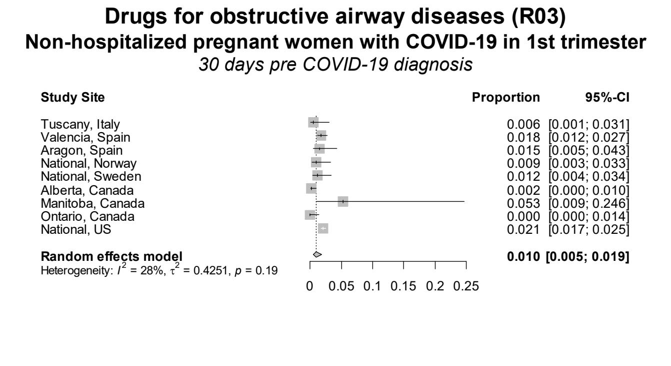

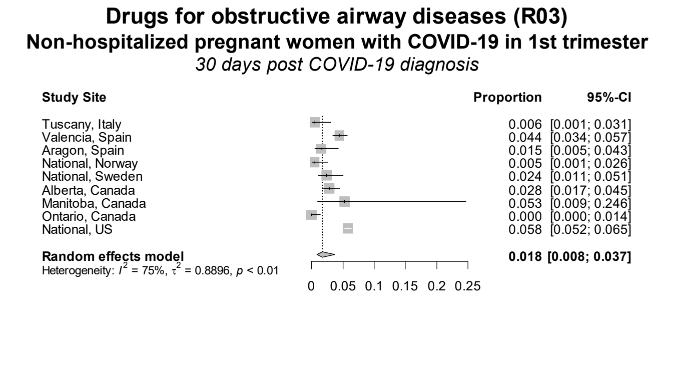

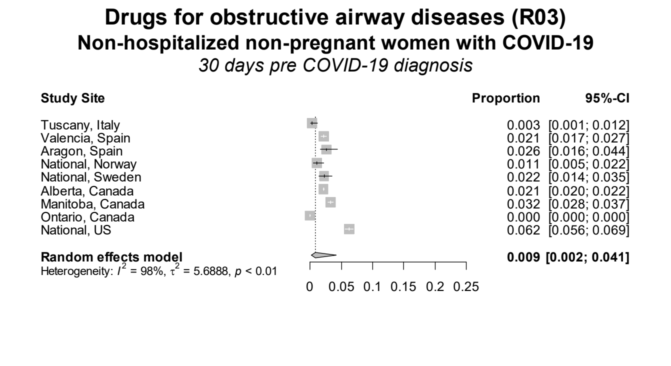

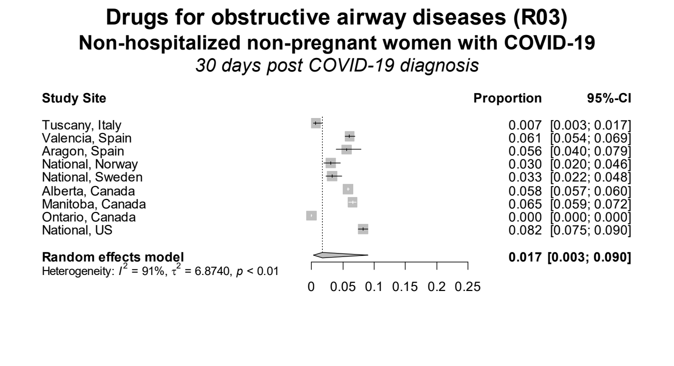

### COVID-19 diagnosis in pregnancy trimester 2

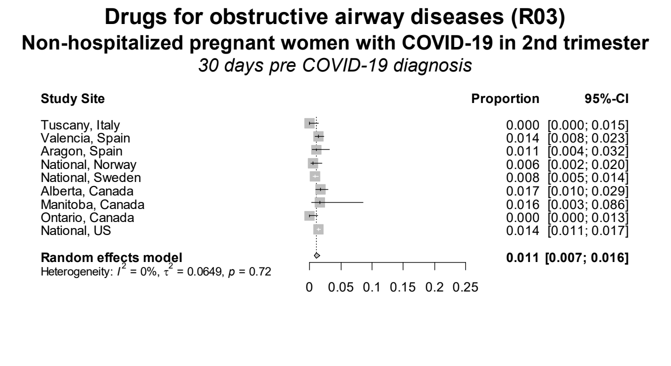

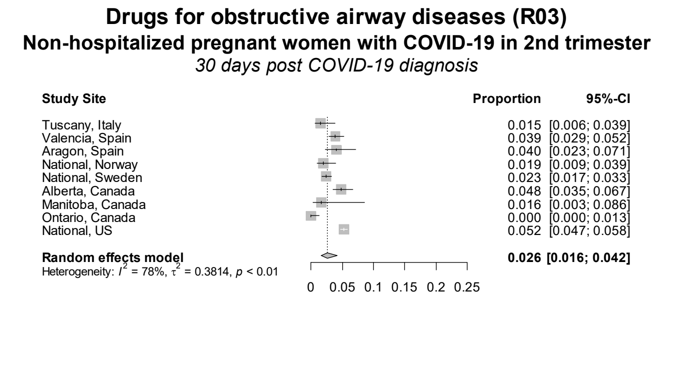

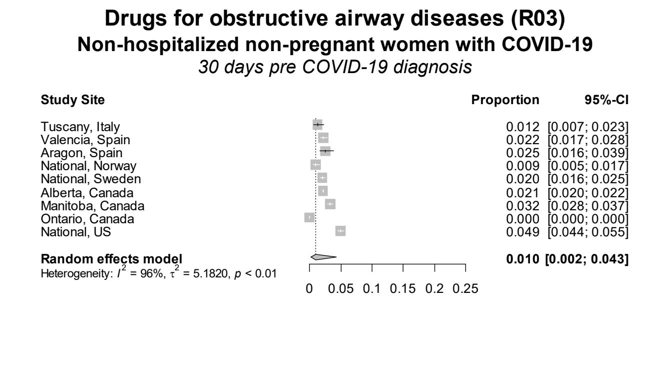

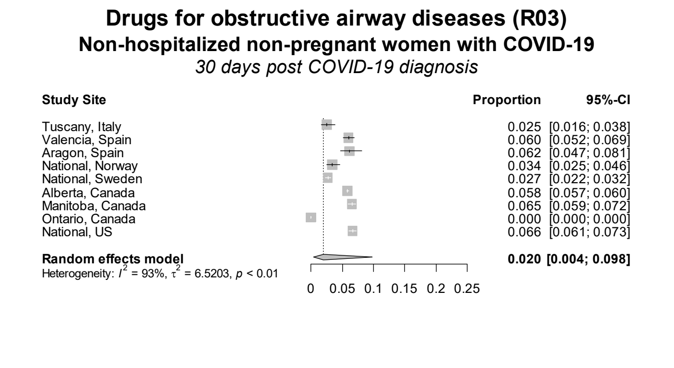

**Figure S37 continued.** Forest plots showing the pooled prevalence of drugs for obstructive airway diseases in the 30 days pre-COVID (left) and 30 days post-COVID (right) in non-hospitalized pregnant women with COVID-19 (upper) and non-hospitalized non-pregnant women with COVID-19 (lower), by pregnancy trimester

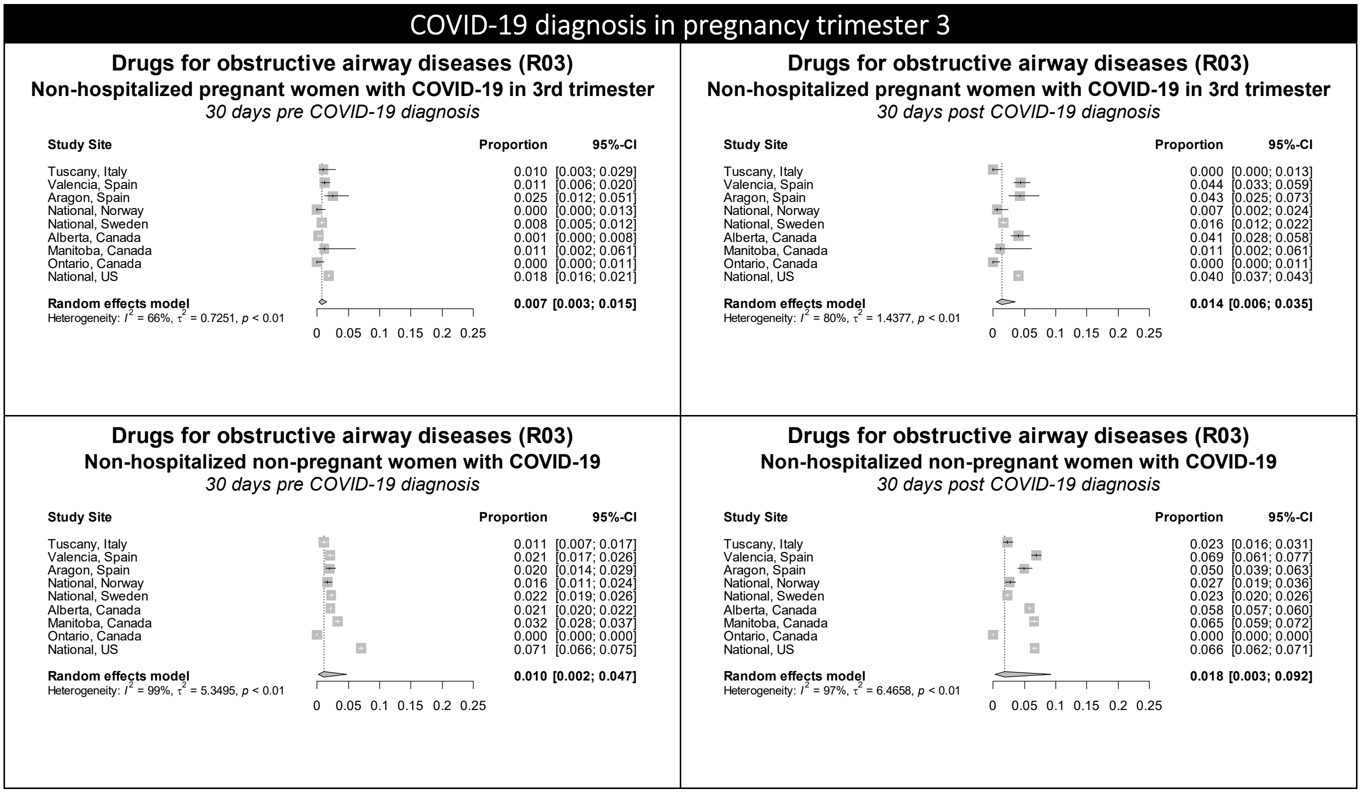

Figure S38. Forest plots showing the pooled prevalence of drugs used in diabetes in the 30 days pre-COVID (left) and 30 days post-COVID (right) in non-hospitalized pregnant women with COVID-19 (upper) and non-hospitalized non-pregnant women with COVID-19 (lower), by pregnancy trimester

### COVID-19 diagnosis in pregnancy trimester 1

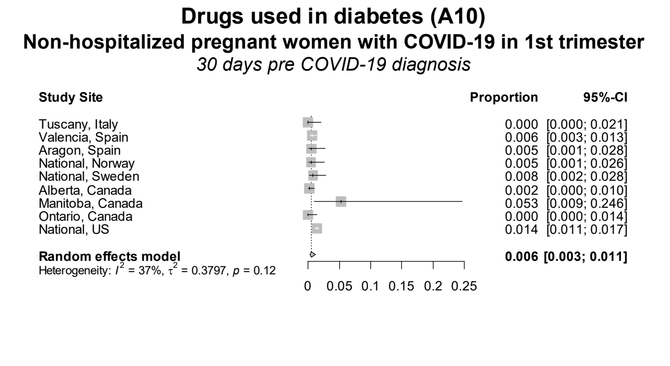

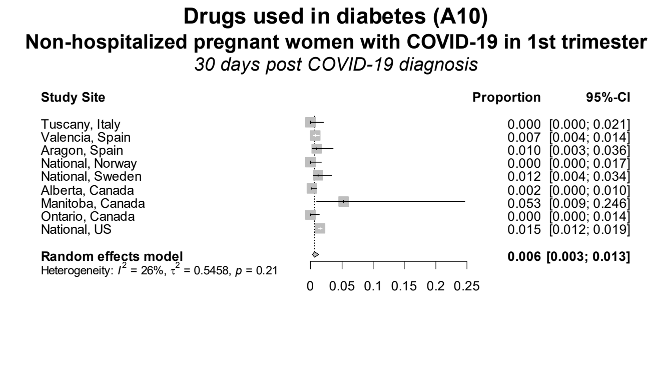

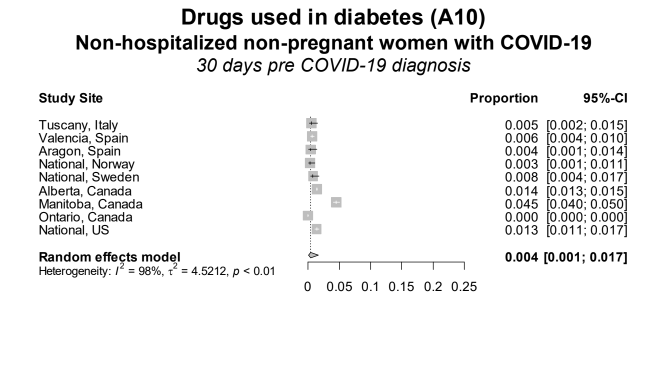

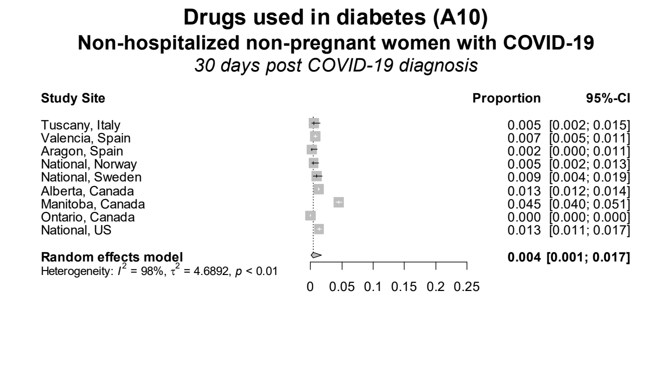

### COVID-19 diagnosis in pregnancy trimester 2

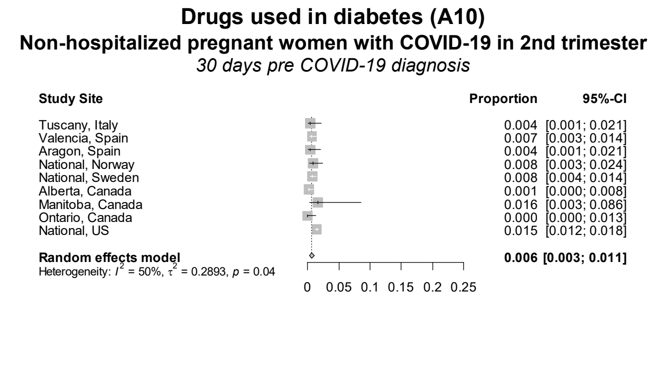

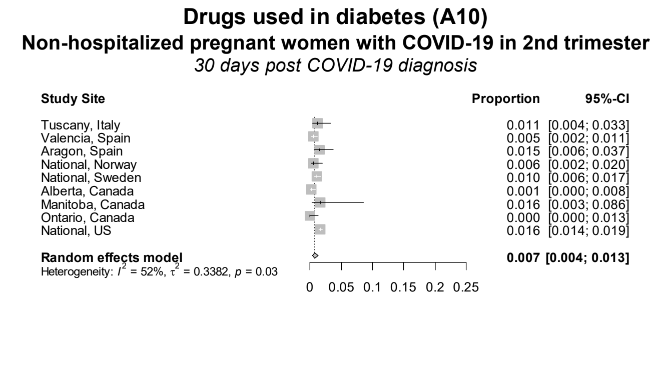

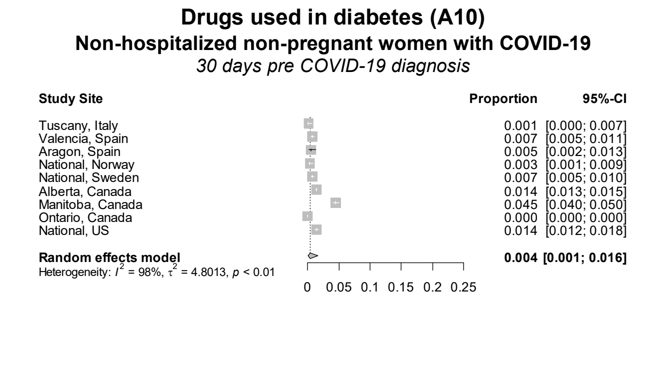

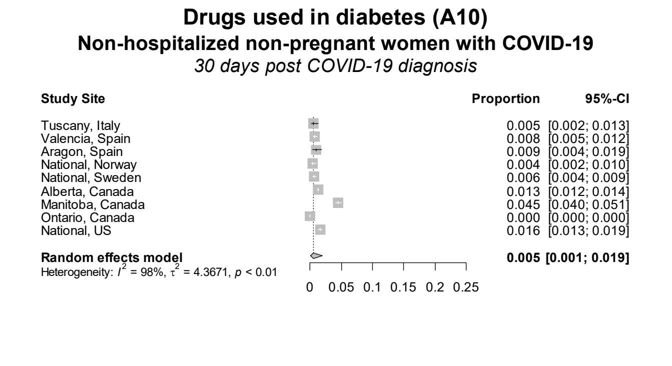

**Figure S38 continued.** Forest plots showing the pooled prevalence of drugs used in diabetes in the 30 days pre-COVID (left) and 30 days post-COVID (right) in non-hospitalized pregnant women with COVID-19 (upper) and non-hospitalized non-pregnant women with COVID-19 (lower), by pregnancy trimester

COVID-19 diagnosis in pregnancy trimester 3

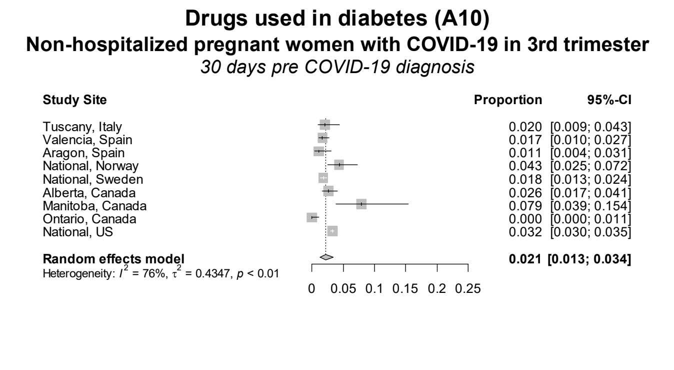

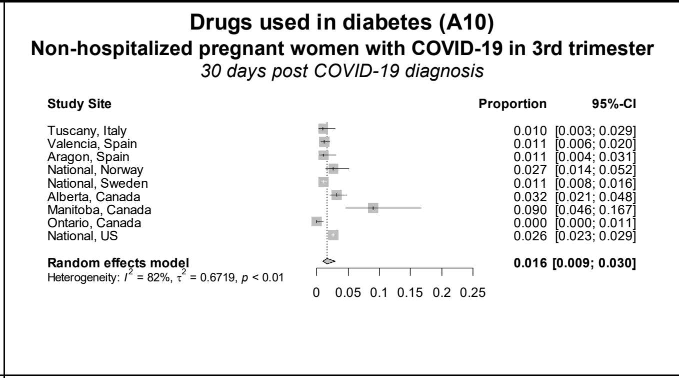

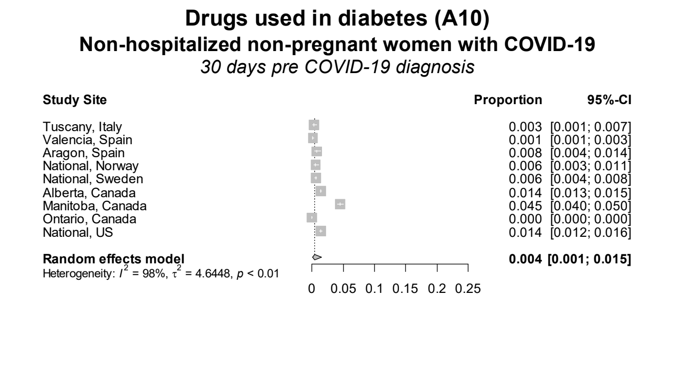

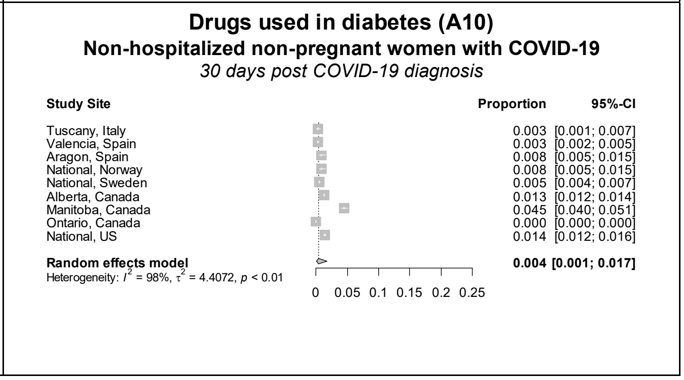

Figure S39. Forest plots showing the pooled prevalence of immune sera and immunoglobulins in the 30 days pre-COVID (left) and 30 days post-COVID (right) in non-hospitalized pregnant women with COVID-19 (upper) and non-hospitalized non-pregnant women with COVID-19 (lower), by pregnancy trimester

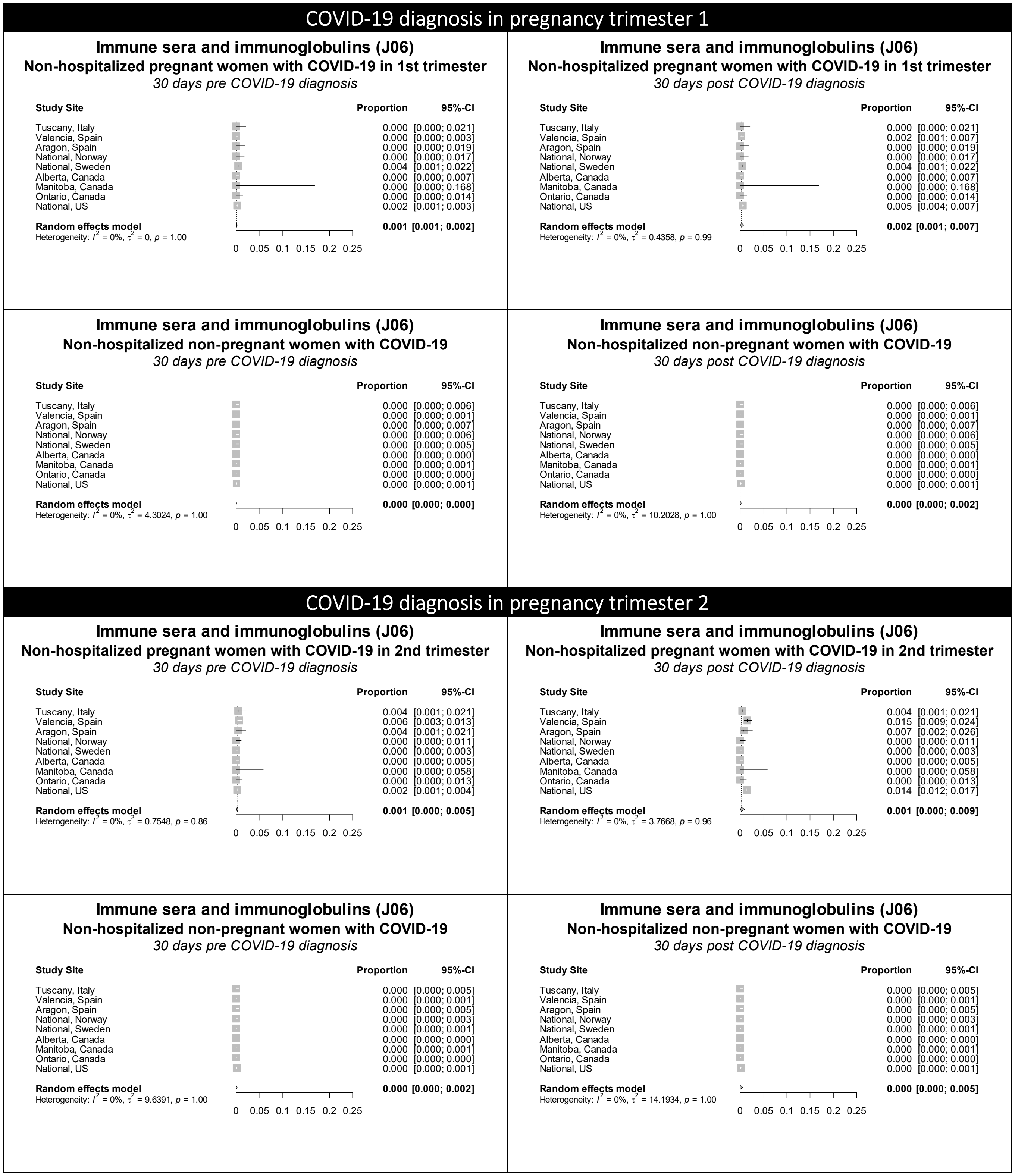

**Figure S39 continued.** Forest plots showing the pooled prevalence of immune sera and immunoglobulins in the 30 days pre-COVID (left) and 30 days post-COVID (right) in non-hospitalized pregnant women with COVID-19 (upper) and non-hospitalized non-pregnant women with COVID-19 (lower), by pregnancy trimester

COVID-19 diagnosis in pregnancy trimester 3

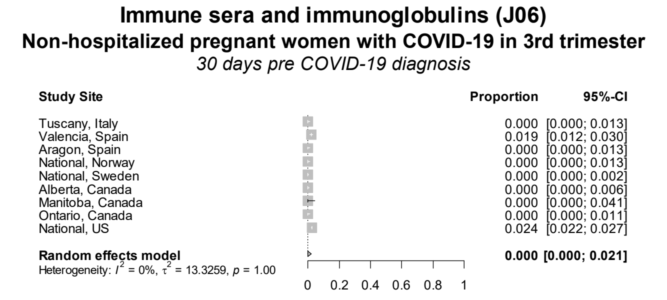

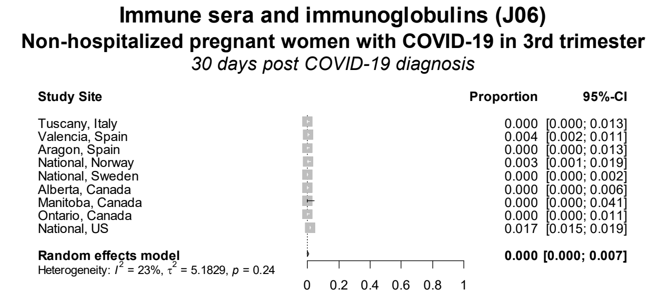

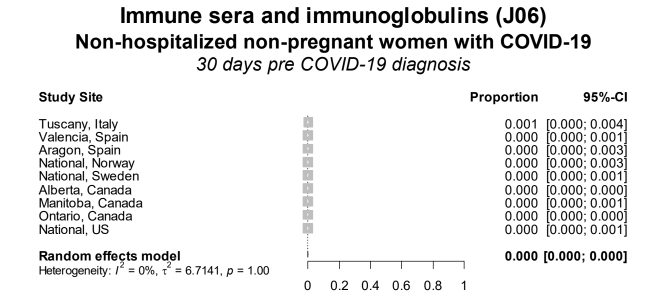

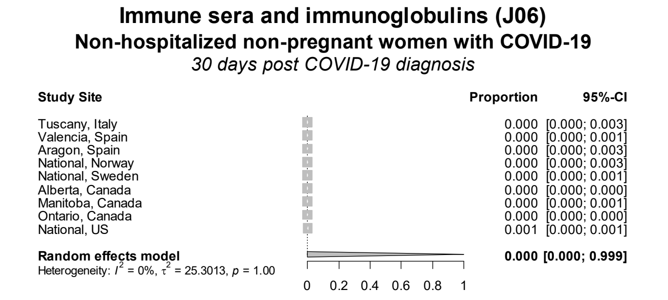

Figure S40. Forest plots showing the pooled prevalence of immunostimulants in the 30 days pre-COVID (left) and 30 days post-COVID (right) in non-hospitalized pregnant women with COVID-19 (upper) and non-hospitalized non-pregnant women with COVID-19 (lower), by pregnancy trimester

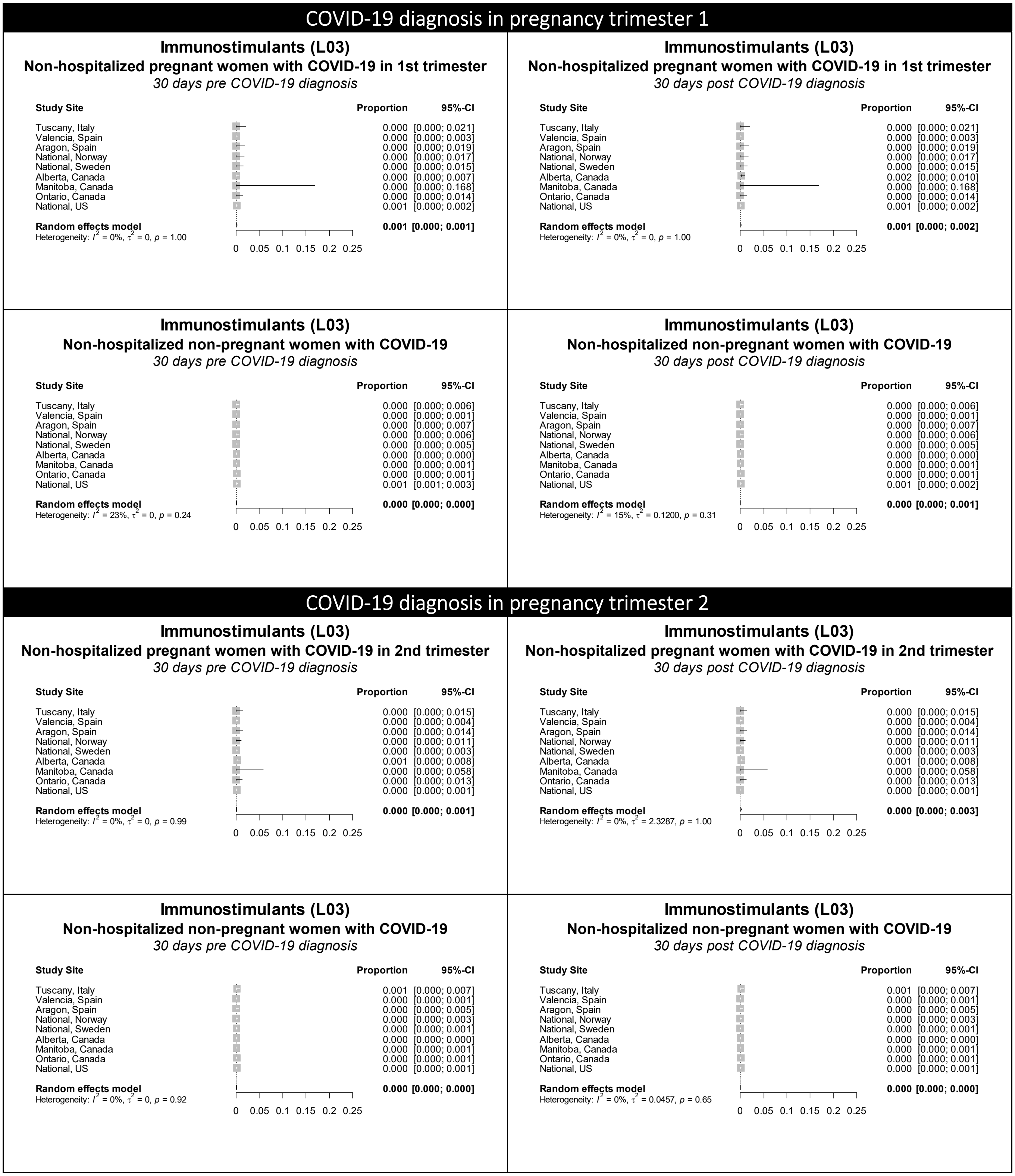

Figure S40 continued. Forest plots showing the pooled prevalence of immunostimulants in the 30 days pre-COVID (left) and 30 days post-COVID (right) in non-hospitalized pregnant women with COVID-19 (upper) and non-hospitalized non-pregnant women with COVID-19 (lower), by pregnancy trimester

### COVID-19 diagnosis in pregnancy trimester 3

#### Immunostimulants (L03)

**Non-hospitalized pregnant women with COVID-19 in 3rd trimester**

*30 days pre COVID-19 diagnosis*

| Study Site | Proportion | 95%-CI |
|---|---|---|
| Tuscany, Italy | 0.000 | [0.000; 0.013] |
| Valencia, Spain | 0.000 | [0.000; 0.004] |
| Aragon, Spain | 0.000 | [0.000; 0.013] |
| National, Norway | 0.000 | [0.000; 0.013] |
| National, Sweden | 0.000 | [0.000; 0.002] |
| Alberta, Canada | 0.000 | [0.000; 0.006] |
| Manitoba, Canada | 0.000 | [0.000; 0.041] |
| Ontario, Canada | 0.000 | [0.000; 0.011] |
| National, US | 0.000 | [0.000; 0.001] |
| **Random effects model** | **0.000** | **[0.000; 0.001]** |

Heterogeneity: $I^2 = 0\%$, $\tau^2 = 0$, $p = 1.00$

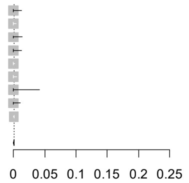

#### Immunostimulants (L03)

**Non-hospitalized pregnant women with COVID-19 in 3rd trimester**

*30 days post COVID-19 diagnosis*

| Study Site | Proportion | 95%-CI |
|---|---|---|
| Tuscany, Italy | 0.000 | [0.000; 0.013] |
| Valencia, Spain | 0.000 | [0.000; 0.004] |
| Aragon, Spain | 0.000 | [0.000; 0.013] |
| National, Norway | 0.000 | [0.000; 0.013] |
| National, Sweden | 0.000 | [0.000; 0.002] |
| Alberta, Canada | 0.000 | [0.000; 0.006] |
| Manitoba, Canada | 0.000 | [0.000; 0.041] |
| Ontario, Canada | 0.000 | [0.000; 0.011] |
| National, US | 0.000 | [0.000; 0.001] |
| **Random effects model** | **0.000** | **[0.000; 0.001]** |

Heterogeneity: $I^2 = 0\%$, $\tau^2 = 0$, $p = 1.00$

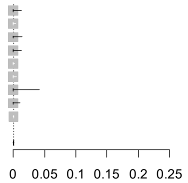

#### Immunostimulants (L03)

**Non-hospitalized non-pregnant women with COVID-19**

*30 days pre COVID-19 diagnosis*

| Study Site | Proportion | 95%-CI |
|---|---|---|
| Tuscany, Italy | 0.001 | [0.000; 0.004] |
| Valencia, Spain | 0.000 | [0.000; 0.001] |
| Aragon, Spain | 0.000 | [0.000; 0.003] |
| National, Norway | 0.001 | [0.000; 0.005] |
| National, Sweden | 0.000 | [0.000; 0.001] |
| Alberta, Canada | 0.000 | [0.000; 0.000] |
| Manitoba, Canada | 0.000 | [0.000; 0.001] |
| Ontario, Canada | 0.000 | [0.000; 0.001] |
| National, US | 0.001 | [0.001; 0.002] |
| **Random effects model** | **0.000** | **[0.000; 0.001]** |

Heterogeneity: $I^2 = 71\%$, $\tau^2 = 0.6909$, $p < 0.01$

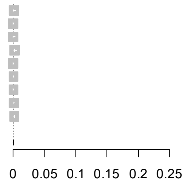

#### Immunostimulants (L03)

**Non-hospitalized non-pregnant women with COVID-19**

*30 days post COVID-19 diagnosis*

| Study Site | Proportion | 95%-CI |
|---|---|---|
| Tuscany, Italy | 0.001 | [0.000; 0.004] |
| Valencia, Spain | 0.000 | [0.000; 0.001] |
| Aragon, Spain | 0.000 | [0.000; 0.003] |
| National, Norway | 0.001 | [0.000; 0.005] |
| National, Sweden | 0.000 | [0.000; 0.001] |
| Alberta, Canada | 0.000 | [0.000; 0.000] |
| Manitoba, Canada | 0.000 | [0.000; 0.001] |
| Ontario, Canada | 0.000 | [0.000; 0.001] |
| National, US | 0.001 | [0.001; 0.002] |
| **Random effects model** | **0.000** | **[0.000; 0.001]** |

Heterogeneity: $I^2 = 76\%$, $\tau^2 = 0.8533$, $p < 0.01$

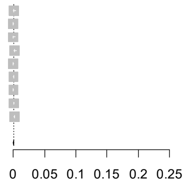

Figure S41. Forest plots showing the pooled prevalence of immunosuppressants in the 30 days pre-COVID (left) and 30 days post-COVID (right) in non-hospitalized pregnant women with COVID-19 (upper) and non-hospitalized non-pregnant women with COVID-19 (lower), by pregnancy trimester

### COVID-19 diagnosis in pregnancy trimester 1

#### Immunosuppressants (L04)
**Non-hospitalized pregnant women with COVID-19 in 1st trimester**
*30 days pre COVID-19 diagnosis*

| Study Site | Proportion | 95%-CI |
|---|---|---|
| Tuscany, Italy | 0.000 | [0.000; 0.021] |
| Valencia, Spain | 0.002 | [0.000; 0.006] |
| Aragon, Spain | 0.000 | [0.000; 0.019] |
| National, Norway | 0.000 | [0.000; 0.017] |
| National, Sweden | 0.000 | [0.000; 0.015] |
| Alberta, Canada | 0.000 | [0.000; 0.007] |
| Manitoba, Canada | 0.000 | [0.000; 0.168] |
| Ontario, Canada | 0.000 | [0.000; 0.014] |
| National, US | 0.004 | [0.003; 0.007] |
| **Random effects model** | **0.001** | **[0.000; 0.007]** |

Heterogeneity: $I^2 = 0\%$, $\tau^2 = 0.9199$, $p = 0.98$

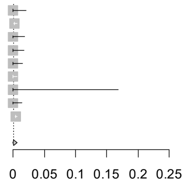

#### Immunosuppressants (L04)
**Non-hospitalized pregnant women with COVID-19 in 1st trimester**
*30 days post COVID-19 diagnosis*

| Study Site | Proportion | 95%-CI |
|---|---|---|
| Tuscany, Italy | 0.000 | [0.000; 0.021] |
| Valencia, Spain | 0.000 | [0.000; 0.003] |
| Aragon, Spain | 0.000 | [0.000; 0.019] |
| National, Norway | 0.000 | [0.000; 0.017] |
| National, Sweden | 0.000 | [0.000; 0.015] |
| Alberta, Canada | 0.000 | [0.000; 0.007] |
| Manitoba, Canada | 0.000 | [0.000; 0.168] |
| Ontario, Canada | 0.000 | [0.000; 0.014] |
| National, US | 0.005 | [0.003; 0.007] |
| **Random effects model** | **0.000** | **[0.000; 0.081]** |

Heterogeneity: $I^2 = 0\%$, $\tau^2 = 8.4140$, $p = 1.00$

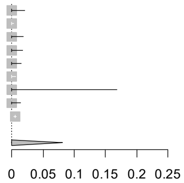

#### Immunosuppressants (L04)
**Non-hospitalized non-pregnant women with COVID-19**
*30 days pre COVID-19 diagnosis*

| Study Site | Proportion | 95%-CI |
|---|---|---|
| Tuscany, Italy | 0.002 | [0.000; 0.009] |
| Valencia, Spain | 0.001 | [0.000; 0.002] |
| Aragon, Spain | 0.004 | [0.001; 0.014] |
| National, Norway | 0.005 | [0.002; 0.013] |
| National, Sweden | 0.003 | [0.001; 0.010] |
| Alberta, Canada | 0.005 | [0.004; 0.005] |
| Manitoba, Canada | 0.000 | [0.000; 0.001] |
| Ontario, Canada | 0.004 | [0.003; 0.005] |
| National, US | 0.008 | [0.006; 0.011] |
| **Random effects model** | **0.002** | **[0.001; 0.005]** |

Heterogeneity: $I^2 = 80\%$, $\tau^2 = 0.9194$, $p < 0.01$

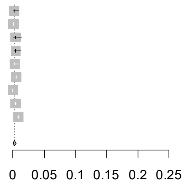

#### Immunosuppressants (L04)
**Non-hospitalized non-pregnant women with COVID-19**
*30 days post COVID-19 diagnosis*

| Study Site | Proportion | 95%-CI |
|---|---|---|
| Tuscany, Italy | 0.005 | [0.002; 0.015] |
| Valencia, Spain | 0.001 | [0.000; 0.002] |
| Aragon, Spain | 0.002 | [0.000; 0.011] |
| National, Norway | 0.003 | [0.001; 0.011] |
| National, Sweden | 0.003 | [0.001; 0.010] |
| Alberta, Canada | 0.005 | [0.004; 0.005] |
| Manitoba, Canada | 0.000 | [0.000; 0.001] |
| Ontario, Canada | 0.004 | [0.004; 0.005] |
| National, US | 0.008 | [0.006; 0.011] |
| **Random effects model** | **0.002** | **[0.001; 0.005]** |

Heterogeneity: $I^2 = 79\%$, $\tau^2 = 1.0240$, $p < 0.01$

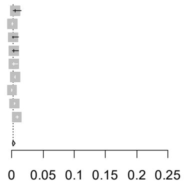

### COVID-19 diagnosis in pregnancy trimester 2

#### Immunosuppressants (L04)
**Non-hospitalized pregnant women with COVID-19 in 2nd trimester**
*30 days pre COVID-19 diagnosis*

| Study Site | Proportion | 95%-CI |
|---|---|---|
| Tuscany, Italy | 0.000 | [0.000; 0.015] |
| Valencia, Spain | 0.000 | [0.000; 0.004] |
| Aragon, Spain | 0.000 | [0.000; 0.014] |
| National, Norway | 0.003 | [0.000; 0.016] |
| National, Sweden | 0.002 | [0.001; 0.006] |
| Alberta, Canada | 0.001 | [0.000; 0.008] |
| Manitoba, Canada | 0.016 | [0.003; 0.086] |
| Ontario, Canada | 0.003 | [0.001; 0.018] |
| National, US | 0.003 | [0.002; 0.005] |
| **Random effects model** | **0.003** | **[0.001; 0.006]** |

Heterogeneity: $I^2 = 0\%$, $\tau^2 = 0.0143$, $p = 0.87$

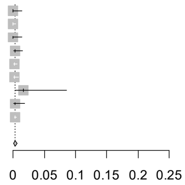

#### Immunosuppressants (L04)
**Non-hospitalized pregnant women with COVID-19 in 2nd trimester**
*30 days post COVID-19 diagnosis*

| Study Site | Proportion | 95%-CI |
|---|---|---|
| Tuscany, Italy | 0.000 | [0.000; 0.015] |
| Valencia, Spain | 0.000 | [0.000; 0.004] |
| Aragon, Spain | 0.000 | [0.000; 0.014] |
| National, Norway | 0.003 | [0.000; 0.016] |
| National, Sweden | 0.001 | [0.000; 0.004] |
| Alberta, Canada | 0.001 | [0.000; 0.008] |
| Manitoba, Canada | 0.016 | [0.003; 0.086] |
| Ontario, Canada | 0.003 | [0.001; 0.018] |
| National, US | 0.003 | [0.002; 0.004] |
| **Random effects model** | **0.002** | **[0.001; 0.004]** |

Heterogeneity: $I^2 = 0\%$, $\tau^2 = 0.0856$, $p = 0.70$

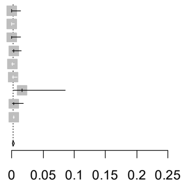

#### Immunosuppressants (L04)
**Non-hospitalized non-pregnant women with COVID-19**
*30 days pre COVID-19 diagnosis*

| Study Site | Proportion | 95%-CI |
|---|---|---|
| Tuscany, Italy | 0.002 | [0.001; 0.009] |
| Valencia, Spain | 0.002 | [0.001; 0.004] |
| Aragon, Spain | 0.003 | [0.001; 0.009] |
| National, Norway | 0.004 | [0.002; 0.010] |
| National, Sweden | 0.003 | [0.001; 0.005] |
| Alberta, Canada | 0.005 | [0.004; 0.005] |
| Manitoba, Canada | 0.000 | [0.000; 0.001] |
| Ontario, Canada | 0.004 | [0.003; 0.005] |
| National, US | 0.007 | [0.005; 0.009] |
| **Random effects model** | **0.003** | **[0.002; 0.005]** |

Heterogeneity: $I^2 = 77\%$, $\tau^2 = 0.5766$, $p < 0.01$

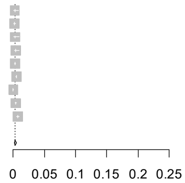

#### Immunosuppressants (L04)
**Non-hospitalized non-pregnant women with COVID-19**
*30 days post COVID-19 diagnosis*

| Study Site | Proportion | 95%-CI |
|---|---|---|
| Tuscany, Italy | 0.001 | [0.000; 0.007] |
| Valencia, Spain | 0.001 | [0.000; 0.003] |
| Aragon, Spain | 0.003 | [0.001; 0.009] |
| National, Norway | 0.003 | [0.001; 0.009] |
| National, Sweden | 0.003 | [0.002; 0.006] |
| Alberta, Canada | 0.005 | [0.004; 0.005] |
| Manitoba, Canada | 0.000 | [0.000; 0.001] |
| Ontario, Canada | 0.004 | [0.004; 0.005] |
| National, US | 0.007 | [0.005; 0.009] |
| **Random effects model** | **0.002** | **[0.001; 0.005]** |

Heterogeneity: $I^2 = 75\%$, $\tau^2 = 0.7385$, $p < 0.01$

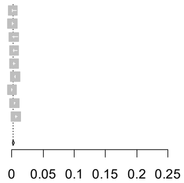

**Figure S41 continued.** Forest plots showing the pooled prevalence of immunosuppressants in the 30 days pre-COVID (left) and 30 days post-COVID (right) in non-hospitalized pregnant women with COVID-19 (upper) and non-hospitalized non-pregnant women with COVID-19 (lower), by pregnancy trimester

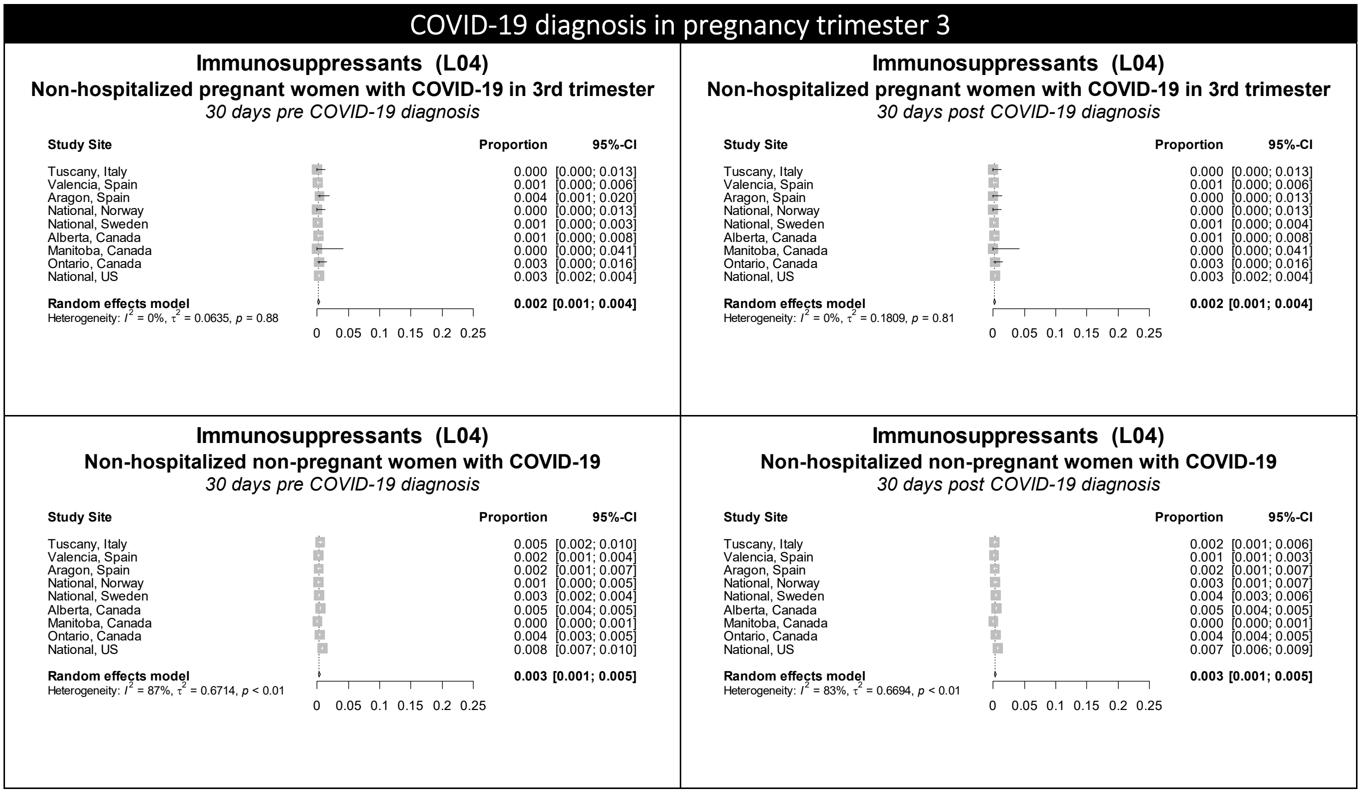

Figure S42. Forest plots showing the pooled prevalence of nasal preparations in the 30 days pre-COVID (left) and 30 days post-COVID (right) in non-hospitalized pregnant women with COVID-19 (upper) and non-hospitalized non-pregnant women with COVID-19 (lower), by pregnancy trimester

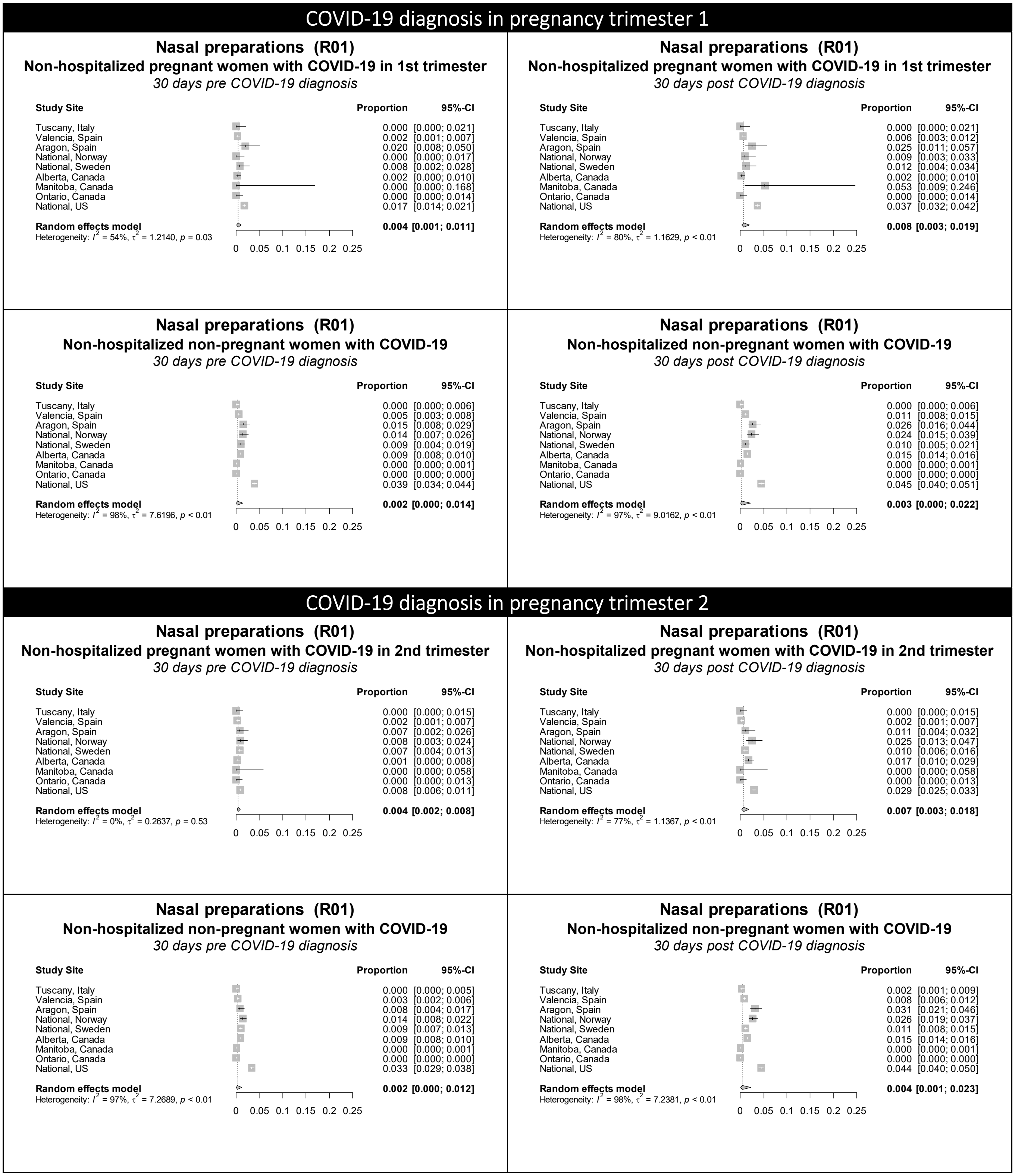

Figure S42 continued. Forest plots showing the pooled prevalence of nasal preparations in the 30 days pre-COVID (left) and 30 days post-COVID (right) in non-hospitalized pregnant women with COVID-19 (upper) and non-hospitalized non-pregnant women with COVID-19 (lower), by pregnancy trimester

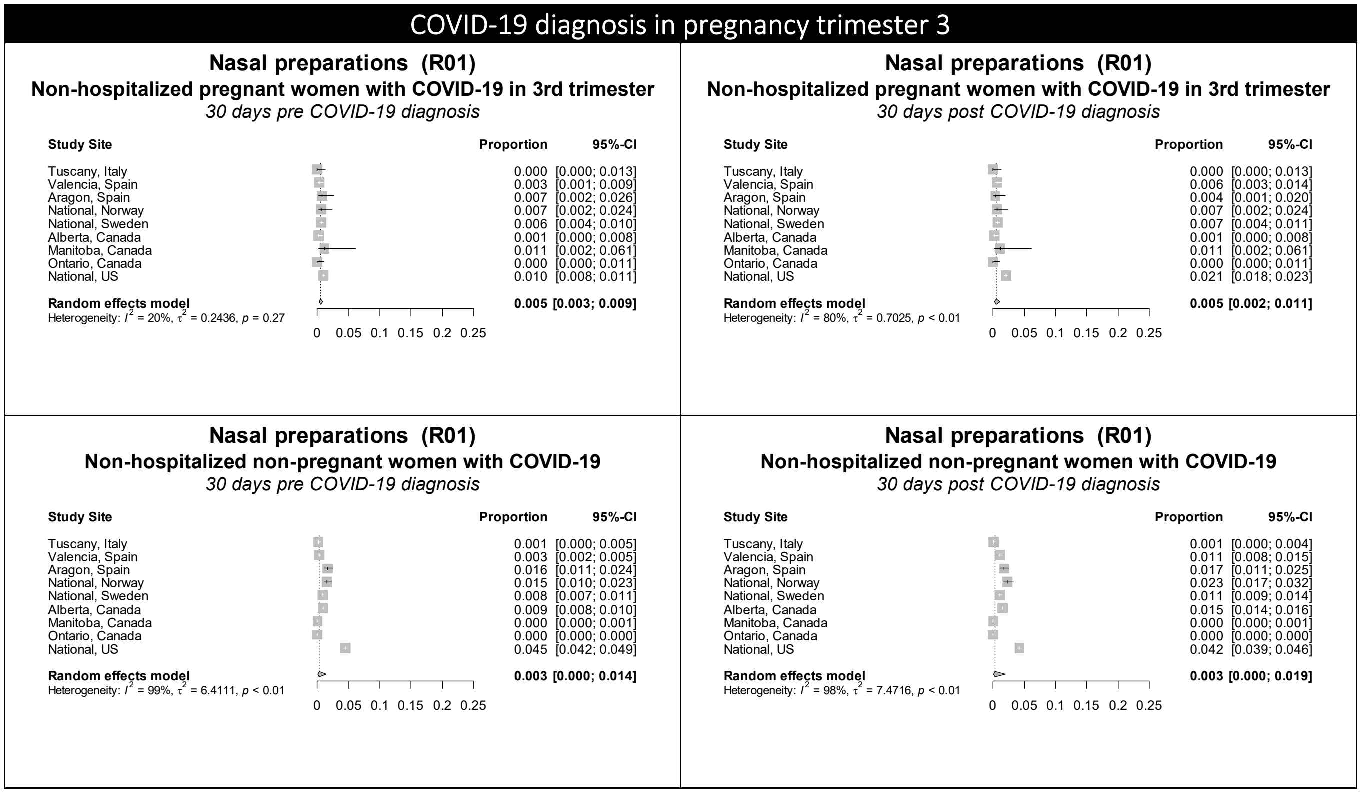

Figure S43. Forest plots showing the pooled prevalence of psychoanaleptics in the 30 days pre-COVID (left) and 30 days post-COVID (right) in non-hospitalized pregnant women with COVID-19 (upper) and non-hospitalized non-pregnant women with COVID-19 (lower), by pregnancy trimester

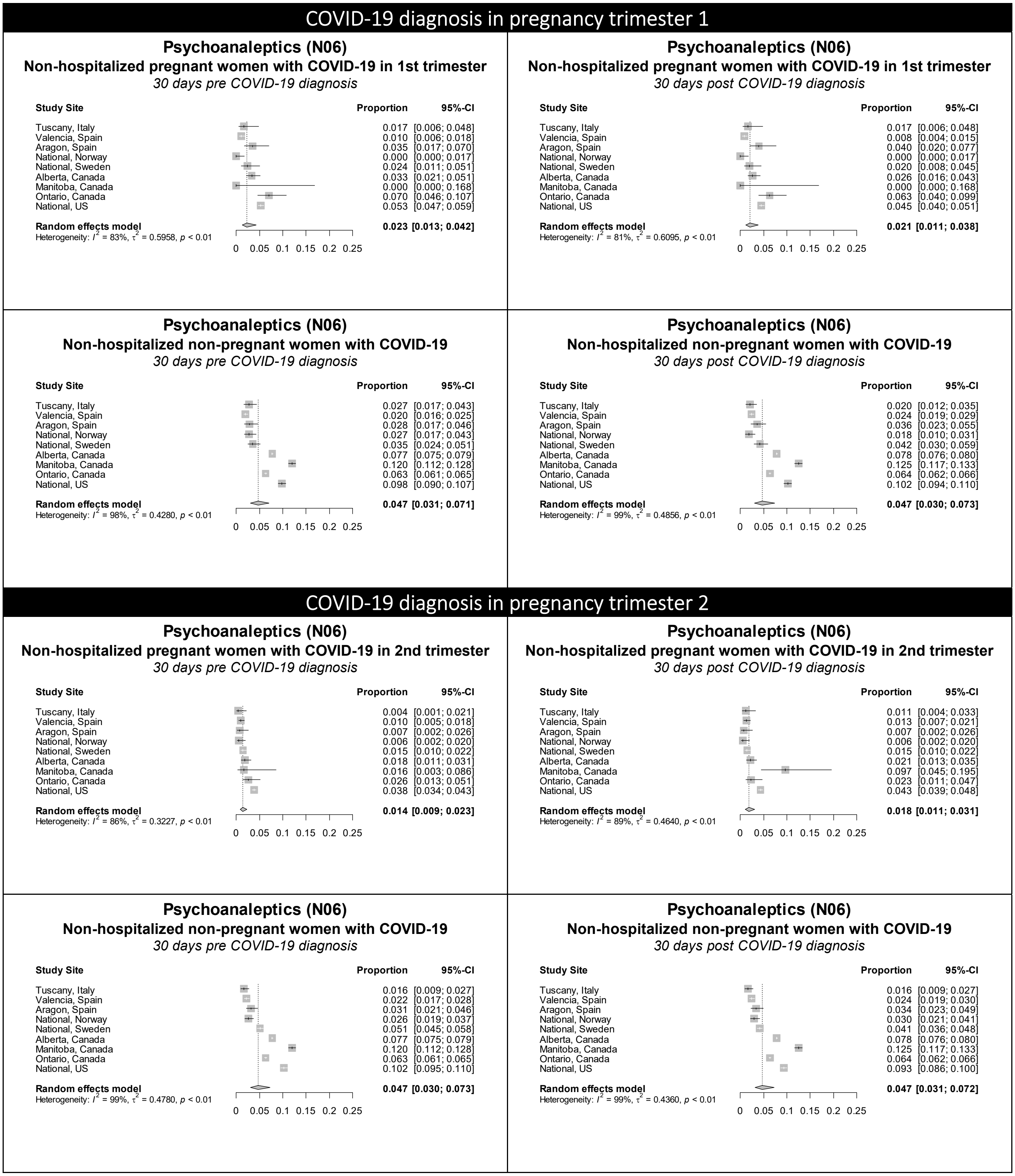

**Figure S43 continued.** Forest plots showing the pooled prevalence of psychoanaleptics in the 30 days pre-COVID (left) and 30 days post-COVID (right) in non-hospitalized pregnant women with COVID-19 (upper) and non-hospitalized non-pregnant women with COVID-19 (lower), by pregnancy trimester

### COVID-19 diagnosis in pregnancy trimester 3

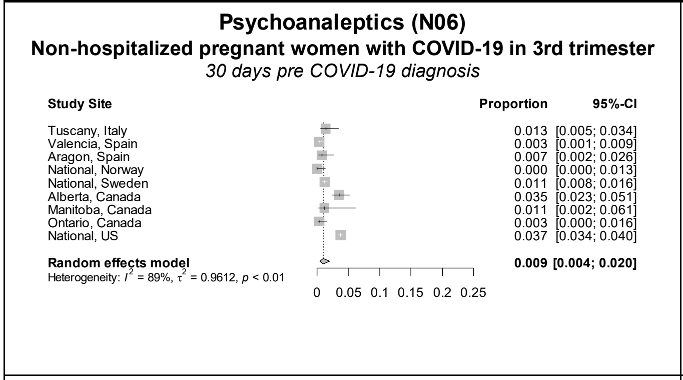

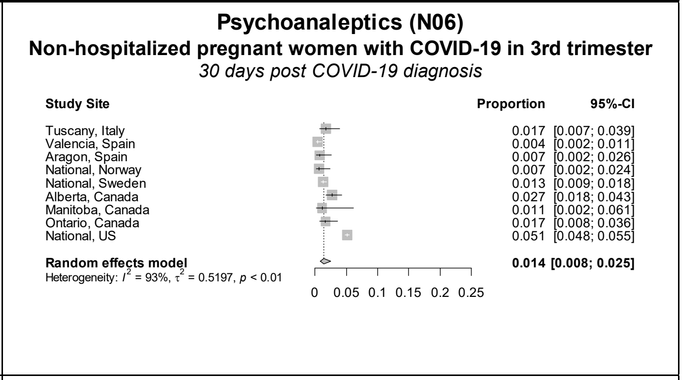

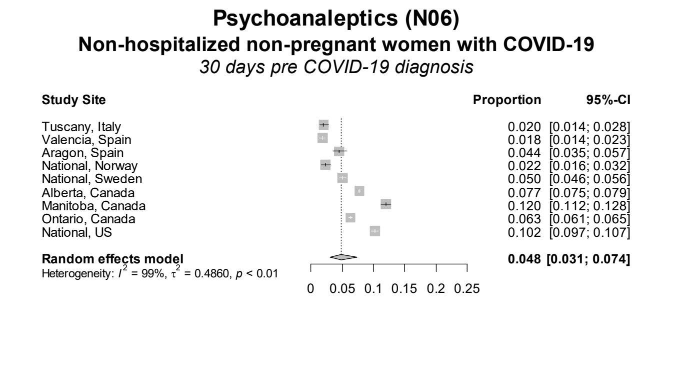

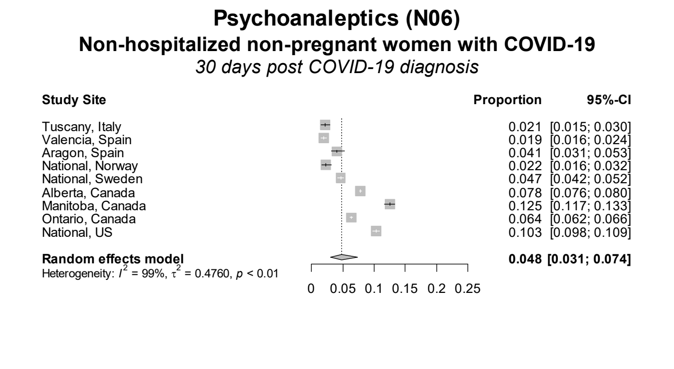

Figure S44. Forest plots showing the pooled prevalence of psycholeptics in the 30 days pre-COVID (left) and 30 days post-COVID (right) in non-hospitalized pregnant women with COVID-19 (upper) and non-hospitalized non-pregnant women with COVID-19 (lower), by pregnancy trimester

### COVID-19 diagnosis in pregnancy trimester 1

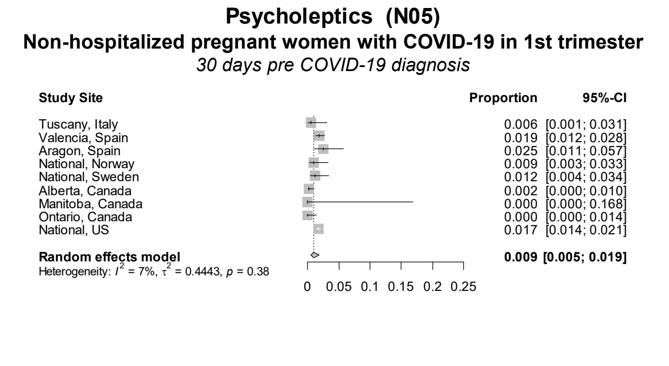

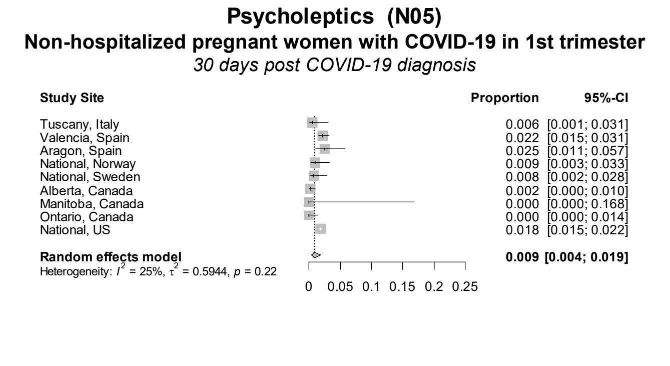

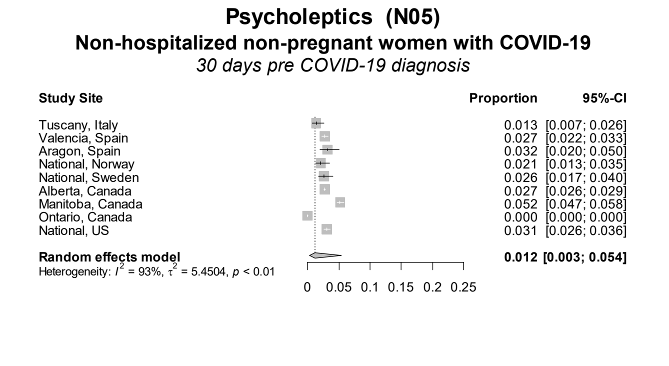

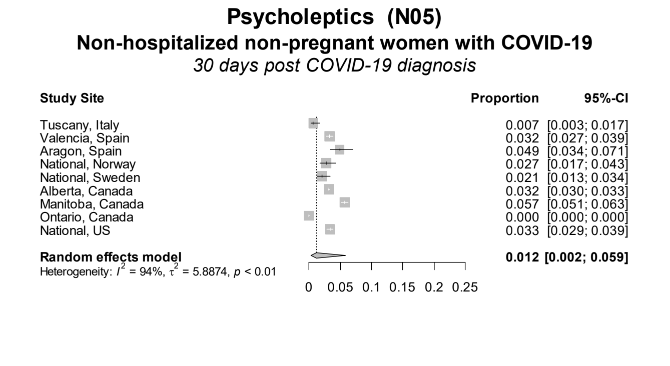

### COVID-19 diagnosis in pregnancy trimester 2

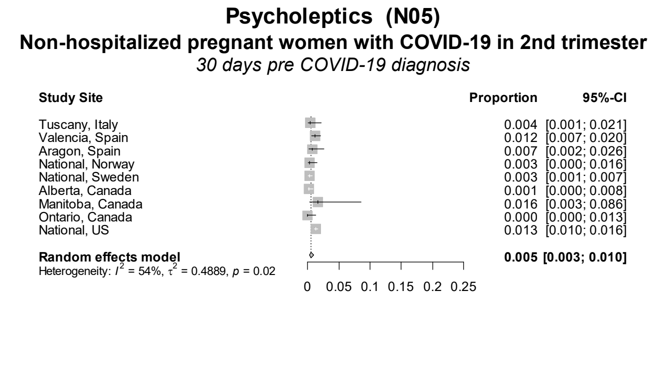

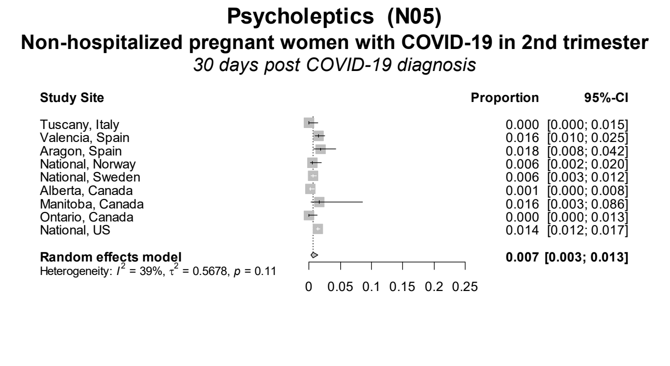

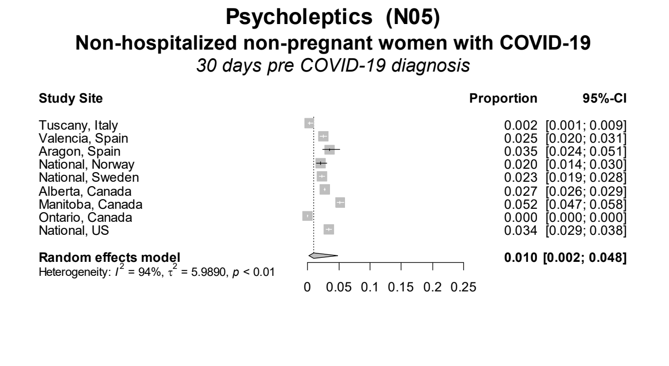

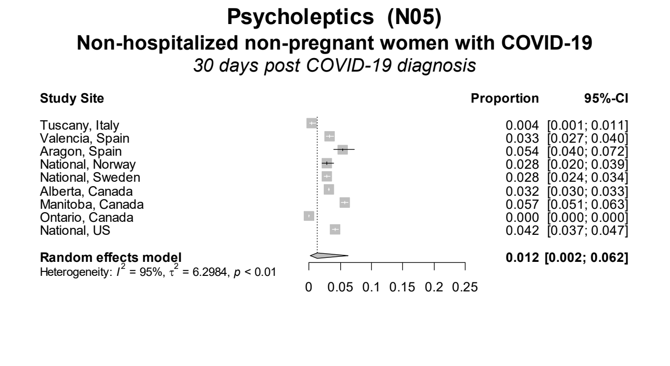

Figure S44 continued. Forest plots showing the pooled prevalence of psycholeptics in the 30 days pre-COVID (left) and 30 days post-COVID (right) in non-hospitalized pregnant women with COVID-19 (upper) and non-hospitalized non-pregnant women with COVID-19 (lower), by pregnancy trimester

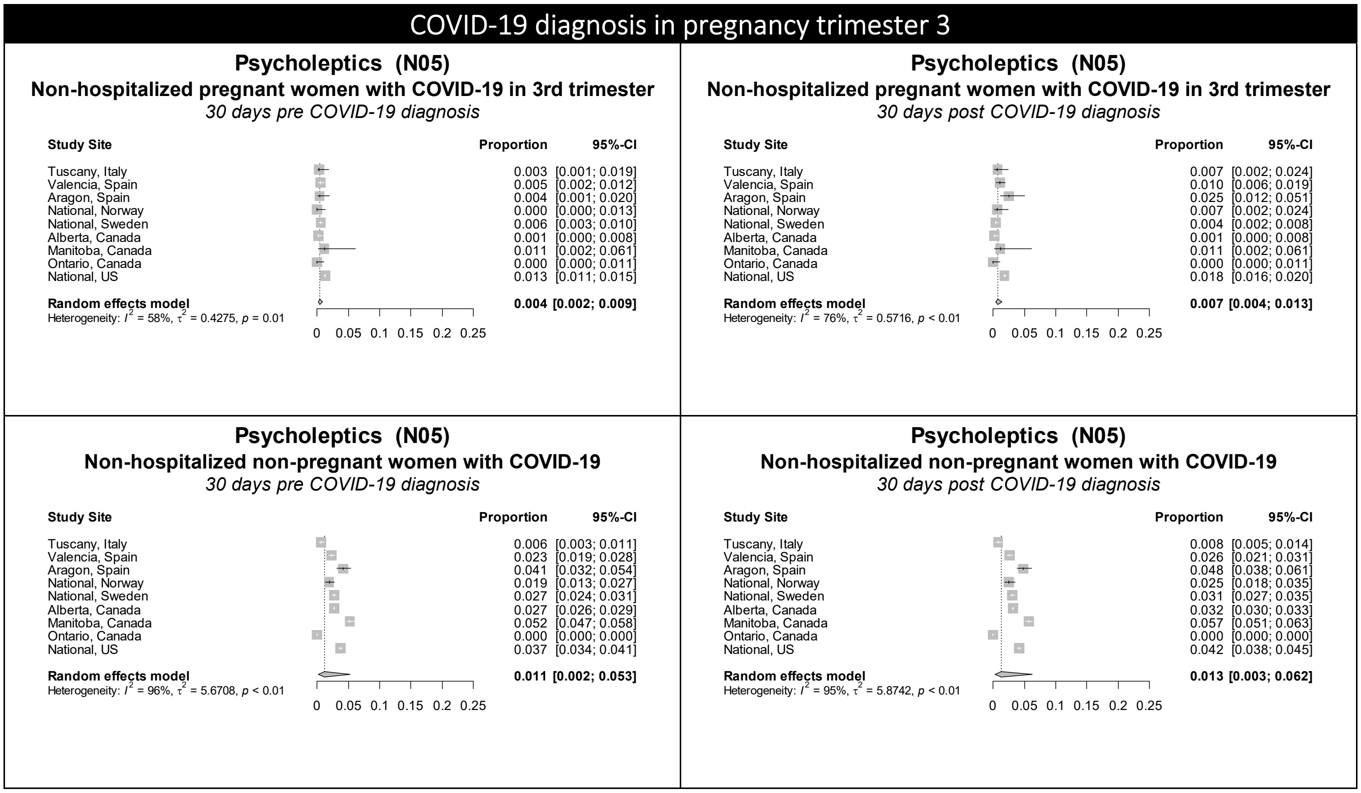
